# New insights on the genetic etiology of Alzheimer’s and related dementia

**DOI:** 10.1101/2020.10.01.20200659

**Authors:** Céline Bellenguez, Fahri Küçükali, Iris Jansen, Victor Andrade, Sonia Moreno-Grau, Najaf Amin, Adam C. Naj, Benjamin Grenier-Boley, Rafael Campos-Martin, Peter A. Holmans, Anne Boland, Luca Kleineidam, Vincent Damotte, Sven J. van der Lee, Teemu Kuulasmaa, Qiong Yang, Itziar de Rojas, Joshua C. Bis, Amber Yaqub, Ivana Prokic, Marcos R Costa, Julien Chapuis, Shahzad Ahmad, Vilmantas Giedraitis, Mercè Boada, Dag Aarsland, Pablo García-González, Carla Abdelnour, Emilio Alarcón-Martín, Montserrat Alegret, Ignacio Alvarez, Victoria Álvarez, Nicola J. Armstrong, Anthoula Tsolaki, Carmen Antúnez, Ildebrando Appollonio, Marina Arcaro, Silvana Archetti, Alfonso Arias Pastor, Beatrice Arosio, Lavinia Athanasiu, Henri Bailly, Nerisa Banaj, Miquel Baquero, Ana Belén Pastor, Luisa Benussi, Claudine Berr, Céline Besse, Valentina Bessi, Giuliano Binetti, Alessandra Bizzarro, Daniel Alcolea, Rafael Blesa, Barbara Borroni, Silvia Boschi, Paola Bossù, Geir Bråthen, Catherine Bresner, Keeley J. Brookes, Luis Ignacio Brusco, Katharina Bûrger, María J. Bullido, Vanessa Burholt, William S. Bush, Miguel Calero, Carole Dufouil, Ángel Carracedo, Roberta Cecchetti, Laura Cervera-Carles, Camille Charbonnier, Caterina Chillotti, Henry Brodaty, Simona Ciccone, Jurgen A.H.R. Claassen, Christopher Clark, Elisa Conti, Anaïs Corma-Gómez, Emanuele Costantini, Carlo Custodero, Delphine Daian, Maria Carolina Dalmasso, Antonio Daniele, Efthimios Dardiotis, Jean-François Dartigues, Peter Paul de Deyn, Katia de Paiva Lopes, Lot D. de Witte, Stéphanie Debette, Jürgen Deckert, Teodoro del Ser, Nicola Denning, Anita DeStefano, Martin Dichgans, Janine Diehl-Schmid, Mónica Diez-Fairen, Paolo Dionigi Rossi, Srdjan Djurovic, Emmanuelle Duron, Emrah Düzel, Sebastiaan Engelborghs, Valentina Escott-Price, Ana Espinosa, Dolores Buiza-Rueda, Michael Ewers, Fabrizio Tagliavini, Sune Fallgaard Nielsen, Lucia Farotti, Chiara Fenoglio, Marta Fernández-Fuertes, John Hardy, Raffaele Ferrari, Catarina B Ferreira, Evelyn Ferri, Bertrand Fin, Peter Fischer, Tormod Fladby, Klaus Fließbach, Juan Fortea, Silvia Fostinelli, Nick C. Fox, Emlio Franco-Macías, Ana Frank-García, Lutz Froelich, Daniela Galimberti, Jose Maria García-Alberca, Sebastian Garcia-Madrona, Guillermo García-Ribas, Geneviève Chene, Roberta Ghidoni, Ina Giegling, Giorgio Giaccone, Oliver Goldhardt, Antonio González-Pérez, Caroline Graff, Giulia Grande, Emma Green, Timo Grimmer, Edna Grünblatt, Tamar Guetta-Baranes, Annakaisa Haapasalo, Georgios Hadjigeorgiou, Jonathan L. Haines, Kara L. Hamilton-Nelson, EADB, Gra@ce, ADGC, Charge, DemGen, FinnGen, EADI, GERAD, Harald Hampel, Olivier Hanon, Annette M. Hartmann, Lucrezia Hausner, Janet Harwood, Stefanie Heilmann-Heimbach, Seppo Helisalmi, Michael T. Heneka, Isabel Hernández, Martin J. Herrmann, Per Hoffmann, Clive Holmes, Henne Holstege, Raquel Huerto Vilas, Marc Hulsman, Jack Humphrey, Geert Jan Biessels, Charlotte Johansson, Patrick G. Kehoe, Lena Kilander, Anne Kinhult Ståhlbom, Miia Kivipelto, Anne Koivisto, Johannes Kornhuber, Mary H. Kosmidis, Pavel P. Kuksa, Brian W. Kunkle, Carmen Lage, Erika J Laukka, Alessandra Lauria, Chien-Yueh Lee, Jenni Lehtisalo, Claudia L. Satizabal, Ondrej Lerch, Alberto Lleó, Rogelio Lopez, Oscar Lopez, Adolfo Lopez de Munain, Seth Love, Malin Löwemark, Lauren Luckcuck, Juan Macías, Catherine A. MacLeod, Wolfgang Maier, Francesca Mangialasche, Marco Spallazzi, Marta Marquié, Rachel Marshall, Eden R. Martin, Angel Martín Montes, Carmen Martínez Rodríguez, Carlo Masullo, Richard Mayeux, Simon Mead, Patrizia Mecocci, Miguel Medina, Alun Meggy, Silvia Mendoza, Manuel Menéndez-González, Pablo Mir, Maria Teresa Periñán, Merel Mol, Laura Molina-Porcel, Laura Montrreal, Laura Morelli, Fermín Moreno, Kevin Morgan, Markus M. Nöthen, Carolina Muchnik, Benedetta Nacmias, Tiia Ngandu, Gael Nicolas, Børge G. Nordestgaard, Robert Olaso, Adelina Orellana, Michela Orsini, Gemma Ortega, Alessandro Padovani, Paolo Caffarra, Goran Papenberg, Lucilla Parnetti, Florence Pasquier, Pau Pastor, Alba Pérez-Cordón, Jordi Pérez-Tur, Pierre Pericard, Oliver Peters, Yolande A.L. Pijnenburg, Juan A Pineda, Gerard Piñol-Ripoll, Claudia Pisanu, Thomas Polak, Julius Popp, Danielle Posthuma, Josef Priller, Raquel Puerta, Olivier Quenez, Inés Quintela, Jesper Qvist Thomassen, Alberto Rábano, Innocenzo Rainero, Inez Ramakers, Luis M Real, Marcel J.T. Reinders, Steffi Riedel-Heller, Peter Riederer, Eloy Rodriguez-Rodriguez, Arvid Rongve, Irene Rosas Allende, Maitée Rosende-Roca, Jose Luis Royo, Elisa Rubino, Dan Rujescu, María Eugenia Sáez, Paraskevi Sakka, Ingvild Saltvedt, Ángela Sanabria, María Bernal Sánchez-Arjona, Florentino Sanchez-Garcia, Shima Mehrabian, Pascual Sánchez-Juan, Raquel Sánchez-Valle, Sigrid B Sando, Michela Scamosci, Nikolaos Scarmeas, Elio Scarpini, Philip Scheltens, Norbert Scherbaum, Martin Scherer, Matthias Schmid, Anja Schneider, Jonathan M. Schott, Geir Selbæk, Jin Sha, Alexey A Shadrin, Olivia Skrobot, Gijsje J. L. Snijders, Hilkka Soininen, Vincenzo Solfrizzi, Alina Solomon, Sandro Sorbi, Oscar Sotolongo-Grau, Gianfranco Spalletta, Annika Spottke, Alessio Squassina, Juan Pablo Tartari, Lluís Tárraga, Niccolo Tesí, Anbupalam Thalamuthu, Thomas Tegos, Latchezar Traykov, Lucio Tremolizzo, Anne Tybjærg-Hansen, Andre Uitterlinden, Abbe Ullgren, Ingun Ulstein, Sergi Valero, Christine Van Broeckhoven, Aad van der Lugt, Jasper Van Dongen, Jeroen van Rooij, John van Swieten, Rik Vandenberghe, Frans Verhey, Jean-Sébastien Vidal, Jonathan Vogelgsang, Martin Vyhnalek, Michael Wagner, David Wallon, Li-San Wang, Ruiqi Wang, Leonie Weinhold, Jens Wiltfang, Gill Windle, Bob Woods, Mary Yannakoulia, Yi Zhao, Miren Zulaica, Manuel Serrano-Rios, Davide Seripa, Eystein Stordal, Lindsay A. Farrer, Bruce M. Psaty, Mohsen Ghanbari, Towfique Raj, Perminder Sachdev, Karen Mather, Frank Jessen, M. Arfan Ikram, Alexandre de Mendonça, Jakub Hort, Magda Tsolaki, Margaret A. Pericak-Vance, Philippe Amouyel, Julie Williams, Ruth Frikke-Schmidt, Jordi Clarimon, Jean-François Deleuze, Giacomina Rossi, Sudha Seshadri, Ole A. Andreassen, Martin Ingelsson, Mikko Hiltunen, Kristel Sleegers, Gerard D. Schellenberg, Cornelia M. van Duijn, Rebecca Sims, Wiesje M. van der Flier, Agustín Ruiz, Alfredo Ramirez, Jean-Charles Lambert

## Abstract

Alzheimer’s disease (AD) is a severe and incurable neurodegenerative disease, and the failure to find effective treatments suggests that the underlying pathology remains poorly understood. Due to its strong heritability, deciphering the genetic landscape of AD and related dementia (ADD) is a unique opportunity to advance our knowledge. We completed a meta-analysis of genome-wide association studies (39,106 clinically AD-diagnosed cases, 46,828 proxy-ADD cases and 401,577 controls) with the most promising signals followed-up in 25,392 independent AD cases and 276,086 controls. We report 75 risk loci for ADD, including 42 novel ones. Pathway-enrichment analyses confirm the involvement of amyloid/Tau pathways, highlight the role of microglia and its potential interaction with APP metabolism. Numerous genes exhibited differential expression or splicing in AD-related conditions and gene prioritization implies EGFR signaling and TNF-α pathway through LUBAC complex. We also generated a novel polygenic risk score strongly associated with the risk of future dementia or progression from mild cognitive impairment to dementia. In conclusion, by more than doubling the number of loci associated with ADD risk, our study offers new insights into the pathophysiological processes underlying AD and offers additional therapeutic entry-points and tools for translational genomics.

## Main

Alzheimer’s disease (AD) is the most common form of dementia. It exhibits a high heritability estimated between 60 and 80%^1^. This strong genetic component presents an opportunity to decipher the pathophysiological processes in AD and related dementia (ADD) and to identify novel biology, new prognosis/diagnosis markers and novel treatment targets (translational genomics). Characterizing the genetic risk factors of ADD is therefore a major objective and following the advent of the high-throughput genomic approaches, many loci/genes have been associated with the risk of ADD^2^. However, much of the underlying heritability remains unexplained and increasing the sample size of genome-wide association studies (GWAS) is an obvious route, already successfully applied in other common, complex diseases like diabetes, to facilitate the characterization of new genetic risk factors.

### GWAS analysis

The European Alzheimer’s Disease BioBank (EADB) consortium brings together the various European GWAS consortia already working on AD and a new dataset of 20,464 AD cases and 22,244 controls collated from 15 European countries. The EADB GWAS results were meta-analysed with a proxy-ADD GWAS performed in the UK Biobank (UKBB; the UKBB proxy-ADD designation is based on questionnaire data asking if parents had AD which is less specific than clinically or pathological diagnosed AD). The EADB Stage I (GWAS meta-analysis) was based on 39,106 clinically diagnosed AD cases, 46,828 proxy-ADD cases (as defined in the Supplementary Information) and 401,577 controls (Supplementary Tables 1 and 2) and on 21,101,114 variants that passed quality control measures (Fig.1 and see Supplementary Fig. 1 for QQ plot and genomic inflation factors). We selected all variants with *P* value less than 1×10^−5^ in Stage I. We defined non-overlapping regions around those variants, excluded the region corresponding to *APOE*, and examined the remaining variants in a large follow-up sample that included clinically diagnosed AD cases and controls from the ADGC, FinnGen and CHARGE consortia (Stage II; 25,392 AD cases and 276,086 controls). A signal was considered as genome-wide significant if nominally associated (*P*≤0.05) in Stage II, with same direction of association in the Stage I and Stage II analyses and if associated with ADD risk with *P*≤5×10^−8^ in the Stage I + Stage II meta-analysis. In addition, we applied a PLINK clumping procedure^3^ to define potential independent hits from the Stage I results (see Methods). After validation by conditional analyses (see Supplementary Information and Supplementary Tables 3 and 4), this approach led us to define 39 signals in 33 loci already known to be associated with the risk of developing ADD^4–10^ and to propose 42 new loci (Table 1, Supplementary Table 5 and Supplementary Fig. 2-29). Of the 42 new loci, 17 had *P*≤5×10^−8^ in Stage I and 25 were associated with *P*≤5×10^−8^ after follow-up (Stage I + Stage II meta-analysis including ADGC, CHARGE and FinnGen data). We also identified 6 loci with *P*≤5×10^−8^ in the Stage I + Stage II analysis, but failing replication in Stage II (Supplementary Table 6). Of note, the magnitude of associations in Stage I did not change substantially if we restricted the Stage I analyses to clinically-diagnosed AD cases (Supplementary Table 7 and Supplementary Fig. 30). Similarly, none of the signals observed seems to be especially driven by the UKBB data (Supplementary Table 7 and Forest plots in Supplementary Fig. 2-29). We finally provide a detailed analysis of the HLA locus (Supplementary Fig. 31-32 and Supplementary Tables 8-9).

**Table 1:**
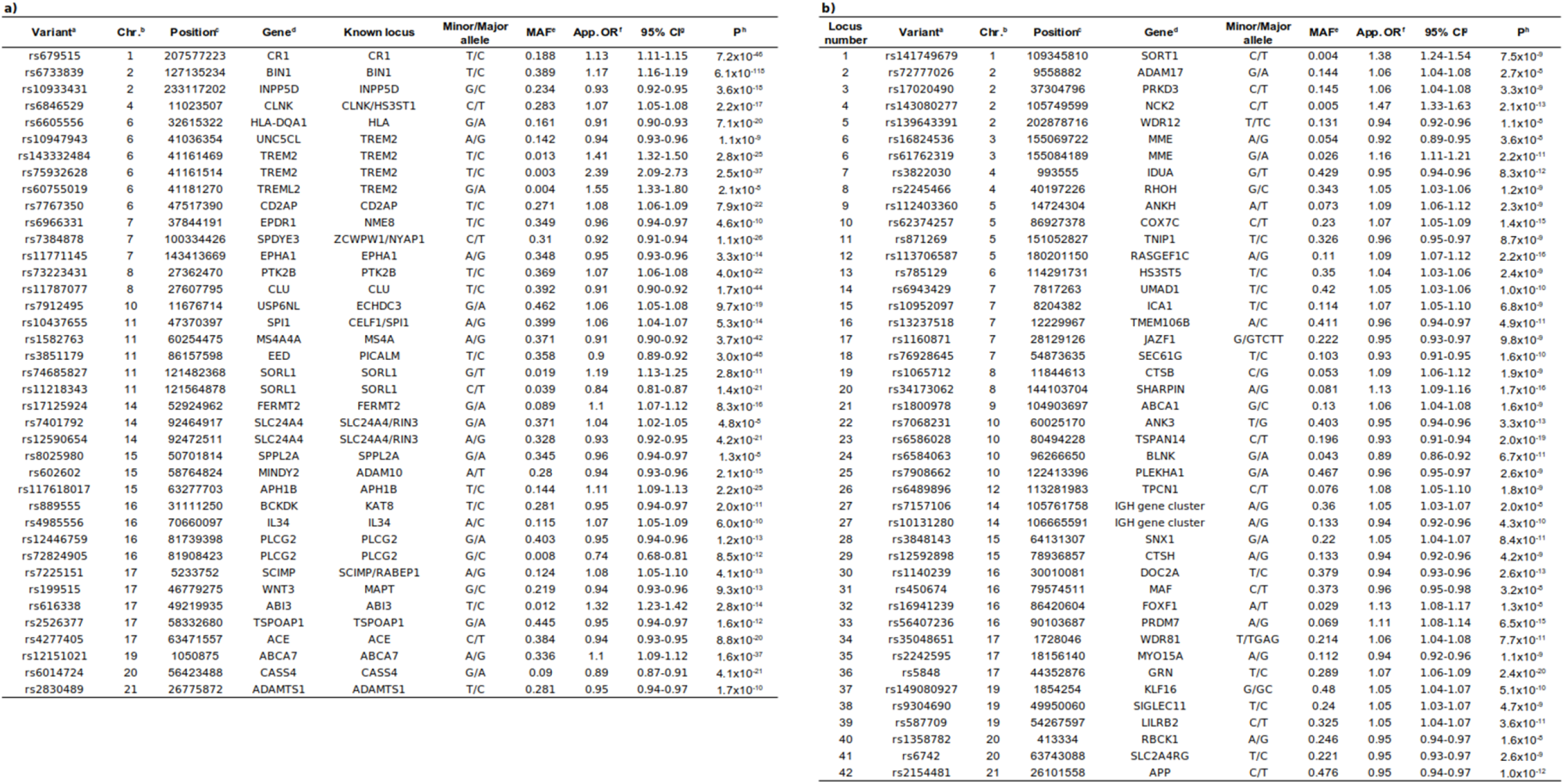
Summary of association results in Stage I + II for (a) known loci and (b) new loci with a genome-wide significant signal. ^a^Reference SNP (rs) number according to dbSNP build 153; ^b^Chromosome; ^c^GRCh38 assembly; ^d^Nearest protein coding gene according to Gencode release 33; ^e^Weighted average of minor allele frequency across discovery studies; ^f^Approximate odds-ratio calculated with respect to the minor allele; ^g^95% confidence interval; ^h^P value.

**Figure 1:**
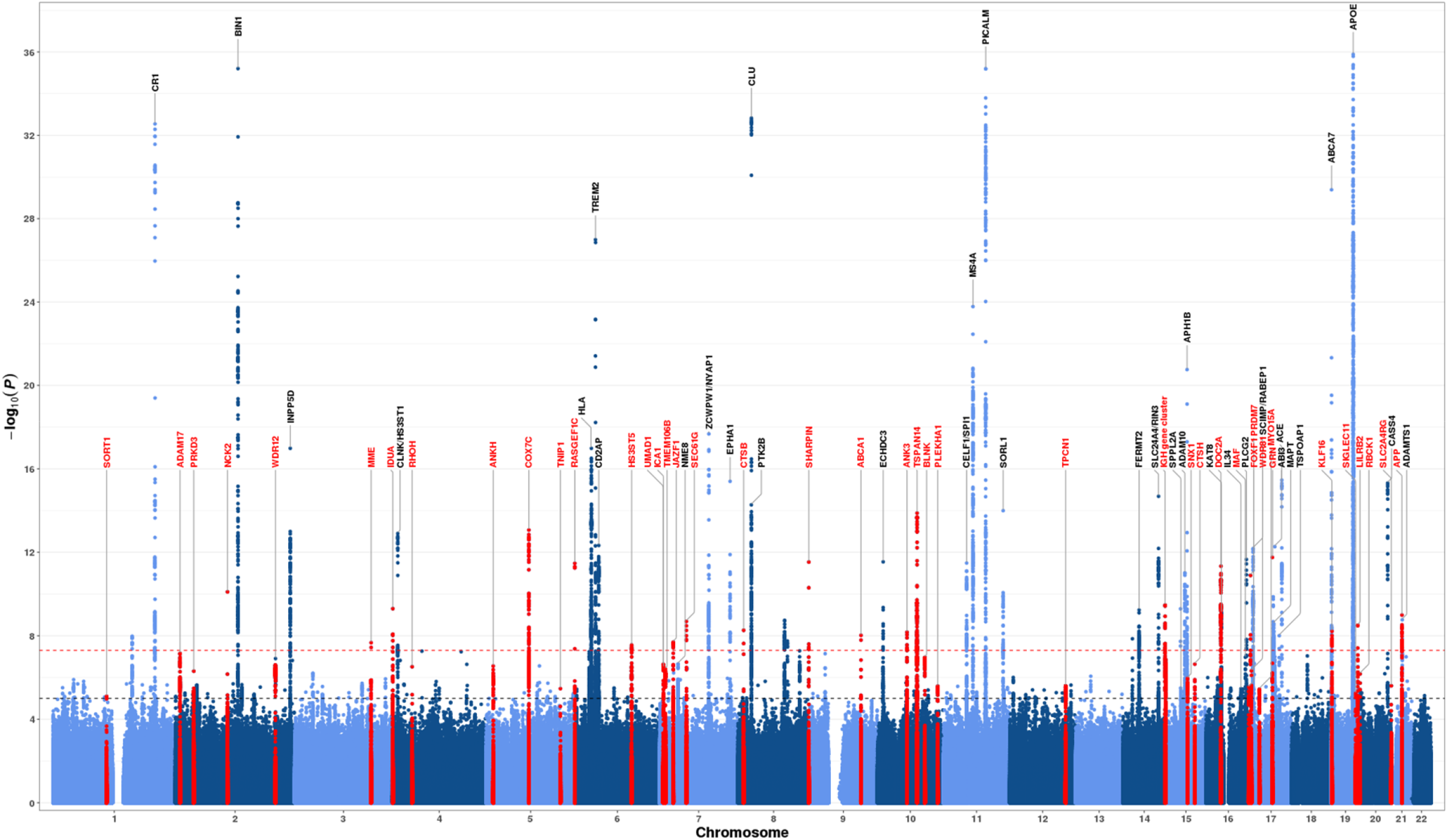
Manhattan plot of the Stage I results. Variants with P value below 1×10^−36^ are not shown. Loci with a genome-wide significant signal are annotated (known loci in black and new loci in red). Variants in new loci are highlighted in red. The red dotted line represents the genome-wide significance level (*P*=5×10^−8^), while the black dotted line represents the suggestive significance level (*P*=1×10^−5^).

We tested the association of the lead variants within our novel loci with the risk of developing other neurodegenerative diseases or AD-related disorders (Supplementary Fig. 33 and Supplementary Tables 10-12) and performed colocalization analyses for three loci known to be associated with Parkinson’s disease (PD) (*IDUA*) or frontotemporal lobar degeneration with TAR DNA binding protein (TDP-43) inclusions (FTLD-TDP) (*TMEM106B* and *GRN*). The *IDUA* signal in PD is independent of the signal in AD (coloc PP3=99.9%) while the *TMEM106B* and *GRN* signals in FTLD-TDP probably share the same causal variants with AD (coloc PP4=99.8% and PP4=80.1% respectively, Supplementary Tables 13-14).

### Pathway analyses

To better understand the novel biological insights emerging from this newly expanded AD genetic landscape, we first performed pathway enrichment analyses in Stage I association results. 150 gene sets remained significant after multiple testing correction (*q*≤0.05, see Methods, Supplementary Table 15). The most significant gene sets relate to amyloid and tau; other significant gene sets relate to lipids and immunity, including macrophage and microglial cell activation.

We then performed a single-cell expression enrichment analysis using two complementary measures in different human brain cell types: average gene expression per nucleus (Av. exp) and percentage of nuclei in a cell type expressing each gene (% Cell exp). Only microglial expression of these genes reached significance across the two measurements after correcting for multiple testing (Supplementary Table 16), with increased expression corresponding to more significant association with ADD. A similar result was also observed using the mouse single cell dataset (Supplementary Table 17). We finally tested whether the relationship between microglial expression and association with ADD risk was specific to particular areas of biology (Supplementary Table 18). The most significant interaction was detected between GO:1902991 (regulation of amyloid precursor protein catabolic process) and gene expression level in microglia (*q*=1.2×10^−12^ and 8.3×10^−8^ for % Cell Exp. and Av. Exp. respectively). Thus, genes in the amyloid-beta pathways with highest microglial expression show the strongest association with ADD, suggesting a functional relationship between microglia and APP/Aβ pathways (Supplementary Table 19).

### Gene prioritization

We next attempted to identify those genes most likely to be responsible for the ADD association signal at each locus. All the prioritized genes are summarized in Fig. 2 with a full description of the genes analyzed in the Supplementary Information (see also Supplementary Tables 20-30 and Supplementary Fig. 34-42).

**Figure 2:**
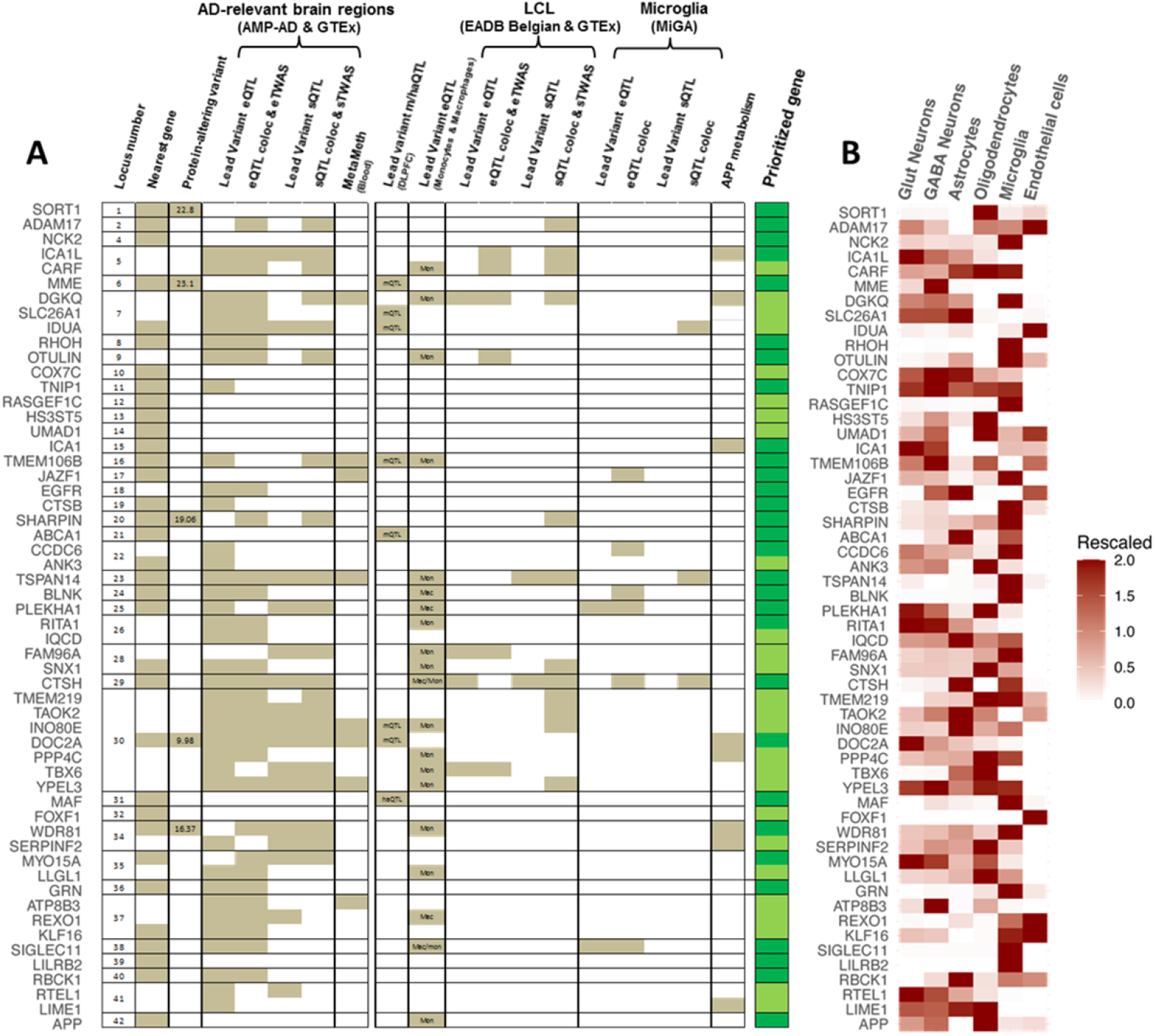
(A) Top candidate genes in the 42 new genome-wide-significant loci and (B) their level of expression in different brain cell types. In order to prioritize candidate genes in the new loci, we considered the nearest protein-coding gene from the lead variant and the genes exhibiting AD-related modulations within a region of 1 Mb around the lead variant. The average expression of each gene expressed by at least 10% of cells (pct.exp > 0.1) was rescaled from 0 to 2, allowing the identification of genes expressed by unique or multiple cell classes. On the left panel, the brown squares represent significant hits for the respective columns. The prioritized genes in each locus are marked with dark green colour, meanwhile light green colour is used when several candidate genes were retained in the same locus and/or the overall evidence is weaker than the evidence for the prioritized genes with the dark green squares. CADD (v1.6) PHRED scores of protein-altering lead variants are shown in the respective column. The columns demonstrating lead variant m/haQTL effects in DLPFC and lead variant eQTL effects in naive state monocyte, macrophages and microglia are annotated for the type of association (mQTL: methylation QTL, haQTL: histone acetylation QTL, mon: monocyte eQTL, mac: macrophage eQTL).

For ten of the novel loci, brain molecular quantitative trait loci (QTL) analyses and transcriptome-wide association studies (TWAS) in AD-relevant brain regions, blood genetics-driven methylation (MetaMeth) analyses and/or APP metabolism results supported the genes closest to the lead variant: *TNIP1* (L11), *ICA1* (L15), *TMEM106B* (L16), *JAZF1* (L17), *CTSB* (L19), *ABCA1* (L21), *CTSH* (L29), *MAF* (L31), *SIGLEC11* (L38) and *RBCK1* (L40). Five other genes in the new loci can be prioritized as likely to be causal since the lead variants correspond to protein-altering variants within the gene itself: *SORT1* (L1), *MME* (L6), *SHARPIN* (L20), *DOC2A* (L30) and *WDR81* (L34). For *SHARPIN, DOC2A*, and *WDR81*, we found additional evidences that ADD risk is associated with expression and/or splicing events. Of note, while *DOC2A* and *WDR81* are the most likely candidates, these loci also include other plausible genes (Fig. 2).

For seven loci: *NCK2* (L4), *COX7C* (L10), *RASGEF1C* (L12), *HS3HT5* (L13), *UMAD1* (L14), *FOXF1* (L32) and *APP* (L42), none of the genes presented any AD-related modulations (see Methods), therefore we considered that their proximity to the lead variant was in favor of their prioritization but at this stage at a lower level of confidence, except for *APP* (an obvious candidate for ADD) and *NCK2* (where the lead variant is rare).

The remaining twenty novel loci present a more complicated pattern since several genes exhibit AD-related modulations at the same locus, and/or the prioritized gene is not the nearest protein-coding gene. We could efficiently prioritize candidate risk genes in twelve additional loci: *ADAM17* (L2), *ICA1L* (L5), *RHOH* (L8), *OTULIN* (L9), *EGFR* (L18), *CCDC6* (L22), *TSPAN14* (L23), *BLNK* (L24), *PLEKHA1* (L25), *RITA1* (L26), *MYO15A* (L35) and *GRN* (L36). Some of these loci also include other genes of potential interest. In locus 18, *EGFR* is a likely candidate gene: its eQTL signals colocalize with the ADD risk association signal, and its fine-mapped eTWAS hits (with FOCUS posterior inclusion probability values of ∼1) associate a predicted increased *EGFR* expression with increased ADD risk (Fig. 3, Supplementary Fig. 39). In the complex locus 23, *TSPAN14* was identified as exhibiting numerous AD-related expression and splicing modulations, including novel cryptic complex splicing events that we identified and experimentally confirmed (Fig. 3, Supplementary Fig. 40). In the remaining eight loci, we did not clearly identify a single candidate but current evidence points towards the following genes: *DGKQ, SLC26A1* and *IDUA* in locus 7, *FAM96A* and *SNX1* in locus 28, *ATP8B3, REXO1* and *KLF16* in locus 37, *RTEL1* and *LIME1* in locus 41. For the locus 39, we consider *LILRB2* as the plausible risk gene based on literature review^13,14^. Finally, we were not able to prioritize a gene in the complex IGH cluster (L27), nor in the *PRKD3* (L3) and *PRDM7* (L33) loci.

**Figure 3:**
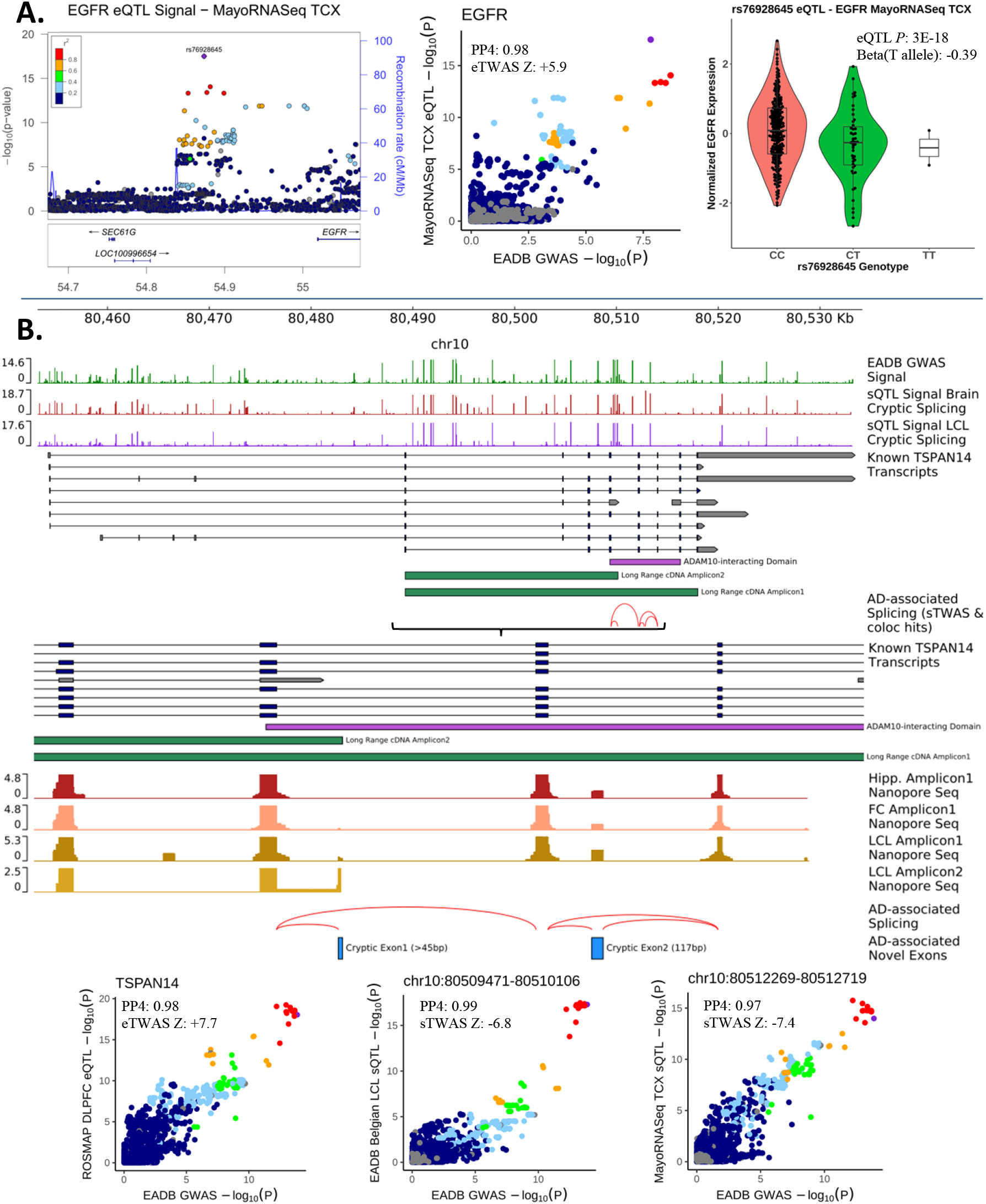
QTL mapping, QTL colocalization, and TWAS results on SEC61G and TSPAN14 loci. (a) An intergenic, distant, and low-frequency cis-eQTL for *EGFR* locus colocalizes (PP4: 0.98) with EADB GWAS signal where genetic downregulation of *EGFR* is significantly associated with lower ADD risk. (b) Elucidation of *TSPAN14* locus: through e/sQTLs, the protective signal is associated with decreased *TSPAN14* expression and increased cryptic splicing (within ADAM10-interacting domain) that are confirmed by long-read single-molecule sequencing on cDNA derived from hippocampus (Hipp.), frontal cortex (FC), lymphoblastoid cell lines (LCL). GWAS and e/sQTL association signals are shown as – log_10_(*P*) and cumulative coverage tracks are in log_10_ scale. The purple dots show the lead variants in the investigated loci, and LD r^2^ values were calculated with respect to these lead variants using the 1000 Genomes non-Finnish European LD reference panel prepared for the TWAS analyses.

We performed protein-protein interaction (PPI) analyses based on previously known GWAS genes, our prioritized novel genes (dark green in Figure 2 and Supplementary Table 20) and a combination of both lists (see Supplementary Information section 11 for a description of the method). The largest network was 14, 8 and 31 proteins respectively (Supplementary Fig. 43). These networks were larger than expected by chance (*P*<2×10^−5^, *P*<3.9×10^−3^, *P*<2×10^−5^ respectively). Notably, the number of interactions between our prioritized genes and previously known genes is also significantly larger than expected (*P*=1×10^−4^), indicating the biological relevance of the newly prioritized genes to AD pathology. No such enrichment (p=0.77) was observed for the remaining genes in the new loci, highlighting again the value of our prioritization approach.

### Polygenic risk score

We explored whether the genetic ADD burden as measured by a polygenic risk score (PRS) generated from our genome-wide significant variants (n=83, see Methods and Supplementary Table 31) might influence rate of progression to dementia starting from either normal cognition or mild cognitive impairment (MCI) using several longitudinal population-based and memory clinic cohorts (Supplementary Table 32).

In a meta-analysis of population-based studies, the PRS was significantly associated with risk of progression to all-causes of dementia (HR=1.05 per average risk variant, 95%CI [1.03-1.06], *P*=1.2×10^−13^) and to AD (HR=1.06 per average risk variant, 95%CI [1.04-1.07], *P*=1.1×10^−12^) (Figure 4). Results were confirmed in the Rotterdam population-based study: a PRS based on 77 variants (due to missing variant information) was also significantly associated with risk of progression to all-causes of dementia (HR=1.09 per average risk variant, 95%CI [1.08-1.11], *P*=7.6×10^−32^) or with the risk of progression to AD (HR=1.10 per average risk variant, 95%CI [1.08-1.12], *P*=1.0×10^−27^).

**Figure 4:**
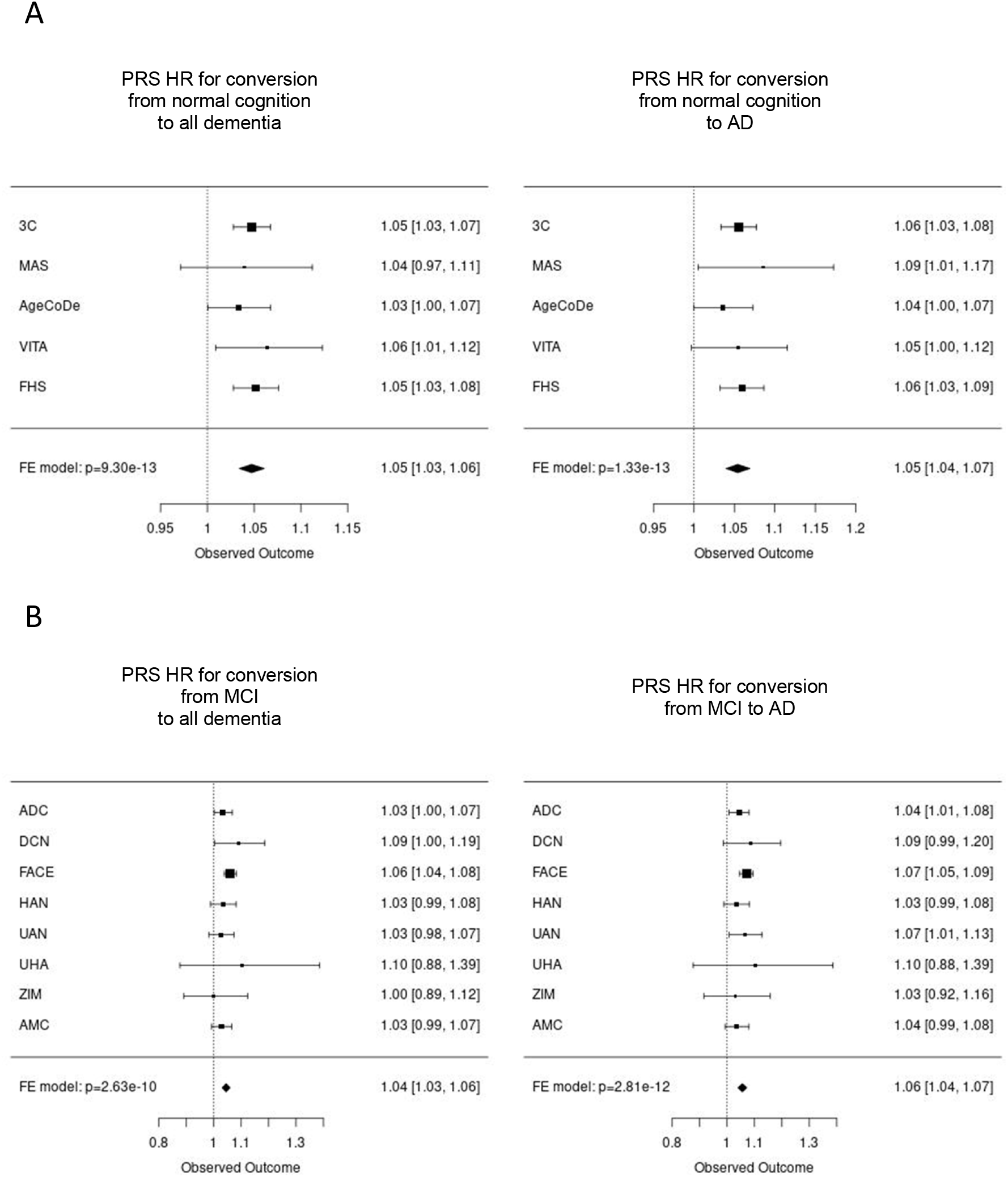
Association of PRS with the risk of progression to dementia starting from either (A) normal cognition or (B) mild cognitive impairment (MCI). Polygenic risk score (PRS) was based on the genetic data of 83 variants (see Methods and Supplementary Table 31). HR: Hazard Ratio; MCI: Mild cognitive impairment; FE: fixed-effect; *P* het: heterogeneity *P* value

Similarly, in MCI patients from memory clinics, we observed a significant association of this PRS with the risk of progressing to all-causes of dementia (HR=1.04 per average risk variant, 95%CI [1.03-1.06], *P*=2.6×10^−10^) or AD (HR=1.06 per average risk variant, 95%CI [1.04-1.07], *P*=2.8×10^−12^) (Figure 4). All analyses were adjusted for age, sex, principal components and the number of APOE-ε4 and APOE-ε2 alleles (Figure 4, Supplementary Table 33).

Association of the PRS with the risk of dementia progression was not modified by APOE-ε4 (interaction *P*=0.95 and 0.44 in population-based or MCI studies respectively; see also APOE-ε4 strata analyses in Supplementary Fig. 44-45). As previously reported, APOE-ε4 influenced progression to all-causes of dementia (Supplementary Table 33).

## DISCUSSION

This meta-analysis combining a new large case-control study and previous GWAS identified 75 independent genetic risk factors for ADD, 33 previously reported and 42 corresponding to novel signals, greatly expanding our knowledge of the complex genetic landscape of AD. A short description of the prioritized genes and their potential significance in AD pathophysiology is presented in the Supplementary Information.

Our meta-analysis and the characterization of these new loci refine and expand our current understanding of AD etiology. The pathway enrichment analyses remove ambiguities concerning the involvement of Tau binding proteins and APP/Aβ metabolism in late-onset AD processes. In addition, we implicated for the first time *ADAM17*, a gene whose protein product is known to carry α-secretase activity as ADAM10^15^. This observation affirms that a deregulation of the non-amyloidogenic pathway of APP metabolism is also important in the AD process. In addition, we identified four highly plausible candidate genes likely to modulate APP metabolism based on biological evidence (*ICA1L, ICA1, DOC2A* and *WDR81)*.

These pathway enrichment analyses also confirmed the involvement of innate immunity and microglial activation in ADD (Supplementary Table 15). In addition, single-cell expression enrichment analysis also highlighted genes expressed in microglia (Supplementary Tables 16 and 17). Finally, ten of our prioritized genes appeared to be primarily expressed in microglia (Fig. 2 and Supplementary Table 34). Several publications have already demonstrated involvement of the corresponding proteins in microglia function/activation (see Supplementary Information) and importantly three, i.e. *GRN, SIGLEC11* and *LILRB2* have also been linked to Aβ peptides/amyloid plaques^13,16,17^. However, it is also important to keep in mind that six of our prioritized genes are expressed at only a very low level in microglia (Supplementary Table 34), emphasizing that ADD results from a complex crosstalk between different cell-types in the brain^18,19^. Of note, we report here the implication of the EGFR but also TNF-alpha signaling through the LUBAC complex (three out of its four complements are coded by ADD genetic risk factors, i.e. *OTULIN, SHARPIN*, and *RBCK1*) reinforcing the role of neuroinflammation in ADD^20^.

Of note, several novel loci were also associated with the risk of developing FTLD, i.e. *MAPT, GRN*, and *TMEM106B*. This may be related to the misclassification in the diagnosis of clinical AD and the proxy-ADD diagnoses in the UKBB. However, both *GRN* and *TMEM106B* are associated with AD neuropathological features and *MAPT* encodes tau, the protein in AD neurofibrillary tangles. Our results may thus also emphasize a continuum between AD and FTD20–22.

Increasing our understanding of the pathophysiological processes operating in AD will lead to better and more specific therapies. Doubling the number of genetic loci associated with ADD risk also gave us the opportunity to create a novel PRS. There are differing opinions regarding the best strategy to define PRS and the model used to evaluate prediction^24–28^. In our approach, we found an association of the PRS with the risk of incident dementia in a healthy population, as well as with progression from MCI to dementia. These results suggest that ADD susceptibility variants, in general, have a similar effect on progression at different clinical stages (independent of age and *APOE*). We also observed that inclusion of confirmed low frequency and rare variants could improve the power of our PRS (we compare HR between PRSs including different MAF thresholds in Supplementary Table 33).

In conclusion, our work doubles the number of ADD associated loci, expanding current knowledge of AD pathophysiological processes, presenting novel opportunities for the development of gene-specific treatments and PRS, further clarifying a path to translational genomics and personalized medicine.

## METHODS Online

### Samples

All discovery meta-analysis samples are from the following consortia/datasets: EADB, GR@ACE, EADI, GERAD/PERADES, DemGene, Bonn, the Rotterdam study, the CCHS study, NxC and the UK Biobank. Of note, in the UK Biobank, individual who did not report dementia or any family history of dementia were used as controls: analysis included 2,447 diagnosed cases, 46,828 proxy cases of dementia and 338,440 controls. Summary demographics of these case-control studies are described in Supplementary Table 1 and more detailed descriptions are available in the Supplementary Information. Replication samples are from the ADGC, CHARGE and FinnGen consortia (Supplementary Table 1 and Supplementary Information) and fully described elsewhere^5,6,9,10,29–31^. Written informed consent was obtained from study participants or, for those with substantial cognitive impairment, from a caregiver, legal guardian, or other proxy. Study protocols for all cohorts were reviewed and approved by the appropriate institutional review boards.

### Quality control and imputation

Standard quality control was performed on variants and samples on all datasets individually. The samples were then imputed with the Trans-Omics for Precision Medicine (TOPMed) reference panel^32,33^. The Haplotype Reference Consortium (HRC) panel^34^ was also used for some datasets (Supplementary Table 2). For the UK Biobank, we used the provided imputed data, generated from a combination of the 1000 Genomes (1000G), HRC and UK10K reference panels. See Supplementary Information for details.

### Stage I analyses

Association tests between ADD clinical or proxy status and autosomal genetic variant were conducted separately in each dataset using logistic regression assuming an additive genetic model as implemented in SNPTEST^35^ or in PLINK^3^, except in the UK Biobank where a logistic mixed model as implemented in SAIGE^36^ was considered. Analyses were performed on the genotype probabilities in SNPTEST (newml method) and on dosages in PLINK and SAIGE. Analyses were adjusted for principal components and genotyping centers when necessary (Supplementary Table 2). For the UK Biobank dataset, effect sizes and standard errors were corrected by a factor of two to take into account that proxy cases were analysed^7^. We filtered out duplicated variants and variants with (i) missing effect size, standard error or P value, (ii) absolute value of effect size above 5, (iii) imputation quality less than 0.3, (iv) the product of the minor allele count (MAC) and the imputation quality (mac-info score) less than 20. In the UK Biobank dataset, only variants with minor allele frequency (MAF) above 0.01% and MAC above 3 were analyzed. For datasets not imputed with the TOPMed reference panel, we also excluded (i) variants for which conversion of position or alleles from the GRCh37 assembly to the GRCh38 assembly was not possible or problematic, or (ii) variants with very large difference of frequency between the TOPMed reference panel and the reference panels used to perform imputation. Results were then combined across studies with a fixed-effect meta-analysis using the inverse variance weighted approach as implemented in the METAL software^37^. We filtered (i) variants with heterogeneity P value below 5×10^−8^, (ii) variants analyzed in less than 20% of the total number of cases and (iii) variants with frequency amplitude above 0.4 (defined as the difference between the maximum and minimum frequency across studies). We further excluded variants analyzed in the UK Biobank only or variants not analyzed in the EADB-TOPMed dataset.

Genomic inflation factor lambda was computed with the R package GenABEL^38^ using the median approach after exclusion of the *APOE* region (44 Mb to 46 Mb on chromosome 19 in GRCh38). The linkage disequilibrium (LD) score (LDSC) regression intercept was computed with the LDSC software using the “baselineLD” LD scores built from 1000 Genomes Phase 3 ^39^. The analysis was restricted to HapMap 3 variants and excluded multi-allelic variants, variants without an rsID and variants in the *APOE* region.

### Definition of associated loci

A region of +/-500kb was defined around each variant with a Stage I P value below 1×10^−5^. Those regions were then merged with the bedtools software to define non-overlapping regions. The region corresponding to the APOE locus was excluded. We then applied the PLINK clumping procedure to define independent hits in each of those regions. The clumping procedure was applied on all variants with a Stage I P value below 1×10^−5^. It is an iterative process beginning with the variant with the lowest P value, named index variant. Variants with a Stage I P value below 1×10^−5^, located within 500 kb of this index variant, and in LD with the index variant (r^2^ above 0.001) are assigned to the clump of this index variant. The clumping procedure is then applied on all the remaining variants, until no variant is left. LD was computed in the EADB-TOPMed dataset using high quality (probability ≥ 0.8) imputed genotypes.

### Stage II analyses

Variants with a Stage I P value below 1×10^−5^ were followed up (see Supplementary Information). A fixed-effect meta-analysis was performed with METAL (inverse variance weighted approach) to combine the results across Stages I and II. In each clump, we then reported the replicated variant (same direction of effects between Stage I and Stage II, with a Stage II P value below 0.05) with the lowest P value in the meta-analysis. Those variants were considered associated at the genome-wide significance level if they had a P value below 5×10^−8^ in the Stages I + II meta-analysis. Among them, we excluded the variant chr6:32657066:G:A because its frequency amplitude was large.

### Pathway analysis

The assignment of Gene Ontology (GO) terms to human genes was obtained from the “gene2go” file, downloaded from NCBI on March 11^th^ 2020. “Parent” GO terms were assigned to genes using the ontology file also downloaded on the same date. GO terms were assigned to genes based on experimental or curated evidence of a specific type, so evidence codes IEA (electronic annotation), NAS (non-traceable author statement), RCA (inferred from reviewed computational analysis) were excluded. Pathways were downloaded from the Reactome website on April 26^th^ 2020. Biocarta, KEGG and Pathway Interaction Database (PID) pathways were downloaded from v7.1 (March 2020) of the Molecular Signatures Database. Analysis was restricted to GO terms containing between 10 and 2000 genes. No size restrictions were placed on the other gene sets, since there were fewer of them. This resulted in a total of 10,271 gene sets for analysis. Gene set enrichment analyses were performed in MAGMA^40^, correcting for the number of variants in each gene, linkage disequilibrium (LD) between variants and LD between genes. LD was computed in the EADB-TOPMed dataset using high quality (probability ≥ 0.9) imputed genotypes. The measure of pathway enrichment is the MAGMA “competitive” test (where the association statistic for genes in the pathway is compared to those of all other protein-coding genes), as recommended by De Leeuw et al.^41^. We used the “mean” test statistic, which uses the sum of -log(variant P value) across all genes. The primary analysis assigned variants to genes if they lie within the gene boundaries, but a secondary analysis used a window of 35kb upstream and 10kb downstream to assign variants to genes, as in Kunkle et al^5^. The primary analysis used all variants with imputation quality above 0.8. We used q-values^42^ to account for multiple testing. Enrichment analyses were performed on single cell expression data from human (Allen Brain Atlas) or mouse (Skene et al^43^) by using average gene expression per nucleus or percentage of nuclei in a cell type expressing each gene as quantitative covariates in a MAGMA gene property analysis.

### QTLs/TWAS/MetaMeth

In order to prioritize candidate genes in the new loci, we employed several approaches: (i) expression quantitative trait loci (eQTLs) and colocalization (eQTL coloc) analyses combined with expression transcriptome-wide association studies (eTWAS) in AD-relevant brain regions; (ii) splicing quantitative trait loci (sQTL) and colocalization (sQTL coloc) analyses combined with splicing transcriptome-wide association studies (sTWAS) in AD-relevant brain regions; (iii) genetic-driven methylation as a biological mediator of genetic signals in blood (MetaMeth). In our regions of interest, we systematically searched if a gene has a significant e/sQTL, colocalization e/sTWAS and/or MetaMeth signal(s) within a region of 1 Mb around the lead variant. In addition to the “nearest” genes from the lead variant, we kept for further analyses those exhibiting such AD-related modulations. We then added several additional approaches: (i) data from a genome-wide, high-content siRNA screening approach to assess the functional impact of gene under-expression on APP metabolism^44^, (ii) methylation QTL (mQTL) and histone acetylation QTL (haQTL) effects of the lead variants in DLPFC^45^, (iii) eQTL effects of the lead variants in monocytes and macrophages^46–51^, (iv) e/sQTL effects of lead variants in LCL along with e/sQTL coloc and e/sTWAS analyses in LCL, and (v) e/sQTL effects of lead variants in microglia along with e/sQTL coloc analyses^52^. A full description of how the genes were prioritized is reported in the Supplementary Information (see also Supplementary Tables 20-30 and Supplementary Fig. 34-42).

### Cell type expression

Assignment of newly identified ADD risk genes to specific cell classes of the adult brain was performed as previously described^53^. Briefly, middle temporal gyrus (MTG) single-nucleus transcriptomes (15,928 total nuclei derived from 8 human tissue donors ranging in age from 24-66 year), were used to annotate and select 6 main cell classes using S*eurat 3*.*1*.*1*^54^: Glutamatergic Neurons, GABAergic Neurons, Astrocytes, Oligodendrocytes, Microglia and Endothelial cells.

### PRS analysis

Eight longitudinal MCI cohorts and six population-based studies were included in the analysis and are fully described in the Supplementary Information and Supplementary Table 32.

PRS were calculated as previously described^24^. Briefly, we considered variants with genome-wide significant evidence of association with ADD in our study and used three different MAF thresholds (Fig. 1, Table 1 and Supplementary Table 33). Variants were directly genotyped or imputed (R^2^ ≥ 0.3). We did not include any *APOE* variants in the PRS. The PRS (based on dosage) was calculated as the weighted average of the number of risk increasing alleles for each variant using dosages. Weights were based on the respective log(OR) obtained in the Stage II. The PRS was then multiplied by the number of included variants. Thus, an increase in HR corresponds to carry one additional average risk allele.

All principal components (PCs) used were generated per cohort, using the same variants that were used on the case/control study PCA. The number of *APOE*-e4 alleles was obtained based on direct genotyping or, if missing, based on genotypes (with probability > 0.8) derived from the TOPMed imputations.

The association of the PRS with risk of progression to dementia in cognitively normal persons or in patients with MCI was assessed using Cox proportional hazards models, initially for all-cause dementia and next, using analysis focused on conversion to AD. Herein, converter to non-AD dementias were coded as censored.

Each Cox-regression analysis was first performed unadjusted for covariates and then repeated, adjusted for age, sex and the first four PCs to correct for potential population stratification. In the 3C study, analysis was adjusted on age, sex, the two first PCs and center. Analyses were additionally controlled for the number of *APOE*-e4 and *APOE*-e2 alleles (assuming an additive effect) to assess the independence of the PRS from *APOE*. The interaction between the PRS and the number of *APOE*-e4 alleles was tested. Analyses were repeated stratified by the presence or absence of at least one *APOE*-e4 allele. Results from individual cohorts were meta-analyzed using fixed effects meta-analysis.

Imputations in Rotterdam were performed using the HRC imputation panel and 6 variants were missing. This study was therefore not included in the meta-analysis.

### URLs

Bedtools: https://bedtools.readthedocs.io/en/latest/

BCFtools: http://samtools.github.io/bcftools/bcftools.html

Samtools: http://www.htslib.org/doc/samtools.html

gene2go: ftp://ftp.ncbi.nlm.nih.gov/gene/DATA/

Gene Ontology: http://geneontology.org/docs/download-ontology/

Reactome: https://reactome.org/download-data

KEGG and Pathway Interaction Database (PID) pathways: https://www.gsea-msigdb.org/gsea/msigdb/index.jsp

AMP-AD rnaSeqReprocessing Study: https://www.synapse.org/#!Synapse:syn9702085

MayoRNAseq WGS VCFs: https://www.synapse.org/#!Synapse:syn11724002

ROSMAP WGS VCFs: https://www.synapse.org/#!Synapse:syn11724057

MSBB WGS VCFs: https://www.synapse.org/#!Synapse:syn11723899

GTEx pipeline: https://github.com/broadinstitute/gtex-pipeline

Leafcutter: https://github.com/davidaknowles/leafcutter

RegTools: https://github.com/griffithlab/regtools

Enhanced version of FastQTL: https://github.com/francois-a/fastqtl

Picard: https://broadinstitute.github.io/picard/

eQTLGen: https://www.eqtlgen.org/

eQTL Catalogue database: https://www.ebi.ac.uk/eqtl/

Brain xQTL serve: http://mostafavilab.stat.ubc.ca/xqtl/

GTEx v8 eQTL and sQTL catalogues: https://www.gtexportal.org/

coloc: https://github.com/chr1swallace/coloc

FUSION: https://github.com/gusevlab/fusion_twas

GTEx v8 expression and splicing prediction models: http://predictdb.org/

MetaXcan: https://github.com/hakyimlab/MetaXcan

FOCUS: https://github.com/bogdanlab/focus

qcat: https://github.com/nanoporetech/qcat

minimap2: https://github.com/lh3/minimap2

NanoStat: https://github.com/wdecoster/nanostat

mosdepth: https://github.com/brentp/mosdepth

ggplot2: https://ggplot2.tidyverse.org/

LocusZoom: https://github.com/statgen/locuszoom-standalone

pyGenomeTracks: https://github.com/deeptools/pyGenomeTracks

BECon website: https://redgar598.shinyapps.io/BECon/

VCFs of phased biallelic SNV and INDEL variants of 1KG samples (de novo called on GRCh38):

ftp://ftp.1000genomes.ebi.ac.uk/vol1/ftp/data_collections/1000_genomes_project/release/20190312_biallelic_SNV_and_INDEL/

MiGA eQTLs: https://doi.org/10.5281/zenodo.4118605

MiGA sQTLs: https://doi.org/10.5281/zenodo.4118403

MiGA Meta-analysis: https://doi.org/10.5281/zenodo.4118676

## Data Availability

summary statistics will be made available followoing publication of the article

## EADB

We thank the numerous participants, researchers, and staff from many studies who collected and contributed to the data. We thank the high performance computing service of the University of Lille. We thank all the CEA-CNRGH staff who contributed to sample preparation and genotyping for their excellent technical assistance. This research has been conducted using the UK Biobank Resource *under Application Number* 61054.

This work was supported by a grant (European Alzheimer DNA BioBank, EADB) from the EU Joint Programme – Neurodegenerative Disease Research (JPND). Inserm UMR1167 is also funded by Inserm, Institut Pasteur de Lille, the Lille Métropole Communauté Urbaine, the French government’s LABEX DISTALZ program (development of innovative strategies for a transdisciplinary approach to Alzheimer’s disease).

## Additional support for EADB cohorts was provided by

The work for this manuscript was further supported by the CoSTREAM project (www.costream.eu) and funding from the European Union’s Horizon 2020 research and innovation programme under grant agreement No 667375. Italian Ministry of Health (Ricerca Corrente); Ministero dell’Istruzione, dell’Università e della Ricerca–MIUR project “Dipartimenti di Eccellenza 2018–2022” to Department of Neuroscience “Rita Levi Montalcini”, University of Torino (IR), and AIRAlzh Onlus-ANCC-COOP (SB); Partly supported by “Ministero della Salute”, I.R.C.C.S. Research Program, Ricerca Corrente 2018-2020, Linea n. 2 “Meccanismi genetici, predizione e terapie innovative delle malattie complesse” and by the “5 x 1000” voluntary contribution to the Fondazione I.R.C.C.S. Ospedale “Casa Sollievo della Sofferenza”; and RF-2018-12366665, Fondi per la ricerca 2019 (Sandro Sorbi). Copenhagen General Population Study (CGPS): We thank staff and participants of the CGPS for their important contributions. Karolinska Institutet AD cohort: Dr. Graff and co-authors of the Karolinska Institutet AD cohort report grants from Swedish Research Council (VR) 2015-02926, 2018-02754, 2015-06799, Swedish Alzheimer Foundation, Stockholm County Council ALF and resarch school, Karolinska Institutet StratNeuro, Swedish Demensfonden, and Swedish brain foundation, during the conduct of the study. ADGEN: This work was supported by Academy of Finland (grant numbers 307866); Sigrid Jusélius Foundation; the Strategic Neuroscience Funding of the University of Eastern Finland; EADB project in the JPNDCO-FUND program (grant number 301220). CBAS: Supported by the project no. LQ1605 from the National Program of Sustainability II (MEYS CR), Supported by Ministry of Health of the Czech Republic, grant nr. NV19-04-00270 (All rights reserved), Grant Agency of Charles University Grants No. 693018 and 654217; the Ministry of Health, Czech Republic-conceptual development of research organization, University Hospital Motol, Prague, Czech Republic Grant No. 00064203; the Czech Ministry of Health Project AZV Grant No. 16-27611A; and Institutional Support of Excellence 2. LF UK Grant No. 699012. CNRMAJ-Rouen: This study received fundings from the Centre National de Référence Malades Alzheimer Jeunes (CNRMAJ). The Finnish Geriatric Intervention Study for the Prevention of Cognitive Impairment and Disability (FINGER) data collection was supported by grants from the Academy of Finland, La Carita Foundation, Juho Vainio Foundation, Novo Nordisk Foundation, Finnish Social Insurance Institution, Ministry of Education and Culture Research Grants, Yrjö Jahnsson Foundation, Finnish Cultural Foundation South Osthrobothnia Regional Fund, and EVO/State Research Funding grants of University Hospitals of Kuopio, Oulu and Turku, Seinäjoki Central Hospital and Oulu City Hospital, Alzheimer’s Research & Prevention Foundation USA, AXA Research Fund, Knut and Alice Wallenberg Foundation Sweden, Center for Innovative Medicine (CIMED) at Karolinska Institutet Sweden, and Stiftelsen Stockholms sjukhem Sweden. FINGER cohort genotyping was funded by EADB project in the JPND CO-FUND (grant number 301220). Research at the Belgian EADB site is funded in part by the Alzheimer Research Foundation (SAO-FRA), The Research Foundation Flanders (FWO), and the University of Antwerp Research Fund. FK is supported by a BOF DOCPRO fellowship of the University of Antwerp Research Fund. SNAC-K is financially supported by the Swedish Ministry of Health and Social Affairs, the participating County Councils and Municipalities, and the Swedish Research Council. BDR Bristol: We would like to thank the South West Dementia Brain Bank (SWDBB) for providing brain tissue for this study. The SWDBB is part of the Brains for Dementia Research programme, jointly funded by Alzheimer’s Research UK and Alzheimer’s Society and is supported by BRACE (Bristol Research into Alzheimer’s and Care of the Elderly) and the Medical Research Council. BDR Manchester: We would like to thank the Manchester Brain Bankfor providing brain tissue for this study. The Manchester Brain Bank is part of the Brains for Dementia Research programme, jointly funded by Alzheimer’s Research UK and Alzheimer’s Society. BDR KCL: Human post-mortem tissue was provided by the London Neurodegenerative Diseases Brain Bank which receives funding from the UK Medical Research Council and as part of the Brains for Dementia Research programme, jointly funded by Alzheimer’s Research UK and the Alzheimer’s Society. The CFAS Wales study was funded by the ESRC (RES-060-25-0060) and HEFCW as ‘Maintaining function and well-being in later life: a longitudinal cohort study’, (Principal Investigators: R.T Woods, L.Clare, G.Windle, V. Burholt, J. Philips, C. Brayne, C. McCracken, K. Bennett, F. Matthews). We are grateful to the NISCHR Clinical Research Centre for their assistance in tracing participants and in interviewing and in collecting blood samples, and to general practices in the study areas for their cooperation. MRC: We thank all individuals who participated in this study. Cardiff University was supported by the Alzheimer’s Society (AS; grant RF014/164) and the Medical Research Council (MRC; grants G0801418/1, MR/K013041/1, MR/L023784/1) (R. Sims is an AS Research Fellow). Cardiff University was also supported by the European Joint Programme for Neurodegenerative Disease (JPND; grant MR/L501517/1), Alzheimer’s Research UK (ARUK; grant ARUK-PG2014-1), the Welsh Assembly Government (grant SGR544:CADR), Brain’s for dementia Research and a donation from the Moondance Charitable Foundation. Cardiff University acknowledges the support of the UK Dementia Research Institute, of which J. Williams is an associate director. Cambridge University acknowledges support from the MRC. Patient recruitment for the MRC Prion Unit/UCL Department of Neurodegenerative Disease collection was supported by the UCLH/UCL Biomedical Centre and NIHR Queen Square Dementia Biomedical Research Unit. The University of Southampton acknowledges support from the AS. King’s College London was supported by the NIHR Biomedical Research Centre for Mental Health and the Biomedical Research Unit for Dementia at the South London and Maudsley NHS Foundation Trust and by King’s College London and the MRC. ARUK and the Big Lottery Fund provided support to Nottingham University. Alfredo Ramirez: Part of the work was funded by the JPND EADB grant (German Federal Ministry of Education and Research (BMBF) grant: 01ED1619A). German Study on Ageing, Cognition and Dementia in Primary Care Patients (AgeCoDe): This study/publication is part of the German Research Network on Dementia (KND), the German Research Network on Degenerative Dementia (KNDD; German Study on Ageing, Cognition and Dementia in Primary Care Patients; AgeCoDe), and the Health Service Research Initiative (Study on Needs, health service use, costs and health-related quality of life in a large sample of oldestold primary care patients (85+; AgeQualiDe)) and was funded by the German Federal Ministry of Education and Research (grants KND: 01GI0102, 01GI0420, 01GI0422, 01GI0423, 01GI0429, 01GI0431, 01GI0433, 01GI0434; grants KNDD: 01GI0710, 01GI0711, 01GI0712, 01GI0713, 01GI0714, 01GI0715, 01GI0716; grants Health Service Research Initiative: 01GY1322A, 01GY1322B, 01GY1322C, 01GY1322D, 01GY1322E, 01GY1322F, 01GY1322G). VITA study: The support of the Ludwig Boltzmann Society and the AFI Germany have supported the VITA study. The former VITA study group should be acknowledged: W. Danielczyk, G. Gatterer, K Jellinger, S Jugwirth, KH Tragl, S Zehetmayer. Vogel Study: This work was financed by a research grant of the ‘‘Vogelstiftung Dr. Eckernkamp’’. HELIAD study: This study was supported by the grants: IIRG-09-133014 from the Alzheimer’s Association, 189 10276/8/9/2011 from the ESPA-EU program Excellence Grant (ARISTEIA) and the ΔΥ2β/οικ.51657/14.4.2009 of the Ministry for Health and Social Solidarity (Greece). Biobank Department of Psychiatry, UMG: Prof. Jens Wiltfang is supported by an Ilídio Pinho professorship and iBiMED (UID/BIM/04501/2013), and FCT project PTDC/DTP_PIC/5587/2014 at the University of Aveiro, Portugal. Lausanne study: This work was supported by grants from the Swiss National Research Foundation (SNF 320030_141179). PAGES study: Harald Hampel is an employee of Eisai Inc. During part of this work he was supported by the AXA Research Fund, the “Fondation partenariale Sorbonne Université” and the “Fondation pour la Recherche sur Alzheimer”, Paris, France. Mannheim, Germany Biobank: Department of geriatric Psychiatry, Central Institute for Mental Health, Mannheim, University of Heidelberg, Germany. Genotyping for the Swedish Twin Studies of Aging was supported by NIH/NIA grant R01 AG037985. Genotyping in TwinGene was supported by NIH/NIDDK U01 DK066134. WvdF is recipient of Joint Programming for Neurodegenerative Diseases (JPND) grants PERADES (ANR-13-JPRF-0001) and EADB (733051061). Gothenburg Birth Cohort (GBC) Studies: We would like to thank UCL Genomics for performing the genotyping analyses. The studies were supported by The Stena Foundation, The Swedish Research Council (2015-02830, 2013-8717), The Swedish Research Council for Health, Working Life and Wellfare (2013-1202, 2005-0762, 2008-1210, 2013-2300, 2013-2496, 2013-0475), The Brain Foundation, Sahlgrenska University Hospital (ALF), The Alzheimer’s Association (IIRG-03-6168), The Alzheimer’s Association Zenith Award (ZEN-01-3151), Eivind och Elsa K:son Sylvans Stiftelse, The Swedish Alzheimer Foundation. Clinical AD, Sweden: We would like to thank UCL Genomics for performing the genotyping analyses. Barcelona Brain Biobank: Brain Donors of the Neurological Tissue Bank of the Biobanc-Hospital Clinic-IDIBAPS and their families for their generosity. Hospital Clínic de Barcelona Spanish Ministry of Economy and Competitiveness-Instituto de Salud Carlos III and Fondo Europeo de Desarrollo Regional (FEDER), Unión Europea, “Una manera de hacer Europa” grants (PI16/0235 to Dr. R. Sánchez-Valle and PI17/00670 to Dr. A.Antonelli). AA is funded by Departament de Salut de la Generalitat de Catalunya, PERIS 2016-2020 (SLT002/16/00329). Work at JP-T laboratory was possible thanks to funding from Ciberned and generous gifts from Consuelo Cervera Yuste and Juan Manuel Moreno Cervera. Sydney Memory and Ageing Study (Sydney MAS): We gratefully acknowledge and thank the following for their contributions to Sydney MAS: participants, their supporters and the Sydney MAS Research Team (current and former staff and students). Funding was awarded from the Australian National Health and Medical Research Council (NHMRC) Program Grants (350833, 568969, 109308). AddNeuroMed consortium was led by Simon Lovestone, Bruno Vellas, Patrizia Mecocci, Magda Tsolaki, Iwona Kłoszewska, Hilkka Soininen. This work was supported by InnoMed (Innovative Medicines in Europe), an integrated project funded by the European Union of the Sixth Framework program priority (FP6-2004-LIFESCIHEALTH-5). Oviedo: This work was partly supported by Grant from Fondo de Investigaciones Sanitarias-Fondos FEDER EuropeanUnion to Victoria Alvarez PI15/00878. Pascual Sánchez-Juan is supported by CIBERNED and Carlos III Institute of Health, Spain (PI08/0139, PI12/02288, and PI16/01652), jointly funded by Fondo Europeo de Desarrollo Regional (FEDER), Unión Europea, “Una manera de hacer Europa”. Project MinE: The ProjectMinE study was supported by the ALS Foundation Netherlands and the MND association (UK) (Project MinE, www.projectmine.com). The SPIN cohort: We are indebted to patients and their families for their participation in the “Sant Pau Initiative on Neurodegeneration cohort”, at the Sant Pau Hospital (Barcelona). This is a multimodal research cohort for biomarker discovery and validation that is partially funded by Generalitat de Catalunya (2017 SGR 547 to JC), as well as from the Institute of Health Carlos III-Subdirección General de Evaluación and the Fondo Europeo de Desarrollo Regional (FEDER-“Una manera de Hacer Europa”) (grants PI11/02526, PI14/01126, and PI17/01019 to JF; PI17/01895 to AL), and the Centro de Investigación Biomédica en Red Enfermedades Neurodegenerativas programme (Program 1, Alzheimer Disease to AL). We would also like to thank the Fundació Bancària Obra Social La Caixa (DABNI project) to JF and AL; and Fundación BBVA (to AL), for their support in funding this follow-up study. Adolfo López de Munain is supported by Fundación Salud 2000 (PI2013156), CIBERNED and Diputación Foral de Gipuzkoa (Exp.114/17).

### Gra@ce

The Genome Research @ Fundació ACE project (GR@ACE) is supported by Grifols SA, Fundación bancaria ‘La Caixa’, Fundació ACE, and CIBERNED (Centro de Investigación Biomédica en Red Enfermedades Neurodegenerativas (Program 1, Alzheimer Disease to MB and AR)). A.R. and M.B. receive support from the European Union/EFPIA Innovative Medicines Initiative Joint undertaking ADAPTED and MOPEAD projects (grant numbers 115975 and 115985, respectively). M.B. and A.R. are also supported by national grants PI13/02434, PI16/01861, PI17/01474 and PI19/01240. Acción Estratégica en Salud is integrated into the Spanish National R + D + I Plan and funded by ISCIII (Instituto de Salud Carlos III)–Subdirección General de Evaluación and the Fondo Europeo de Desarrollo Regional (FEDER–’Una manera de hacer Europa’). Some control samples and data from patients included in this study were provided in part by the National DNA Bank Carlos III (www.bancoadn.org, University of Salamanca, Spain) and Hospital Universitario Virgen de Valme (Sevilla, Spain); they were processed following standard operating procedures with the appropriate approval of the Ethical and Scientific Committee. The present work has been performed as part of the doctoral program of I. de Rojas at the Universitat de Barcelona (Barcelona, Spain).

### EADI

This work has been developed and supported by the LABEX (laboratory of excellence program investment for the future) DISTALZ grant (Development of Innovative Strategies for a Transdisciplinary approach to ALZheimer’s disease) including funding from MEL (Metropole européenne de Lille), ERDF (European Regional Development Fund) and Conseil Régional Nord Pas de Calais. This work was supported by INSERM, the National Foundation for Alzheimer’s disease and related disorders, the Institut Pasteur de Lille and the Centre National de Recherche en Génomique Humaine, CEA, the JPND PERADES, the Laboratory of Excellence GENMED (Medical Genomics) grant no. ANR-10-LABX-0013 managed by the National Research Agency (ANR) part of the Investment for the Future program, and the FP7 AgedBrainSysBio. The Three-City Study was performed as part of collaboration between the Institut National de la Santé et de la Recherche Médicale (Inserm), the Victor Segalen Bordeaux II University and Sanofi-Synthélabo. The Fondation pour la Recherche Médicale funded the preparation and initiation of the study. The 3C Study was also funded by the Caisse Nationale Maladie des Travailleurs Salariés, Direction Générale de la Santé, MGEN, Institut de la Longévité, Agence Française de Sécurité Sanitaire des Produits de Santé, the Aquitaine and Bourgogne Regional Councils, Agence Nationale de la Recherche, ANR supported the COGINUT and COVADIS projects. Fondation de France and the joint French Ministry of Research/INSERM “Cohortes et collections de données biologiques” programme. Lille Génopôle received an unconditional grant from Eisai. The Three-city biological bank was developed and maintained by the laboratory for genomic analysis LAG-BRC -Institut Pasteur de Lille.

### GERAD/PERADES

We thank all individuals who participated in this study. Cardiff University was supported by the Wellcome Trust, Alzheimer’s Society (AS; grant RF014/164), the Medical Research Council (MRC; grants G0801418/1, MR/K013041/1, MR/L023784/1), the European Joint Programme for Neurodegenerative Disease (JPND, grant MR/L501517/1), Alzheimer’s Research UK (ARUK, grant ARUK-PG2014-1), Welsh Assembly Government (grant SGR544:CADR), a donation from the Moondance Charitable Foundation, UK Dementia’s Platform (DPUK, reference MR/L023784/1), and the UK Dementia Research Institute at Cardiff. Cambridge University acknowledges support from the MRC. ARUK supported sample collections at the Kings College London, the South West Dementia Bank, Universities of Cambridge, Nottingham, Manchester and Belfast. King’s College London was supported by the NIHR Biomedical Research Centre for Mental Health and Biomedical Research Unit for Dementia at the South London and Maudsley NHS Foundation Trust and Kings College London and the MRC. Alzheimer’s Research UK (ARUK) and the Big Lottery Fund provided support to Nottingham University. Ulster Garden Villages, AS, ARUK, American Federation for Aging Research, NI R&D Office and the Royal College of Physicians/Dunhill Medical Trust provided support for Queen’s University, Belfast. The University of Southampton acknowledges support from the AS. The MRC and Mercer’s Institute for Research on Ageing supported the Trinity College group. DCR is a Wellcome Trust Principal Research fellow. The South West Dementia Brain Bank acknowledges support from Bristol Research into Alzheimer’s and Care of the Elderly. The Charles Wolfson Charitable Trust supported the OPTIMA group. Washington University was funded by NIH grants, Barnes Jewish Foundation and the Charles and Joanne Knight Alzheimer’s Research Initiative. Patient recruitment for the MRC Prion Unit/UCL Department of Neurodegenerative Disease collection was supported by the UCLH/UCL Biomedical Research Centre and their work was supported by the NIHR Queen Square Dementia BRU, the Alzheimer’s Research UK and the Alzheimer’s Society. LASER-AD was funded by Lundbeck SA. The AgeCoDe study group was supported by the German Federal Ministry for Education and Research grants 01 GI 0710, 01 GI 0712, 01 GI 0713, 01 GI 0714, 01 GI 0715, 01 GI 0716, 01 GI 0717. Genotyping of the Bonn case-control sample was funded by the German centre for Neurodegenerative Diseases (DZNE), Germany. The GERAD Consortium also used samples ascertained by the NIMH AD Genetics Initiative. HH was supported by a grant of the Katharina-Hardt-Foundation, Bad Homburg vor der Höhe, Germany. The KORA F4 studies were financed by Helmholtz Zentrum München; German Research Center for Environmental Health; BMBF; German National Genome Research Network and the Munich Center of Health Sciences. The Heinz Nixdorf Recall cohort was funded by the Heinz Nixdorf Foundation (Dr. Jur. G.Schmidt, Chairman) and BMBF. We acknowledge use of genotype data from the 1958 Birth Cohort collection and National Blood Service, funded by the MRC and the Wellcome Trust which was genotyped by the Wellcome Trust Case Control Consortium and the Type-1 Diabetes Genetics Consortium, sponsored by the National Institute of Diabetes and Digestive and Kidney Diseases, National Institute of Allergy and Infectious Diseases, National Human Genome Research Institute, National Institute of Child Health and Human Development and Juvenile Diabetes Research Foundation International. The project is also supported through the following funding organisations under the aegis of JPND -www.jpnd.eu (United Kingdom, Medical Research Council (MR/L501529/1; MR/R024804/1) and Economic and Social Research Council (ES/L008238/1)) and through the Motor Neurone Disease Association. This study represents independent research part funded by the National Institute for Health Research (NIHR) Biomedical Research Centre at South London and Maudsley NHS Foundation Trust and King’s College London. Prof Jens Wiltfang is supported by an Ilídio Pinho professorship and iBiMED (UID/BIM/04501/2013), at the University of Aveiro, Portugal.

### Rotterdam study

Rotterdam (RS). This study was funded by the Netherlands Organisation for Health Research and Development (ZonMW) as part of the Joint Programming for Neurological Disease (JPND)as part of the PERADES Program (Defining Genetic Polygenic, and Environmental Risk for Alzheimer’s disease using multiple powerful cohorts, focused Epigenetics and Stem cell metabolomics), Project number 733051021. This work was funded also by the European Union Innovative Medicine Initiative (IMI) programme under grant agreement No. 115975 as part of the Alzheimer’s Disease Apolipoprotein Pathology for Treatment Elucidation and Development (ADAPTED, https://www.imi-adapted.eu);and the European Union’s Horizon 2020 research and innovation programme as part of the Common mechanisms and pathways in Stroke and Alzheimer’s disease CoSTREAM project (www.costream.eu, grant agreement No. 667375). The current study is supported by the Deltaplan Dementie and Memorabel supported by ZonMW (Project number 733050814) and Alzheimer Nederland. The Rotterdam Study is funded by Erasmus Medical Center and Erasmus University, Rotterdam, Netherlands Organization for the Health Research and Development (ZonMw), the Research Institute for Diseases in the Elderly (RIDE), the Ministry of Education, Culture and Science, the Ministry for Health, Welfare and Sports, the European Commission (DG XII), and the Municipality of Rotterdam. The authors are grateful to the study participants, the staff from the Rotterdam Study and the participating general practitioners and pharmacists. The generation and management of GWAS genotype data for the Rotterdam Study (RS-I, RS-II, RS-III) was executed by the Human Genotyping Facility of the Genetic Laboratory of the Department of Internal Medicine, Erasmus MC, Rotterdam, The Netherlands. The GWAS datasets are supported by the Netherlands Organization of Scientific Research NWO Investments (Project number 175.010.2005.011, 911-03-012), the Genetic Laboratory of the Department of Internal Medicine, Erasmus MC, the Research Institute for Diseases in the Elderly (014-93-015; RIDE2), the Netherlands Genomics Initiative (NGI)/Netherlands Organization for Scientific Research (NWO) Netherlands Consortium for Healthy Aging (NCHA), project number 050-060-810. We thank Pascal Arp, Mila Jhamai, Marijn Verkerk, Lizbeth Herrera and Marjolein Peters, MSc, and Carolina Medina-Gomez, MSc, for their help in creating the GWAS database, and Karol Estrada, PhD, Yurii Aulchenko, PhD, and Carolina Medina-Gomez, MSc, for the creation and analysis of imputed data.

### DemGene

The project has received funding from The Research Council of Norway (RCN) Grant Nos. 213837, 223273, 225989, 248778, and 251134 and EU JPND Program ApGeM RCN Grant No. 237250, the South-East Norway Health Authority Grant No. 2013-123, the Norwegian Health Association, and KG Jebsen Foundation. The RCN FRIPRO Mobility grant scheme (FRICON) is co-funded by the European Union’s Seventh Framework Programme for research, technological development and demonstration under Marie Curie grant agreement No 608695. European Community’s grant PIAPP-GA-2011-286213 PsychDPC.

### Bonn study

This group would like to thank Dr. Heike Koelsch for her scientific support. The Bonn group was funded by the German Federal Ministry of Education and Research (BMBF): Competence Network Dementia (CND) grant number 01GI0102, 01GI0711, 01GI042

### ADGC

The National Institutes of Health, National Institute on Aging (NIH-NIA) supported this work through the following grants: ADGC, U01 AG032984, RC2 AG036528; Samples from the National Cell Repository for Alzheimer’s Disease (NCRAD), which receives government support under a cooperative agreement grant (U24 AG21886) awarded by the National Institute on Aging (NIA), were used in this study. We thank contributors who collected samples used in this study, as well as patients and their families, whose help and participation made this work possible; Data for this study were prepared, archived, and distributed by the National Institute on Aging Alzheimer’s Disease Data Storage Site (NIAGADS) at the University of Pennsylvania (U24-AG041689-01); NACC, U01 AG016976; NIA LOAD (Columbia University), U24 AG026395, U24 AG026390, R01AG041797; Banner Sun Health Research Institute P30 AG019610; Boston University, P30 AG013846, U01 AG10483, R01 CA129769, R01 MH080295, R01 AG017173, R01 AG025259, R01 AG048927, R01AG33193, R01 AG009029; Columbia University, P50 AG008702, R37 AG015473, R01 AG037212, R01 AG028786; Duke University, P30 AG028377, AG05128; Einstein Aging Study NIA grant at Albert Einstein College of Medicine, P01 AG03949. Emory University, AG025688; Group Health Research Institute, UO1 AG006781, UO1 HG004610, UO1 HG006375, U01 HG008657; Indiana University, P30 AG10133, R01 AG009956, RC2 AG036650; Johns Hopkins University, P50 AG005146, R01 AG020688; Massachusetts General Hospital, P50 AG005134; Mayo Clinic, P50 AG016574, R01 AG032990, KL2 RR024151; Mount Sinai School of Medicine, P50 AG005138, P01 AG002219; New York University, P30 AG08051, UL1 RR029893, 5R01AG012101, 5R01AG022374, 5R01AG013616, 1RC2AG036502, 1R01AG035137; North Carolina A&T University, P20 MD000546, R01 AG28786-01A1; Northwestern University, P30 AG013854; Oregon Health & Science University, P30 AG008017, R01 AG026916; Rush University, P30 AG010161, R01 AG019085, R01 AG15819, R01 AG17917, R01 AG030146, R01 AG01101, RC2 AG036650, R01 AG22018; TGen, R01 NS059873; University of Alabama at Birmingham, P50 AG016582; University of Arizona, R01 AG031581; University of California, Davis, P30 AG010129; University of California, Irvine, P50 AG016573; University of California, Los Angeles, P50 AG016570; University of California, San Diego, P50 AG005131; University of California, San Francisco, P50 AG023501, P01 AG019724; University of Kentucky, P30 AG028383, AG05144; University of Michigan, P30 AG053760 and AG063760; University of Pennsylvania, P30 AG010124; University of Pittsburgh, P50 AG005133, AG030653, AG041718, AG07562, AG02365; University of Southern California, P50 AG005142; University of Texas Southwestern, P30 AG012300; University of Miami, R01 AG027944, AG010491, AG027944, AG021547, AG019757; University of Washington, P50 AG005136, R01 AG042437; University of Wisconsin, P50 AG033514; Vanderbilt University, R01 AG019085; and Washington University, P50 AG005681, P01 AG03991, P01 AG026276. HP was supported by AG025711. ER was supported by CCNA. The Kathleen Price Bryan Brain Bank at Duke University Medical Center is funded by NINDS grant # NS39764, NIMH MH60451 and by Glaxo Smith Kline. Support was also from the Alzheimer’s Association (LAF, IIRG-08-89720; MP-V, IIRG-05-14147), the US Department of Veterans Affairs Administration, Office of Research and Development, Biomedical Laboratory Research Program, and BrightFocus Foundation (MP-V, A2111048). P.S.G.-H. is supported by Wellcome Trust, Howard Hughes Medical Institute, and the Canadian Institute of Health Research. Genotyping of the TGEN2 cohort was supported by Kronos Science. The TGen series was also funded by NIA grant AG041232 to AJM and MJH, The Banner Alzheimer’s Foundation, The Johnnie B. Byrd Sr. Alzheimer’s Institute, the Medical Research Council, and the state of Arizona and also includes samples from the following sites: Newcastle Brain Tissue Resource (funding via the Medical Research Council, local NHS trusts and Newcastle University), MRC London Brain Bank for Neurodegenerative Diseases (funding via the Medical Research Council),South West Dementia Brain Bank (funding via numerous sources including the Higher Education Funding Council for England (HEFCE), Alzheimer’s Research Trust (ART), BRACE as well as North Bristol NHS Trust Research and Innovation department and DeNDRoN), The Netherlands Brain Bank (funding via numerous sources including Stichting MS Research, Brain Net Europe, Hersenstichting Nederland Breinbrekend Werk, International Parkinson Fonds, Internationale Stiching Alzheimer Onderzoek), Institut de Neuropatologia, Servei Anatomia Patologica, Universitat de Barcelona. ADNI data collection and sharing was funded by the National Institutes of Health Grant U01 AG024904 and Department of Defense award number W81XWH-12-2-0012. ADNI is funded by the National Institute on Aging, the National Institute of Biomedical Imaging and Bioengineering, and through generous contributions from the following: AbbVie, Alzheimer’s Association; Alzheimer’s Drug Discovery Foundation; Araclon Biotech; BioClinica, Inc.; Biogen; Bristol-Myers Squibb Company; CereSpir, Inc.; Eisai Inc.; Elan Pharmaceuticals, Inc.; Eli Lilly and Company; EuroImmun; F. Hoffmann-La Roche Ltd and its affiliated company Genentech, Inc.; Fujirebio; GE Healthcare; IXICO Ltd.; Janssen Alzheimer Immunotherapy Research & Development, LLC.; Johnson & Johnson Pharmaceutical Research & Development LLC.; Lumosity; Lundbeck; Merck & Co., Inc.; Meso Scale Diagnostics, LLC.; NeuroRx Research; Neurotrack Technologies; Novartis Pharmaceuticals Corporation; Pfizer Inc.; Piramal Imaging; Servier; Takeda Pharmaceutical Company; and Transition Therapeutics. The Canadian Institutes of Health Research is providing funds to support ADNI clinical sites in Canada. Private sector contributions are facilitated by the Foundation for the National Institutes of Health (www.fnih.org). The grantee organization is the Northern California Institute for Research and Education, and the study is coordinated by the Alzheimer’s Disease Cooperative Study at the University of California, San Diego. ADNI data are disseminated by the Laboratory for Neuro Imaging at the University of Southern California. We thank Drs. D. Stephen Snyder and Marilyn Miller from NIA who are *ex-officio* ADGC members. FTLD-TDP GWAS: National Institute on Aging (AG101024, AG066597 and AG017586)

### CHARGE

#### Cardiovascular Health Study (CHS)

This CHS research was supported by NHLBI contracts HHSN268201200036C, HHSN268200800007C, HHSN268201800001C, N01HC55222, N01HC85079, N01HC85080, N01HC85081, N01HC85082, N01HC85083, N01HC85086, 75N92021D00006; and NHLBI grants U01HL080295, U01HL130114, R01HL087652, R01HL105756, R01HL103612, R01HL120393 and 75N92021D00006 with additional contribution from the National Institute of Neurological Disorders and Stroke (NINDS). Additional support was provided through R01AG023629, R01AG033193, R01AG15928, R01AG20098, and U01AG049505 from the National Institute on Aging (NIA). A full list of principal CHS investigators and institutions can be found at CHS-NHLBI.org. The provision of genotyping data was supported in part by the National Center for Advancing Translational Sciences, CTSI grant UL1TR001881, and the National Institute of Diabetes and Digestive and Kidney Disease Diabetes Research Center (DRC) grant DK063491 to the Southern California Diabetes Endocrinology Research Center. <u>Framingham Heart Study</u>. This work was supported by the National Heart, Lung, and Blood Institute’s Framingham Heart Study (contracts N01-HC-25195 and HHSN268201500001I). This study was also supported by grants from the National Institute on Aging: R01AG033193, U01AG049505, U01AG52409, R01AG054076, RF1AG0059421 (S. Seshadri). S. Seshadri and A.L.D. were also supported by additional grants from the National Institute on Aging (R01AG049607, R01AG033040, RF1AG0061872, U01AG058589) and the National Institute of Neurological Disorders and Stroke (R01-NS017950, NS100605). The content is solely the responsibility of the authors and does not necessarily represent the official views of the US National Institutes of Health.

### FinnGen

The FinnGen project is funded by two grants from Business Finland (HUS 4685/31/2016 and UH 4386/31/2016) and the following industry partners: AbbVie Inc., AstraZeneca UK Ltd, Biogen MA Inc., Celgene Corporation, Celgene International II Sàrl, Genentech Inc., Merck Sharp & Dohme Corp, Pfizer Inc., GlaxoSmithKline Intellectual Property Development Ltd., Sanofi US Services Inc., Maze Therapeutics Inc., Janssen Biotech Inc, and Novartis AG. Following biobanks are acknowledged for the project samples: Auria Biobank (www.auria.fi/biopankki), THL Biobank (www.thl.fi/biobank), Helsinki Biobank (www.helsinginbiopankki.fi), Biobank Borealis of Northern Finland (https://www.ppshp.fi/Tutkimus-ja-opetus/Biopankki/Pages/Biobank-Borealis-briefly-in-English.aspx), Finnish Clinical Biobank Tampere (www.tays.fi/en-US/Research_and_development/Finnish_Clinical_Biobank_Tampere), Biobank of Eastern Finland (www.ita-suomenbiopankki.fi/en), Central Finland Biobank (www.ksshp.fi/fi-FI/Potilaalle/Biopankki), Finnish Red Cross Blood Service Biobank (www.veripalvelu.fi/verenluovutus/biopankkitoiminta) and Terveystalo Biobank (www.terveystalo.com/fi/Yritystietoa/Terveystalo-Biopankki/Biopankki/). All Finnish Biobanks are members of BBMRI.fi infrastructure (www.bbmri.fi).

### QTLs/TWAS analyses

The results published here are in whole or in part based on data obtained from the AD Knowledge Portal (https://adknowledgeportal.synapse.org/). For MayoRNAseq, the study data were provided by the following sources: The Mayo Clinic Alzheimers Disease Genetic Studies, led by Dr. Nilufer Ertekin-Taner and Dr. Steven G. Younkin, Mayo Clinic, Jacksonville, FL using samples from the Mayo Clinic Study of Aging, the Mayo Clinic Alzheimers Disease Research Center, and the Mayo Clinic Brain Bank. Data collection was supported through funding by NIA grants P50 AG016574, R01 AG032990, U01 AG046139, R01 AG018023, U01 AG006576, U01 AG006786, R01 AG025711, R01 AG017216, R01 AG003949, NINDS grant R01 NS080820, CurePSP Foundation, and support from Mayo Foundation. Study data includes samples collected through the Sun Health Research Institute Brain and Body Donation Program of Sun City, Arizona. The Brain and Body Donation Program is supported by the National Institute of Neurological Disorders and Stroke (U24 NS072026 National Brain and Tissue Resource for Parkinsons Disease and Related Disorders), the National Institute on Aging (P30 AG19610 Arizona Alzheimers Disease Core Center), the Arizona Department of Health Services (contract 211002, Arizona Alzheimers Research Center), the Arizona Biomedical Research Commission (contracts 4001, 0011, 05-901 and 1001 to the Arizona Parkinson’s Disease Consortium) and the Michael J. Fox Foundation for Parkinsons Research. For ROSMAP, the study data were provided by the Rush Alzheimer’s Disease Center, Rush University Medical Center, Chicago. Data collection was supported through funding by NIA grants P30AG10161 (ROS), R01AG15819 (ROSMAP; genomics and RNAseq), R01AG17917 (MAP), R01AG30146, R01AG36042 (5hC methylation, ATACseq), RC2AG036547 (H3K9Ac), R01AG36836 (RNAseq), R01AG48015 (monocyte RNAseq) RF1AG57473 (single nucleus RNAseq), U01AG32984 (genomic and whole exome sequencing), U01AG46152 (ROSMAP AMP-AD, targeted proteomics), U01AG46161(TMT proteomics), U01AG61356 (whole genome sequencing, targeted proteomics, ROSMAP AMP-AD), the Illinois Department of Public Health (ROSMAP), and the Translational Genomics Research Institute (genomic). Additional phenotypic data can be requested at www.radc.rush.edu. For MSBB, the data were generated from postmortem brain tissue collected through the Mount Sinai VA Medical Center Brain Bank and were provided by Dr. Eric Schadt from Mount Sinai School of Medicine.

This work was supported by grants from the US National Institutes of Health (NIH NIA R21-AG063130, NIA R01-AG054005, NIA R56-AG055824, and NIA U01-AG068880).

## Competing Interests Statement

HH is an employee of Eisai Inc. This work has been performed during his previous position at Sorbonne University, Paris, France. At Sorbonne University HH was supported by the AXA Research Fund, the “*Fondation partenariale Sorbonne Université*” and the “*Fondation pour la Recherche sur Alzheimer*”, Paris, France.

## Author contributions

**EADB Coordination**: A.B., K.M., F.J., M.T., R.F-S., J.C., J-F.D., O.A.A., M.I., M.H., K.S., C.VD., R.S., W.M.VDF., A.Ruiz., A.Ramirez., J-C.L; **Data Analyses** : C.B., F.K., I.J. V.A. S.M-G., N.A., B.G-B., P.A.H., R.C-M., L.K., V.D., S.J.VDL., T.K., I.R., J.C., M.C., P.G-G.; **ADGC analysis and coordination**: A.C.N, W.S.B, L.A.F., J.L.H., K.L.H-N., P.P.K., B.W.K., C-Y.L., E.R.M., R.M., M.A.P-V., J.S., L-S.W., Y.Z., G.D.S.; **Charge analysis and coordination**: Q.Y., J.C.B., A.DS., C.S., B.P., R.W., O.L., S.S.; **FinnGen analysis**: T.K., M.H.; **Rotterdam Analysis**: A.Y., I.P-N, M.G., A.I.; **Core writing group**: C.B., F.K., V.A., B.G-B., P.A.H., R.C-M., L.K., S.J.VDL., K.S., A.Ruiz., A.Ramirez., J-C.L. All other authors contributed data and all authors critically reviewed the paper.

## Data Availability Statement

Summary statistics will be made available upon publication.

All the data used in the gene prioritization are publically available:

AMP-AD rnaSeqReprocessing Study: https://www.synapse.org/#!Synapse:syn9702085

MayoRNAseq WGS VCFs: https://www.synapse.org/#!Synapse:syn11724002

ROSMAP WGS VCFs: https://www.synapse.org/#!Synapse:syn11724057

MSBB WGS VCFs: https://www.synapse.org/#!Synapse:syn11723899

GTEx pipeline: https://github.com/broadinstitute/gtex-pipeline

eQTLGen: https://www.eqtlgen.org/

eQTL Catalogue database: https://www.ebi.ac.uk/eqtl/

Brain xQTL serve: http://mostafavilab.stat.ubc.ca/xqtl/

GTEx v8 eQTL and sQTL catalogues: https://www.gtexportal.org/

GTEx v8 expression and splicing prediction models: http://predictdb.org/

MiGA eQTLs: https://doi.org/10.5281/zenodo.4118605

MiGA sQTLs: https://doi.org/10.5281/zenodo.4118403

MiGA Meta-analysis: https://doi.org/10.5281/zenodo.4118676

## Notes

### Author Declarations

Written informed consent was obtained from study participants or, for those with substantial cognitive impairment, from a caregiver, legal guardian or other proxy. Study protocols for all cohorts were reviewed and approved by the appropriate institutional review boards.

