## supplementary data for "New insights on the genetic etiology of Alzheimer’s and related dementia"

Céline Bellenguez[1,*,#], Fahri Küçükali[2,3,4,*], Iris Jansen[5,6,*], Victor Andrade[7,8,*], Sonia Moreno-Grau[9,10,*], Najaf Amin[11,12,*], Adam C. Naj[13,*], Benjamin Grenier-Boley[1], Rafael Campos-Martin[7], Peter A. Holmans[14], Anne Boland[15], Luca Kleineidam[7,8,16], Vincent Damotte[1], Sven J. van der Lee[5,17], Teemu Kuulasmaa[18], Qiong Yang[19,20], Itziar de Rojas[9,10], Joshua C. Bis[21], Amber Yaqub[11], Ivana Prokic[11], Marcos R, Costa[1,22], Julien Chapuis[1], Shahzad Ahmad[11,23], Vilmantas Giedraitis[24], Mercè Boada[9,10], Dag Aarsland[25,26], Pablo García-González[9,10], Carla Abdelnour[9,10], Emilio Alarcón-Martín[9,27], Montserrat Alegret[10,11], Ignacio Alvarez[28,29], Victoria Álvarez[30,31], Nicola J. Armstrong[32], Anthoula Tsolaki [33,34], Carmen Antúnez[35], Ildebrando Appollonio[36,37], Marina Arcaro[38], Silvana Archetti[39], Alfonso Arias Pastor[40,41], Beatrice Arosio[42,43], Lavinia Athanasiu[44], Henri Bailly[45], Nerisa Banaj[46], Miquel Baquero[47], Ana Belén Pastor[48], Luisa Benussi[49], Claudine Berr[50], Céline Besse[15], Valentina Bessi[51,52], Giuliano Binetti[49,53], Alessandra Bizzarro[54], Daniel Alcolea[10,55], Rafael Blesa[10,55], Barbara Borroni[39], Silvia Boschi[56], Paola Bossù[57], Geir Bråthen[58,59], Catherine Bresner[14], Keeley J. Brookes[60], Luis Ignacio Brusco[61,62,63], Katharina Bûrger[64,65], María J. Bullido[10,66,67], Vanessa Burholt[68,69], William S. Bush[70], Miguel Calero[10,48,71], Carole Dufouil[72,73], Ángel Carracedo[74,75], Roberta Cecchetti[76], Laura Cervera-Carles[10,55], Camille Charbonnier[77], Caterina Chillotti[78], Henry Brodaty[32,79], Simona Ciccone[43], Jurgen A.H.R. Claassen[80], Christopher Clark[14], Elisa Conti[36], Anaïs Corma-Gómez[81], Emanuele Costantini[82], Carlo Custodero[83], Delphine Daian[15], Maria Carolina Dalmasso[7], Antonio Daniele[82], Efthimios Dardiotis[84], Jean-François Dartigues[85], Peter Paul de Deyn[86], Katia de Paiva Lopes[87,88,89,90], Lot D. de Witte[91], Stéphanie Debette[85,92], Jürgen Deckert[93], Teodoro del Ser[48], Nicola Denning[94], Anita DeStefano[19,20,95], Martin Dichgans[64,65,96], Janine Diehl-Schmid[97], Mónica Diez-Fairen[28,29], Paolo Dionigi Rossi[43], Srdjan Djurovic[44], Emmanuelle Duron[45], Emrah Düzel[98,99], Sebastiaan Engelborghs[100 ,101,102,103], Valentina Escott-Price[14,94], Ana Espinosa[9,10], Dolores Buiza-Rueda[10,104], Michael Ewers[64,65], Fabrizio Tagliavini[105], Sune Fallgaard Nielsen[106], Lucia Farotti[107], Chiara Fenoglio[108], Marta Fernández-Fuertes[81], John Hardy[109], Raffaele Ferrari[109], Catarina B Ferreira[110], Evelyn Ferri[43], Bertrand Fin[15], Peter Fischer[111], Tormod Fladby[112], Klaus Fließbach[8,16], Juan Fortea[10,55], Silvia Fostinelli[49], Nick C. Fox[113], Emlio Franco-Macías[114], Ana Frank-García[10,67,115], Lutz Froelich[116], Daniela Galimberti[38,108], Jose Maria García-Alberca[10,117], Sebastian Garcia-Madrona[118], Guillermo García-Ribas[118], Geneviève Chene[72,73], Roberta Ghidoni[49], Ina Giegling[119], Giorgio Giaccone[105], Oliver Goldhardt[97], Antonio González-Pérez[120], Caroline Graff[121], Giulia Grande[122], Emma Green[123], Timo Grimmer[97], Edna Grünblatt[124,125], Tamar Guetta-Baranes[126], Annakaisa Haapasalo[127], Georgios Hadjigeorgiou[128], Jonathan L. Haines[70], Kara L. Hamilton-Nelson[129], EADB, Gra@ce, ADGC, Charge, DemGen, FinnGen, EADI, GERAD, Harald Hampel[130], Olivier Hanon[45], Annette M. Hartmann[119], Lucrezia Hausner[116], Janet Harwood[14], Stefanie Heilmann-Heimbach[131], Seppo Helisalmi[132,133], Michael T. Heneka[8,16], Isabel Hernández[9,10], Martin J. Herrmann[93], Per Hoffmann[131], Clive Holmes[134], Henne Holstege[5,17], Raquel Huerto Vilas[40,41], Marc Hulsman[5,17], Jack Humphrey[87,88,89,90], Geert Jan Biessels[135], Charlotte Johansson[121], Patrick G. Kehoe[136], Lena Kilander[24], Anne Kinhult Ståhlbom[121], Miia Kivipelto[137,138,139,140], Anne Koivisto[132,141,142], Johannes Kornhuber[143], Mary H. Kosmidis[144], Pavel P. Kuksa[145], Brian W. Kunkle[129], Carmen Lage[146], Erika J Laukka[122,147], Alessandra Lauria[54], Chien-Yueh Lee[145], Jenni Lehtisalo[132,148], Claudia L. Satizabal[20,95,149], Ondrej Lerch[150,151], Alberto Lleó[10,55], Rogelio Lopez[9], Oscar Lopez[21], Adolfo Lopez de Munain[10,152], Seth Love[135], Malin Löwemark[24], Lauren Luckcuck[14], Juan Macías[81], Catherine A. MacLeod[153], Wolfgang Maier[8,16], Francesca Mangialasche[137], Marco Spallazzi[154], Marta Marquié[10,11], Rachel Marshall[14], Eden R. Martin[129], Angel Martín Montes[10,67,115], Carmen Martínez Rodríguez[31], Carlo Masullo[155], Richard Mayeux[156,157], Simon Mead[158], Patrizia Mecocci[76], Miguel Medina[10,48], Alun Meggy[94], Silvia Mendoza[117], Manuel Menéndez-González[31], Pablo Mir[10,104], Maria Teresa Periñán[10,104], Merel Mol[159], Laura Molina-Porcel[160,161], Laura Montrreal[9], Laura Morelli[162], Fermín Moreno[10,152], Kevin Morgan[163], Markus M. Nöthen[131], Carolina Muchnik[61,164], Benedetta Nacmias[51,165], Tiia Ngandu[148], Gael Nicolas[77], Børge G. Nordestgaard[106,166], Robert Olaso[15], Adelina Orellana[9,10], Michela Orsini[82], Gemma Ortega[9,10], Alessandro Padovani[39], Paolo Caffarra[154], Goran Papenberg[122], Lucilla Parnetti[107], Florence Pasquier[167], Pau Pastor[28,29], Alba Pérez-Cordón[9], Jordi Pérez-Tur[10,168,169],, Pierre Pericard[170], Oliver Peters[171,172], Yolande A.L. Pijnenburg[5], Juan A Pineda[81], Gerard Piñol-Ripoll[40,41], Claudia Pisanu[78], Thomas Polak[93], Julius Popp[173,174,175], Danielle Posthuma[6], Josef Priller[172,176], Raquel Puerta[9], Olivier Quenez[77], Inés Quintela[74], Jesper Qvist Thomassen[106],

Alberto Rábano[48], Innocenzo Rainero[56], Inez Ramakers[177], Luis M Real[81,178], Marcel J.T. Reinders[179], Steffi Riedel-Heller[180], Peter Riederer[111], Eloy Rodriguez-Rodriguez[10,146], Arvid Rongve[181,182], Irene Rosas Allende[30,31], Maitée Rosende-Roca[9,10], Jose Luis Royo[183], Elisa Rubino[184], Dan Rujescu[119], María Eugenia Sáez[120], Paraskevi Sakka[185], Ingvild Saltvedt[59,186], Ángela Sanabria[9,10], María Bernal Sánchez-Arjona[114], Florentino Sanchez-Garcia[187], Shima Mehrabian[188], Pascual Sánchez-Juan[10,146], Raquel Sánchez-Valle[189], Sigrid B Sando[58,59], Michela Scamosci[76], Nikolaos Scarmeas[156,190], Elio Scarpini[38,108], Philip Scheltens[5], Norbert Scherbaum[191], Martin Scherer[192], Matthias Schmid[16,193], Anja Schneider[9,16], Jonathan M. Schott[113], Geir Selbæk[112,194], Jin Sha[13], Alexey A Shadrin[44], Olivia Skrobot[136], Gijsje J. L. Snijders[91], Hilkka Soininen[132], Vincenzo Solfrizzi[83], Alina Solomon[132,137], Sandro Sorbi[51,165], Oscar Sotolongo-Grau[9], Gianfranco Spalletta[46], Annika Spottke[16], Alessio Squassina[78], Juan Pablo Tartari[9], Lluís Tárraga[9,10], Niccolo Tesí[5,17], Anbupalam Thalamuthu[32], Thomas Tegos[33,34], Latchezar Traykov[158], Lucio Tremolizzo[36,37], Anne Tybjærg-Hansen[106,166], Andre Uitterlinden[195], Abbe Ullgren[121], Ingun Ulstein[194], Sergi Valero[9,10], Christine Van Broeckhoven[2,3,196], Aad van der Lugt[197], Jasper Van Dongen[2,3,4], Jeroen van Rooij[159,195], John van Swieten[159], Rik Vandenberghe[198,199], Frans Verhey[177], Jean-Sébastien Vidal[45], Jonathan Vogelgsang[200,201], Martin Vyhnalek[150,151], Michael Wagner[9,16], David Wallon[202], Li-San Wang[145], Ruiqi Wang[19,20], Leonie Weinhold[193], Jens Wiltfang[200,203,204], Gill Windle[153], Bob Woods[153], Mary Yannakoulia[205], Yi Zhao[145], Miren Zulaica[10,206], Manuel Serrano-Rios[207], Davide Seripa[208], Eystein Stordal[209], Lindsay A. Farrer[19,95,210,211], Bruce M. Psaty[21,212], Mohsen Ghanbari[11], Towfique Raj[87,88,89,90], Perminder Sachdev[32], Karen Mather[32], Frank Jessen[16,213], M. Arfan Ikram[11], Alexandre de Mendonça[110], Jakub Hort[150,151], Magda Tsolaki[33,34], Margaret A. Pericak-Vance[129], Philippe Amouyel[1], Julie Williams[14,94], Ruth Frikke-Schmidt[106,166], Jordi Clarimon[10,55], Jean-François Deleuze[15], Giacomina Rossi[105], Sudha Seshadri[20,95,149], Ole A. Andreassen[44,214], Martin Ingelsson[24], Mikko Hiltunen[18,**], Kristel Sleegers[2,3,4,**], Gerard D. Schellenberg[145,**], Cornelia M. van Duijn[11,12,**], Rebecca Sims[14,**], Wiesje M. van der Flier[5,**], Agustín Ruiz[9,10,**], Alfredo Ramirez[7,8,16,215,**], Jean-Charles Lambert[1,**,#]

*  co-first
** co-last

\# corresponding author:

Céline Bellenguez:
Jean-Charles Lambert:

1. Univ. Lille, Inserm, CHU Lille, Institut Pasteur de Lille, U1167-RID-AGE facteurs de risque et déterminants moléculaires des maladies liés au vieillissement, Lille, France
2. Complex Genetics of Alzheimer's Disease Group, VIB Center for Molecular Neurology, VIB, Antwerp, Belgium
3. Laboratory of Neurogenetics, Institute Born - Bunge, Antwerp, Belgium
4. Department of Biomedical Sciences, University of Antwerp, Neurodegenerative Brain Diseases Group, Center for Molecular Neurology, VIB, Antwerp, Belgium
5. Alzheimer Center Amsterdam, Department of Neurology, Amsterdam Neuroscience, Vrije Universiteit Amsterdam, Amsterdam UMC, Amsterdam, The Netherlands
6. Department of Complex Trait Genetics, Center for Neurogenomics and Cognitive Research, Amsterdam Neuroscience, Vrije University, Amsterdam, The Netherlands.
7. Division of Neurogenetics and Molecular Psychiatry, Department of Psychiatry and Psychotherapy, University of Cologne, Medical Faculty, Cologne, Germany.
8. Department of Neurodegenerative Diseases and Geriatric Psychiatry, University Hospital Bonn, Bonn, Germany
9. Research Center and Memory clinic Fundació ACE, Institut Català de Neurociències Aplicades, Universitat Internacional de Catalunya, Barcelona, Spain
10. CIBERNED, Network Center for Biomedical Research in Neurodegenerative Diseases, National Institute of Health Carlos III, Madrid, Spain
11. Department of Epidemiology, ErasmusMC, Rotterdam, The Netherlands
12. Nuffield Department of Population Health Oxford University, Oxford, UK
13. Department of Biostatistics, Epidemiology, and Informatics; Penn Neurodegeneration Genomics Center, Department of Pathology and Laboratory Medicine, University of Pennsylvania Perelman School of Medicine, Philadelphia, Pennsylvania, USA

14. MRC Centre for Neuropsychiatric Genetics and Genomics, Division of Psychological Medicine and Clinical Neuroscience, School of Medicine, Cardiff University, Cardiff, UK
15. Université Paris-Saclay, CEA, Centre National de Recherche en Génomique Humaine, 91057, Evry, France
16. German Center for Neurodegenerative Diseases (DZNE Bonn), Bonn, Germany
17. Department of Clinical Genetics, VU University Medical Centre, Amsterdam, The Netherlands
18. Institute of Biomedicine, University of Eastern Finland, Kuopio, Finland
19. Department of Biostatistics, Boston University School of Public Health, Boston, MA, USA.
20. Framingham Heart Study, Framingham, MA, USA.
21. Cardiovascular Health Research Unit, Department of Medicine, University of Washington, Seattle, WA, USA.
22. Brain Institute, Federal University of Rio Grande do Norte, Av. Nascimento de Castro 2155 Natal, Brazil
23. LACDR, Leiden, The Netherlands
24. Dept.of Public Health and Carins Sciences / Ger+iatrics, Uppsala University
25. Centre of Age-Related Medicine, Stavanger University Hospital, Norway
26. Institute of Psychiatry, Psychology & Neuroscience, PO 70, 16 De Crespigny Park, London, UK
27. Department of Surgery, Biochemistry and Molecular Biology, School of Medicine, University of Málaga, Málaga, Spain.
28. Fundació Docència i Recerca Mútua de Terrassa, Terrassa 08221, Barcelona, Spain
29. Memory Disorders Unit, Department of Neurology, Hospital Universitari Mútua de Terrassa, Terrassa, Barcelona, Spain.
30. Laboratorio de Genética. Hospital Universitario Central de Asturias, Oviedo, Spain
31. Servicio de Neurología HOspital Universitario Central de Asturias- Oviedo and Instituto de Investigación Biosanitaria del Principado de Asturias, Oviedo, Spain
32. Centre for Healthy Brain Ageing, School of Psychiatry, Faculty of Medicine, University of New South Wales, Sydney, Australia
33. 1st Department of Neurology, Medical school, Aristotle University of Thessaloniki, Thessaloniki, Makedonia, Greece
34. Alzheimer Hellas, Thessaloniki, Makedonia, Greece
35. Unidad de Demencias, Hospital Clínico Universitario Virgen de la Arrixaca, Spain
36. School of Medicine and Surgery, University of Milano-Bicocca, Italy
37. Neurology Unit, "San Gerardo" hospital, Monza, Italy
38. Fondazione IRCCS Ca' Granda, Ospedale Policlinico, Milan, Italy
39. Department of Laboratory Diagnostics, III Laboratory of Analysis, Brescia Hospital, Brescia, Italy
40. Unitat Trastorns Cognitius, Hospital Universitari Santa Maria de Lleida, Lleida, Spain
41. Institut de Recerca Biomedica de Lleida (IRBLLeida), Lleida, Spain
42. Department of Clinical Sciences and Community Health, University of Milan, Italy
43. Geriatic Unit, Fondazione Cà Granda, IRCCS Ospedale Maggiore Policlinico, Milan, Italy
44. NORMENT Centre, University of Oslo, Oslo, Norway
45. Université de Paris, EA 4468, APHP, Hôpital Broca, Paris, France
46. Laboratory of Neuropsychiatry, Department of Clinical and Behavioral Neurology, IRCCS Santa Lucia Foundation, Rome, Italy
47. Servei de Neurologia, Hospital Universitari i Politècnic La Fe, Valencia, Spain.
48. CIEN Foundation/Queen Sofia Foundation Alzheimer Center, Madrid, Spain
49. Molecular Markers Laboratory, IRCCS Istituto Centro San Giovanni di Dio Fatebenefratelli, Brescia, Italy
50. Univ. Montpellier, Inserm U1061, Neuropsychiatry: epidemiological and clinical research, PSNREC, Montpellier, France
51. Department of Neuroscience, Psychology, Drug Research and Child Health University of Florence, Florence Italy
52. Azienda Ospedaliero-Universitaria Careggi. Largo Brambilla, 3, 50134, Florence, Italy
53. MAC - Memory Clinic, IRCCS Istituto Centro San Giovanni di Dio Fatebenefratelli, Brescia, Italy
54. Geriatrics Unit Fondazione Policlinico A. Gemelli IRCCS, Rome, Italy
55. Department of Neurology, II B Sant Pau, Hospital de la Santa Creu i Sant Pau, Universitat Autònoma de Barcelona, Barcelona, Spain.
56. Department of Neuroscience “Rita Levi Montalcini”, University of Torino, Torino, Italy
57. Experimental Neuro-psychobiology Laboratory,Department of Clinical and Behavioral Neurology, IRCCS Santa Lucia Foundation, Rome, Italy
58. Department of Neurology and Clinical Neurophysiology, University Hospital of Trondheim, Trondheim, Norway

59. Department of Neuromedicine and Movement Science, Norwegian University of Science and Technology, Trondheim, Norway
60. Biosciences, School of Science and Technology, Nottingham Trent University, Nottingham, UK
61. Centro de Neuropsiquiatría y Neurología de la Conducta (CENECON), Facultad de Medicina, Universidad de Buenos Aires (UBA), C.A.B.A, Buenos Aires, Argentina.
62. Departamento Ciencias Fisiológicas UAII, Facultad de Medicina, UBA, C.A.B.A, Buenos Aires, Argentina.
63. Hospital Interzonal General de Agudos Eva Perón, San Martín, Buenos Aires, Argentina.
64. Institute for Stroke and Dementia Research (ISD), University Hospital, LMU Munich, Munich, Germany
65. German Center for Neurodegenerative Diseases (DZNE, Munich), Munich, Germany.
66. Centro de Biología Molecular Severo Ochoa (UAM-CSIC), Madrid, Spain
67. Instituto de Investigacion Sanitaria ‘Hospital la Paz’ (IdIPaz), Madrid, Spain
68. Faculty of Medical & Health Sciences, University of Auckland, New Zealand
69. Wales Centre for Ageing & Dementia Research, Auckland, New Zealand
70. Department of Population & Quantitative Health Sciences, Case Western Reserve University, Cleveland, Ohio, USA
71. UFIEC, Instituto de Salud Carlos III, Madrid, Spain
72. Inserm, Bordeaux Population Health Research Center, UMR 1219, Univ. Bordeaux, ISPED, CIC 1401-EC, Univ Bordeaux, Bordeaux, France
73. CHU de Bordeaux, Pole santé publique, Bordeaux, France
74. Grupo de Medicina Xenómica, Centro Nacional de Genotipado (CEGEN-PRB3-ISCIII). Universidade de Santiago de Compostela, Santiago de Compostela, Spain.
75. Fundación Pública Galega de Medicina Xenómica- CIBERER-IDIS, University of Santiago de Compostela, Santiago de Compostela, Spain.
76. Institute of Gerontology and Geriatrics, Department of Medicine and Surgery, University of Perugia Perugia, Italy
77. Normandie Univ, UNIROUEN, Inserm U1245 and Rouen University Hospital, Department of Genetics and CNR-MAJ, F 76000, Normandy Center for Genomic and Personalized Medicine, Rouen, France
78. Unit of Clinical Pharmacology, University Hospital of Cagliari, Cagliari, Italy
79. Centre for Healthy Brain Ageing, School of Psychiatry, Faculty of Medicine, University of New South Wales, Sydney, Australia
80. Radboudumc Alzheimer Center, Department of Geriatrics, Radboud University Medical Center, Nijmegen, the Netherlands
81. Unidad Clínica de Enfermedades Infecciosas y Microbiología. Hospital Universitario de Valme, Sevilla, Spain
82. Department of Neuroscience, Catholic University of Sacred Heart, Fondazione Policlinico Universitario A. Gemelli IRCCS, Rome, Italy
83. University of Bari, “A. Moro”, Bari, Italy
84. School of Medicine, University of Thessaly, Larissa, Greece
85. University Bordeaux, Inserm, Bordeaux Population Health Research Center, France
86. Department of Neurology, University Medical Center Groningen, the Netherlands
87. Nash Family Department of Neuroscience & Friedman Brain Institute, Icahn School of Medicine at Mount Sinai, New York, USA
88. Ronald M. Loeb Center for Alzheimer’s disease, Icahn School of Medicine at Mount Sinai, New York, USA
89. Department of Genetics and Genomic Sciences & Icahn Institute for Data Science and Genomic Technology, Icahn School of Medicine at Mount Sinai, New York, USA
90. Estelle and Daniel Maggin Department of Neurology, Icahn School of Medicine at Mount Sinai, New York, USA
91. Department of Psychiatry, Icahn School of Medicine at Mount Sinai, New York
92. Department of Neurology, Bordeaux University Hospital, Bordeaux, France
93. Department of Psychiatry, Psychosomatics and Psychotherapy, Center of Mental Health, University Hospital, Wuerzburg
94. UKDRI@ Cardiff, School of Medicine, Cardiff University, Cardiff, UK
95. Department of Neurology, Boston University School of Medicine, Boston, MA, USA.
96. Munich Cluster for Systems Neurology (SyNergy), Munich, Germany.
97. Technical University of Munich, School of Medicine, Klinikum rechts der Isar, Department of Psychiatry and Psychotherapy, Munich, Germany
98. Institute of Cognitive Neurology and Dementia Research (IKND), Otto-Von-Guericke University, Magdeburg, Germany.

99. German Center for Neurodegenerative Diseases (DZNE), Magdeburg, Germany.
100. Center for Neurosciences, Vrije Universiteit Brussel (VUB), Brussels, Belgium
101. Reference Center for Biological Markers of Dementia (BIODEM), Institute Born-Bunge, University of Antwerp, Antwerp, Belgium
102. Institute Born-Bunge, University of Antwerp, Antwerp, Belgium
103. Department of Neurology, UZ Brussel, Brussels, Belgium
104. Unidad de Trastornos del Movimiento, Servicio de Neurología y Neurofisiología. Instituto de Biomedicina de Sevilla (IBiS), Hospital Universitario Virgen del Rocío/CSIC/Universidad de Sevilla, Seville, Spain
105. Fondazione IRCCS, Istituto Neurologico Carlo Besta, Milan Italy
106. Department of Clinical Biochemistry, Herlev and Gentofte Hospital, Herlev Denmark
107. Centre for Memory Disturbances, Lab of Clinical Neurochemistry, Section of Neurology, University of Perugia, Italy
108. University of Milan, Milan, Italy
109. Laboratory of Neurogenetics, Department of Internal Medicine, Texas Tech University Health Science Center, Lubbock, Texas, USA; Reta Lila Weston Research Laboratories, Department of Molecular
110. Faculty of Medicine, University of Lisbon, Portugal
111. Department of Psychiatry, Social Medicine Center East- Donauspital, Vienna, Austria
112. Institute of Clinical Medicine, University of Oslo, Oslo, Norway.
113. Dementia Research Centre, UCL Queen Square Institute of Neurology, London, United Kingdom
114. Unidad de Demencias, Servicio de Neurología y Neurofisiología. Instituto de Biomedicina de Sevilla (IBiS), Hospital Universitario Virgen del Rocío/CSIC/Universidad de Sevilla, Seville, Spain
115. Hospital Universitario la Paz, Madrid, Spain
116. Department of geriatric Psychiatry, Central Institute for Mental Health, Mannheim, University of Heidelberg, Germany
117. Alzheimer Research Center & Memory Clinic, Andalusian Institute for Neuroscience, Málaga, Spain.
118. Hospital Universitario Ramon y Cajal, IRYCIS, Madrid
119. Martin-Luther-University Halle-Wittenberg, University Clinic and Outpatient Clinic for Psychiatry, Psychotherapy and Psychosomatics, Halle (Saale), Germany
120. CAEBI, Centro Andaluz de Estudios Bioinformáticos, Sevilla, Spain.
121. Unit for Hereditary Dementias, Theme Aging, Karolinska University Hospital-Solna, Stockholm Sweden
122. Aging Research Center, Department of Neurobiology, Care Sciences and Society, Karolinska Institutet and Stockholm University, Stockholm, Sweden
123. Institute of Public Health, University of Cambridge, UK
124. Department of Child and Adolescent Psychiatry and Psychotherapy, University Hospital of Psychiatry Zurich, University of Zurich, Zurich, Switzerland
125. Neuroscience Center Zurich, University of Zurich and ETH Zurich, Switzerland
126. Human Genetics, School of Life Sciences, Life Sciences Building, University Park, University of Nottingham, Nottingham, UK
127. A.I Virtanen Institute for Molecular Sciences, University of Eastern Finland, Kuopio, Finland
128. Department of Neurology, Medical School, University of Cyprus, Cyprus
129. The John P. Hussman Institute for Human Genomics, University of Miami, Miami, Florida, USA
130. Sorbonne University, GRC n° 21, Alzheimer Precision Medicine Initiative (APMI), AP-HP, Pitié-Salpêtrière Hospital, Boulevard de l'hôpital, F-75013, Paris, France; Eisai Inc., Neurology Business Group, 100 Tice Blvd, Woodcliff Lake, NJ 07677, USA
131. Institute of Human Genetics, University of Bonn, School of Medicine & University Hospital Bonn, Bonn, Germany
132. Institute of Clinical Medicine - Neurology, University of Eastern, Kuopio, Finland
133. Institute of Clinical Medicine – Internal Medicine, University of Eastern Finland, Kuopio, Finland
134. Clinical and Experimental Science, Faculty of Medicine, University of Southampton, Southampton, UK
135. Department of Neurology, UMC Utrecht Brain Center, Utrecht, the Netherlands
136. Translational Health Sciences, Bristol Medical School, University of Bristol, Bristol, UK
137. Division of Clinical Geriatrics, Center for Alzheimer Research, Care Sciences and Society (NVS), Karolinska Institutet, Stockholm, Sweden
138. Institute of Public Health and Clinical Nutrition, University of Eastern Finland, Kuopio, Finland
139. Neuroepidemiology and Ageing Research Unit, School of Public Health, Imperial College London, London, United Kingdom
140. Stockholms Sjukhem, Research & Development Unit, Stockholm, Sweden
141. Department of Neurology, Kuopio University Hospital, Kuopio, Finland

142. Department of Neurosciences, University of Helsinki and Department of Geriatrics, Helsinki University Hospital, Helsinki, Finland
143. Department of Psychiatry and Psychotherapy, Universitätsklinikum Erlangen, and Friedrich-Alexander Universität Erlangen-Nürnberg, Erlangen, Germany.
144. Laboratory of Cognitive Neuroscience, School of Psychology, Aristotle University of Thessaloniki, Thessaloniki, Greece
145. Penn Neurodegeneration Genomics Center, Department of Pathology and Laboratory Medicine, University of Pennsylvania Perelman School of Medicine, Philadelphia, Pennsylvania, USA
146. Neurology Service, Marqués de Valdecilla University Hospital (University of Cantabria and IDIVAL), Santander, Spain.
147. Stockholm Gerontology Research Center, Stockholm, Sweden
148. Public Health Promotion Unit, Finnish Institute for Health and Welfare, Helsinki, Finland
149. Glenn Biggs Institute for Alzheimer's and Neurodegenerative Diseases, San Antonio, TX, USA
150. Memory Clinic, Department of Neurology, Charles University, 2nd Faculty of Medicine and Motol University Hospital, Czech Republic
151. International Clinical Research Center, St. Anne's University Hospital Brno, Brno, Czech Republic
152. Department of Neurology. Hospital Universitario Donostia. OSAKIDETZA-Servicio Vasco de Salud, San Sebastian, Spain
153. School of Health Sciences, Bangor University, UK
154. Unit of Neurology, University of Parma and AOU, Parma, Italy
155. Institute of Neurology, Catholic University of the Sacred Heart, Rome, Itlay
156. Taub Institute on Alzheimer's Disease and the Aging Brain, Department of Neurology, Columbia University, New York, New York, USA
157. Gertrude H. Sergievsky Center, Columbia University, New York, New York, USA
158. MRC Prion Unit at UCL, UCL Institute of Prion Diseases, London, UK
159. Department of Neurology, ErasmusMC, Rotterdam, The Netherlands
160. Neurological Tissue Bank of the Biobanc-Hospital Clinic-IDIBAPS, Institut d'Investigacions Biomèdiques August Pi i Sunyer, Barcelona, Spain.
161. Alzheimer's disease and other cognitive disorders Unit. Neurology Department, Hospital Clinic, Barcelona, Spain
162. Laboratory of Brain Aging and Neurodegeneration- FIL-CONICET, Buenos Aires, Argentina
163. Human Genetics, School of Life Sciences, University of Nottingham, Nottinghma, UK
164. Laboratorio de Bioquímica Molecular, Facultad de Medicina, Instituto de Investigaciones Médicas A. Lanari, UBA, C.A.B.A, Buenos Aires, Argentina.
165. IRCCS Fondazione Don Carlo Gnocchi, Florence, Italy
166. Department of Clinical Medicine, University of Copenhagen, Copenhagen, Denmark
167. Univ Lille Inserm 1171, CHU Clinical and Research Memory Research Centre (CMRR) of Distalz Lille France.
168. Institut de Biomedicina de València-CSIC, CIBERNED, Valencia, valència, Spain
169. Unitat Mixta de de Neurología y Genética, Institut d'Investigació Sanitària La Fe, València, Spain
170. Univ. Lille, CNRS, Inserm, CHU Lille, Institut Pasteur de Lille, US 41-UMS 2014-PLBS, bilille, Lille, France.
171. Institute of Psychiatry and Psychotherapy, Charité-Universitätsmedizin Berlin, Corporate Member of Freie Universität Berlin, Humboldt-Universität Zu Berlin, and Berlin Institute of Health, Berlin, Germany.
172. German Center for Neurodegenerative Diseases (DZNE), Berlin, Germany.
173. CHUV, Old Age Psychiatry, Department of Psychiatry, Lausanne, Switzerland
174. Old Age Psychiatry, Department of Psychiatry, Lausanne University Hospital, Lausanne, Switzerland
175. Department of Geriatric Psychiatry, University Hospital of Psychiatry Zürich, Zürich, Switzerland
176. Department of Neuropsychiatry and Laboratory of Molecular Psychiatry, Charité, Charitéplatz 1, 10117 Berlin, Germany
177. Maastricht University, Department of Psychiatry & Neuropsychologie, Alzheimer Center Limburg, Maastricht, the Netherlands
178. Depatamento de Especialidades Quirúrgicas, Bioquímica e Inmunología. Facultad de Medicina. Universidad de Málaga. Málaga, Spain
179. Delft Bioinformatics Lab, Delft University of Technology, Delft, The Netherlands
180. Institute of Social Medicine, Occupational Health and Public Health, University of Leipzig, 04103 Leipzig, Germany.
181. Department of Research and Innovation, Helse Fonna, Haugesund Hospital, Haugesund, Norway.
182. The University of Bergen, Institute of Clinical Medicine (K1), Bergen Norway

183. Departamento de Especialidades Quirúrgicas, Bioquímicas e Inmunología, School of Medicine, University of Málaga, Málaga, Spain.
184. Department of Neuroscience and Mental Health, AOU Città della Salute e della Scienza di Torino, Torino, Italy
185. Athens Association of Alzheimer's disease and Related Disorders, Athens, Greece
186. Department of Geriatrics, St. Olav's Hospital, Trondheim University Hospital, Norway
187. Department of Immunology, Hospital Universitario Doctor Negrín, Las Palmas de Gran Canaria, Spain.
188. Clinic of Neurology, UH "Alexandrovska", Medical University - Sofia, Sofia, Bulgaria
189. Neurology department-Hospital Clínic, IDIBAPS, Universitat de Barcelona, Barcelona, Spain.
190. 1st Department of Neurology, Aiginition Hospital, National and Kapodistrian University of Athens, Medical School, Greece
191. LVR-Hospital Essen, Department of Psychiatry and Psychotherapy, Medical Faculty, University of Duisburg-Essen, Essen, Germany
192. Department of Primary Medical Care, University Medical Centre Hamburg-Eppendorf, Hamburg, Germany.
193. Institute of Medical Biometry, Informatics and Epidemiology, University Hospital of Bonn, Bonn, Germany.
194. Department of Geriatric Medicine, Oslo University Hospital, Oslo, Norway
195. Department of Internal medicine and Biostatistics, ErasmusMC, Rotterdam, The Netherlands
196. Neurodegenerative Brain Diseases Group, VIB Center for Molecular Neurology, VIB, Antwerp, Belgium
197. Department of Radiology&Nuclear medicine, ErasmusMC, Rotterdam, The Netherlands
198. Laboratory for Cognitive Neurology, Department of Neurosciences, University of Leuven, Belgium
199. Neurology Department, University Hospitals Leuven, Leuven, Belgium
200. Department of Psychiatry and Psychotherapy, University Medical Center Goettingen, Goettingen, Germany
201. Department of Psychiatry, Harvard Medical School, McLean Hospital, Belmont, MA, USA
202. Normandie Univ, UNIROUEN, Inserm U1245 and Rouen University Hospital, Department of Neurology and CNR-MAJ, F 76000, Normandy Center for Genomic and Personalized Medicine, Rouen, France
203. German Center for Neurodegenerative Diseases (DZNE), Goettingen, Germany
204. Medical Science Department, iBiMED, Aveiro, Portugal
205. Department of Nutrition and Diatetics, Harokopio University, Athens, Greece
206. Neurosciences Area. Instituto Biodonostia. San Sebastian, Spain
207. Centro de Investigación Biomédica en Red de Diabetes y Enfermedades Metabólicas Asociadas, CIBERDEM, Spain, Hospital Clínico San Carlos, Madrid, Spain.
208. Laboratory for Advanced Hematological Diagnostics, Department of Hematology and Stem Cell Transplant, Lecce, Italy
209. Department of Psychiatry, Namsos Hospital, Namsos, Norway
210. Department of Epidemiology, Boston University, Boston, Massachusetts, USA
211. Departments of Medicine (Biomedical Genetics) and Ophthalmology, Boston University, Boston, Massachusetts, USA
212. Department of Epidemiology, University of Washington, Seattle, WA; and Department of Health Services, University of Washington, Seattle, WA, USA
213. Department of Psychiatry and Psychotherapy, University of Cologne, Medical Faculty, Cologne, Germany
214. Oslo University Hospital, Kirkeveien 166, 0407 Oslo, Norway
215. Department of Psychiatry and Glenn Biggs Institute for Alzheimer's and Neurodegenerative Diseases, San Antonio, TX, USA

**TABLE OF CONTENTS**


**1. Sample description 8**
1.1. Stage I samples 8
1.2. Stage II samples 18
1.3. Longitudinal studies 26

**2. Quality control 26**
2.1. EADB 26
2.2. Other datasets 30

**3. Imputations 32**

**4. GRCh37/GRCh38 conversion 33**

**5. Stage II analyses 33**

**6. Conditional analyses 35**

**7. HLA analyses 35**

**8. PheWAS 36**

**9. GWAS signal colocalization analyses 37**

**10. Gene prioritization 38**
10.1. Gene prioritization methods 38
10.2. Gene prioritization results 43
10.3. Short description of some functions of the prioritized genes and their potential implication in AD 50

**11. STRING protein interaction analysis 55**

**12. List of URLs 55**

**13. Supplementary References 56**

**14. Supplementary List of Authors 68**

**15. Supplementary Figures 77**

### 1. Sample description

#### 1.1. Stage I samples

***The European Alzheimer's Disease DNA Biobank dataset (EADB)***

This consortium groups together 20,464 Alzheimer's disease (AD) cases and 22,244 controls after quality controls from 15 European countries (Belgium, Bulgaria, Czech Republic, Denmark, Finland, France, Germany, Greece, Italy, Portugal, Spain, Sweden, Switzerland, The Netherlands and the UK). These samples were genotyped in three independent centers (France, Germany and the Netherlands) leading to define three nodes: EADB-France, EADB-Germany and EADB-Netherlands. In addition, EADB also included Australian partners.

**EADB-France**

In the France node, samples were collected from nine countries (39 centers/studies), and after quality controls (QCs), we obtained 13,867 AD cases and 15,310 controls. All these samples were genotyped at the Centre National de Recherche en Génomique Humaine (CNRGH, Evry, France).

Belgium: The participants were part of a large prospective cohort[1] of Belgian AD patients and healthy elderly control individuals. The patients were ascertained at the memory clinic of Middelheim and Hoge Beuken (Hospital Network Antwerp, Belgium) and at the memory clinic of the University Hospitals of Leuven, Belgium. The control individuals were the partners of the patients or volunteers from the Belgian community. The study protocols were approved by the ethics committees of the Antwerp University Hospital and the participating neurological centers at the different hospitals of the BELNEU consortium and by the University of Antwerp.

Czech Republic: The Czech Brain Aging Study (CBAS)[2] is a longitudinal memory-clinic–based study recruiting subjects at risk of dementia (subjects referred for cognitive complaints-SCD, MCI). The CBAS+ study is a cross-sectional study of patients in the early stages of dementia. All subjects signed informed consent and both studies were approved by the local ethics committee.

Denmark: The Copenhagen General Population Study (CGPS) is a prospective study of the Danish general population initiated in 2003 and still recruiting. Individuals were selected randomly based on the national Danish Civil Registration System to reflect the adult Danish population aged 20-100. Data were obtained from a self-administered questionnaire reviewed together with an investigator at the day of attendance, a physical examination, and from blood samples including DNA extraction.

Finland: *The ADGEN cohort*[3]: a clinic-based collection of AD patients from Eastern and Northern Finland examined in the Department of Neurology in Kuopio University Hospital and the Department of Neurology in Oulu University Hospital. All the patients were diagnosed with probable AD according to the criteria of the National Institute of Neurological and Communicative Disorders and Stroke and the Alzheimer's disease and Related Disorders Association (NINCDS–ADRDA). The study was approved by the ethics committee of Kuopio University Hospital, Finland (420/2016). *The FINGER study*[4]: a Finnish multi-domain lifestyle RCT enrolling 1,260 older adults with an increased risk of dementia from the general population. The intensive lifestyle intervention lasted for two years, and follow-up extends currently up to seven years. The FINGER study was approved by the coordinating ethics committee of the Hospital District of Helsinki and Uusimaa (94/13/03/00/2009 and HUS/1204/2017), and all the participants gave written informed consent.

France: *The BALTAZAR multicenter (23 memory centers) prospective study*[5]: 1,040 participants from September 2010 to April 2015. They were classified as AD cases (n = 501) according to DSM IV-TR and NINCDS–ADRDA criteria as well as amnestic mild cognitive impairment (MCI) cases (a MCI, n = 417) and non-amnestic MCI cases (na MCI, n = 122) according to Petersen's criteria. A comprehensive battery of cognitive tests was performed, including MMSE, verbal fluency, and FCSRT. All the participants or their legal guardians gave written informed consent. The study was approved by the Paris ethics committee (CPP Ile de France IV Saint Louis Hospital). *MEMENTO*: a clinic-based study[6] aimed at better understanding the natural history of AD, dementia, and related diseases. Between 2011 and 2014, 2,323 individuals presenting either recently diagnosed MCI or isolated cognitive complaints were enrolled in 26 memory centers in France. This study was performed in accordance with the guidelines of the Declaration of Helsinki. The MEMENTO study protocol has been approved by the local ethics committee (Comité de Protection des Personnes Sud-Ouest et Outre Mer III; approval number 2010-A01394-35). All the participants provided written informed consent. *The CNRMAJ-Rouen study*[7]: early onset AD patients (n = 870). The patients or their legal guardians provided written informed consent. This study was approved by the ethics committee of CPP Ile de France II.

Italy: The AD cases and controls were collated through Italy in different centers: Brescia, Cagliari, Florence, Milan, Rome, Pertugia, San Giovani Rotondo and Torino. AD cases were diagnosed according to DSM III-R,IV and NINCDS–ADRDA criteria. Controls were defined a minima as subjects without DMS-III-R dementia criteria and with integrity of their cognitive functions (MMS>25).

Spain: The Dementia Genetic Spanish Consortium (DEGESCO) is a national consortium comprising 23 research centers and hospitals across the country, that holds the institutional coverage of The Network Center for Biomedical Research in Neurodegenerative Diseases (CIBERNED). Created in 2013, DEGESCO's objective is the promotion and conduction of genetic studies aimed at understanding the genetic architecture of neurodegenerative dementias in the Spanish population and participates in coordinated actions in national and international frameworks. All DNA samples are in compliance with the Law of Biomedical Research (Law 14/2007) and the Royal Decree on Biobanks (RD 1716/2011). Patients included in the present study met clinical criteria for probable or possible disease established by the National Institute of Neurological and Communication Disorders and Stroke and the Alzheimer Disease and Related Disorders Association (NINCDS-ADRDA). Cognitively healthy controls were unrelated individuals who had a documented MMSE in the normal range. Contributing centers in the France node genotyping were Centro de Biología Molecular Severo Ochoa (CSIC-UAM (Madrid), the Institute Biodonostia, University of Basque Contry (EHU-UPV, San Sebastián), Institut de Biomedicina de Valencia CSIC (València), and Sant Pau Biomedical Research Institute (Barcelona).

Sweden: *Upsala.* The Swedish AD patients were ascertained at the Memory Disorder Unit at Uppsala University Hospital. For all patients, the diagnosis was established according to the National Institute on Neurological Disorders and Stroke, and the Alzheimer's Disease and Related Disorders Association (NINDS-ADRDA) guidelines[8]. Healthy control subjects were recruited from the same geographic region following advertisements in local newspapers and displayed no signs of dementia upon Mini Mental State Examination (MMSE). *Swedish National Study on Aging and Care in Kungsholmen (SNAC-K)* data was collected. The original SNAC-K population consisted of 4590 living and eligible persons who lived on the island of Kungsholmen in Central Stockholm, belonged to pre-specified age strata, and were randomly selected to take part in the study. Between 2001 and 2004, 3363 persons participated in the baseline assessment. They belonged to the age cohorts 60, 66, 72, 78, 81, 84, 87, 90, 93, and 96 years and 99 years and older. The examination consists of three parts: a nurse interview, a medical examination, and a neuropsychological testing session. Altogether, the examination takes about six hours. The participants are reexamined each time they reach the next age cohort. All parts of the SNAC-K project have been approved by the ethical committee at Karolinska Institutet or the regional ethical review board. Informed consent was collected from all the participants or, if the person was severely cognitively impaired, from their next of kin.

The UK: *MRC*. The sample set comprises individuals with AD and healthy controls recruited across the MRC Centre for Neuropsychiatric Genetics and Genomics, Cardiff University, Cardiff, UK; Institute of Psychiatry, London, UK; University of Cambridge,

Cambridge, UK. The collection of the samples was through multiple channels, including specialist NHS services and clinics, research registers and Join Dementia Research (JDR) platform. The participants were assessed at home or in research clinics along with an informant, usually a spouse, family member or close friend, who provided information about and on behalf of the individual with dementia. Established measures were used to ascertain the disease severity: Bristol activities of daily living (BADL), Clinical Dementia Rating scale (CDR), Neuropsychiatric Inventory (NPI) and Global Deterioration Scale (GIDS). Individuals with dementia completed the Addenbrooke's Cognitive Examination (ACE-r), Geriatric Depression Scale (GeDS) and National Adult Reading Test (NART) too. Control participants were recruited from GP surgeries and by means of self-referral (including existing studies and Joint Dementia Research platform). For all other recruitment, all AD cases met criteria for either probable (NINCDS-ADRDA, DSM-IV) or definite (CERAD) AD. All elderly controls were screened for dementia using the Mini Mental State Examination (MMSE) or ADAS-cog, were determined to be free from dementia at neuropathological examination or had a Braak score of 2.5 or lower. Control samples were chosen to match case samples for age, gender, ethnicity and country of origin. Informed consent was obtained for all study participants, and the relevant independent ethical committees approved study protocols. *SOTON, University of Southampton, Southampton, UK*. All AD cases met criteria for either probable (NINCDS-ADRDA, DSM-IV) or definite (CERAD) AD. All elderly controls were screened for dementia using the MMSE or ADAS-cog, were determined to be free from dementia at neuropathological examination or had a Braak score of 2.5 or lower. *Nottingham and Manchester*, *University of Nottingham, Nottingham, UK and Manchester Brain Bank*. All AD cases met criteria for either probable (NINCDS-ADRDA, DSM-IV) or definite (CERAD) AD. All elderly controls were screened for dementia using the MMSE or ADAS-cog, were determined to be free from dementia at neuropathological examination or had a Braak score of 2.5 or lower. *KCL, London Neurodegenerative Diseases Brain Bank*. All AD cases met criteria for either probable (NINCDS-ADRDA, DSM-IV) or definite (CERAD) AD. All elderly controls were screened for dementia using the MMSE or ADAS-cog, were determined to be free from dementia at neuropathological examination or had a Braak score of 2.5 or lower. *PRION,* All AD cases met criteria for either probable (NINCDS-ADRDA, DSM-IV) or definite (CERAD) AD. All elderly controls were screened for dementia using the MMSE or ADAS-cog, were determined to be free from dementia at neuropathological examination or had a Braak score of 2.5 or lower. *CFAS Wales*, The Cognitive Function and Ageing Study Wales (CFAS-Wales) is a longitudinal population-based study of people aged 65 years and over in rural and urban areas of Wales that aims to investigate physical and cognitive health in older age and examine the interactions between health, social networks, activity, and participation. Individuals aged 65 years and over were randomly sampled from general medical practice lists between 2011 and 2013, stratified by age to ensure equal numbers in two age groups, 65-74 years and 75 and over. The baseline sample included 3593 older people and included those living in care homes as well as those living at home. Those who provided written consent to join the study were interviewed in their own homes by trained interviewers and could choose to have the interview conducted through the medium of either English or Welsh. Participants were followed up 2 years later. All AD cases met criteria for either probable (NINCDS-ADRDA, DSM-IV) or definite (CERAD) AD. All elderly controls were screened for dementia using the MMSE or CAMCOG, and were determined to be free from dementia. *UCL-DRC*. the UCL Alzheimer's disease cohort of the Dementia Research Centre (UCL - EOAD DRC) included patients seen at the Cognitive Disorders Clinics at The National Hospital for Neurology and Neurosurgery (Queen Square), or affiliated hospitals. Individuals were assessed clinically and diagnosed as having probable Alzheimer's disease based on contemporary clinical criteria in use at the time, including imaging and neuropsychological testing where appropriate.

**EADB-Germany**

In the German node, samples were collected from seven countries (11 centers/studies) and after QCs, we obtained 4,159 AD cases and 4,545 controls. All these samples were genotyped at Life&brain (Bonn, Germany).

Germany: *The VOGEL study*: The VOGEL study is a prospective, observational, long-term follow-up study with three time points of investigation within 6–8 years. This cohort includes dementia and healthy subjects. Residents of the city of Würzburg born between 1936 and 1941 were recruited. Every participant underwent physical, psychiatric, and laboratory examinations and performed intense neuropsychological testing as well as VSEP and NIRS according to the published procedures. A total of 604 subjects were included. *The Heidelberg/Mannheim memory clinic sample:* This cohort includes 61 subjects from whom 40 MCI patients were recruited and assessed between 2012 and 2016. Some of those patients converted to dementia by AD or other dementias. *The PAGES study:* This study includes 301 subjects. AD patients were recruited at the memory clinic of the Department of Psychiatry, University of Munich, Germany. Participants in whom dementia associated with AD was diagnosed fulfilled the criteria for probable AD according to the NINCDS–ADRDA. The control group included participants who were randomly selected from the general population of Munich. Controls who had central nervous system diseases or psychotic disorders or who had first-degree relatives with psychotic disorders were excluded. *The Technische Universität München study:* This cohort includes 359 healthy, AD, and other dementias patients recruited from the Centre for Cognitive Disorders. All the participants provided written informed consent. A biobank was submitted to the ethics committee of the Technical University of Munich, School of Medicine (Munich, Germany), which raised no objections and approved the biobank (reference number 347-14). *The Göttingen Universität study:* This study includes 111 in- and outpatients with a healthy or AD dementia status from the Department of Psychiatry of the University of Göttingen. The study's ethical statement was provided locally at the Göttingen University Medical Centre. *The German Dementia Competence Network (DCN) cohort:* Individuals from the DCN cohort were recruited from 14 university hospital memory clinics across Germany between 2003 and 2005[9]. The study was approved by the respective ethics committees, and written informed consent was obtained from all the participants prior to inclusion. *The German Study on Aging, Cognition, and Dementia (AgeCoDe):* The AgeCoDe study is a general practice (GP) registry-based longitudinal study in elderly individuals that recruited patients aged 75 years and above in six German cities from 2003 to 2004[10]. The study was approved by the respective ethics committees, and written informed consent was obtained from all the participants prior to inclusion.

Greece: *the HELIAD study*, comprising 49 AD cases and 1,150 controls. HELIAD is a population-based, multidisciplinary, collaborative study designed to estimate, in the Greek population over the age of 64 years, the prevalence and incidence of MCI, AD, other forms of dementia, and other neuropsychiatric conditions of aging and to investigate associations between nutrition and cognitive dysfunction or age-related neuropsychiatric diseases. The participants were selected through random sampling from the records of two Greek municipalities, Larissa and Marousi. All the participants signed informed consent in Greek.

Portugal: *the Lisbon study* from Portugal, totaling 78 AD cases and 74 controls. This cohort was recruited in 2008–2009 to investigate the connections between oxidative stress and lipid dyshomeostasis in AD. The project includes 190 subjects and was approved by the local ethics committee, and all the participants provided written informed consent. This study includes healthy and dementia-by-AD subjects.

Spain: Those samples are part of DEGESCO. DEGESCO Centers from whom DNA samples were genotyped in the German node (1,778 cases and 470 controls) were the Alzheimer Research Center and Memory Clinic, Fundació ACE, Institut Català de Neurociències Aplicades (Barcelona), the Neurology Service at University Hospital Marqués de Valdecilla (Santander), the Alzheimer's disease and other cognitive disorders, Neurology Department, at Hospital Clínic, IDIBAPS (Barcelona), the Molecular Genetics Laboratory, at the Hospital Universitario Central de Asturias (Oviedo), and Fundació Docència i Recerca

Mútua de Terrassa and Movement Disorders Unit, Department of Neurology, University Hospital Mútua de Terrassa (Barcelona).

Switzerland: Two datasets from Switzerland and Austria were combined, totaling 182 AD cases and 388 controls. *The Lausanne study*: This study includes 137 community-dwelling participants aged 55+ years with cognitive impairment (memory clinic patients with MCI, dementia) or normal cognition (recruited by advertisement, word of mouth). The study's ethical statement was provided locally at the Department of Psychiatry, Geneva University Centre, Switzerland. *The VITA study:* This is a longitudinal study of 606 individuals (Vienna, Austria) who were 75 years old in 2000, followed up every 30–90 months. This cohort includes dementia and healthy subjects. All the participants gave written informed consent. The study conformed to the latest version of the Declaration of Helsinki and was approved by the ethics committee of the City of Vienna, Austria

**EADB-Netherlands**

In the Dutch node, samples were collected from six organizations in the Netherlands and after QCs, we obtained 2,438 AD cases and 2,389 controls. All these samples were genotyped at the Erasmus Medical University (Rotterdam, The Netherlands). The Medical Ethics Committee (METC) of the local institutes approved the studies. All the participants and/or their legal guardians gave written informed consent for participation in the clinical and genetic studies. Samples from the following institutes were included. 1) *Erasmus Medical Center:* most individuals were selected from population studies from the epidemiology department and accounted for most of the controls, while a smaller subset of samples originated from the neurology department, where AD was diagnosed according to the National Institute of Neurological and Communicative Disorders and Stroke-Alzheimer's Disease and Related Disorders Association (NINCDS-ADRDA) criteria for AD[11]. 2) *The Amsterdam Dementia Cohort (ADC)*[12]: This cohort comprises patients who visit the memory clinic of the VU University Medical Centre, the Netherlands. The diagnosis of probable AD is based on the clinical criteria formulated by the NINCDS–ADRDA and based on the NIA–AA. Diagnosis of MCI was made according to Petersen and NIA-AA. Controls presented with subjective cognitive decline at the memory clinic, but performed within normal limits on all clinical investigations. 3) *The 100-Plus study:* This study includes Dutch-speaking individuals who (i) can provide official evidence for being aged 100 years or older, (ii) self-report to be cognitively healthy, which is confirmed by a proxy, (iii) consent to the donation of a blood sample, (iv) consent to (at least) two home visits from a researcher, and (v) consent to undergo an interview and neuropsychological test battery[13]. 4) *Parelsnoer Institute:* a collaboration between 8 Dutch University Medical Centers in which clinical data and biomaterials from patients suffering from chronic diseases (so called "Pearls") are collected according to harmonized protocols. The Pearl Neurodegenerative Diseases[14] includes individuals diagnosed with dementia, mild cognitive impairment, and controls with subjective memory complaints. 5) *The Netherlands Brain Bank:* a non-profit organization that collects human brain tissue of donors with a variety of neurological and psychiatric disorders, but also of non-diseased donors. A clinical diagnosis of AD is based on the clinical criteria of probable AD[8,15]. The selected AD patients for this study all received a definitive diagnosis which was based on autopsy. 6) *Maastricht University Medical Center*: a subset of individuals that were referred to the memory clinic for cognitive complaints were included if they participated in the BioBank-Alzheimer Centrum Limburg (BB-ACL)[16]. Diagnosis of MCI was made according to the criteria of Petersen, and diagnosis of AD-type dementia was made according to the criteria of the DSM-4[17], and the NINCDS-ADRDA[8].

**EADB-Australia**

The Sydney MAS study: a longitudinal study investigating MCI, related syndromes, and age-related cognitive change. Older adults (70–90 years old) were randomly recruited from the community in Sydney, Australia (n = 1,037). An extensive interview was undertaken and questionnaire data collected, including demographics, cognitive performance, and medical history. The majority of participants provided blood samples for genetic analysis.

Neuroimaging was performed on a subset of participants. Ethics approval for the study was provided by the ethics committee of the University of New South Wales and the Illawarra Area Health Service Human Research Ethics Committee. All the participants provided written informed consent to join the study. More information is provided in Sachdev et al[18]. In our study, there were 43 AD cases and 215 controls. Due to the low sample size, the study was not considered in the meta-analysis. However, samples from Sydney MAS study were included in the evaluation of the association of the polygenic risk score with conversion to all-dementia and AD dementia.

***GR@ACE***

The GR@ACE study[19] recruited Alzheimer's disease (AD) patients from Fundació ACE, Institut Català de Neurociències Aplicades (Catalonia, Spain), and control individuals from three centers: Fundació ACE (Barcelona, Spain), Valme University Hospital (Seville, Spain), and the Spanish National DNA Bank–Carlos III (University of Salamanca, Spain) (http://www.bancoadn.org). Additional cases and controls were obtained from dementia cohorts included in the Dementia Genetics Spanish Consortium (DEGESCO)[20]. At all sites, AD diagnosis was established by a multidisciplinary working group—including neurologists, neuropsychologists, and social workers—according to the DSM-IV criteria for dementia and the National Institute on Aging and Alzheimer's Association's (NIA–AA) 2011 guidelines for diagnosing AD. In our study, we considered as AD cases any individuals with dementia diagnosed with probable or possible AD at any point in their clinical course.
Genotyping was conducted using the Axiom 815K Spanish biobank array (Thermo Fisher) at the Spanish National Centre for Genotyping (CeGEN, Santiago de Compostela, Spain). The genotyping array not only is an adaptation of the Axiom biobank genotyping array but also contains rare population-specific variations observed in the Spanish population.

***The Rotterdam Study***

The Rotterdam Study is a prospective population-based middle-aged and elderly cohort that started in 1990 in the district of Ommoord, in Rotterdam, The Netherlands. The study includes 14,926 participants and has three subcohorts[21]. At start of the study, all inhabitants of the district of Ommoord who were aged 55 years and older were invited to participate. At baseline, in 1990-1993, of the 10,215 invited inhabitants, 7,983 agreed to participate in the baseline examination (response rate 78%). In 2000, the cohort was extended with 3,011 participants (67% of invitees). This extension consisted of all persons living in the study district who had become 55 years and older or had moved into the study district. A second extension was initiated in 2006, in which 3,932 participants (65% of invitees) who were 45 years and older were included. Study rounds consist of a home interview and visits with extensive investigations at the dedicated research centre. Rounds are repeated every 4-6 years. Participants are continuously monitored for diseases and mortality through linkage of the medical records from the general practitioners and municipality records. The Rotterdam Study has been approved by the Medical Ethics Committee of the Erasmus MC and by the Ministry of Health, Welfare and Sport of The Netherlands. All participants provided written informed consent to participate in the study and to obtain information from their treating physicians.
A total of 11,496 participants from the three subcohorts who were genotyped passed genotyping quality control (92% of all subjects with genotyping)[22]. Exclusion criteria were a call rate <98%, Hardy–Weinberg p-value $<10^{-6}$, minor allele frequency <0.01%, excess autosomal heterozygosity >0.336, sex mismatch, and outlying identity-by-state clustering estimates. Imputations were performed using the Haplotype Reference Consortium (HRC) panel[23].
Dementia ascertainment involved cognitive screening at the study research centre. We further assessed individuals with a Mini-mental state examination (MMSE) score <26 or Geriatric Mental State Schedule organic level >0[24], by administering the Cambridge Mental Disorders of the Elderly Examination by a research physician. We also interviewed spouses or informants. A consensus panel headed by a consultant neurologist established the final

diagnosis according to standard criteria. We studied the outcomes of all-cause dementia (DSM-III-R), and Alzheimer's disease (NINCDS–ADRDA). For the assessment of dementia, and type of dementia, the latest follow-up information with available data was used to determine the disease state. Follow-up for dementia was near complete until 1st January 2016. Within this period, participants were censored at date of dementia diagnosis, death, or loss to follow-up. For this study, we included participants from the first, second and third subcohort (N=11,070).

***European Alzheimer's Disease Initiative (EADI) Consortium***

All the 2,400 Alzheimer's disease cases were ascertained by neurologists from Bordeaux, Dijon, Lille, Montpellier, Paris and Rouen. Clinical diagnosis of probable Alzheimer's disease was established according to the DSM-III-R and NINCDS-ADRDA criteria. Controls were selected from the 3C Study[25]. This cohort is a population-based, prospective study of the relationship between vascular factors and dementia. It has been carried out in three French cities: Bordeaux (southwest France), Montpellier (southeast France) and Dijon (central eastern France). A sample of non-institutionalized, over-65 subjects was randomly selected from the electoral rolls of each city. Between January 1999 and March 2001, 9,686 subjects meeting the inclusion criteria agreed to participate. Following recruitment, 392 subjects withdrew from the study. Thus, 9,294 subjects were finally included in the study (2,104 in Bordeaux, 4,931 in Dijon and 2,259 in Montpellier). The AD status was defined based on 12 years follow-up for Dijon participants, 14-15 years follow-up for Montpellier participants and 17-18 years follow-up for Bordeaux participants. The Alzheimer's disease cases from 3C were included as cases in the EADI discovery dataset and the other individuals were retained as controls. Genomic DNA samples of 7,200 individuals were transferred to the French Centre National de Génotypage (CNG). First stage samples that passed DNA quality control were genotyped with Illumina Human 610-Quad BeadChips.

***Genetic and Environmental Risk in AD (GERAD) Consortium/Defining Genetic, Polygenic, and Environmental Risk for Alzheimer's Disease (PERADES) Consortium***

The GERAD/PERADES sample comprises 3,177 Alzheimer's disease cases and 7,277 controls with available age and gender data[26]. Cases and elderly screened controls were recruited by the Medical Research Council (MRC) Genetic Resource for Alzheimer's disease (Cardiff University; Institute of Psychiatry, London; Cambridge University; Trinity College Dublin), the Alzheimer's Research Trust (ART) Collaboration (University of Nottingham; University of Manchester; University of Southampton; University of Bristol; Queen's University Belfast; the Oxford Project to Investigate Memory and Ageing (OPTIMA), Oxford University); Washington University, St Louis, United States; MRC PRION Unit, University College London; London and the South East Region Alzheimer's disease project (LASER-AD), University College London; Competence Network of Dementia (CND) and Department of Psychiatry, University of Bonn, Germany; the National Institute of Mental Health (NIMH) Alzheimer's disease Genetics Initiative. 6129 population controls were drawn from large existing cohorts with available GWAS data, including the 1958 British Birth Cohort (1958BC) (http://www.b58cgene.sgul.ac.uk), the KORA F4 Study and the Heinz Nixdorf Recall Study. All Alzheimer's disease cases met criteria for either probable (NINCDS-ADRDA, DSM-IV) or definite (CERAD) Alzheimer's disease. All elderly controls were screened for dementia using the MMSE or ADAS-cog, were determined to be free from dementia at neuropathological examination or had a Braak score of 2.5 or lower. Genotypes from all cases and 4617 controls were previously included in the AD GWAS by Harold and colleagues[26]. Genotypes for the remaining 2660 population controls were obtained from WTCCC2.

***The Norwegian DemGene Network***

This is a Norwegian network of clinical sites collecting cases from memory clinics based on a standardized examination of cognitive, functional, and behavioral measures and data on the progression of most patients. The Norwegian DemGene Network includes 2,224 cases and 3,089 healthy controls from different studies described elsewhere[27]. The cases were diagnosed according to recommendations from the NIA–AA, the NINCDS–ADRDA criteria, or the ICD-10 research criteria. The controls were screened with a standardized interview and cognitive tests. Additional controls from blood donors of the Oslo University Hospital, Ulleval Hospital, were included (n=4992, age between 18-65 years, 48% female). They were thoroughly screened for diseases and medication, and provided blood for DNA analysis, in line with approval from the Regional Committee for Medical and Health Research Ethics. Individuals from the DemGene study and blood donors were genotyped using either the Human Omni Express-24 v1.1 chip (Illumina Inc., San Diego, CA) or the DeCodeGenetics_V1_20012591_A1 chip at deCODE Genetics (Reykjavik, Iceland).

***The Neocodex–Murcia study (NxC)***

This study includes 324 sporadic AD patients and 754 controls of unknown cognitive status from the Spanish general population collected by Neocodex[28,29]. AD patients were diagnosed as having possible or probable AD in accordance with the NINCDS–ADRDA criteria.

***The Copenhagen City Heart Study (CCHS)***

CCHS is a prospective study of the Danish general population initiated in 1976-78 with follow-up examinations in 1981-83, 1991-94, 2001-03, and 2011-13. Individuals were selected randomly based on the national Danish Civil Registration System to reflect the adult Danish population aged 20-100. Data were obtained from a self-administered questionnaire reviewed together with an investigator at the day of attendance, a physical examination, and from blood samples including DNA extraction. Genotypes were available on 8,118 individuals from the 1991-94 and 2001-03 examinations following genotyping on the Illumina Metabochip and/or the Illumina HumanExome.

***Bonn studies***

DietBB: The DietBB sample included in this GWAS is a subsample extracted from the AgeCoDe cohort in the context of an ongoing genome-wide methylation analysis for dementia. In addition to methylation, the DietBB samples has genome-wide genotype data which was included in this study. The German study on aging, cognition and dementia (AgeCoDe)[10,30] study is a general practice (GP) registry-based longitudinal study in elderly individuals on the identification of predictors of dementia. Participants were recruited in six German cities (Bonn, Dusseldorf, Hamburg, Leipzig, Mannheim, and Munich) with a total of 138 GPs connected to the study sites. The inclusion criteria for this study were an age of 75 years and older, absence of dementia according to GP judgment, and at least one contact with the GP within the past 12 months. Exclusion criteria were GP consultations by home visits only, living in a nursing home, severe illness with an anticipated fatal outcome within 3 months, language barrier, deafness or blindness, and lack of ability to provide informed consent. Baseline recruitment was performed in 2002 and 2003. The study was approved by the local ethical committees of the Universities of Bonn, Hamburg, Dusseldorf, Heidelberg/Mannheim, and Leipzig, and the Technical University of Munich. A total of 3327 subjects provided informed consent for participation after being provided with a complete description of the study protocol. The study assessments were performed by trained interviewers at the subjects' home. Seventy individuals were excluded after baseline interview because of the presence of dementia according to standard assessment, and 40 subjects were excluded for age less than 75 years. In AgeCoDe, dementia was diagnosed according to the criteria set of DSM-IV in a consensus conference with the interviewer and an experienced geriatrician or geriatric psychiatrist. The etiological diagnosis of dementia in AD was established according to the National Institute of Neurological and Communicative Diseases and Stroke/Alzheimer's Disease and Related Disorders Association (NINCDS-ADRDA) criteria for probable AD[8]. Mixed dementia was diagnosed in cases of cerebrovascular events without temporal relationship to cognitive decline. Mixed dementia and dementia in AD were combined. Dementia diagnosis in subjects who were not interviewed personally was based on the Global Deterioration Scale[31] (score ≥4 points). In these cases, an etiological diagnosis was established only if the information provided was sufficient to judge etiology according to the criteria just described. For DietBB, cohort participants where included if they were dementia-free at baseline and available biomaterial for DNA analysis is available. This criterion led to the selection of 320 participants. In 120 of these participants, dementia of the AD-type occurred at any follow up. The additional 200 remain free of dementia until last follow up of AgeCoDe.

Bonn OMNI cohort: the Bonn OMNI cohort consists of AD patients and controls derived from a larger German GWAS cohort which was recruited from the following three sources: (i) the German Dementia Competence Network; (ii) the German study on Aging, Cognition, and Dementia in primary care patients (AgeCoDe); and (iii) the interdisciplinary Memory Clinic at the University Hospital of Bonn. The control sample comprised of individuals from the population-based study Heinz Nixdorf Recall (HNR) study cohort. This sample was previously used for replication in Lambert et al.[32]. *The German study on aging, cognition and dementia (AgeCoDe)*: see description above. *The German competence network cohort (DCN)*: The DCN cohort includes 1,095 patients with mild cognitive impairment (MCI) and 648 cases with mild Alzheimer's disease (AD) clinical dementia syndrome that were recruited from 14 university hospital memory clinics across Germany between 2003 and 2005[9]. Exclusion criteria were substance abuse or dependence, insufficient German language skills, multi-morbidity, comorbid condition with excess mortality, circumstances that would have made regular attendance at follow-up visits questionable and lack of an informant. The diagnosis of mild dementia according to ICD-10 criteria required a decline of cognitive ability (at least 1 SD) from a previous level in at least 2 domains as evidenced by age-corrected standardized tests, impairment in activities of daily living (i.e. B-ADL > 6), changes in personality, drive, social behavior or control of emotion but no clouding of consciousness. These changes must have persisted for at least 3 months. The etiological diagnosis of AD was assigned according to NINCDS-ADRDA criteria[8]. *Memory clinic Bonn*: The interdisciplinary Memory Clinic of the Department of Psychiatry and Department of Neurology at the University Hospital in Bonn provided further patients. Diagnoses were assigned according the NINCDS/ADRDA criteria[8] and on the basis of clinical history, physical examination, neuropsychological testing (using the CERAD neuropsychological battery, including the MMSE), laboratory assessments, and brain imaging. *Control sample:* In the Heinz Nixdorf Recall (Risk Factors, Evaluation of Coronary Calcification, and Lifestyle) study, participants were randomly sampled in three cities in Germany. The study design has previously been described[33,34]. Briefly, 4814 participants aged 45 to 75 years were enrolled between 2000 and 2003 (t0, baseline). Cognitive performance of participants was evaluated at follow up scheduled 5 years after baseline (t1, n = 4157, 2005–2008) and then again at follow up 5 years after t1 (t2, n = 3087, 2010–2015). Controls sample was selected if participant did not present cognitive impairment as reported at the last available evaluation. Cognitive evaluation has been described extensively previously[35,36]. Herein, cognitive impairment at t1 was defined as a performance of one standard deviation (SD) below the age- and education-adjusted mean except for the clock-drawing test, where a performance ≥3 was rated as impaired (for a detailed description, see the study by Winkler et al.[35]). The study was approved by the University of Duisburg-Essen Institutional Review Board and followed established guidelines of good epidemiological practice.

***UK Biobank***

AD/Dementia cases were extracted from UK Biobank (data release Feb 2020) self-report, ICD10 diagnoses and ICD10 cause of death. Proxy AD/Dementia cases included all participants who reported at least one biological relative (parents and siblings) affected with dementia either at baseline or follow up. Participants who answered “Do not know” or “Prefer not to answer” were excluded from analyses. Individual who did not report dementia or any family history of dementia were used as controls. Our analysis included 2,447 diagnosed cases, 46,828 proxy cases of dementia and 338,440 controls.

##### 1.2. Stage II samples

***Alzheimer's Disease Genetics Consortium (ADGC)***

The ADGC dataset comprises subjects from 35 datasets including two waves of the Adult Changes in Thought (ACT) cohort study [ACT1/ACT2]; ten waves of cases and cognitively normal controls from the National Institute on Aging (NIA) Alzheimer Disease Centers (ADCs); the Alzheimer Disease Neuroimaging Initiative (ADNI); the Biomarkers of Cognitive Decline Among Normal Individuals (BIOCARD) Cohort; two waves of the Religious Orders Study/Memory and Aging Project (ROSMAP1-2) and the Chicago Health and Aging Project (CHAP) cohort studies at Rush University; the Einstein Aging Study (EAS); the Multi-Site Collaborative Study for Genotype-Phenotype Associations in Alzheimer's Disease (GenADA) Study by GlaxoSmithKline; Mayo Clinic Jacksonville (MAYO) and Rochester (RMAYO) case-control datasets; the Multi-Institutional Research in Alzheimer's Genetic Epidemiology (MIRAGE) study; the NIA Late-Onset Alzheimer's Disease (LOAD) Family Study (NIA-LOAD); the Netherlands Brain Bank (NBB) case-control dataset; the Oregon Health and Science University (OHSU) case-control dataset; the Pfizer case-control dataset; the Texas Alzheimer's Research and Care Consortium (TARCC) dataset; the Translational Genomics Research Institute series 2 (TGEN2) dataset; the University of Miami (UM)/ Case Western Reserve University (CWRU)/ Mt. Sinai School of Medicine (MSSM) and UM/CWRU/TARCC wave 2 datasets [UM/CWRU/MSSM and UM/CWRU/TARCC2]; the Universitatsklinikum Saarlandes (UKS) case-control dataset; the University of Pittsburgh (UPITT) case-control dataset; Washington University (WASHU) wave 1 and 2 case-control datasets [WASHU1/WASHU2]; and the Washington Heights-Inwood Community Aging Project (WHICAP) study datasets.

Descriptions of the ACT1, ADC waves 1-7, ADNI, BIOCARD, CHAP, EAS, GenADA, MAYO, MIRAGE, NBB, NIA-LOAD, OHSU, PFIZER, RMAYO, ROSMAP1, ROSMAP2, TARCC, TGEN2, UKS, UM/CWRU/MSSM, UM/CWRU/TARCC2, UPITT, WASHU1, WASHU2, and WHICAP cohorts have been provided in previous ADGC and IGAP studies[32,37–41]. Here we update descriptions of these studies, where applicable, and provide descriptions for ACT2, ADC wave 8-10, as well as the Combined Small Datasets Collection (CSDC). The CSDC is a harmonized collection of merged small datasets (comprising the existing datasets of ACT2, BIOCARD, CHAP2, EAS, NBB, RMAYO, ROSMAP2, and WASHU2) used in common variant analyses to deal with small case and control counts within the individual studies. All analyses were restricted to individuals of European ancestry. All subjects were recruited under protocols approved by the appropriate Institutional Review Boards (IRBs).

<u>ACT1/ACT2:</u> The ACT cohort is an urban and suburban elderly population from a stable HMO that includes 2,581 cognitively intact subjects age ≥ 65 who were enrolled between 1994 and 1998[42,43]. An additional 811 subjects were enrolled in 2000-2002 using the same methods except oversampling clinics with more minorities. More recently, a Continuous Enrollment strategy was initiated in which new subjects are contacted, screened, and enrolled to keep 2,000 active at-risk person-years accruing in each calendar year. This resulted in an enrollment of 4,146 participants as of May 2009. All clinical data are reviewed at a consensus conference. Dementia onset is assigned half-way between the prior biennial and the exam that diagnosed dementia. A waiver of consent was obtained from the IRB to enroll deceased ACT participants. In total, ACT contributed data on 553 individuals with probable or possible Alzheimer's disease (70 with autopsy-confirmation) and on 1,579 cognitively normal elders (CNEs, 155 with autopsy-confirmation) who were included in the analyses, with 2,103 cases/1,571 CNEs in the first wave (ACT1) and 29 cases/8 CNEs in the second wave (ACT2).

<u>NIA ADC Samples (ADC1-10)</u>: The NIA ADC cohort included subjects ascertained and evaluated by the clinical and neuropathology cores of the 32 NIA-funded ADCs. Data collection is coordinated by the National Alzheimer's Coordinating Center (NACC). NACC coordinates collection of phenotype data from the 32 ADCs, cleans all data, coordinates implementation of definitions of Alzheimer's disease cases and controls, and coordinates collection of samples. The complete ADC cohort consists of 3,311 autopsy-confirmed and

2,889 clinically-confirmed Alzheimer's disease cases, and 247 cognitively normal elders (CNEs) with complete neuropathology data who were older than 60 years at age of death, and 3,687 living CNEs evaluated using the Uniform dataset (UDS) protocol[44,45] who were documented to not have mild cognitive impairment (MCI) and were between 60 and 100 years of age at assessment. Based on the data collected by NACC, the ADGC Neuropathology Core Leaders Subcommittee derived inclusion and exclusion criteria for Alzheimer's disease and control samples. All autopsied subjects were age ≥ 60 years at death. Based on the data collected by NACC, the ADGC Neuropathology Core Leaders Subcommittee derived inclusion and exclusion criteria for Alzheimer's disease and control samples. All autopsied subjects were age ≥ 60 years at death. Alzheimer's disease cases were demented according to NINCDS-ADRDA/DSMIV-V criteria or Clinical Dementia Rating (CDR) ≥ 137[8,11]. Neuropathologic stratification of cases followed NIA/Reagan criteria explicitly or used a similar approach when NIA/Reagan criteria were coded as not done, missing, or unknown. Cases were intermediate or high likelihood by NIA/Reagan criteria with moderate to frequent amyloid plaques[46] and neurofibrillary tangle (NFT) Braak stage of III-VI[47,48]. Persons with Down's syndrome, non-Alzheimer's disease tauopathies and synucleinopathies were excluded. All autopsied controls had a clinical evaluation within two years of death. Controls did not meet NINCDS-ADRDA/DSMIV-V criteria for dementia, did not have a diagnosis of mild cognitive impairment (MCI), and had a CDR of 0, if performed. Controls did not meet or were low-likelihood Alzheimer's disease by NIA/Reagan criteria, had sparse or no amyloid plaques, and a Braak NFT stage of 0 – II. ADCs sent frozen tissue from autopsied subjects and DNA samples from some autopsied subjects and from living subjects to the ADCs to the National Cell Repository for Alzheimer's Disease (NCRAD). DNA was prepared by NCRAD for genotyping and sent to the genotyping site at Children's Hospital of Philadelphia. ADC samples were genotyped and analyzed in separate batches (waves 1-10). The ADC data used in the analyses (ADC1-10) consist of 6,292 cases and 4,980 CNEs in total.

ADNI: ADNI is a longitudinal, multi-site observational study including Alzheimer's disease, mild cognitive impairment (MCI), and elderly individuals with normal cognition assessing clinical and cognitive measures, MRI and PET scans (FDG and 11C PIB) and blood and CNS biomarkers. For this study, ADNI contributed data on 268 Alzheimer's disease cases with MRI confirmation of Alzheimer's disease diagnosis and 173 healthy controls with Alzheimer's disease-free status confirmed as of most recent follow-up. Alzheimer's disease subjects were between the ages of 55–90, had an MMSE score of 20–26 inclusive, met NINCDS-ADRDA criteria for probable Alzheimer's disease[8,11], and had an MRI consistent with the diagnosis of Alzheimer's disease. Control subjects had MMSE scores between 28 and 30 and a Clinical Dementia Rating of 0 without symptoms of depression, MCI or other dementia and no current use of psychoactive medications. According to the ADNI protocol, subjects were ascertained at regular intervals over 3 years, but for the purpose of our analysis we only used the final ascertainment status to classify case-control status. Additional details of the study design are available elsewhere[40,49,50].

BIOCARD: The BIOCARD study is supported by a grant jointly funded by the National Institute on Aging (NIA) and the National Institute of Mental Health (NIMH). The overarching goal of the BIOCARD Study is to identify biomarkers associated with progression from normal cognitive status to cognitive impairment or dementia, with a particular focus on Alzheimer's Disease. Please see Albert et al.[51] for a detailed description of the study. A total of 354 individuals were initially enrolled in the study. Recruitment was conducted by the staff of the Geriatric Psychiatry Branch (GPB) of the intramural program of the NIMH, beginning in 1995 and ending in 2005. The domains of information collected as part of the study include: cognitive testing, magnetic resonance imaging (MRI), cerebrospinal fluid (CSF), amyloid imaging (using PET-PiB), and blood specimens. Investigators at the Johns Hopkins University School of Medicine began evaluating participants in 2009, and subjects are seen annually. At each visit there are assessments of medical and cognitive status, as well as acquisition of MRI, CSF, PET-PiB, and blood. Each subject in the analyses received a consensus diagnosis by a team of neurologists, neuropsychologists, research nurses and research assistants of the BIOCARD Clinical Core at Johns Hopkins with diagnoses based on evidence of clinical or cognitive dysfunction (i.e., individuals with a CDR score > 0 and/or evidence of decline on cognitive testing). To the extent possible, this diagnosis did not use the cognitive test scores. In brief, (1) clinical data relating to the medical, neurologic and psychiatric status of the subject were examined, (2) reports of changes in cognition by the subject and other sources were examined, and (3) decline in cognitive performance was established. Cognitive test scores were used to: (1) determine whether the subject had become cognitively impaired, and (2) determine the likely etiology of such impairment. These diagnostic procedures are comparable to those implemented in the Alzheimer's Disease Centers (ADC) program, supported by the NIA. For this study, BIOCARD contributed data on 6 Alzheimer's disease cases and 112 healthy controls with Alzheimer's disease-free status confirmed as of most recent follow-up.

CHAP: CHAP is an on-going community based study of individuals from a geographically defined community of 3 neighborhoods in Chicago, Illinois (Morgan Park, Washington Heights, and Beverly), with 6,158 participants in the first phase of the study (78.7% overall; 80.5% of the blacks, 74.6% of the whites)[52]. Data were collected in cycles of approximately 3 years; each consisting of an in-home interview of all participants and clinical evaluation of a random, stratified sample. The baseline cycle measured disease prevalence and provided risk factor data prior to incident disease onset. A cohort of 3,838 persons free of Alzheimer's disease was identified; 729 persons were sampled for baseline clinical evaluation. Persons in the disease-free cohort had either good cognitive function at baseline, or if cognitive function was intermediate or poor, were free from Alzheimer's disease at the baseline clinical evaluation. This disease-free cohort was evaluated for incident disease after an average of 4.1 years. Sampling for incident clinical evaluation was based on age, sex, race, and change in cognitive function (i.e., stable or improved, small decline, or large decline). The sample set available in the ADGC for genetic analyses included 27 Alzheimer's disease cases and 144 persons free of Alzheimer's disease at time of last assessment. All subjects were age 65 years or older at last assessment.

EAS: Based at the Albert Einstein College of Medicine, the EAS is an ongoing community based cohort study of cognitive aging and Alzheimer's disease in the elderly which began over four decades ago. Please see Barzilai et al.[53] and Katz et al.[54] for details. The EAS cohort has employed systematic recruiting methods to reduce the selection biases that arise from clinic-based samples and to capture the racial diversity within the Bronx community. Since 1993, a total of 1,944 participants have been enrolled. Between 1993 and 2004, Health Care Financing Administration/Centers for Medicaid and Medicare Services (HCFA/CMS) rosters of Medicare eligible persons aged 70 and above were used to develop sampling frames of community residing participants in Bronx County. Since 2004, New York City Board of Elections registered voter lists for the Bronx have been used due to changes in policies for release of HCFA/CMS rosters. Individuals were mailed introductory letters regarding the study and were then telephoned to complete a brief screening interview. Eligible participants were at least 70 years of age, Bronx residents, non-institutionalized, and English speaking. Exclusion criteria included visual or auditory impairments that preclude neuropsychological testing, active psychiatric symptomatology that interfered with the ability to complete assessments, and non-ambulatory status. Written informed consent was obtained at the initial clinic visit. In-person evaluations were completed at baseline and at subsequent 12-month intervals. Functional status was assessed by the self-administered CERAD C1-ALT, a cognitive/functional impairment instrument, and the Instrumental Activities of Daily Living scale (IADL), a subscale on the Lawton Brody Activities of Daily Living Scale. The score on the IADL was based on 5 domains of function that were common to both elderly men and women. Scores for each domain were dichotomized as impaired vs. not impaired and then the domain scores were summed. If the participant agreed, an informant completed the CERAD C2-ALT, a cognitive/functional impairment instrument, and the Informant Questionnaire on Cognitive Decline in the Elderly (IQ-CODE) 14 forms. The standard neurological physical examination was adapted from the Unified Parkinson's Disease Rating Scale. The evaluation assessed the participant's memory for significant recent events in the news and personal events. The coherence and focus of responses, repetitiveness, and language were determined. When possible, informants were interviewed to ascertain whether they noted any cognitive changes in the participant, and to assess accuracy of the participant's responses. The neurologist also assessed each participant for abnormal behaviors, fluctuation in cognition, and history of sleep disturbance and visual/auditory hallucinations. The neurologist assigned an Hachinski Ischemic Score (HIS), the Clinical Dementia Rating (CDR), and provided a clinical impression of presence or absence of dementia. A diagnosis of dementia was based on standardized clinical criteria from the Diagnostic and Statistical Manual, Fourth Edition (DSM-IV) and required impairment in memory plus at least one additional cognitive domain, accompanied by evidence of functional decline. Diagnoses were assigned at consensus case conferences, which included comprehensive review of cognitive test results, relevant neurological signs and symptoms, and functional status. Memory impairment was defined as scores in the impaired range on any of the memory tests in the neuropsychological battery. (FCSRT ≤ 2430 or 1.5 standard deviations (SD) below the age-adjusted mean on Logical Memory) Functional decline was determined at case conference based on information from self or informant report, impairment score on the IADL Lawton Brody Scale, clinical evaluation, and informant questionnaires. Alzheimer's disease was diagnosed in participants with dementia meeting clinical criteria for probable or possible disease established by the NINCDS-ADRDA[8,11]. Incident dementia and Alzheimer's disease were diagnosed in persons free of dementia at baseline who met criteria at follow-up. A subset of individuals who participated in the clinical studies of the EAS came to autopsy, providing an important quality control for diagnostic accuracy. A clinical diagnosis of dementia had a positive predictive value (PPV) of 96% for significant pathology upon autopsy. A clinical diagnosis of possible or probable Alzheimer's disease had a PPV of 79% for the presence of NIA-Reagan intermediate or high likelihood Alzheimer type pathology based on an autopsy sample of 175. For this study, EAS contributed data on 9 Alzheimer's disease cases and 141 healthy controls with Alzheimer's disease-free status confirmed as of most recent follow-up.

GenADA: GenADA study data analyzed included 666 Alzheimer's disease cases and 712 CNEs ascertained from nine memory referral clinics in Canada between 2002 and 2005. Patients and CNEs were of non-Hispanic White (NHW) ancestry from Northern Europe. All patients with Alzheimer's disease satisfied NINCDS-ADRDA and DSM-IV criteria for probable Alzheimer's disease with Global Deterioration Scale scores of 3-7[8,11]. CNEs had MMSE test scores higher than 25 (mean 29.2 ± 1.1), a Mattis Dementia Rating Scale score of ≥ 136, a Clock Test without error, and no impairments on seven instrumental activities of daily living questions from the Duke Older American Resources and Services Procedures test. Data were collected under an academic-industrial grant from Glaxo-Smith-Kline, Canada by Principal Investigator P. St George-Hyslop. Detailed characteristics of this cohort have been described previously[55].

MAYO/RMAYO: All 671 cases and 1,279 controls consisted of NHW subjects from the United States ascertained at the Mayo Clinic. All subjects were diagnosed by a neurologist at the Mayo Clinic in Jacksonville, Florida or Rochester, Minnesota. The neurologist confirmed a Clinical Dementia Rating score of 0 for all controls; cases had diagnoses of possible or probable Alzheimer's disease made according to NINCDS-ADRDA criteria[8,11]. Autopsy-confirmed samples (221 cases, 216 CNEs) came from the brain bank at the Mayo Clinic in Jacksonville, FL and were evaluated by a single neuropathologist. In clinically-identified cases, the diagnosis of definite Alzheimer's disease was made according to NINCDS-ADRDA criteria. All Alzheimer's disease brains analyzed in the study had a Braak score of 4.0 or greater. Brains employed as controls had a Braak score of 2.5 or lower but often had brain pathology unrelated to Alzheimer's disease and pathological diagnoses that included vascular dementia, frontotemporal dementia, dementia with Lewy bodies, multi-system atrophy, amyotrophic lateral sclerosis, and progressive supranuclear palsy.

MIRAGE: The MIRAGE study is a family-based genetic epidemiology study of Alzheimer's disease that enrolled Alzheimer's disease cases and unaffected sibling controls at 17 clinical centers in the United States, Canada, Germany, and Greece (details elsewhere[56]), and contributed 1,229 subjects (491 Alzheimer's disease cases and 738 CNEs), a subset of the cases and controls that were incorporated into our prior studies[37,40] which met more stringent QC criteria for this study. Briefly, families were ascertained through a proband meeting the NINCDS-ADRDA criteria for definite or probable Alzheimer's disease[8,11]. Unaffected sibling controls were verified as cognitively healthy based on a Modified Telephone Interview of Cognitive Status score ≥ 86[57].

UM/CWRU/TARCC2: The UM/CWRU/TARCC2 sample included 256 cases and 189 controls from the University of Miami, Case Western Reserve University, and the Texas Alzheimer's Research Care Consortium (wave 2). All Alzheimer's disease cases had onset of disease symptoms after age 65 years and met NINCDS-ADRDA criteria for probable or possible Alzheimer's disease[8,11]. Controls were adjudicated to have MMSE scores greater than 28 and no clinically identified signs of cognitive impairment. Additional details of subject recruitment at these sites are described in the UM/CWRU/MSSM (formerly UM/VU/MSSM) and TARCC cohort descriptions in this supplement and elsewhere[37,39,58].

NIA-LOAD: The NIA LOAD Family Study[59] recruited families with two or more affected siblings with LOAD and unrelated, CNEs similar in age and ethnic background. A total of 1,819 cases and 1,969 CNEs from 1,802 families were recruited through the NIA LOAD study, NCRAD, and the University of Kentucky, with 1,798 cases and 1,568 CNEs included for analysis. One case per family was selected after determining the individual with the strictest diagnosis (definite > probable > possible LOAD). If there were multiple individuals with the strictest diagnosis, then the individual with the earliest age of onset was selected. The controls included only those samples that were neurologically evaluated to be normal and were not related to a study participant.

NBB: The Netherlands Brain Bank, which has been previously described elsewhere[58], is a department of the Netherlands Institute for Neuroscience, an institute of the Royal Netherlands Academy of Arts and Sciences. The NBB is a non-profit organization that collects human brain tissue from donors with a variety of neurological and psychiatric disorders and brain tissue from non-diseased donors, as well as anonymized summaries of donors' medical records to be made available for neuroscience research[60]. The sample set available in the ADGC for genetic analyses included 80 pathologically-confirmed Alzheimer's disease cases and 48 subjects free of Alzheimer's pathology at autopsy. All cases were age 65 years or older at time of diagnosis, and all controls were age 65 years or older at time of death.

OHSU: The OHSU dataset includes 132 autopsy-confirmed Alzheimer's disease cases and 153 deceased controls that were evaluated for dementia within 12 months prior to death (age at death > 65 years), which are a subset of the 193 cases and 451 controls examined in our previous study[40] meeting more stringent QC criteria in this study. Subjects were recruited from aging research cohorts at 10 NIA-funded ADC and did not overlap other samples assembled by the ADGC. A more extensive description of control samples can be found elsewhere[61].

Pfizer: The Pfizer sample collection comprises Alzheimer's disease cases taken from the Lipitor's Effect in Alzheimer's Disease (LEADe) trial, including subjects who converted to Alzheimer's disease after ascertainment as MCI, as well as 216 probable Alzheimer's disease subjects enrolled by PrecisionMed for a case-control study and 149 subjects from a Phase II trial (#A3041005) of CP-457920 (a selective α5 GABAA receptor inverse agonist) in Alzheimer's disease. Samples were collected from multiple clinical sites, and with appropriate IRB/ethics committee approvals at each individual site, with written and informed consent given by subjects for use in follow-up studies. All subjects were diagnosed with probable or possible Alzheimer's disease if they met NINCDS-ADRDA and/or DSM-IV criteria, and had Mini-Mental Status Exam (MMSE) scores < 25 at baseline[8,11]. The control group included subjects from two studies: 1) the PrecisionMed case-control study (#A9010012), which recruited elderly subjects free of neurological or psychiatric conditions, and 2) 999-GEN-0583-001, which obtained a reference population of cognitively, neurologically, and psychiatrically normal subjects. Controls have no neuropsychiatric conditions or diseases and had MMSE>27 at the time of enrollment. For Alzheimer's disease analysis, all cases with age-at-onset (AAO) less than 65 years were removed to exclude early-onset Alzheimer's disease subjects. All controls were re-matched with remaining cases according to gender, age (all controls are older than cases), and ethnicity (only individuals with NHW background were analyzed). The final Pfizer Alzheimer's disease case-control GWAS dataset included 696 cases and 762 controls. Cases from the PrecisionMed/ A3041005 and LEADe studies and age-matched controls were genotyped using the Illumina HumanHap550 array. *APOE* genotypes were determined from genotypes for rs429358 and rs7412 obtained using Taqman assays.

ROSMAP: ROSMAP are two community-based cohort studies. The ROS has been ongoing since 1993, with a rolling admission. Through July of 2010, 1,139 older nuns, priests, and brothers from across the United States initially free of dementia who agreed to annual clinical evaluation and brain donation at the time of death completed their baseline evaluation. The MAP has been on-going since 1997, also with a rolling admission. Through July of 2010, 1,356 older persons from across northeastern Illinois initially free of dementia who agreed to annual clinical evaluation and organ donation at the time of death completed their baseline evaluation. Details of the clinical and neuropathologic evaluations have been previously reported[62–64]. A total of 1,064 persons passed genotyping QC. Of these, 295 met clinical criteria for Alzheimer's disease at the time of their last clinical evaluation or time of death and met neuropathologic criteria for Alzheimer's disease for those on whom neuropathologic data were available, and 769 were without dementia or MCI at the time of their last clinical evaluation or time of death and did not meet neuropathologic criteria for Alzheimer's disease for those on whom neuropathologic data were available. A second wave of ROSMAP (referred to as ROSMAP2 in this study) included 59 persons who met clinical criteria for Alzheimer's disease at the time of their last clinical evaluation or time of death and met neuropathologic criteria for Alzheimer's disease for those on whom neuropathologic data were available, and 217 persons who were without dementia or MCI at the time of their last clinical evaluation or time of death and did not meet neuropathologic criteria for Alzheimer's disease for those on whom neuropathologic data were available.

TARCC: The TARCC is a collaborative Alzheimer's research effort directed and funded by the Texas Council on Alzheimer's Disease and Related Disorders (the Council), as part of the Darrell K Royal Texas Alzheimer's Initiative. Composed of Baylor College of Medicine (BCM), Texas Tech University Health Sciences Center (TTUHSC), University of North Texas Health Science Center (UNTHSC), the UT Southwestern Medical Center at Dallas (UTSW), University of Texas Health Science Center at San Antonio (UTHSCSA), Texas A&M Health Science Center (TAMHSC), and the University of Texas at Austin (UTA), this consortium was created to establish a comprehensive research cohort of well characterized subjects to address better diagnosis, treatment, and ultimately prevention of Alzheimer's disease[65]. The resulting prospective cohort, the Texas Harris Alzheimer's Research Study, contains clinical, neuropsychiatric, genetic, and blood biomarker data on more than 3,000 participants diagnosed with Alzheimer's disease, mild cognitive impairment (MCI), and cognitively normal individuals. Longitudinal data/sample collection and follow-up on participants occurs on an annual basis. Two waves of case-control data from TARCC were examined as part of genetic analyses in the ADGC. Data from the TARCC included 323 cases and 181 controls in the first wave (included in the TARCC1 cohort), with 84 cases and 115 controls in the second wave (included in the UM/CWRU/TARCC2 cohort). All TARCC subjects were greater than 65 years of age at disease onset (cases) or at last disease-free exam (non-cases).

TGEN2: Among the TGEN2 data analyzed were 668 clinically- and neuropathologically-characterized brain donors, and 365 CNEs without dementia or significant Alzheimer's disease pathology. Of these cases and CNEs, 667 were genotyped as a part of the TGEN1 series[66]. Samples were obtained from twenty-one different National Institute on Aging-supported Alzheimer's disease Center brain banks and from the Miami Brain Bank as previously described[66–69]. Additional individual samples from other brain banks in the United States, United Kingdom, and the Netherlands were also obtained in the same manner. The criteria for inclusion were as follows: self-defined ethnicity of European descent, neuropathologically confirmed Alzheimer's disease or neuropathology present at levels consistent with status as a control, and age of death greater than 65. Autopsy diagnosis was performed by board-certified neuropathologists and was based on the presence or absence of the characterization of probable or possible Alzheimer's disease. Where possible, Braak staging and/or CERAD classification were employed. Samples derived from subjects with a clinical history of stroke, cerebrovascular disease, comorbidity with any other known neurological disease, or with the neuropathological finding of Lewy bodies were excluded.

UKS: The UKS cohort is a thoroughly diagnosed case-control cohort from Universitätsklinikum des Saarlandes, consisting of individuals clinically diagnosed with sporadic Alzheimer's disease (N = 596; mean age onset, 72.2 ± 6.6 years) and cognitively healthy, age-, gender-, and ethnicity-matched population-based controls (N = 170; 64.1 ± 3.0 years).

UM/CWRU/MSSM: The UM/CWRU/MSSM dataset (formerly UM/VU/MSSM[70–73]) contains 1,177 cases and 1,126 CNEs ascertained at the University of Miami, Case Western Reserve University and Mt. Sinai School of Medicine, including 409 autopsy-confirmed cases and 136 controls, primarily from the Mt. Sinai School of Medicine[74]. An additional 16 cases were included and 34 controls excluded from the data analyzed in the Jun et al. 2010 study[40]. Each affected individual met NINCDS-ADRDA criteria for probably or definite Alzheimer's disease[8,11] with age at onset greater than 60 years as determined from specific probe questions within the clinical history provided by a reliable family informant or from documentation of significant cognitive impairment in the medical record. Cognitively healthy controls were unrelated individuals from the same catchment areas and frequency matched by age and gender, and had a documented MMSE or 3MS score in the normal range. Cases and controls had similar demographics: both had similar ages-at-onset/ages-at-exam of 71.1 (±17.4 SD) for cases and 73.5 (±10.6 SD) for controls, and cases and controls were 64.5% and 61.3% female, respectively.

UPITT: The University of Pittsburgh dataset contains 1,255 NHW Alzheimer's disease cases (of which 277 were autopsy-confirmed) recruited by the University of Pittsburgh Alzheimer's Disease Research Center, and 829 NHW, CNEs ages 60 and older (2 were autopsy-confirmed). All Alzheimer's disease cases met NINCDS-ADRDA criteria for probable or definite Alzheimer's disease[8,11]. Additional details of the cohort used for GWAS have been previously published[75].

WASHU: An NHW LOAD case-control dataset consisting of 377 cases and 281 healthy elderly controls was used in analyses for this study. This dataset was split between two analysis datasets (WASHU1 and WASHU2). Participants were recruited as part of a longitudinal study of healthy aging and dementia. Diagnosis of dementia etiology was made in accordance with standard criteria and methods[45]. Severity of dementia was assessed using the Clinical Dementia Rating scale[76].

WHICAP: WHICAP is a community-based longitudinal study of aging and dementia among elderly, urban-dwelling residents[77,78]. Beginning enrolment in 1989, WHICAP has followed more than 5,900 residents over 65 years of age, including white, African American, and Hispanic participants. Detailed clinical assessments were performed at approximately 24-month intervals over the 7 years of the initial study. All interviews were conducted in either English or Spanish. The choice of language was decided by the subject to ensure the best performance, and the majority of assessments were performed in the subject's home, which included medical, neurological, and neuropsychological evaluations. Results of the neurological, psychiatric, and neuropsychological assessments were reviewed in a consensus conference comprised of neurologists, psychiatrists, and neuropsychologists. Based on this review all participants were assigned to one of three categories: dementia, cognitive impairment, or normal cognitive function. The sample set available in the ADGC for genetic analyses included 73 Alzheimer's disease cases and 560 subjects with normal cognitive function.

CSDC: The Combined Small Datasets Collection is a harmonized dataset including all data from eight separately ascertained datasets or waves of datasets already described above (ACT2, BIOCARD, CHAP2, EAS, NBB, RMAYO, ROSMAP2, and WASHU2). None of these datasets were separately incorporated into the analyses, and were only analyzed in the CSDC. Datasets or waves were incorporated into the CSDC if they included fewer than 100 cases and/or 100 CNEs. As all datasets were genotyped separately but imputed to the same dataset, genotyped single nucleotide polymorphisms (SNPs) overlapping all of the high-density genotyping platforms used were extracted and a set of ~20,000 variants were used to estimate both population substructure and explore potential heterogeneity between datasets using EIGENSTRAT/EIGENSOFT[79,80]. Data-set level association analyses similar to those described for all other cohorts and datasets were performed, though covariate adjustment additionally included indicator variables for study to adjust for residual batch effects not captured in PCs. Association results among common variants (minor allele frequency [MAF]>0.01) in the CSDC were similar to other datasets in the ADGC with very modest deflation (λ=0.9828) while rare variant associations demonstrated extreme patterns of association with a high degree of genomic inflation (λ=1.227). For this reason, only common variant association results from the CSDC were incorporated into common variant meta-analysis, while rare variant association results were excluded from rare variant meta-analysis (none of the eight individual datasets composing the CSDC were used in rare variant meta-analysis).

***Cohorts for Heart and Aging Research in Genomic Epidemiology (CHARGE)***

CHS: The Cardiovascular Health Study (CHS) is a population-based cohort study of risk factors for coronary heart disease and stroke in adults ≥65 years conducted across four field centers[81]. The original predominantly European ancestry cohort of 5,201 persons was recruited in 1989-1990 from random samples of the Medicare eligibility lists; subsequently, an additional predominantly African-American cohort of 687 persons was enrolled for a total sample of 5,888. Genotyping was performed using the Illumina 370CNV BeadChip system (for European ancestry participants, in 2007) or the Illumina HumanOmni1-Quad_v1 BeadChip system (for African-American participants, in 2010). CHS was approved by institutional review committees at each field center and individuals in the present analysis had available DNA and gave informed consent including consent to use of genetic information for the study of cardiovascular disease.

FHS: Framingham heart study (FHS) samples consist of 4350 well genotyped individuals from Original and Offspring cohorts[82,83]. The details of recruitment and surveillance of AD, and genotyping in FHS have been detailed previously[84]. Briefly, the Original cohort of the FHS has been evaluated biennially since 1948, was screened for prevalent dementia and AD since 1974-76. The Offspring cohort (offspring of original cohorts and spouse of offspring), recruited in 1971 and examined once every 4 years, have been screened for prevalent dementia with a neuropsychological battery and brain MRI. The AD status used in this study was taken from surveillance up to 2017. FHS participants had DNA extracted and provided consent for genotyping in the 1990s. Genotyping using the Affymetrix GeneChip® Human Mapping 500K Array Set and 50K Human Gene Focused Panel.® was attempted in 5293 Original and Offspring cohort participants.

***FinnGen***

FinnGen is a public-private partnership project that aggregates genotype data from Finnish biobanks (https://www.finngen.fi/en). The latest FinnGen release (Data Freeze 6) consists of 260,405 samples after quality control with population outliers excluded via principal component analysis based on genetic data. The samples have been linked with harmonized data from several national healthcare related registries. AD (Alzheimer's disease, wide definition, N=7,329) cases were identified from hospital discharge and cause of death registries having G30 (International Classification of Diseases (ICD)-10) or 29010 (ICD-8) codes, from Finnish-specific Social Insurance Institute (KELA) reimbursement registry having 307 or G30 (ICD-10) codes, and from medicine purchase registry having N06D (Anatomical Therapeutic Chemical, ATC) code. The same criteria were used as exclusion

criteria for controls (N=252,879). The overlap between Finnish EADB and FinnGen controls is 0.3%.
Patients and control subjects in FinnGen provided informed consent for biobank research, based on the Finnish Biobank Act. Alternatively, older research cohorts, collected prior the start of FinnGen (in August 2017), were collected based on study-specific consents and later transferred to the Finnish biobanks after approval by Fimea, the National Supervisory Authority for Welfare and Health. Recruitment protocols followed the biobank protocols approved by Fimea. The Coordinating Ethics Committee of the Hospital District of Helsinki and Uusimaa (HUS) approved the FinnGen study protocol Nr HUS/990/2017.
The FinnGen study is approved by Finnish Institute for Health and Welfare (permit numbers: THL/2031/6.02.00/2017, THL/1101/5.05.00/2017, THL/341/6.02.00/2018, THL/2222/6.02.00/2018, THL/283/6.02.00/2019, THL/1721/5.05.00/2019, THL/1524/5.05.00/2020, and THL/2364/14.02/2020), Digital and population data service agency (permit numbers: VRK43431/2017-3, VRK/6909/2018-3, VRK/4415/2019-3), the Social Insurance Institution (permit numbers: KELA 58/522/2017, KELA 131/522/2018, KELA 70/522/2019, KELA 98/522/2019, KELA 138/522/2019, KELA 2/522/2020, KELA 16/522/2020 and Statistics Finland (permit numbers: TK-53-1041-17 and TK-53-90-20).
The Biobank Access Decisions for FinnGen samples and data utilized in FinnGen Data Freeze 6 include: THL Biobank BB2017_55, BB2017_111, BB2018_19, BB_2018_34, BB_2018_67, BB2018_71, BB2019_7, BB2019_8, BB2019_26, BB2020_1, Finnish Red Cross Blood Service Biobank 7.12.2017, Helsinki Biobank HUS/359/2017, Auria Biobank AB17-5154, Biobank Borealis of Northern Finland_2017_1013, Biobank of Eastern Finland 1186/2018, Finnish Clinical Biobank Tampere MH0004, Central Finland Biobank 1-2017, and Terveystalo Biobank STB 2018001.

##### 1.3. Longitudinal studies

13 longitudinal cohorts were included in the polygenic risk score (PRS) analysis (Supplementary Table 32). We used MCI patients from: the Dutch Amsterdam dementia cohort (ADC)[85], the German dementia competence network cohort (DCN)[9], two cohorts from the Spanish Fundacio ACE memory clinic cohort (FACE, AMC)[86], the French Balthazar cohort (HAN)[87], the Belgian memory clinic cohort from the Hospital Network Antwerp (UAN)[88], the German memory clinic of Halle (UHA), and the German memory clinic of Mannheim (ZIM)[89]. From the population based studies we used patients from: the German study on aging, cognition and dementia (AgeCoDe)[90], the Austrian VITA study[91], the Australian Sydney Memory and Ageing Study[18,92], the Three City study (3C)[25] and the Rotterdam study[24,93]. All selection criteria for the MCI patients and criteria used to define conversion to dementia are provided in the respective references.

#### 2. Quality control

##### 2.1. EADB

***Genotyping***

EADB genomic DNA samples were transferred to 3 genotyping centers: the Centre National de Recherche en Génomique Humaine, Evry, France (CNRGH), the Erasmus Medical Center, Rotterdam, the Netherlands (Erasmus MC) and the LIFE & BRAIN Center, Bonn, Germany (LIFE & BRAIN Gmbh). Samples that passed the DNA quality control (QC) were genotyped with the Illumina Infinium Global Screening Array (GSA, GSAsharedCUSTOM_24+v1.0). Raw probe intensities were shared with the CNRGH, which performed the genotype calling on all samples using the same custom cluster file obtained with the GenTrain 3.0 clustering algorithm (https://www.illumina.com/content/dam/illumina-marketing/documents/products/technotes/gentrain3-technical-note-370-2016-015.pdf). Of note, insertion and deletion polymorphisms were excluded from this process and only single nucleotide polymorphisms were called. During the genotyping QC process, three genotyping batches were considered: (1) 49 genotyping chips were identified as possibly problematic and thus were considered as a separate batch (denoted possibly problematic chips batch or PPC batch in the next sections), (2) a batch of samples was genotyped and processed after all other samples (denoted last genotyped batch or LGB batch in the next sections) and (3) the main batch including all other samples.

##### ***Chip assessment***

Prior to the initial QC, positions and alleles of variants were assessed. First, using the Illumina support files (https://support.illumina.com/array/array_kits/infinium-global-screening-array/downloads.html), variants for which the position was erroneous in the first version of the manifest had their positions corrected and variants which are part of the removed markers list provided by Illumina (i.e., exclusion because of multi-mapping, poor clustering, non-validated correlation against 1000 Genomes data, multinucleotide variants or discrepant rsID) were excluded. Then, variants' probes were aligned against both Human reference genome assemblies GRCh37 (GRCh37.p13) and GRCh38 (GRCh38.p12) using the bwa software v0.7.17 with the BWA-MEM algorithm[94]. Only variants for which the full-length probe(s) aligned uniquely on the genome without any mismatch were retained. Next, the GRCh37 coordinates of variants were remapped to the GRCh38 assembly (GCF_000001405.26) using the NCBI Remapping Service (https://www.ncbi.nlm.nih.gov/genome/tools/remap). Variants which were unmapped, not mapped on a primary contig of the Primary Assembly (chromosomes 1-22, X, Y, MT), or with a discordant position according to the previous alignment step on GRCh38, were excluded. Last, a normalization process of the alleles was performed on both GRCh37 and GRCh38 assemblies using the bcftools software (http://www.htslib.org/doc/bcftools.html) in order to obtain alleles expressed on the plus strand. Variants showing incompatible alleles against the reference assembly were removed. Finally, only variants passing in all steps, for both GRCh37 and GRCh38 assemblies, were included and their coordinates and alleles were set according to the GRCh38 assembly for the rest of the pipeline.

##### ***Variant Intensity Quality Control***

A QC on the intensity metrics extracted from the Illumina GenomeStudio software v2.0.3 (https://www.illumina.com/techniques/microarrays/array-data-analysis-experimental-design/genomestudio.html) was then performed on all autosomes and chromosome X variants. Only the metrics from the main batch were used in this QC. The involved intensity metrics and their thresholds were adapted from the CHARGE Consortium HumanExome BeadChip quality control paper[95]. Variants were removed based on the exclusion criteria described in the Supplementary Table 35.

##### ***Sample Quality Control***

To be consistent with the main batch and PPC batches, the sample QC for the LGB batch, performed afterwards, used the same thresholds (including the steps which require the computation of a metric mean or median).

Pre-quality control. First, a pre-quality control was performed on both autosomes and the chromosome X variants. Variants having a p-value <1e-15 for the Hardy-Weinberg equilibrium test in controls in at least one genotyping center or globally, or showing a missingness >0.05 in at least one genotyping center or >0.025 globally, were excluded prior to all the following sample quality control steps.

Heterozygosity and missingness. Sample missingness was computed using all autosomal variants while the sample heterozygosity was computed at the pruned set of autosomal variants (maximum $r^2$ of linkage disequilibrium (LD) set to 0.2 with a window of 500kb) using plink v1.9 (https://www.cog-genomics.org/plink2/). Samples showing a missingness >0.05 or showing a heterozygosity metric (Method-of-moments F coefficient estimate) outside the interval mean ±6 standard deviation (sd) were removed.

Population outliers. In order to identify population outliers, a principal component analysis (PCA) was performed. The 1000 Genomes Phase3 data (1000GP3) called on the GRCh38 assembly was used as the reference (http://ftp.1000genomes.ebi.ac.uk/vol1/ftp/data_collections/1000_genomes_project/release/20190312_biallelic_SNV_and_INDEL/). First, a subset of variants was selected to be included in the PCA: variants in common between 1000GP3 and the GSA variants passing the variant QC, having a minor allele frequency (MAF) >0.01 in both 1000GP3 and EADB, not ambiguous (i.e., A/T and C/G variants) and not located in high LD regions as described here[96] as well as LCT (2q21), HLA and 2 inversion regions (8p23 and 17q21.31) following the TOPMed analysis pipeline described here (https://github.com/UW-GAC/analysis_pipeline). Variants were then pruned using PLINK (maximum r² of LD set to 0.2 using a window of 500kb). Principal components (PCs) were computed on the 1000GP3 samples and the EADB samples were projected onto these PCs using the FlashPCA2 software[97]. Samples falling outside the interval median ±12 median absolute deviation (mad), computed using only the EADB samples, on PC1 or on PC2 were flagged as population outliers.

Sex-check. A sex-check of samples was performed to identify discordances between the genetic and the clinical sex information. All chromosome X variants of the non pseudoautosomal region, passing the variant pre-quality control, were pruned (maximum r² of LD set to 0.2 with a window of 500kb) and then used in the sex-check function of PLINK. Using default parameters of PLINK for exclusion on the resulting inbreeding coefficient F (i.e., F <0.8 for males, F >0.2 for females), samples with a discordant genetic and clinical sex were removed. Samples who had no clinical sex available were not excluded and their imputed genetic sex was used as a replacement in latter analyses. Also, plates showing more than 30% of failed sex-check samples were excluded.

Relatedness. An analysis was performed to infer the relatedness between samples using the GENESIS package[98]. This package provides methods to infer relatedness by taking into account the population structure. The pipeline described on the package documentation (https://bioconductor.org/packages/release/bioc/vignettes/GENESIS/inst/doc/pcair.html) was followed. All samples already excluded by a previous sample QC step or flagged as population outlier were excluded of this analysis. The included variants were the same as the ones included in the population outlier analysis minus the ones failing the variant QC described below (missingness, differential missingness test, Hardy-Weinberg test or frequency test). The default parameters of the pipeline were used at the exception of the kinship and divergence thresholds, used to assign relatives and samples of divergent ancestry in the PCAir step, which were set to 2^(-9/2) and -2^(-9/2) respectively, following the TOPMed analysis pipeline. Also, 21 PCs were used in the PC-Relate step of the pipeline which computes the final kinship coefficients. All pairs with a kinship >0.09375 (representing the mean between the 2nd and 3rd degrees) were then selected and processed in 3 different categories: duplicate samples (samples with a pairwise kinship >0.45), multi-related samples (samples related to more than one sample) and samples related to only one other sample. First, the duplicate samples were processed, excluding all involved samples if there was a status or sex mismatch (not considering a missing variable as discordant). If the clinical variables matched, samples were excluded in the following order: (1) the sample from the PPC batch, (2) the sample from cohorts not being imputed with TOPMed, (3) the sample with a missing status or (4) the sample with the highest missingness. Concerning the multi-related samples, samples were sorted by considering first the one with the most related samples (sorting also alphanumerically by the sample identifier in case of multiple samples with the same number of related samples for reproducibility) and this sample was excluded. All pairs involving this excluded sample were then discarded and the process was repeated until no more multi-related samples remain. Finally, for the remaining pairs, samples were excluded in the following order: (1) the sample from the PPC batch, (2) the sample with a pathogenic mutation, (3) the sample from cohorts not being imputed with TOPMed, (4) the control over the case or the sample with a missing status over the control or (5) the sample with the highest missingness.

Possibly problematic chips batch. After all the sample QC steps, we assessed the impact of the samples present in the PPC batch by performing a Genome-Wide Association study (GWAs) on all the EADB GSA samples and noticed many false positives signals coming from these samples so that this batch was entirely excluded.

***Variant Quality control***

For the variant QC, the initial set of autosomal variants passing the variant intensity quality control was used (re-integrating the variants failing the pre-quality control of the sample QC). All samples failing the sample QC were removed prior to the variant QC.

Missingness and Hardy-Weinberg equilibrium. The variants missingness, the p-value of the differential missingness test between cases and controls and the Hardy-Weinberg equilibrium test P value were computed using PLINK. Variants showing a missingness >0.05 in at least one genotyping center or having a P value of the Fisher's exact test on cases/controls missing calls <1e-10, were excluded. The Hardy-Weinberg equilibrium tests were performed only in controls and for each genotyping center/country pair separately. A variant was excluded if at least one center/country test showed a P value <5e-8.

Frequency checks. Two frequency tests of variants were performed to compare the frequency in the EADB GSA samples, excluding population outliers, against two reference panels: (1) the Haplotype Reference Consortium[99] (HRC) and (2) the Genome Aggregation Database[100] (gnomAD). For HRC, the release r1.1 was used and the frequency extracted was the one excluding 1000 Genome samples. For gnomAD, the release v3 was used as this version contains more than 70,000 whole genomes and thus has a better estimation for non exonic variants than the v2.1.1 which mostly contains whole exomes. Finnish and non-Finnish allele counts and frequencies were extracted from gnomAD sites having a PASS filter with more than 50% of the genomes called. To perform the frequency test, a Pearson chi-square test was performed on the allelic counts. After graphical review of the chi-squared test statistics ($\chi^2$) distribution, variants having (1) $\chi^2$ >3,000 in both HRC and gnomAD, (2) $\chi^2$ >3,000 in HRC and not present in gnomAD or (3) $\chi^2$ >3,000 in gnomAD and not present in HRC were excluded because of large difference of frequency. When the gnomAD and HRC tests disagreed (i.e., <3,000 in one and > 3,000 in the other), the variant was kept due to the uncertainty. Finally, when no frequency information was available from both HRC and gnomAD, the variant was excluded if it showed a different minor allele compared to the TOPMed reference panel freeze5[101] with a MAF difference higher than 0.2. Finally, to assess the frequency difference between genotyping centers, genome wide association studies (GWAS) were performed between controls across genotyping centers after excluding population outliers and related samples. First, PCs for all controls were computed with flashPCA2 using the same variants included in the relatedness estimation. Then we performed the analyses to compare controls from one genotyping center to controls from another genotyping center (3 analyses in total). The center was thus converted into a binary variable and used as the analysis phenotype. Adjustments included PCs associated significantly with the center and the analyses were performed with the SNPTEST software[102], v2.5.4-beta3 using an additive model with the new model-fitting functionality (newml method). All variants having a Likelihood Ratio Test P value <1e-5 were excluded.

Ambiguous variants. All ambiguous variants (e.i: A/T and C/G) showing a MAF >0.4 were excluded.

Duplicated variants. Concerning the duplicated variants of the chip, only the copy with the minimum missingness was kept.

Last genotyped batch supplementary steps. Since this batch was available after the variant QC of the main batch, only the variants passing all QC steps in the main batch were used as the initial set of variants prior to the batch variant QC. All steps described in this section were then performed for that batch separately. Also, since this batch contains only German samples, one supplementary step was performed: a GWAS between controls from this batch and German controls from the main batch following the same pipeline described in the Frequency checks section.

***Analysis principal components computation***
The principal components used as adjustment in the analysis were computed using AD cases and controls only, on the same variants which were retained for the relatedness estimation. These PCs were computed using the flashPCA2 software.

***Clinical data QC***
After QC of the genotyping data, we additionally excluded controls with age below 30 and individuals with known pathogenic mutations.

The EADB study finally included 20,464 AD cases and 22,244 controls for 606,881 autosomal variants (Supplementary Fig. 46 and 47). Among those, 20,301 AD cases and 21,839 controls were imputed with the TOPMed reference panel (EADB-TOPMed) while 163 AD cases and 405 controls were imputed with the HRC reference panel (EADB-HRC) as described in the Imputations section.

##### 2.2. Other datasets

***European Alzheimer's Disease Initiative (EADI) Consortium***
The EADI chip assessment only included the alignment, remapping and normalization step. The sample QC was already detailed here[84] and only the relatedness step was redone using the methodology used for the EADB GSA samples. The variant QC followed the same pipeline and steps than the EADB GSA chip with the same metrics and thresholds at the exception of the Frequency tests where (1) only allele counts from non-Finnish samples were extracted from the gnomAD reference panel and (2) the $\chi^2$ threshold used was set to 1,500 because of the EADI sample size. Also, no controls GWAS across controls/centers step was performed because not applicable for this study. After QC and exclusion of individuals with known pathogenic mutations, the EADI study included 2,400 AD cases and 6,338 controls for 523,431 autosomal variants.

***Genetic and Environmental Risk in AD (GERAD) Consortium***
We removed individuals with missing genotype rates > 0.01. We also applied a filter based on mean autosomal heterozygosity, inconsistencies between reported gender and genotype-determined gender. All individuals passing these QC filters were examined for potential genetic relatedness by calculating identity-by-descent (IBD) estimates for all possible pairs of individuals in PLINK, and removing one of each pair with an IBD estimate $\geqslant$0.125. We assessed population structure within the data using principal components analysis as implemented in EIGENSTRAT[79] to infer continuous axes of genetic variation. Eigenvectors were calculated based on LD-pruned SNPs common to all arrays. The EIGENSTRAT program also identifies genetic outliers, which are defined as individuals whose ancestry is at least 6 s.d. from the mean on one of the top ten axes of variation. Individuals either were genotyped on the Illumina 610-quad chip or on the Illumina HumanHap550 array. We assessed the effects of different missing data rates and Hardy-Weinberg filters, aiming to remove poorly performing SNPs without excluding markers that may show genuine association with Alzheimer's disease. Markers were excluded if they had a minor allele frequency (MAF) <0.01 or a Hardy-Weinberg $P \geqslant 1 \times 10^{-5}$ in either cases or controls. SNPs with a MAF $\geqslant$0.05 were excluded if they had a genotype missing rate of >0.03 in either cases or controls; for SNPs with a MAF between 0.01 and 0.05, a more stringent genotype missing rate threshold of 0.01 was used. To minimize inter-chip and inter-cohort differences minor allele frequencies were compared between controls in the different groups using logistic regression analysis, incorporating the top four PCs as covariates as previously described. Comparisons were performed only between individuals from the same geographical region (that is, British Isles, Germany or USA). For each of the four categories

of SNPs, a quantile-quantile (Q-Q) plot was produced for each cohort control comparison, and the significance threshold used to exclude SNPs was based on where the observed $\chi^2$ statistics departed from the null expectation. Finally, we applied a variant QC following the same pipeline and steps than the EADB GSA chip with the same metrics and thresholds at the exception of the Frequency tests where (1) only allele counts from non-Finnish samples were extracted from the gnomAD reference panel and (2) the $\chi^2$ threshold used was set to 1,500 because of the GERAD sample size. After QC, the GERAD study included 3,030 AD cases and 7,153 controls for 418,258 autosomal variants.

***Bonn studies***

DietBB: The dietBB chip assessment only included the alignment, remapping and normalization step. The sample and variant QCs followed the same pipeline and steps than the EADB GSA chip with the same metrics and thresholds at the exception of the Frequency tests where (1) only allele counts from non-Finnish samples were extracted from the gnomAD reference panel and (2) the $\chi^2$ threshold used was set to 250 because of the dietBB sample size. Also, no control GWAs across controls/centers step was performed because not applicable for this study. After QC, the dietBB study included 139 AD cases and 177 controls for 630,058 autosomal variants.

Bonn OMNI cohort: The Bonn OMNI chip assessment only included the alignment, remapping and normalization. The sample and variant QCs followed the same pipeline and steps than the EADB GSA chip with the same metrics and thresholds at the exception of the Frequency tests where (1) only allele counts from non-Finnish samples were extracted from the gnomAD reference panel and (2) the $\chi^2$ threshold used was set to 500 because of the Bonn OMNI sample size. Also, no controls GWAs across controls/centers step was performed because not applicable for this study. After QC, the Bonn OMNI study included 496 AD cases and 1,033 controls for 789,359 autosomal variants.

***DemGene (DG) Consortium***

deCODE chip batch: The deCODE chip assessment only included the alignment, remapping and normalization step. The sample and variant QCs followed the same pipeline and steps than the EADB GSA chip with the same metrics and thresholds at the exception of Hardy-Weinberg tests were all samples were included in the test, instead of only controls, because of the low number of controls. The Frequency test was also modified so that (1) only allele counts from non-Finnish samples were extracted from the gnomAD reference panel and (2) the $\chi^2$ threshold used was set to 250 because of its sample size. Also, no controls GWAs across controls/centers step was performed because not applicable for this study. After QC, the deCODE chip batch included 300 AD cases and 11 controls for 638,952 autosomal variants.

Omni chip batch: The Omni chip assessment only included the alignment, remapping and normalization step. The sample and variant QCs followed the same pipeline and steps than the EADB GSA chip with the same metrics and thresholds at the exception of the Frequency test where (1) only allele counts from non-Finnish samples were extracted from the gnomAD reference panel and (2) the $\chi^2$ threshold used was set to 1,500 because of its sample size. Moreover, there were multiple batches for this Omni chip which were taken into account for the differential missingness and the GWAs across controls. For the differential missingness, the test was performed globally as well as for the 4 batches including both cases and controls with enough sample sizes and, following the EADB GSA pipeline, a variant was excluded if it failed in at least one test. For the GWAs across controls, we selected the 4 batches with more than 400 controls and performed the GWAs following the same pipeline and thresholds than the EADB GSA chip. After QC, the Omni chip batch included 1,393 AD cases and 5,915 controls for 654,313 autosomal variants.

***The Copenhagen City Heart Study (CCHS, Denmark)***

The CCHS chips assessments were performed in parallel for both chips and only included the alignment, remapping and normalization step. Since common samples were genotyped on both chips, the sample and variant QCs were performed in parallel for both chips and followed the same pipeline and steps than the EADB GSA chip with the same metrics and thresholds at the exception of the population outlier QC where samples falling outside the interval median ±4 mad, computed using only CCHS samples, on PC1 or on PC2 were flagged as population outliers and the Frequency test where (1) only allele counts from non-Finnish samples were extracted from the gnomAD reference panel and (2) the $\chi^2$ threshold used was set to 1,500 because of the sample size. After the quality control steps performed for both chips, we merged the samples and variants passing individual chip QCs using PLINK where discordant genotypes between the 2 chips were set to missing. After the merging process, all the sample and variants QCs steps were performed again using the same pipeline except for the relatedness estimation for which the KING software[103] was used because of the absence of population structure. Finally, after performing the PCA analysis on all CCHS samples, some outliers were identified on the first PC and were excluded (PC1 <-0.05). After QC, the CCHS study included 365 AD cases and 6,106 controls for 467,446 autosomal variants.

***GR@ACE***
The sample and variant QCs followed similar pipeline and steps than the EADB QC. Duplicated samples between GR@ACE and EADB were excluded.

***Relatedness across studies***
Relatedness was inferred across each pair of the following studies: Bonn, CCHS, EADB, EADI, DemGene and GERAD. The process followed the same methods described in section 2, restricting the initial set of variants to variants in common between the two studies. Using the same thresholds, one sample of each related pair was excluded in one study, while the other sample was retained in the other study. The number of related and duplicate samples between processed studies can be found in Supplementary Table 36.

#### 3. Imputations

***TOPMed imputations***
All samples and variants passing the QC were used as the input of the imputation process. The imputation was performed by the Michigan Imputation Server[104] where the TOPMed Freeze5 reference panel was granted to the EADB consortium. The server version used was the 1.2.4 with Eagle v2.4[105] as the phasing software and Minimac4 v4-1.0.2 as the imputation software. Due to the limitation in terms of maximum number of samples per job (20,000), the GSA samples were split into 5 batches (4 for the main batch and one for the LGB batch). After the imputation process of all batches, in order to have a global imputation quality, a merged imputation quality was recomputed including all samples using the bcftools impute_info plugin.

***HRC imputations***
Samples which were imputed with the HRC reference panel were sent to the Sanger Imputation server (https://imputation.sanger.ac.uk/) where the HRC reference panel r1.1 was used. The data were phased using the Eagle v2.4 software and the imputation process was performed by PBWT.
For studies imputed with the HRC panel, we excluded from the meta-analysis variants with a very large difference of frequency between the HRC and TOPMed panels ($\chi^2$ >15,000). For the UK Biobank study, we further excluded variants with a very large difference of frequency between the UK10K+1000G or UK10K or 1000G panels and the TOPMed panel ($\chi^2$ >5,000).

#### 4. GRCh37/GRCh38 conversion

Prior to the meta-analysis, a conversion for variants from GRCh37 assemblies (i.e., studies not imputed with the TOPMed reference panel) was performed. First, variant positions were lifted from the GRCh37 assembly to the GRCh38 assembly using the UCSC liftover software (https://genome.ucsc.edu/cgi-bin/hgLiftOver). Variants failing the lift process were removed. Then, a normalization followed by a left-alignment process of these GRCh38 positions was performed using the bcftools software to obtain the reference alleles in the GRCh38 assembly. Comparing the reference alleles between GRCh37 and GRCh38 allowed to define if the GRCh37 alleles needed a flip, a swap or both in order to be represented in the GRCh38 assembly. For ambiguous variants, we also compared the flanking sequences (10 bases) of the variant in order to decide if a flip or a swap of alleles was needed. When results of the comparison was too complex to interpret or when the flanking sequences show more than 1 mismatch between the 2 assemblies, the variant was discarded.

#### 5. Stage II analyses

***Alzheimer disease genomic consortium (ADGC)***

Variant- and sample-level quality control (QC). Standard QC was performed on individual datasets using PLINK v1.9[106–108] and including filtering and re-estimating all quality metrics after excluding variants with a missingness rate of >10% of genotype calls. QC filters included exclusions on SNPs with call rates below 98% for Illumina and 95% for Affymetrix panels; SNPs with departure from Hardy-Weinberg Equilibrium (HWE) of $P<10^{-6}$ among cognitively-normal elders (CNEs, either non-cases or controls) for variants of MAF>0.01; and SNPs with informative missingness by case-CNE status of $P<10^{-6}$. Samples were dropped if the individual call rate was <95%; if X chromosome heterozygosity indicated inconsistency between predicted and reported sex; or if population substructure analyses (described below) indicated the sample did not cluster with 1000 Genomes Phase 3 populations of European ancestry.

Relatedness Check. Relatedness was assessed using the "--genome" function of PLINK v1.9. Using ~20,000 LD-pruned SNPs sampled from among genotyped variants, pi-hat (the proportion of alleles shared IBD) was estimated across all pairs of subjects across all ADGC datasets. Among pairs of subjects with no known familial relationships, one sample was excluded among pairs with pi-hat>0.95 if phenotype and covariate data matched, otherwise both samples were excluded; among all pairs with pi-hat>0.4 but less than 0.95, one sample was kept giving preference to cases over CNEs, age (earlier age-at-onset among case pairs, later age-at-exam among CNE pairs). Pairs of relatives were dropped from family datasets if pi-hat differed substantially from expectation based on their reported relationships.

Populations substructure. To identify samples of non-European ancestry, we performed a principal components (PCs) analysis using 'smartpca' in EIGENSOFT[79,80] on the subset of ~20,000 LD-pruned SNPs used for relatedness checks on genotypes from all samples within each individual dataset and from the 1000 Genomes Phase III reference panels. Subjects not clustering with European ancestry groups were excluded from analysis. To account for the effects of population substructure in our analysis, a second PC analysis was performed using only the remaining subjects in each dataset. PCs 1-10 were examined for association with AD case-control status and eigenvector loading, and only PCs showing nominal association with AD (P<0.05) and eigenvector loadings >3 were used in covariate adjustment for populations substructure (average number of PCs used is 3; range: 2-4).

Imputation. For each dataset, SNPs not directly genotyped were imputed on the Michigan Imputation Server (MIS)[104] using samples of all ancestries available on the Haplotype Reference Consortium (HRC) 1.1 reference panel[99], which includes 39,235,157 SNPs observed on 64,976 haplotypes (from 32,488 subjects), all with an estimated minor allele count (MAC)≥5 and observed in samples from at least two separately-ascertained data sources. Phasing on the MIS was done with EAGLE[105] while imputation was performed using Minimac3[104]. Quality of imputation for all variants was assessed using $R^2$ for imputation quality, although all variants were retained and not filtered prior to analysis. For rare variants, a global average of $R^2$ across all datasets weighted by sample size was considered.

Single-variant Association Analysis and Meta-analysis for Common Variants (MAF>0.01). Single variant-based association analysis on datasets of unrelated cases and CNEs were performed in SNPTEST[102] using score-based logistic regression under an additive model, with adjustment for PCs only. Family-based datasets were analyzed using the GWAF package in R[109] which implemented a generalized estimating equation (GEE) approach to account for correlation between subjects. For each study, we filtered (i) variants with missing effect size, standard error or P value, (ii) variants with absolute value of effect size above 5, (iii) variants with imputation quality below 0.3. Within-study association results were meta-analyzed using a fixed-effects approach with inverse variance-weighting using METAL[110].

Single-variant Association Analysis and Meta-analysis for Rare Variants (MAF≤0.01). Rare variant association and meta-analysis was performed for individual variants using the SeqMeta package in R[111,112]. SeqMeta performs a score-based logistic regression, estimating scores in individuals using 'prepScores()' and performing meta-analysis using 'singleSNPMeta()'. Family-based datasets were analyzed by selecting a maximally-informative subset of unrelated individuals for analysis, and no datasets with fewer than 100 cases and/or CNEs were analyzed (including the CSDC which demonstrated extreme association patterns and genomic inflation suggesting potential bias). As in common variant analyses, models evaluated included covariate adjustment for PCs. After meta-analysis, we filtered (i) variants with missing effect size, standard error or P value, (ii) variants with absolute value of effect size above 5, (iii) variants with average imputation quality below 0.3.

GRCh38/GRCh37 conversion. We converted variant positions and alleles from the GRCh37 assembly to the GRCh38 assembly (see above), and excluded variants for which conversion was not possible or problematic.

***Cohorts for Heart and Aging Research in Genomic Epidemiology (CHARGE)***

CHS: In CHS, the following exclusions were applied to identify a final set of 306,655 autosomal SNPs: call rate < 97%, HWE P < 10-5, > 2 duplicate errors or Mendelian inconsistencies (for reference CEPH trios), heterozygote frequency = 0, SNP not found in imputation reference panel. Imputation to the TOPMed Freeze5 panel was performed on the Michigan imputation server. SNPs were excluded for variance on the allele dosage ≤0.01. These analyses were limited to the 2152 European ancestry participants from the CHS Memory Study[113] with successful genotyping.

FHS: 4425 persons met QC criteria (call rate>97%, no extreme heterozygosity or high Mendelian error rate). Imputation to the TOPMed Freeze5 panel was performed on the Michigan imputation server. GWAS was carried out using a logistic regression model fitted via generalized estimating equations, with each family as a cluster, minimally adjusting for cohort status and the first and ninth PCs that were associated with the outcome.

CHARGE meta-analysis: For each study, we filtered (i) variants with missing effect size, standard error or P value, (ii) variants with absolute value of effect size above 5, (iii) variants with imputation quality below 0.3. FHS and CHS results were then combined with a fixed-effect meta-analysis (inverse variance weighted approach) as implemented in METAL to obtain the CHARGE results.

***FinnGen***

Detailed description of the FinnGen analysis pipeline can be found on the FinnGen website (https://finngen.gitbook.io/documentation/methods/phewas). Briefly, genome statistics were analyzed using Scalable and Accurate Implementation of Generalized mixed model

(SAIGE), which uses saddle point approximation (SPA) to calibrate unbalanced case-control ratios[114]. The first ten genetic PCs, sex, age, and genotyping batch were used as covariates.

***Stage II meta-analysis***

A fixed-effect meta-analysis was performed with METAL (inverse variance weighted approach) to combine ADGC, CHARGE and FinnGen results.

#### 6. Conditional analyses

In some regions, several variants were identified associated at the genome-wide significance level. In each of those regions, we performed an approximate conditional analysis of each associated variant conditionally on each other variant. The analyses were run with the GCTA-COJO approach[115,116], using the same EADB-TOPMed LD reference panel as in the PLINK clumping procedure. We repeated those analyses by performing exact conditional analyses as implemented in SNPTEST on the EADB-TOPMed dataset.
Those conditional analyses were also run between 1) the OARD1 and WWOX variants detected by Kunkle et al[84], and our top variants in the TREM2 and MAF loci respectively; 2) the TRIP4 variant detected by Ruiz et al[117] and our top variant in the SNX1 locus; 3) the ABCA7 variant with P value < $5x10^{-8}$ in the Stage I + II analysis but failing replication (see Supplementary Table 6) and our top variant in the ABCA7 locus.
The results are provided in Supplementary Tables 3 and 4. According to those conditional analyses, the following pairs of loci can be considered as independent: i) UMAD1 and ICA1; ii) CLU and PTK2B; iii) APH1B and SNX1; iv) SNX1 and TRIP4; v) DOC2A and KAT8; vi) WWOX and MAF; vii) ABCA7 and KLF16; viii) APP and ADAMTS1. Besides, we identified several independent signals in the MME, TREM2, SORL1, IGH gene cluster, and PLCG2 loci. In the ABCA7 loci, the two tested signals are also independent. However, the clumping procedure identified several signals in the CELF1/SPI1 and MAPT loci respectively, which are not independent according to those conditional analyses.

After validation by conditional analyses (Supplementary Tables 3 and 4), this approach led us to define 39 signals in 33 loci already known to be associated with the risk of developing AD and related dementia (ADD) and to propose 42 new loci (Table 1, Supplementary Table 5 and Supplementary Fig. 2-29). Six of these loci (*APP*, *ANK3/CCDC6, NCK2, PRKD3, TSPAN14* and *SHARPIN*) have already been reported in two preprints that examined some of the GWAS data included in our study[118,119].

#### 7. HLA analyses

The analyses were restricted to diagnosed cases and to the following datasets: EADB-TOPMed, GR@ACE/DEGESCO, GERAD, EADI, DemGene, Bonn, CCHS and EADB-HRC.

***Imputation of HLA alleles***

Two-field resolution alleles of *HLA-A*, *HLA-B*, *HLA-C* class I genes, and *HLA-DPB1*, *HLA-DQA1*, *HLA-DQB1*, and *HLA-DRB1* class II genes were imputed using R package HIBAG v1.4[120]. When available, European training sets specific to the genotyping arrays were used. Alleles with an imputation posterior probability lower than 0.5 were considered as undetermined as recommended by HIBAG developers.

***HLA analysis - Alleles***

In addition to the individuals excluded from the single variant analysis, individuals with more than 20% of missing genotypes were excluded from the HLA alleles analysis. Analysis was performed on a total of 34,067 cases and 54,361 controls. HLA imputed genotypes were converted to PLINK binary format files, considering each HLA allele as a SNP, and analyzed with SNPTEST. Same covariates as specified for the single variant analysis were used. HLA alleles with an effect size higher than 5 were excluded. Results were then meta-analyzed with a fixed-effect meta-analysis using the inverse variance weighted approach as implemented in the METAL software. Only alleles with a frequency higher than 1% were considered, representing 111 alleles. Adjusted P values were computed using the FDR method and the R p.adjust function, and applied to the meta-analysis P values. The FDR threshold was set to 0.05.

***HLA analysis - Haplotypes***

Only individuals with non-missing genotypes were included in this analysis. Three-locus HLA class I or class II haplotypes were determined using the haplo.em function from the R haplo.stats package. Haplotypes analysis was performed on a total of 28,253 cases and 46,005 controls. HLA haplotypes were converted to PLINK binary format files, considering each haplotype as a SNP, and analyzed with SNPTEST. Same covariates as specified for the single variant analysis were used. Haplotypes with an effect size higher than 5 were excluded. Results were then meta-analyzed with a fixed-effect meta-analysis using the inverse variance weighted approach as implemented in the METAL software. Only haplotypes with a frequency higher than 1% were considered, representing 36 three-locus haplotypes. Adjusted P values were computed using the FDR method and the R p.adjust function, and applied to the meta-analysis P values. The FDR threshold was set to 0.05.

***Results***

Association analysis of imputed HLA alleles showed an association of three HLA class II risk alleles (DQA1*01:01, DQB1*05:01, DRB1*01:01), three class II protective alleles (DQA1*03:01, DQB1*03:02, DRB1*04:04), and two HLA class I risk alleles (A*02:01, B*57:01) (Supplementary Table 8 and Supplementary Fig. 31). The associated HLA class II alleles form two distinct three-locus haplotypes that also showed association with ADD (the risk haplotype DQA1*01:01~DQB1*05:01~DRB1*01:01, OR=1.10 [1.06-1.14] and the protective haplotype DQA1*03:01~DQB1*03:02~DRB1*04:04, OR=0.87 [0.82-0.93]). One class I haplotype containing the risk allele B*57:01 also showed association with ADD (A*01:01~B*57:01~C*06:02, OR=1.15 [1.05-1.27]) (Supplementary Table 9 and Supplementary Fig. 32).

#### 8. PheWAS

We searched for the effects of the 83 ADD genome-wide significant variants (Table 1) in the GWAS of neurodegenerative and AD-related diseases. The effects and significance of the following diseases were provided by the corresponding authors of the following studies: Creutzfeldt-Jakob disease (CJD)[121], Dementia with Lewy-bodies (DLB)[122], amyotrophic lateral sclerosis (ALS)[123], Frontotemporal dementia (FTD)[124], Parkinson's disease (PD)[125], ischemic brain infarcts (MRI defined)[126] and ischemic stroke (clinical)[127], white matter hyperintensities (WMH)[128]. Reported effects and significance were transformed into Z-scores. The Z-scores for the risk allele for ADD from the current manuscript are reported (Supplementary Table 10). We were able to look-up 74 out of 83 ADD associated variants present in more than half of the explored traits. For these variants 91% (539/592) of the variant-trait associations were present in the GWAS. In Supplementary Fig. 33, we show these associations for the allele that increases the risk of ADD and clustered the variants. There were more variant-disease associations (P value<0.05) that increased ADD risk and associated with an increased disease risk (N=42), compared to associations opposite to the effect in ADD. Inspecting the clusters, there was a cluster of ADD associated variants that

was also associated with increased risk of Parkinson's disease (*HLA*, *CLU*, *NME8*, *SPPL2A*, *KAT8*). A second cluster associated with increased risk of FTD, ALS, and PD (*MAPT*, *MAF*, *CTSB*, *GRN*). A third cluster of loci associated with increased risk of almost all associated traits (*TNIP1*, *PICALM*, *HS3ST5*, *PLCG2*, *PLEKHA1*, *PRKD3*, *SHARPIN*, and *DOC2A*).
Next we conducted a PheWAS using the 'phewas' function of the R-package 'ieugwasr'[129,130]. This function searches traits that associate with a list of variants, with a P value lower than a given value in all GWAS harmonized summary statistics in the MRC IEU OpenGWAS data infrastructure[130]. We chose to search for the 83 ADD-associated variants (Table 1) only for association with $P<1x10^{-5}$. This resulted in 1980 significant associations. We included GWAS with priority (priority=0), excluded eQTLs, excluded results from Japan biobank GWAS, excluded all GWAS with '_raw' in them (as they are the duplicate of those with 'irnt'), removed duplicates and we removed all GWAS of AD or history of AD. After this cleaning, 660 associations remain. We report these in Supplementary Tables 11 and 12. In total 27 variants had no other trait than ADD associated with them (or variants were not present in the public GWAS), 9 had 1 single trait, 17 had between 2 and 5, 24 had between 6 and 20 and 6 were very pleiotropic with over 21 traits associated. Interesting traits that appear more than once are IGF-1 (8 loci), systolic and diastolic blood pressure (8 loci), Aspartate aminotransferase (8 loci), Apoliprotein A (5 loci), Albumin (7 loci), Alkaline phosphatase (6 loci) and Cystatin C (6 loci).

#### 9. GWAS signal colocalization analyses

For loci known to be associated with other neurodegenerative disorders, colocalization analyses were performed. For each locus, we performed the analysis in two steps: (1) a fine mapping analysis in Stage I to see whether the signal was due to only one causal variant and (2) a colocalization analysis to see if this signal was shared with the other neurodegenerative disorder.

***Fine mapping***
To assess whether the locus contained multiple independent signals, all the imputed variants having a MAF ≥0.005 and which were analyzed in the meta-analysis in ≥95% of the total number of samples were considered. Those variants were extracted from the EADB-TOPMed imputations, restricted to the samples included in the analysis, and converted into a hard-called genotypes PLINK format using a probability cutoff of 0.8. A joint analysis using the Stage I results, along with the EADB-TOPMed extracted genotypes for the linkage disequilibrium estimation, was performed using the GCTA-cojo method[131]. We used the stepwise model selection procedure of GCTA-cojo with a P value threshold of $10^{-5}$.

***Colocalization***
The colocalization analysis between Alzheimer's disease and the other neurodegenerative trait was performed for each locus independently. For each region, only the common variants between the Stage I results and the summary statistics of the other trait were considered. When the summary statistics of the other trait was expressed on another build than GRCh38, the variant alleles and positions were converted according to the method previously described. Then, only variants having a MAF ≥0.005 in Stage I and analyzed in ≥95% of the total number of samples in both traits were extracted. The colocalization analysis was performed with the coloc R package v4.0-4 using the enumeration of configurations under a single causal variant assumption method[132]. The analysis was performed on regression coefficients and their variance using defaults priors but also using a p12 prior (prior probability of any random variant in the region is associated with both traits) of $5x10^{-6}$ for sensitivity analysis.

***Loci assessed.***
Colocalization analyses were performed for 3 loci:

- IDUA: the summary statistics from a Parkinson's disease (PD) GWAS[125] was used and the region tested was restricted to chr4:643555-1243555 (GRCh38).
- GRN: we used the summary statistics from a Frontotemporal Dementia (FTD) GWAS[133] (all type of FTDs) and a Frontotemporal lobar degeneration with TAR DNA binding protein (TDP-43) inclusions (FTLD-TDP) GWAS[134]. The region tested was restricted to chr17:44102876-44602876 (GRCh38).
- TMEM106B: We used the same summary statistics as for the GRN locus. The region tested was restricted to chr7:11961934-12461934 (GRCh38).

#### 10. Gene prioritization

##### 10.1. Gene prioritization methods

***Description of genetic and transcriptomic datasets and cohorts.***
Two RNA-sequencing (RNA-seq) data sources were used in this study. First, through the Accelerating Medicines Partnership AD (AMP-AD) Knowledge Portal, we used uniformly processed AD-relevant brain RNA-seq datasets from the Mayo RNAseq Study (MayoRNAseq)[135], The Religious Orders Study and Memory and Aging Project (ROSMAP)[136,137], and The Mount Sinai Brain Bank study (MSBB)[138] available under consortium study "AMP-AD Cross-Study RNAseq Harmonization" (accessed in December 2019). Briefly, for MayoRNAseq and ROSMAP RNA-seq datasets, PolyA-selected libraries were sequenced on an Illumina HiSeq2000 platform (101 bp paired-end); for the MSBB RNA-seq dataset, rRNA-depleted libraries were sequenced on an Illumina HiSeq250 (100 bp single-end). Of note, among these three RNA-seq datasets, only ROSMAP dataset was stranded. Inclusion criteria were: (i) RIN value ≥ 5, (ii) availability of whole-genome sequencing (WGS) genotypes, and (iii) passing RNA-seq QC checks performed by the respective studies such as expression principle component analysis (samples should be positioned within the mean PC1 and/or PC2 ± 3xSD range), gene body coverage (a ratio of <3 between read number values at the $80^{th}$ and $20^{th}$ percentile), no sample swaps, concordance with genetic information and clinical metadata available. Additionally, in case multiple QCed RNA-seq samples were available on the same brain region of an individual, the RNA-seq sample selection was prioritized first based on the higher number of mapped reads and then based on the lower rRNA ratio.
Furthermore, through the AMP-AD Knowledge Portal, we accessed cohort-specific multi-sample WGS VCFs of MayoRNAseq, ROSMAP and MSBB studies that were generated by running GATK[139] (v3.4) HaplotypeCaller on 150 bp paired-end reads (sequenced on an Illumina HiSeq X) aligned to GRCh37 human reference genome. For each cohort-specific WGS VCF, we only selected autosomal variants that are passing Variant Quality Score Recalibration (VQSR) filters. Multiallelic variants were split and indels were left-aligned using BCFtools (v1.9) norm function. Moreover, we applied genotype level QC by assigning individual genotypes as missing if genotype quality (GQ) < 20 (using BCFtools) or if allele depth ratios of heterozygous genotypes exceeding 1:3 ratio, or if allele depth ratios of homozygous genotypes are within 1:9 ratio (using vcffilterjdk[140] with a custom filtering java code that is available upon request). We then removed the variants missing in more than 85% of the cohort and/or the variants deviating from Hardy-Weinberg Equilibrium (HWE $P < 10^{-6}$). For sample QC, we excluded the samples with call rate < 95%. For each cohort, PLINK was first used to select non-missing (variant missingness ≤ 0.02), common (MAF ≥ 1), and LD-pruned (PLINK parameters: "--indep-pairwise 500kb 1 0.2") variants that are out of following long-range LD loci that are likely to confound genomic scans: LCT (2q21), HLA (including MHC), 8p23 & 17q21.31 inversions, and 24 other long-range LD regions[96]. Using

these variants, we then estimated identity-by-descent (IBD) and heterozygosity ratios across the samples and excluded them based on relatedness (for pairs with PI-HAT > 0.2, we kept the sample with higher call rate or higher Genomic Quality Number [GQN]) and on excess heterozygosity (out of mean ± 3xSD range). Furthermore, using these LD-pruned high-quality common variants, for each cohort we calculated genetic principal components for subsequent downstream analyses including molecular quantitative trait locus (QTL) mapping.

Finally, as a result of the above genetic and transciptomic QC and selection criteria, in our study we included a total of 1067 QCed unique WGS samples with 1552 QCed unique RNA-seq samples derived from six different brain regions in AD-relevant frontal and temporal lobes. These regions and their respective studies are as below: *n*=259 temporal cortex (TCX) RNA-seq samples from MayoRNAseq (31% AD, 31% progressive supranuclear palsy, 11% pathological aging diagnoses, 27% healthy controls; 51% female, mean age at death at >80 years) study, *n*=560 dorsolateral prefrontal cortex (DLPFC) samples from ROSMAP (spectrum of clinical diagnosis at death: 32% no cognitive impairment [CI], 34% AD with no other CI, 34% other MCI and dementia types; 64% female, mean age at death at >87 years) study, and *n*=248 individuals in MSBB (spectrum of clinical dementia rating scores: 14% no dementia, 13% MCI, rest 73% are dementia at different stages) 66% female, mean age at death at >84 years) study with RNA-seq data available from frontal pole (*n*=207; Brodmann area [BA] 10), superior temporal gyrus (*n*=186; BA22), parahippocampal gyrus (*n*=162; BA36), and inferior frontal gyrus (*n*=178; BA44). We downloaded RNA-seq BAM files (aligned to genome indexes generated from human genome GRCh38 and GENCODE 24 with STAR RNA-seq aligner v2.5[141].1b) for these selected QCed samples for further processing and data analyses.

Second, we used a cohort of 70 EADB Flanders-Belgian samples with lymphoblastoid cell line (LCL) RNA sequencing (RNA-seq) and TOPMed-imputed genetic information from EADB project available (referred to as “EADB Belgian LCL” cohort). EADB Belgian LCL cohort individuals consisted of 51 AD patients (49% female, mean age at blood sampling 75.5 ± 4.4 years), 17 healthy controls (47% women, mean age at blood sampling 74.9 ± 6.7 years), and 2 individuals with mild cognitive impairment (50% female, mean age at blood sampling 74 ± 1.4 years).

Lymphoblast cells were cultured at 37 °C with 5% CO2 in RPMI1640 medium supplemented with 15% fetal bovine serum, 1% Glutamax, 1% Sodium pyruvate and 1% Penicillin/Streptomycin. Total RNA was isolated from Epstein - Barr virus (EBV) immortalized lymphoblasts derived from whole blood lymphocytes for all included samples. RNA isolation was performed using $10^7$ lymphoblast cells for each sample with the RNeasy mini kit (Qiagen Inc., Valencia, CA) according to manufacturer's protocol. Depletion of genomic DNA from the RNA sample was performed by turbo DNase treatment (Life Technologies, Carlsbad, CA, USA). RNA concentration was measured by dropsense 16 (Trinean, Gentbrugge, Belgium). RNA integrity number (RIN) values were obtained using the Agilent Technologies 2100 Bioanalyzer (Agilent Technologies, Santa Clara, CA, USA). RIN values were between 6.6 and 9.7 with an average value of 8.4 ± 0.8 Sequence libraries were constructed using the TruSeq Stranded mRNA Library Prep Kit (Illumina, San Diego, CA) using 1µg total RNA for each sample. Library preparation included RNA poly-A selection, RNA fragmentation, and random-hexamer-primed reverse transcription cDNA synthesis. Sequencing of prepared libraries was performed using an Illumina HiSeq2000 sequencer at the Macrogen NGS sequencing core, Seoul, Rep. of Korea, generating an average of $72x10^6 \pm 6x10^6$ 101 base-pair (bp) paired-end sequence reads. RNA-seq reads were mapped to genome indexes generated from human reference genome GRCh38 and GENCODE 32 using STAR RNA-seq aligner (v2.7.3a). These 70 LCL RNA-seq samples were paired with their corresponding TOPMed-imputed genetic data (genotyped on Illumina Global Screening Array platform within the framework of EADB project). The genetic principal components for the genetic data of these 70 individuals were computed using PLINK (v1.9)[107] on the same subset of high-quality LD-pruned variants used to calculate principal components in EADB GWAS.

***Expression & splicing quantification and cis-e/sQTL mapping***

To quantify gene expression for mapping expression QTLs (eQTLs) we followed the GTEx pipeline[142] with adaptations. First, we downloaded GENCODE GTF files (v24 for AMP-AD datasets and v32 for EADB Belgian LCL dataset), patched chromosome prefixes and created collapsed gene models with the Python script "collapse_annotation.py" that merged known transcripts into a single transcript model for each gene. Then RNASeQC (v2.3.5; "--legacy" parameter was used to enable compatibility with RNASeQC v1.1.9) was used to quantify expression on seven different RNA-seq datasets[143]. We added the "--unpaired" parameter for allowing quantification of single-end reads of MSBB RNA-seq datasets and "--stranded rf" parameter was added for stranded RNA-seq datasets (EADB Belgian LCL & ROSMAP). After combining transcript per million (TPM) counts and gene counts per sample in each RNA-seq dataset, we created normalized gene expression matrices using "eqtl_prepare_expression.py" Python script that (i) first filtered the genes with <0.1 TPM & <6 reads in at least 20% of samples in each dataset, (ii) normalized expression values for each sample using trimmed mean of M values (TMM) to account for library size, and (iii) normalized gene expression across samples for each gene with inverse normal transformation[144]. We also calculated Probabilistic Estimation of Expression Residuals (PEER) factors for each dataset to account for potential technical confounders for gene expression[145]. The resulting BED files of normalized gene expression matrices were used for eQTL mapping.

For annotation-free splicing quantification in each RNA-seq dataset, we used Leafcutter (v0.2.9) and RegTools (v0.5.1)[146,147]. First, RegTools "junction extract" command was used to extract exon-exon junctions with minimum anchor length of 8 bp, minimum intron size of 50 bp and maximum intron size of $5x10^5$ bp; and "-s 1" parameter was added for stranded RNA-seq datasets (ROSMAP and EADB Belgian LCL). The resulting splice junction quantification files per sample in each dataset were clustered using "leafcutter_cluster_regtools.py" Python script, where we filtered out splicing clusters with less than 50 split reads. We then used "prepare_phenotype_table.py" Python script that (i) performed quantile normalization of the distribution of splice junction usage ratios per sample to a normal distribution, (ii) standardized these ratios across samples, and (iii) calculated splicing principal components (sPCs) for each dataset. The resulting BED files of normalized splice junction usage ratios were used for splicing QTL (sQTL) mapping.

For eQTL and sQTL mapping, an enhanced version of FastQTL was utilized[148]. We used only the common (MAF ≥1%) QCed WGS variants (for AMP-AD cohorts) or common imputed (imputation quality score $R^2 \geq 0.3$ in the EADB cohort that was used in GWAS) variants (for EADB Belgian LCL cohort). Prior to QTL mapping, we lifted-over QCed AMP-AD WGS variants from GRCh37 to GRCh38 genome build by using Picard (v2.22.6) LiftOver tool. All genetic variants were annotated with dbSNPv151 (GRCh38) using BCFtools annotate function. We considered the genetic variants within 1 million bases window from the transcription start sites (TSS) and splice sites, respectively for eQTL and sQTL mapping. Sex and first 3 genetic principal component covariates were included in the linear regression models both for eQTL and sQTL mapping. In addition to these, following the recommendations of GTEx pipeline based on the sample size, for eQTL mapping we also included first 15 PEER factors as covariates for EADB Belgian LCL, 30 PEER factors for MSBB, 45 PEER factors for MayoRNAseq, and 60 PEER factors for ROSMAP datasets; and for sQTL mapping we added first 15 sPCs for all datasets in the linear regression models. We performed linear regression with FastQTL by first (i) generating nominal *P* values for each tested variant-gene or tested variant-splice junction pair, then (ii) using Beta distribution-adjusted empirical *P* values (generated by adaptive permutations with "--permute 1000 10000") of the most significant variant-molecular phenotype pair to calculate *q*-values[149] for estimating false discovery rate (FDR), (iii) applying FDR ≤0.05 filter to identify genes or splice junctions with at least one significant e/sQTL ("eGene" and "sJunction"), and finally (iv) defining all eQTL variant and eGene and sQTL variant and sJunction pairs as significant if their nominal *P* passes the significance threshold defined for each eGene and sJunction by the permutation and FDR procedure.

***Molecular QTL Catalogues***

In this study, in addition to 7 eQTL catalogues and 7 sQTL catalogues that we prepared (as explained above); we also used various publically available molecular QTL catalogues to assess the potential downstream regulatory effects of GWAS variants on these molecular phenotypes, including expression, splicing, and methylation. Our main source for these was GTEx v8 where we utilized cis-e/sQTL catalogues for selected AD-relevant GTEx brain regions (hippocampus [*n*=165], frontal cortex [*n*=175], cortex [*n*=205] and anterior cingulate cortex [*n*=147; BA24]), LCL (*n*=147), and whole blood (*n*=670). As our molecular phenotype quantification and QTL mapping methodology highly overlap with the methodology used to construct these GTEx catalogues, we primarily used these GTEx v8 catalogues to replicate the effects of significant cis-e/sQTL variants in a general population context (compared to our cis-e/sQTL catalogues that are derived from the AD cohorts). Furthermore, we incorporated the results from Microglia Genomics Atlas (MiGA) that is a recently established microglial expression and splicing regulation dataset[150]. We retained significant eQTLs and sQTLs (after a similar permutation and FDR [≤0.05] procedure described above for the other e/sQTL catalogues) in four different brain regions: medial frontal gyrus (MFG [BA9], *n*=63), superior temporal gyrus (STG [BA22], *n*=55), subventricular zone (SVZ, *n*=53) and thalamus (THA, *n*=45). Because the sample size is rather low compared to other bulk RNA-seq based e/sQTL catalogues, as also recommended by the authors, we additionally used the meta-analysis results based on RE2 random effects model as implemented in METASOFT[151] that combined four brain regions assessed (*n*=216). To define the significance of meta-analysis microglial e/sQTL associations, the stringent Bonferroni-corrected *P* value thresholds based on the number of independent tests were used ($\leq 6.58 \times 10^{-10}$ for MiGA Meta eQTLs and $\leq 1.79 \times 10^{-10}$ for MiGA Meta sQTLs).

Moreover, we used the blood cis-eQTL catalogue from eQTLGen project[152] (December 2019 release) that is the largest eQTL catalogue as it analyzed the regulatory effects of variants in over 30K individuals. Furthermore, macrophage[153,154] and monocyte[155–158] (CD14+ or CD16+) cis-eQTLs uniformly prepared by eQTL Catalogue database[159] (release 3 - October 2020) were also used in our analyses, where we used a nominal *P* value threshold of $\leq 10^{-5}$ to define significance for the associations. We mainly prioritized the eQTL effects in the naïve state macrophages and monocytes over the effects in stimulated macrophages and monocytes that were stimulated with various stimulants such as Influenza, Listeria, Salmonella, IFNg (Interferon gamma), LPS (Lipopolysaccharides), Pam3CSK4 (Pam3CysSerLys4), and R848 (Resiquimod) with different lengths of time. Finally, in addition to these e/sQTL catalogues, we also utilized brain methylation QTL (mQTL; *n*=468) and histone acetylation (haQTL; *n*=433) catalogues available at Brain xQTL serve[160] and that were mapped by integrating imputed genotypes with H3K9Ac ChIP-seq data and DNA methylation array data of DLPFC samples from ROSMAP project[161,162].

***e/sQTL colocalization and e/sTWAS***

The genetic colocalization between EADB GWAS and e/sQTL signals were investigated using coloc (v4.0.4)[132] in 12 eQTL and 12 sQTL catalogues for which we have full summary statistics available (AMP-AD, EADB Belgian LCL, and MiGA). We uniformly annotated and matched the variants in the EADB GWAS summary statistics and e/sQTL summary statistics files with the rsIDs from dbSNPv151 (GRCh38), and if no rsID available, with the "CHR_POS_REF_ALT" format using BCFtools annotate function. We selected a list of eGenes and sJunctions whose significant e/sQTLs (in at least one e/sQTL catalogue assessed) are associated with ADD risk in EADB Stage I GWAS minimally at a suggestive significant level (*P* value $\leq 10^{-5}$). We then ran "coloc.abf" (Bayesian colocalization analysis using default priors) on each selected eGene and sJunction considering all tested variants within 1 Mb of the TSS or splice site (except for MiGA sQTLs that used a 100kb window).

The results showed the calculated posterior probabilities (PP) for 5 different hypotheses between two signals compared: H0 (no causal variant for both traits), H1 (causal variant only for EADB GWAS), H2 (causal variant only for e/sQTL), H3 (two distinct causal variants) and H4 (common causal variant shared between EADB GWAS and e/sQTL catalogue). We considered a signal as colocalized in EADB GWAS and e/sQTL catalogue if coloc PP4 is at least 70%.
We investigated the association between predicted expression and splicing and ADD risk by performing a transcriptome-wide association study (TWAS) using EADB summary statistics and expression and splicing reference panels. First, we used FUSION pipeline[163] to create custom expression and splicing reference panels based on AMP-AD and EADB Belgian LCL e/sQTL catalogues generated. To this end, we supplied e/sQTL mapping input files to "FUSION.compute_weights.R" R script for expression and splice weight calculation per study. These prediction weights were computed using BLUP, LASSO, top SNP, and Elastic Net models. We used "--hsq_p 0.05" parameter for heritability *P* to calculate functional weights for genes and splice junctions only if they are significantly heritable features. We generated a custom LD reference data (annotated with dbSNPv151 and excluding variants with HWE $P < 10^{-6}$) by analyzing the phased biallelic SNV and INDEL genetic variants called *de novo* on GRCh38 (for selected *n*=404 unrelated Non-Finnish European (NFE) samples from the 1000 Genomes (1KG) project[164,165]. We restricted TWAS functional weight modelling by only using the variants that are both found in this LD reference data and the QCed genetic data of the cohort. We then ran TWAS by integrating EADB Stage I GWAS summary statistics, custom LD reference data, and custom expression (*n*=7) and splicing (*n*=7) reference panels using "FUSION.assoc_test.R" R script. The TWAS significance thresholds were defined per study based on Bonferroni correction on transcriptome-wide number of tested features (Supplementary Table 21).
Second, we ran additional expression and splicing TWAS of ADD using precalculated MASHR-based expression and splicing prediction models for GTEx v8 datasets[166] using S-PrediXcan[167,168] implemented in MetaXcan tools[167]. We ran S-PrediXcan (with non-default parameters "--keep_non_rsid --model_db_snp_key varID --additional_output –throw) using EADB summary statistics and MASHR model and covariance files for the same GTEx brain regions, cells and tissues chosen for e/sQTL studies: hippocampus, frontal cortex, cortex, BA24, LCL and whole blood. The significance thresholds of S-PrediXcan results were defined per study based on Bonferroni correction on transcriptome-wide number of tested features (Supplementary Table 21).
Finally, we used FOCUS (v0.7) for fine-mapping of expression TWAS results[169]. We imported both FUSION and PrediXcan based expression weights per dataset and ran fine-mapping of TWAS associations in regions of interest (based on 1 Mb extended region of lead variants in each novel EADB GWAS loci) to calculate posterior inclusion probabilities (PIPs) for each association which were later used to define the 90% credible sets of genes (which we accepted as fine-mapped TWAS associations).

***Long-read cDNA Sequencing***

For validating two AD-associated splice junction clusters that contained complex novel cryptic splicing events within *TSPAN14*, we designed an amplicon-based long-read single-molecule nanopore sequencing experiment[170] on cDNA derived from hippocampus, frontal cortex BA10 and LCL of AD patients and cognitively healthy individuals of our Flanders-Belgian cohort (Fig. 3). We first designed the following two amplicons with Primer3[8] (v4.1)[171]: long range cDNA amplicon 1 (forward primer: 5`-CTCTAACGCCAAGGTCAGCT-3`, reverse primer: 5`-CTCCCTCAACTCTGCTCCTC-3`) and long range cDNA amplicon 2 (the same forward primer, reverse primer: 5`-CTGACATGGCCAAGGAGTG-3`). All primers contained tag sequences for nanopore sequencing. All PCR amplifications were performed with 35 cycles using Platinum Taq DNA polymerase (Thermo Fisher). After the reactions, excess primers and nucleotides were cleaned with ExoSAP-IT (Thermo Fisher). Amplicon 1 was generated on *n*=60 LCL (44 AD patients, 15 healthy individuals, 1 MCI patient, 48% female, mean age at blood sampling 75.3 ± 5.3 years), *n*=18 frontal cortex BA10 (50% AD patient,

44% female, mean age at death 79 ± 7.4 years), and *n*=16 hippocampal (50% AD patient, 44% female, mean age at death 79.5 ± 8.3 years) cDNA samples; and barcoded with the PCR Barcoding Expansion 1-96 kit (Oxford Nanopore Technologies) using 20 amplification cycles on a 1/200 diluted template. Amplicon 2 was only run on a pooled unbarcoded sample of 14 LCL cDNA samples (8 AD patients, 5 healthy individuals, 1 MCI patient, 57% female, mean age at blood sampling 74.8 ± 4.2 years) as we only identified these splicing events in LCL. After purification with Agencourt AMPure XP beads (Beckman Coulter, Brea, CA, USA) and concentration measurement with Qubit (Thermo Fisher), amplicons were pooled equimolarly. The sequencing library was prepared as previously described[172]. SQK-LSK109 chemistry and FLO-FLG001 Flongle flow cell adapted into MinION platform were used for sequencing at the VIB-UAntwerp Center for Molecular Neurology, Antwerp, Belgium.
Base calling of the raw reads was performed with ONT basecaller Guppy (v3.2.4) on the Promethion compute device in Antwerp. After demultiplexing of basecalled FASTQ reads with qcat (v1.0.1) alignment of demultiplexed reads to GRCh38 reference genome was performed with minimap2 (v2.17) with parameters "-L -ax splice"[173]. NanoStat (v1.1.2) was used to calculate sequencing statistics which indicated an output of 311 million sequenced bases and 387 thousand reads with a median read length of 903 bp[174]. The resulting number of successfully sequenced cDNA samples were: 59 LCL cDNA, 18 frontal cortex BA10 cDNA, and 16 hippocampal cDNA samples for Amplicon 1; and a pooled LCL cDNA sample for Amplicon 2. We removed secondary alignments and supplementary alignments from the aligned reads using Samtools (v1.9). The aligned reads whose lengths of clipped bases were over 20% of their actual length were excluded using SamJdk[140]. For extracting Amplicon 2 specific aligned reads from the unbarcoded sample, we extracted the aligned reads containing unique reverse primer for this amplicon in the 3' end of the aligned read. The AD-associated cryptic splicing events and cryptic exons in *TSPAN14* were also confirmed by visualization of aligned reads on Integrative Genomics Viewer (IGV; v2.4.17)[175]. We merged the aligned reads of Amplicon 1 and Amplicon 2 based on LCL, frontal cortex and hippocampus categories and then ran mosdepth[43] (v0.2.9) to generate the cumulative coverage tracks of each amplicon per cDNA type for further data visualization[176].

***Genetically driven DNA methylation scan (MetaMeth)***
We tested for association between ADD and genetically driven DNA methylation (DNAm) using the procedures proposed previously by Freytag et al.[177] and Barbeira et al.[167]. The approximate association statistics between the methylation of 5′-C-phosphate- G-3′ (CpG) sites and ADD were computed with the function *MetaMethScan* from the *EstiMeth* (v1.1) software. The approach was applied to the EADB Stage I summary statistics, using the default DNAm estimation models and variant covariance structure, which is inferred from the 1000G European population. We performed a systematic search of CpG association signals within a region of 1 Mb around the lead variants. We considered as significant CpG sites with P value < 0.05, after Bonferroni correction for multiple testing for 77,881 features. Each CpG was paired with its annotated gene(s) and respective positional annotation of CpGs. Significant CpG sites were annotated by their percentile and direction of blood-brain methylation correlation estimates across three brain regions that were obtained from BECon[178].

##### 10.2. Gene prioritization results

The gene prioritization process is described in the Methods. The results for the prioritized genes are summarized in Fig. 2. A results summary for all the protein-coding genes with at least one significant signal for the criteria considered is provided in Supplementary Table 20 (and the results for non-coding genes provided in Supplementary Table 30), with a full description of the results in Supplementary Tables 21-29 and Supplementary Fig. 34-42.

***Prioritized genes that are the closest to the lead variants***

For ten of the novel loci; brain molecular QTL, TWAS, blood MetaMeth, and/or APP metabolism results exclusively supported the protein-coding gene nearest to the lead variant:

- ***TNIP1*** (L11). We identified that the risk allele of the lead variant (that is intronic in *TNIP1*) was significantly associated with increased *TNIP1* expression in DLPFC. (Supplementary Table 22 and Supplementary Fig. 34a).
- ***ICA1*** (L15). The evidence that supported this gene in L15 came from its association with APP metabolism modulation (Supplementary Fig. 42). Interestingly, this gene was a paralog of another prioritized gene, *ICA1L*, in L5 (discussed in below section).
- ***TMEM106B*** (L16). The risk allele of the lead variant in L16 was significantly associated with decreased *TMEM106B* expression in cortex (GTEx) and in naïve and stimulated monocytes (Fairfax dataset) (Supplementary Table 22 and Supplementary Fig. 34). Furthermore, the risk allele was also associated with increased methylation levels of cg09613507 intragenic CpG site in brain (Supplementary Table 29 and Supplementary Fig. 36a); however, when the effect of all variants were considered, MetaMeth implicated this CpG site as potentially protective for ADD when predicted methylation levels are increased in blood (Supplementary Table 28 and Supplementary Fig. 36b). Moreover, sTWAS implicated the increased predicted preference for chr7:12224385-12229679 splice junction (specifically present in longer *TMEM106B* transcripts including the canonical transcript) in cortex as protective for ADD risk (Supplementary Table 27 and Supplementary Fig. 40).
- ***JAZF1*** (L17). This gene was mainly prioritized by the colocalization of its eQTL signals in microglia with ADD genetic association signal. We observed eQTL coloc hits in MFG (PP4 = 77%), STG (PP4 = 89%), and THA (PP4 = 71%) (Supplementary Table 24 and Supplementary Fig. 37). Furthermore, MetaMeth results suggested that increased methylation in two intragenic CpGs can be protective for ADD (Supplementary Table 28 and Supplementary Fig. 36b).
- ***CTSB*** (L19). In this locus, the lead variant is a low-frequency 3' UTR variant in *CTSB*, and the risk allele was found to be associated with decreased CTSB expression in DLPFC (Supplementary Table 22 and Supplementary Fig. 34a).
- ***ABCA1*** (L21). The risk allele of the lead variant in the locus was found to be associated with decreased methylation levels in brain for the CpG site cg14313833 that is 77 bp upstream of the TSS of *ABCA1* (Supplementary Table 29 and Supplementary Fig. 36a). The bibliographical data also strongly support implication of *ABCA1* in AD as ABCA1 overexpression reduces amyloid deposition in an AD-like mouse model[179] and the burden of rare variants in this gene was associated with AD risk[180].
- ***CTSH*** (L29). *CTSH* was prioritized with a high confidence as it was implicated by the numerous AD-driven modulations we observed, including i) the overlap of the lead variant with brain, macrophage, and monocyte eQTLs with considerably large effect sizes (where the risk allele was associated with increased *CTSH* expression) in most of the eQTL catalogues investigated, ii) the overlap of the lead variant with sQTLs in brain controlling *CTSH* splicing, and consequently high coloc PP4 values in these tissues (and in microglia additionally) both for eQTL and sQTL coloc, iii) the fine-mapped eTWAS hits (agreeing on the effect direction of the risk allele) and iv) the significant sTWAS hits for splice junctions in *CTSH* (Supplementary Table 22-27 and Supplementary Fig. 34-35, 37-40).
- ***MAF*** (L31). The risk allele of the intergenic lead variant was associated with decreased histone acetylation (H3K9Ac) levels (indicative for active chromatin) at an intergenic site (~14kb downstream of *MAF*) in brain (Supplementary Table 29 and Supplementary Fig. 36a). We found that this associated H3K9Ac peak was in line with the H3K27ac signature (measured at single-cell level in brain) in microglia and overlapped with a microglia-specific enhancer[181]. Of note, we observed that *MAF* expression was highly enriched in microglia (Supplementary Table 35).
- ***SIGLEC11*** (L38). *SIGLEC11* has very high colocalization probabilities (coloc PP4 93% to 98%) between the GWAS signal in the locus and its brain eQTL signal in six different catalogues investigated and also in microglia (PP4 = 98%), hinting at a possible association

between ADD risk and higher SIGLEC11 expression (Supplementary Table 24 and Supplementary Fig. 37). We also observed the lead variant as a significant eQTL in many brain regions, microglia, macrophages, and monocytes (Supplementary Table 22 and Supplementary Fig. 34). Even though eTWAS signals failed to pass stringent Bonferroni-corrected significance thresholds, statistical fine-mapping mapped *SIGLEC11* to the 90% credible set of plausible candidate genes (with FOCUS PIP values 0.39 to 0.64) in this genomic locus.

- ***RBCK1*** (L40). We identified that the lead variant overlapped with significant eQTLs in temporal and frontal lobe, showing a *RBCK1* expression decreasing effect for the risk allele. Consequently, we observed a fine-mapped significant eTWAS hit (supported also by eQTL coloc PP4 of 99%) in DLPFC suggesting a potential protective effect of increased predicted *RBCK1* expression for ADD (Supplementary Table 22, 24, 26 and Supplementary Fig. 34a, 37, 39).

***<u>Prioritized genes with a protein-altering lead variant</u>***

Moreover, five other nearest genes in the new loci can be prioritized since the lead variant corresponds to a protein-altering variant (predicted deleterious missense or inframe deletion):

- ***SORT1*** (L1, p.Lys302Glu, CADD = 22.8). This variant is located in the β-propeller domain of SORT1 involved in ligand binding. Of interest, it is located in the predicted ligand binding site for GRN[182] (L36). This same rare variant was previously found to be enriched in European patients with FTD[183].
- ***MME*** (L6, rs61762319; p.Met8Val, CADD = 23.1). We observed two independent lead variants in L6, and one of them (rs61762319) was a predicted deleterious missense variant. Interestingly, we identified that the other independent lead variant in *MME*, rs16824536, is a significant mQTL in DLPFC where the risk allele is significantly associated with higher methylated levels of cg25511593 CpG site within *MME* (Supplementary Table 29 and Supplementary Fig. 36a). This might suggest a complementary mechanism of action for both lead variants in a way that both predicted deleterious missense allele of rs61762319 and higher methylation levels (therefore expected potentially lower expression of *MME*) controlled by the risk allele of rs16824536 work potentially in the same direction for the disease risk.
- ***SHARPIN*** (L20; p.Ser17Phe, CADD = 19.06). We found additional evidence that higher ADD risk in the locus is associated with lower *SHARPIN* expression (fine-mapped eTWAS hit in GTEx brain BA24 region; Supplementary Table 26 and Supplementary Fig. 39) and that ADD risk is associated with the regulation of splicing of the first two exons of *SHARPIN* (sTWAS hits in AD-relevant GTEx brain regions; Supplementary Table 27 and Supplementary Fig. 40) which is related to the protective effect of increased preference of splice junctions specific to non-coding transcripts of *SHARPIN*, where the first exon containing the lead variant rs34173062 is spliced out.
- ***DOC2A*** (L30; p.Gly48Ser, CADD = 9.98). *DOC2A* locus was one of the most complex new locus we encountered as (i) it is among the most gene-dense loci, (ii) our methodology listed numerous genes in the locus with differing association patterns in different tissues and regions investigated, and additionally (iii) it is partially overlapping with *BCKDK* locus (that is ~1 Mb downstream) when 1 Mb extended coordinates are considered for analyses. While statistical fine-mapping of eTWAS and sTWAS prioritized numerous genes in different tissues (Supplementary Tables 26-27 and Supplementary Fig. 39-40), genetic driven methylation signals pointed mostly to *DOC2A* as 5 CpGs (3 found in promoter region: cg27151362, cg03890691, cg07041748) have positive blood-brain methylation correlations at least in >50% percentile and point at a protective effect of increased methylation for ADD risk (Supplementary Table 28 and Supplementary Fig. 36b). Altogether, increased *DOC2A* expression, decreased methylation (therefore potentially increased *DOC2A* expression), and increased preference for canonical splicing were all predicted to be associated with increased ADD risk; and in addition, it modulated APP metabolism (Supplementary Fig. 42). However, one should be careful with interpretation of

this complex locus 30 given that several other genes had high posterior probabilities of explaining the eTWAS association signal in the fine-mapping analysis of certain brain regions (Supplementary Fig. 39, 41d), suggesting that multiple risk genes can be potential candidates in this locus, including *YPEL3* and *INO80E* (Supplementary Tables 26-27 and Supplementary Fig. 39-40).

- ***WDR81*** (L34; p.Glu1033del, CADD = 16.37). The lead variant is a deleterious inframe deletion in *WDR81.* The additional evidence was obtained from its APP modulation effect, colocalization of its eQTLs in temporal cortex with ADD genetic association signal (PP4 = 78%), and colocalization of its sQTLs for a cryptic splicing event (chr17:1732831-1733303) in DLPFC with ADD genetic association signal (PP4 = 79%) (Supplementary Tables 24-25 and Supplementary Fig. 37-38, 42). However, we identified that *SERPINF2* (downstream to *WDR81*) also modulates APP, but with a rather lower log2 fold-change in mCherry signal compared to *WDR81* (Supplementary Fig. 42), and the sQTLs for canonical skipping (chr17:1745395-1747019) of exon 5 colocalized with ADD genetic association signal in DLPFC (coloc PP4 = 95%) (Supplementary Table 25 and Supplementary Fig. 38). Of note, we also observed cryptic splicing events in a repetitive non-coding region between *WDR81* and *SERPINF2* whose sQTLs greatly colocalized with ADD genetic association signal (Supplementary Table 25 and Supplementary Fig. 38).

***Prioritized genes not meeting any top priority criteria***

For seven loci, none of the genes presented any top priority criteria, therefore we considered that their proximity to the lead variant was in favor of their prioritization but at this stage at a lower level of confidence for most of them:

- ***NCK2*** (L4), for which evidence is higher as the lead variant is rare.
- ***COX7C*** (L10)
- ***RASGEF1C*** (L12)
- ***HS3HT5*** (L13)
- ***UMAD1*** (L14)
- ***FOXF1*** (L32)
- ***APP*** (L42), an obvious candidate for AD. We could only detect the significant *APP* expression decreasing effect of the risk allele of the intronic lead variant rs2154481 in blood and monocytes (Supplementary Table 22 and Supplementary Fig. 34a, 34b), but not in AD-relevant bulk brain regions or in microglia; instead we identified *CYYR1-AS1,* a non-coding transcript, in between *APP* and *ADAMTS1* as having brain eQTL coloc & fine-mapped eTWAS signals, where its predicted increased expression is associated with increased ADD risk (Supplementary Tables 24, 26 and Supplementary Fig. 37, 39).

***Prioritized genes in complex loci***

We could efficiently prioritize candidate risk genes in twelve additional loci with a more complicated pattern in a way that several genes exhibit AD-related modulations in the same locus, and/or the prioritized gene is not the nearest protein-coding gene:

- ***ADAM17*** (L2). The lead variant is positioned in the promoter region of *ADAM17*. Its eQTL signal colocalized with ADD genetic association signal (coloc PP4 values of 73% both in BA22 and in BA10) (Supplementary Table 24 and Supplementary Fig. 37) meanwhile eTWAS showed a potential protective effect of increased *ADAM17* expression for ADD (though, it could not pass stringent Bonferroni-correction). In addition, the ADD signal colocalized with the sQTLs that are controlling the splicing of the proximal first exons of *ADAM17* in both temporal and frontal lobe, which was also supported by sTWAS results (Supplementary Tables 25, 27 and Supplementary Fig. 38, 40).
- ***ICA1L*** (L5). In this locus, the lead variant is found in 3' UTR of *WDR12* however it is also ~7kb upstream from the transcription start site of *ICA1L*. eQTL mapping, eQTL coloc, and eTWAS results output multiple genes in the locus whose predicted upregulation are seemingly co-regulated by the ADD-associated LD block in the locus, meanwhile fine-mapping of eTWAS results prioritizes *ICA1L*, *WDR12*, *CARF* and *NBEAL1* in the different brain regions assessed (Supplementary Tables 22, 24, 26 and Supplementary Fig. 34a, 37,

39). Two of these genes, *ICA1L* and *CARF*, were additionally prioritized because of the multiple AD-related splicing modulations in sTWAS, meanwhile we also observed the lead variant as a significant sQTL for chr2:202819899-202828848 splice junction in *ICA1L* in TCX and DLPFC, whose sQTL signal also colocalized with ADD association signal (Supplementary Tables 23, 25, 27 and Supplementary Fig. 35, 38, 40). This splice junction is specific to known protein-coding short transcript of *ICA1L* (ENST00000418208.5) that is about 7 times smaller than the canonical isoform, and our data shows an increased ADD risk correlated with the increased predicted preference of this junction (Supplementary Table 27 and Supplementary Fig. 40). Furthermore, we identified that *ICA1L* modulated APP metabolism (Supplementary Fig. 42), and interestingly it is a paralog of another gene in our study, *ICA1* (in L15), that was also prioritized due to its effect on APP metabolism. Taken together, in locus 5, we prioritized *ICA1L* however we also retained *CARF* as a candidate gene but at a lower confidence compared to *ICA1L*.

- ***RHOH*** (L8). We prioritized the nearest gene *RHOH* as the risk allele of the lead variant is significantly associated with decreased *RHOH* expression in TCX and as we observed colocalization of *RHOH* eQTL signals in TCX with ADD genetic association signal (PP4 = 98%) (Supplementary Tables 22, 24 and Supplementary Fig. 34, 37).
- ***OTULIN*** (L9). We could prioritize *OTULIN* with a very high confidence even though it is not the nearest protein-coding gene, because all the results of our assessments in this locus pointed out at *OTULIN* exclusively, including significant eQTL - lead variant overlap, eQTL coloc, and fine-mapped eTWAS results in brain (Supplementary Tables 22, 24, 26 and Supplementary Fig. 34a, 37, 39); all hinting at a correlation between increased ADD risk and predicted higher expression of *OTULIN*.
- ***EGFR*** (L18). In the intergenic signal of L18, *EGFR* is the exclusively prioritized gene because an intergenic, distant, and low-frequency cis-eQTL signal for *EGFR* which colocalizes with the ADD association signal (near ~1 coloc PP4s), and its fine-mapped eTWAS hits (with FOCUS PIP values of ~1) associate predicted increased *EGFR* expression with increased ADD risk both in TCX and DLPFC (see Fig 3; Supplementary Tables 22, 24, 26 and Supplementary Fig. 34a, 37, 39).
- ***CCDC6*** (L22). Between the two genes of interest (*ANK3* and *CCDC6*), *ANK3* is the nearest gene to the lead variant in the locus. The lead variant is a significant eQTL for both genes in DLPFC where the risk allele is associated with increased expression for both genes (Supplementary Table 22 and Supplementary Fig. 34a). However, even though not passing stringent Bonferroni-corrected thresholds, predicted higher expression of *CCDC6* suggestively correlated with higher ADD risk in the locus, and *CCDC6* was also placed among the 90% credible gene set in the fine-mapping of eTWAS results in TCX and DLPFC brain regions. Moreover, importantly, we identified a colocalization between *CCDC6* eQTL and ADD genetic association signal in microglia (MiGA Meta coloc PP4 = 81%) (Supplementary Table 24 and Supplementary Fig. 37). This is in line with the observation in a recent AD GWAS where monocyte eQTL signal of *CCDC6* colocalized with the AD signal[118]. Thus, based on the current evidence, *CCDC6* can be prioritized in this locus at a higher confidence, meanwhile we do not fully rule out the possibility for *ANK3* being the risk gene in the locus.
- ***TSPAN14*** (L23). *TSPAN14* was identified as the candidate risk gene as it exhibited numerous AD-related expression, methylation, and splicing modulations. The protective signal is associated with decreased *TSPAN14* expression and increased preference for cryptic splice junctions (within ADAM10-interacting domain) that we identified and experimentally confirmed by long-read single-molecule sequencing in brain and LCL samples (see Fig. 3; Supplementary Tables 22-28 and Supplementary Fig. 34-40, 41c). TSPAN14 was reported to regulate the trafficking and the function of the metalloprotease ADAM10[184].
- ***BLNK*** (L24). *BLNK* was supported by significant and fine-mapped eTWAS results in brain (FOCUS PIP values of 0.96 to 0.98) with considerable eQTL colocalization in DLPFC (PP4 = 97%) and importantly in microglia (MiGA Meta PP4 = 98%), predicting a risk-

increasing effect of increased *BLNK* expression (Supplementary Tables 22, 24, 26 and Supplementary Fig. 34, 37, 39).

- ***PLEKHA1*** (L25). *PLEKHA1* was supported by eQTL overlap with the lead variant in DLPFC and microglia (the risk allele is associated with increased expression), sQTL overlap (the risk allele is associated with decreased preference) and sQTL coloc hit in DLPFC (PP4 = 82) for its splice junction of chr10:122428316-122429624 that is related to alternative splicing of the last coding exon, and most importantly substantial eQTL coloc hits in microglia (PP4s of 88%, 92%, and 97% respectively in MFG, SVZ, and meta-analysis) (Supplementary Tables 22-25 and Supplementary Fig. 34, 35, 37, 38).
- ***RITA1*** (L26). Even though the risk allele of the lead variant is not frequent, we were able to identify significant and consistent eQTL association of it with decreased *RITA1* expression across all investigated brain regions except for BA10 and across several monocyte eQTL catalogues; and with increased *IQCD* and decreased *TPCN1* expression only in DLPFC. Moreover, eQTL coloc and eTWAS results mainly supported *RITA1* and *IQCD* (where increased ADD risk was correlated with decreased predicted *RITA1* expression and increased predicted *IQCD* expression), however implicated *RITA1* in more brain regions and with stronger probabilities. Therefore, in this locus we prioritized *RITA1* based on the current evidence, but we also accounted for possibility of *IQCD* being the candidate gene in the locus at a lower confidence (Supplementary Tables 22, 24, 26 and Supplementary Fig. 34a, 37, 39).
- ***MYO15A*** (L35). *MYO15A* and *LLGL1* were first prioritized based on the number of significant associations in AD-driven expression and splicing analysis results. We then assigned *MYO15A* as the main candidate risk gene in the locus especially because of consistent eQTL coloc hits for this gene across all brain regions assessed (with between 72% - 85% eQTL coloc PP4), and we retained *LLGL1* as a secondary candidate in the locus (eQTL coloc PP4 of 96% in DLPFC) (Supplementary Tables 22-25 and Supplementary Fig. 34, 35, 37, 38).
- ***GRN*** (L36). Our analyses initially prioritized *GRN*, *FAM171A2*, *ITGA2B* and *PLCD3*. However, we observed a close to 100% probability for *GRN* eQTL signal colocalization with the genetic association signal in 5 out of 6 brain regions investigated, and fine-mapping of eTWAS strongly pointed towards *GRN* as the gene explaining the GWAS signal at this locus (FOCUS PIP=1 in 5 frontal and 1 temporal lobe prediction panels tested), suggesting a correlation between predicted lower *GRN* expression and increased ADD risk (Supplementary Tables 22, 24, 26 and Supplementary Fig. 34a, 37, 39, 41a). Bibliographical data were also clearly in favor of *GRN*: beyond its implication in frontotemporal dementia, *GRN* deficiency significantly reduces diffuse Aβ plaque growth in an AD-like mouse model and it has been proposed that this protective effect is due, in part, to enhanced microglial Aβ phagocytosis[185]. Moreover, rare variants in *GRN* have previously been associated with AD[186,187]. Consequently, *GRN* is highly likely to be the candidate gene in this locus.

***No obvious single prioritized gene in complex loci***

In the remaining eight loci, we did not clearly identify a single candidate; however the current evidence pointed towards certain multiple candidate genes in four loci (L7, L28, L37, and L41), or a single gene was prioritized based on the lack of relevant AD-related modulations and the presence of convincing bibliographical evidence (L39):

- ***SLC26A1***, ***DGKQ***, and ***IDUA*** (L7). Among the 7 genes of interest, the three most proximal genes to the lead variant obtained similarly high number of hits. The lead variant rs3822030 is positioned in a regulatory active promoter region of *SLC26A1* and intron of *IDUA*, and the risk allele was found to be significantly associated with increased methylation levels of a CpG site in proximity (cg21616051) as mQTL (Supplementary Table 29 and Supplementary Fig. 36a). In brain, eQTL coloc & fine-mapped eTWAS results mainly prioritized *SLC26A1* over the other two genes, suggesting a *SLC26A1* expression decreasing effect on the ADD risk in the locus through a plausible mechanism affecting its promoter sequence (Supplementary Tables 22, 24, 26 and Supplementary Fig. 34a, 37, 39). Moreover, *DGKQ* was also of interest as its eQTLs colocalized with ADD genetic association signal uniquely among *SLC26A1*, *DGKQ* and *IDUA* in frontal lobe (near 100% coloc PP4 in DLPFC) and as we also observed significant eTWAS association in EADB Belgian LCL dataset (however in risk-increasing direction contrary to its predicted brain expression effect) (Supplementary Tables 22, 24, 26 and Supplementary Fig. 34a, 37, 39). In addition, DGKQ underexpression was associated with the modulation of the APP metabolism (Supplementary Fig. 42). Furthermore, sTWAS associated 5 splice junctions in *IDUA* (sQTL signal for 3 of these also colocalized with ADD signal) and 2 splice junctions in *DGKQ* with genetic ADD risk in the locus (Supplementary Tables 25, 27 and Supplementary Fig. 38, 40). Taken together, in this complex locus *DGKQ, SLC26A1, IDUA* are likely candidate genes that warrant further investigation.

- ***SNX1*** and ***FAM96A*** (L28). Among the 8 candidate genes presenting AD-related modulations in 1 Mb around the lead variant, *APH1B* (~820kb away from the lead variant) has very significant eQTL coloc, eTWAS and sTWAS hits in our analyses (Supplementary Tables 24, 26, 27 and Supplementary Fig. 37, 39, 40). However, we previously determined that *SNX1* and *APH1B* GWAS signals in this locus were independent (Supplementary Tables 3 and 4). The risk allele of the lead variant rs3848143 is also associated with decreased *SNX1* expression in BA44, corroborated by eTWAS fine mapping for this brain region (Supplementary Tables 22, 26 and Supplementary Fig. 34a, 39). We additionally identified multiple sTWAS hits across different AD-relevant brain regions in proximity of *SNX1* lead variant, including in *FAM96A* and *CSNK1G1* which were both related to canonical exon-skipping splice events that were predicted to be protective for ADD (Supplementary Table 27 and Supplementary Fig. 40). However, between these two genes, we retained *FAM96A* as an additional candidate (together with *SNX1*) because we also observed that the lead variant was also a significant sQTL for regulation of the above mentioned splice junctions in *FAM96A*, additional lead variant eQTL effects for *FAM96A* were found in monocytes and in LCL, and *FAM96A* had a significant hit in GTEx LCL panel of eTWAS (suggesting a protective effect of increased *FAM96A* expression) (Supplementary Tables 22, 23, 26 and Supplementary Fig. 34, 35, 39).
- ***ATP8B3***, ***REXO1***, and ***KLF16*** (L37). The lead variant is an eQTL for these three genes in AD-related brain regions (and additionally in macrophages for *REXO1*) (Supplementary Table 22 and Supplementary Fig. 34a). MetaMeth also implicated a CpG within *ATP8B3* whose decreased predicted methylation is associated with increased ADD risk (Supplementary Table 28 and Supplementary Fig. 36b). Finally, we observed fine-mapped *KLF16* eTWAS associations (Supplementary Table 26 and Supplementary Fig. 39) in TCX and DLPFC that are independent of *POLR2E* eTWAS associations (Supplementary Fig. 41b) that are likely driven by another ADD risk locus that is positioned upstream, *ABCA7* locus. However; similarly, we also observed fine-mapped eTWAS hits in other panels investigated, such as *ATP8B3* in GTEx hippocampus, cortex, and BA24 (where increased predicted *ATP8B3* expression was correlated with increased ADD risk), and *REXO1* in GTEx frontal cortex (where increased predicted *REXO1* expression was correlated with decreased ADD risk). Taken together, the current evidence does not allow us to further prioritize one of these three genes over another.
- ***LILRB2*** (L39). In brain we could only find a considerable eQTL coloc for *MYADM* gene (~400kb from the lead variant) in BA44 region (coloc PP4 = 93%) (Supplementary Table 24 and Supplementary Fig. 37). Moreover, the risk allele of the lead variant rs587709 was associated with decreased expression of *LILRB2* in blood in both GTEx and eQTLGen (Supplementary Fig. 34a). Nevertheless, in L39 we prioritized *LILRB2* as the bibliographical data also supported *LILRB2*, as LilRb2 is an Aβ receptor and its murine homolog PirB is required for the deleterious effect of Aβ oligomers on hippocampal long-term potentiation in an AD-like mouse model[188]. In addition, molecules that inhibit Aβ-LilrB2 interactions in vitro and on the cell surface reduce Aβ cytotoxicity[189].
- ***RTEL1*** and ***LIME1*** (L41). In this *SLC2A4RG* locus, we did not obtain any significant results from e/sQTL coloc, e/sTWAS, and MetaMeth analyses. We thus considered the significant effects of the lead variants in molecular QTL catalogues; and we identified, uniquely in DLPFC, that the risk allele of the lead variant rs6742, a 3' UTR variant for

*SLC2A4RG*, is associated with decreased *LIME1* and *RTEL1* levels and associated with increased preference for the chr20:63689132-63689750 splice junction (a canonical exon skipping event as in the ENST00000425905.5 short transcript) in *RTEL1* (Supplementary Tables 22-23 and Supplementary Fig. 34-35). Moreover, we identified *LIME1* as a modulator of APP metabolism (Supplementary Fig. 42). Taken together, it is challenging to identify a single candidate risk gene, but current evidence points towards *LIME1* and *RTEL1* in this locus.

***No prioritized genes***

For the remaining three loci we did not prioritize any genes:

- ***PRKD3*** (L3). We only observed significant eQTL - lead variant overlap for *EIF2AK2* and *CEBPZOS*, and significant sQTL - lead variant overlap for two splice junctions in *NDUFAF7*. However, based on the lack of other type of QTL overlap, coloc, TWAS, and/or Metameth associations, we concluded that none of these genes could be prioritized at this level even in a suggestive way.
- ***IGH* gene cluster** (L27). Even though the lead variant is only associated with expression of numerous *IGH* cluster genes in blood, we had multiple significant hits, especially in sTWAS analyses. However, we observed that the majority of splice junctions identified in sTWAS are very long, complex, non-canonical splice events; thus likely a consequence of known fusion events[190] in this complex telomeric region of chromosome 14. Therefore, we did not prioritize any genes in this complex IGH cluster locus (Supplementary Tables 22-23, 25-27, Supplementary Fig. 34, 35, 38-40).
- ***PRDM7*** (L33). MetaMeth showed significant association between higher ADD risk and lower predicted methylation of cg06295223 (Supplementary Table 28 and Supplementary Fig. 36b, that is a CpG site whose methylation is positively correlated (50-75% percentile group) between blood and brain and located in the promoter region of *PRDM7*. eTWAS also identified a distant significant association for predicted expression of *CDK10* in GTEx frontal cortex (Supplementary Table 26 and Supplementary Fig. 39). Of note, the *TUBB8P7* pseudogene also presented significant AD-related modulations (Supplementary Table 22, 24, 26, 30 and Supplementary Fig. 34, 37, 39). In conclusion, we were not able to prioritize a single gene with a high confidence or multiple genes at a rather lower confidence in this locus based on the current results.

##### 10.3. Short description of some functions of the prioritized genes and their potential implication in AD

***SORT1*** (Sortilin 1) encodes a member of the VPS10-related sortilin family of proteins which also include SORL1. This protein is involved in the traffic of protein from the Golgi to the endosomes, secretory vesicles, and the cell surface[191]. GWAS revealed an association between *SORT1* and reduced plasma LDL-cholesterol (LDL-C) as well as reduced coronary artery disease (CAD)[192]. In AD, animal model studies suggest that sortilin is a beneficial protein for the reduction of amyloid pathology in APP/PS1 mice by promoting APP degradation[193]. SORT1 was also shown to be a neuronal receptor for GRN, down-regulating GRN extracellular levels under stress conditions[194].

***ADAM17*** (a disintegrin and metalloprotease domain 17) encodes a protein which belongs to the same family than ADAM10. As for the latter, ADAM17 has been proposed to carry an α-secretase activity which leads to the increasing secretion of soluble APP-α fragment and reduction of Aβ generation[195]. In addition, it has been proposed that TREM2 is shed via Adam17 proteolytic activity[196]. Finally, ADAM17 is also known as TACE (TNFα converting enzyme) and is involved in inflammatory processes[197]. A rare loss-of function in *ADAM17* has been associated with familial forms of late-onset AD[198].

***NCK2*** (non-catalytic (region of) tyrosine kinase adaptor protein-2) encodes a protein involved in integrin signaling and as a consequence, signaling to regulate survival, proliferation and cell shape as well as polarity, adhesion, migration and differentiation[199]**.** In neurons, nck2 has been involved in change of neuron morphology[200] and synaptic transmission[201]. NCK2 is also interacting with PSEN2 and EGFR.

***ICA1L*** (islet cell autoantigen 1 like) is a paralog of ICA1. This locus has been associated with small cerebral vessel disease[202]. It has been recently proposed that brain protein abundance of ICAL1 was genetically regulated in AD[203].

***CARF*** (calcium responsive transcription factor) encodes a protein which has been involved in synaptic plasticity[204] and brain development[205].

***MME*** (membrane metalloendopeptidase) encodes neprilysin (NEP). In vivo and cell culture experiments have shown that a decreased NEP level results in an increased Aβ level and vice versa. NEP has been proposed as one of the most prominent degrading Ab enzyme[206].

***DGKQ*** (diacylglycerol kinase theta) encodes a protein that, as other Diacylglycerol kinases, is an important regulator of lipid signaling and, consequently, important regulator of many diglyceride-dependent and phosphatidic acid-dependent proteins[207]. Ablation of the mammalian DGKQ orthologue, DGK-1 in *C. elegans*, prevents serotonin-mediated inhibition of neurotransmitter release at neuromuscular junctions suggesting that this protein is involved in synaptic transmission[208].

***SLC26A1*** (solute carrier family 26 member 1) belongs to the family of sulfate/anion transporter genes. Little is known about its functions in brain.

***IDUA*** (iduronidase alpha-L) encodes an enzyme that hydrolyzes the terminal alpha-L-iduronic acid residues of two glycosaminoglycans, dermatan sulfate and heparan sulfate. In *Idua* -/- mouse, a modulation of the APP metabolism was reported likely through cathepsin B activation[209].

***RHOH*** (ras homolog family member H) encodes a protein which acts as a negative regulator of cell growth and survival. The protein has been mainly involved in cancers[210].

***OTULIN*** encodes a deubiquitinase which is an essential negative regulator of inflammation and autoimmunity[211]. OTULIN causes a potentially fatal autoinflammatory pathology termed OTULIN-related autoinflammatory syndrome (ORAS)[212]. Importantly, overexpression of OTULIN favours microglia activation and neuroinflammation through inhibition of the NF-κB signaling pathway in cerebral ischemia/reperfusion rats[213]. OTULIN is a specific regulator of the LUBAC complex which is a major actor of the TNF signaling[214]. Moreover, OTULIN antagonizes the cargo loading, retromer binding, endosome to plasma membrane trafficking functions of SNX27 (sorting nexin 27)[215] which is a protein known to regulate β-amyloid production potentially in interaction with Sorl1 or Presenilin 1[216,217].

***COX7C*** (cytochrome c oxidase subunit 7C) encodes a protein part of the terminal component of the mitochondrial respiratory chain, and that catalyzes the electron transfer from reduced cytochrome c to oxygen.

***TNIP1*** (NFAIP3 interacting protein 1) encodes a protein which plays a role in autoimmunity and tissue homeostasis through the regulation of nuclear factor kappa-B activation. The TNIP1 locus has been associated with the risk of amyotrophic lateral sclerosis[218] and auto-immune diseases[219].

***RASGEF1C*** (Ras-GEF domain-containing family member 1C) encodes a specific activator of Rap2 which regulates cell-cell adhesion[220]. Little is known about its functions in brain.

***HS3ST5*** (heparan sulfate-glucosamine 3-sulfotransferase 5) encodes a protein involved in post-translational modifications. Heparan sulfate proteaglycans have been involved in multiple pathways in AD from abeta production[221] or Tau seeding[222] to neuroinflammation[223].

***UMAD1*** (UBAP1-MVB12-associated (UMA) domain containing 1) encodes a protein for which we know almost nothing (only 3 publications on PubMed).

***ICA1*** (islet cell autoantigen 1) encodes a protein which has been initially described as an autoantigen associated with autoimmune type 1 diabetes (T1D)[224]. In neurons, ICA1 was shown to be involved in the recruitment of AMPA receptors at the synapses[225].

***TMEM106B*** (transmembrane protein 106B) is well known to be associated with the risk of developing fronto-temporal dementia[134]. The corresponding protein has been involved in lysosomal dysfunction, myelin deficits[226], dendritic trafficking[227] or cell death[228].

***JAZF1*** (JAZF zinc finger 1) encodes a nuclear protein which functions as a transcriptional repressor. This gene has been associated with the risk of developing Type 2 diabetes and the protein regulates glucose and lipid homeostasis and inflammation[229].

***EGFR*** (epidermal growth factor receptor) encodes a cell surface protein that binds to epidermal growth factor. Activation of the EGFR enhances neurite growth and regeneration through SORL1 functions[230] and Presenilin 1 has been shown to regulate EGFR turnover and signaling in the endosomal-lysosomal pathway[231]. EGFR was also proposed as target for treating amyloid-β-induced memory loss[232].

***CTSB*** (cathepsin B) encodes a protein which is a lysosomal cysteine protease with both endopeptidase and exopeptidase activity. CTSB has been described to either participate to the production of pyroglutamate Aβ[233] or degrade amyloid-β in mice expressing APP[234]. In addition, oxidative stress has been proposed to activate NLRP3 through upregulating CTSB activity[235].

***SHARPIN*** (SHANK associated RH domain interactor). The missense variant associated with AD risk has been described to attenuate an inflammatory/immune response that may promote late-onset AD development[236]. A common variant has also been associated with neuroanatomical variation in the limbic system[237]. The corresponding protein has also been described as a novel postsynaptic density protein[238] and interestingly, SHARPIN is an endogenous inhibitor of β1-integrin activation[239] and is part of the core enzymatic LUBAC complex[214].

***ABCA1*** (ATP binding cassette subfamily A member 1) encodes a member of the superfamily of ATP-binding cassette (ABC) transporters which transport various molecules across extra- and intracellular membranes. ABCA1 deficiency affects Basal Cognitive Deficits and Dendritic Density in Mice[240]. In addition, ABCA1 Deficiency was shown to exacerbate blood-brain barrier and white matter damage after stroke[241]. Its overexpression was described to reduce amyloid deposition in an AD-like mouse model[179].

***CCDC6*** (coiled-coil domain containing 6) encodes a protein ubiquitously expressed which may be a tumor suppressor. A chromosomal rearrangement resulting in the expression of a fusion gene containing a portion of this gene with different protooncogenes has been reported[242]. Little is known about its potential function in the brain but this protein has been proposed to be involved in actin cytoskeleton rearrangement[243].

***ANK3*** (Ankyrin 3) encodes a protein (AnkG) which is part of a family that is believed to link the integral membrane proteins to the underlying spectrin-actin cytoskeleton. Neuronal expression of AnkG is higher in AD brains when compared with healthy control subjects. AnkG is present in exosomal vesicles, and it accumulates in β-amyloid plaques[244].

***TSPAN14*** (tetraspanin 14) encodes a protein which regulates the trafficking and function of ADAM10[184]. The TSPAN14 locus has been also associated with periventricular white matter hyperintensities[128].

***BLNK*** (B cell linker) encodes a protein which plays a critical role in B cell development and function[245], and plays an important role in PLCg2 activation, another genetic risk factor of AD[246]. BLNK is also significantly upregulated when exposed to Aβ[247].

***PLEKHA1*** (leckstrin homology domain containing A1) encodes a protein localized to the plasma membrane where it specifically binds phosphatidylinositol 3,4-bisphosphate. This protein may be involved in the formation of signaling complexes in the plasma membrane. The *PLEKHA1* locus has been associated with the risk of macular degeneration[248].

***RITA1*** (RBP-J interacting and tubulin associated) encodes a tubulin-binding protein that acts as a negative regulator of the Notch signaling pathway[249].

***IQCD*** (IQ motif containing D) encodes a protein for which we know almost nothing.

***FAM96A/CIAO2A*** (cytosolic iron-sulfur assembly component 2A) encodes a protein that has been described as a novel pro-apoptotic tumor suppressor[250].

***SNX1*** (sorting nexin 1) encodes a component of the retromer complex and is involved in several stages of intracellular trafficking. In particular, it has been described the participation of SNX1 in Sorl1 sorting[251], Sorl1 being a major genetic risk factor of AD and a major actor of the APP metabolism.

***CTSH*** (cathepsin H) encodes a lysosomal cysteine proteinase important in the overall degradation of lysosomal proteins. Interestingly, the induction of neuronal death by up-regulation of CTSH in microglia following LPS treatment has been reported[252] and CTSH has also been described to be over-expressed in microglia following Aβ exposure.

***TMEM219*** encodes a cell death receptor specific for IGFBP3 and may mediate caspase-8-dependent apoptosis[253].

***TAOK2*** (TAO kinase 2) encodes a serine/threonine protein kinase that is involved in many different processes, including, cell signaling, microtubule organization and stability, and apoptosis. More specifically, in drosophila, the corresponding ortholog activity regulates Dendritic Arborization, Cytoskeletal Dynamics, and Sensory Function[254]. TAOK2 is also involved in dendritic spine maturation[255] and its altered activity causes autism-related neurodevelopmental and cognitive abnormalities[256].

***INO80E*** encodes a protein belonging to a chromatin remodeling complex. Only 5 publications are available on PubMed and this gene has been potentially involved in schizophrenia[257].

***DOC2A*** (double C2 domain alpha) encodes a protein that is mainly expressed in the brain and potentially involved in Ca(2+)-dependent neurotransmitter release[258]. None is known about its potential implication in AD but a copy number variation of this gene has been associated with schizophrenia[259].

***PPP4C*** (protein phosphatase 4 catalytic subunit) encodes a protein that interacts with components of the Survival of Motor Neurons protein complex, which is functionally defective in the hereditary disorder spinal muscular atrophy[260].

***TBX6*** (T-box transcription factor 6) encodes a protein involved in transcription regulation which has been also involved in axon degeneration[261].

***YPEL3*** (yippee like 3) encodes a TP53-regulated protein which induces cellular senescence[262] and that is required for development of oligodendrocyte precursor cells[263].

***MAF*** (alias c-MAF) encodes for a transcriptional factor which appears to be mainly expressed in microglia. In this cell-type, adult microglia from p53-deficient mice have increased expression of this anti-inflammatory transcription factor[264].

***FOXF1*** **(Forkhead box F1)** belongs to the forkhead family of transcription factors. Its function in the brain is unknown.

***WDR81*** encodes a multi-domain transmembrane protein which is predominantly expressed in the brain and is thought to play a role in endolysosomal trafficking[265]. WDR81 regulates adult hippocampal neurogenesis[266].

***SERPINF2*** (serpin family F member 2) encodes a member of the serpin family of serine protease inhibitors. SERPINF2 exhibits CpG associations with AD risk and altered expression in AD brains[162].

***MYO15A*** (myosin XVA) encodes for an unconventional myosin. Mutations in these gene have been associated with hearing loss[267].

***LLGL1*** (LLGL scribble cell polarity complex component 1) encodes a protein that is part of a cytoskeletal network. LLGL1 directly binds to and promotes internalization of N-cadherin. Disruption of the N-cadherin-LLGL1 interaction during cortical development in vivo may lead to malformations of the cerebral cortex[268].

***GRN*** (progranulin) is a gene known to be responsible for monogenic forms of fronto-temporal dementia. GRN is mainly expressed in microglia and as BLNK, its expression is significantly upregulated when exposed to Aβ in microglia[247]. GRN deficiency significantly reduces diffuse Aβ plaque growth in an AD-like mouse model and it has been proposed that this protective effect is due, in part, to enhanced microglial Aβ phagocytosis[185].

***ATP8B3*** (ATPase phospholipid transporting 8B3) encodes a protein mainly expressed in testis and little is known about its function in the brain.

***REXO1*** (RNA exonuclease 1 homolog). Little is known about its function in general.

***KLF16*** (Krüppel-Like Factor 16) encodes a transcription factor that binds GC and GT boxes and displaces Sp1 and Sp3 from these sequences[269]. This transcriptional factor might be involved in dopaminergic transmission in the brain[270].

***SIGLEC11*** (sialic acid binding Ig like lectin 11) encodes a protein belonging to the immunoglobulin superfamily. Siglec-11, which mediates immunosuppressive signals, is only expressed in microglia and is the only Siglec protein expressed in this cell type. Salminen et al. has proposed the following hypothesis: “aggregating amyloid plaques are masked in AD by sialylated glycoproteins and gangliosides. Sialylation and glycosylation of plaques, mimicking the cell surface glycocalyx, can activate the immunosuppressive Siglec-11 receptors, as well as hiding the neuritic plaques, allowing them to evade the immune surveillance of microglial cells. This kind of immune evasion can prevent the microglial cleansing process of aggregating amyloid plaques in AD.”[271].

***LILRB2*** (leukocyte immunoglobulin like receptor B2) encodes a protein thought to control inflammatory responses and cytotoxicity to help focus the immune response and limit autoreactivity. LilRb2 has been described to be an Ab receptor and the murine homolog PirB is required for deleterious effect of Aβ oligomers on hippocampal long-term potentiation in an AD-like mouse model[188]. In addition, molecules that inhibit Aβ-LilrB2 interactions in vitro and on the cell surface, reduce Aβ cytotoxicity[189].

***RBCK1*** (RANBP2-type and C3HC4-type zinc finger containing 1) encodes a protein (HOIL-1) that is part of the core of the LUBAC complex[214]. This complex is the only known E3 ubiquitin-ligating enzyme producing M1 ubiquitin linkages de novo. This complex also involving OTULIN and SHARPIN is a crucial modulator of innate and adaptive immune responses, and act by regulating inflammatory and cell death signaling[214].

***RTEL1*** (telomere elongation helicase 1) encodes a DNA helicase which functions in the stability, protection and elongation of telomeres. Only three publications are available on PubMed.

***LIME1*** (Lck interacting transmembrane adaptor 1) encodes a transmembrane adaptor protein involved in signaling pathways via its association with the Src family kinases Lck and Lyn. LIME1 has been proposed to interact with Grb2[272], a major actor of the APP metabolism (this observation potentially allows to explain its impact on the APP function/metabolism[273]). In addition, this protein has been described to potentially interact with PlCg2[272]. However, since LIME1 is not expressed in microglia, the latter observation precludes its potential implication in AD though this interaction in microglia.

#### 11. STRING protein interaction analysis

The genes from the sets of interest (previous known genes and/or prioritized genes in Fig. 2, light and dark green) were tested for an excess of high-confidence protein-protein interactions as in[38]. First, a list of high-confidence (confidence score >0.7) human protein-protein interactions was downloaded from the latest version (v11.0)[274] of the STRING database (http://string-db.org). Then, a protein interaction network was generated for each of the genes as follows: (i) Choose a gene to start the network (the "seed" gene); (ii) For each remaining gene in the set of significant genes, add it to the network if its corresponding protein shows a high-confidence protein interaction with a protein corresponding to any gene already in the network; (iii) Repeat step 2 until no more gene can be added; (iv) Note the number of genes in the network; (v) Repeat, choosing each of the genes in turn as the seed gene; (vi) Note the size of the largest network.
To test whether the largest network was larger than expected by chance, given the total number of protein-protein interactions for each gene, 50,000 random sets of genes were generated, equal in number to the test set, with each gene chosen to have the same total number of protein-protein interactions as the corresponding gene in the actual data. Protein networks were generated for each gene as described above, and the size of the largest such network compared to that observed in the actual data. Excess interactions between two gene sets were tested by a similar approach.

#### 12. List of URLs

Bedtools: https://bedtools.readthedocs.io/en/latest/
BCFtools: http://samtools.github.io/bcftools/bcftools.html
Samtools: http:/www.htslib.org/doc/samtools.html
gene2go: ftp://ftp.ncbi.nlm.nih.gov/gene/DATA/
Gene Ontology: http://geneontology.org/docs/download-ontology/
Reactome: https://reactome.org/download-data
KEGG and Pathway Interaction Database (PID) pathways: https://www.gsea-msigdb.org/gsea/msigdb/index.jsp
AMP-AD rnaSeqReprocessing Study: https://www.synapse.org/#!Synapse:syn9702085
MayoRNAseq WGS VCFs: https://www.synapse.org/#!Synapse:syn11724002
ROSMAP WGS VCFs: https://www.synapse.org/#!Synapse:syn11724057
MSBB WGS VCFs: https://www.synapse.org/#!Synapse:syn11723899
GTEx pipeline: https://github.com/broadinstitute/gtex-pipeline
Leafcutter: https://github.com/davidaknowles/leafcutter
RegTools: https://github.com/griffithlab/regtools
Enhanced version of FastQTL: https://github.com/francois-a/fastqtl
Picard: https://broadinstitute.github.io/picard/
eQTLGen: https://www.eqtlgen.org/

eQTL Catalogue database: https://www.ebi.ac.uk/eqtl/
Brain xQTL serve: http://mostafavilab.stat.ubc.ca/xqtl/
MiGA eQTLs: https://doi.org/10.5281/zenodo.4118605
MiGA sQTLs: https://doi.org/10.5281/zenodo.4118403
MiGA Meta-analysis: https://doi.org/10.5281/zenodo.4118676
GTEx v8 eQTL and sQTL catalogues: https://www.gtexportal.org/
coloc: https://github.com/chr1swallace/coloc
FUSION: https://github.com/gusevlab/fusion_twas
GTEx v8 expression and splicing prediction models: http://predictdb.org/
MetaXcan: https://github.com/hakyimlab/MetaXcan
FOCUS: https://github.com/bogdanlab/focus
qcat: https://github.com/nanoporetech/qcat
minimap2: https://github.com/lh3/minimap2
NanoStat: https://github.com/wdecoster/nanostat
mosdepth: https://github.com/brentp/mosdepth
ggplot2: https://ggplot2.tidyverse.org/
LocusZoom: https://github.com/statgen/locuszoom-standalone
pyGenomeTracks: https://github.com/deeptools/pyGenomeTracks
BECon website: https://redgar598.shinyapps.io/BECon/
VCFs of phased biallelic SNV and INDEL variants of 1KG samples (de novo called on GRCh38):
ftp://ftp.1000genomes.ebi.ac.uk/vol1/ftp/data_collections/1000_genomes_project/release/20190312_biallelic_SNV_and_INDEL/

#### 14. Supplementary List of Authors

##### EADB

Jan Laczo[1,2], Vaclav Matoska[3], Maria Serpente[4], Francesca Assogna[5], Fabrizio Piras[5], Federica Piras[5], Valentina Ciullo[5], Jacob Shofany[5], Carlo Ferrarese[6,7], Simona Andreoni[6], Gessica Sala[6], Chiara Paola Zoia[6], Maria Del Zompo[8], Alberto Benussi[9], Patrizia Bastiani[10], Mari Takalo[11], Tiina Laatikainen[12,13,14], Jaakko Tuomilehto[12,15,16], Riitta Antikainen[17,18], Timo Strandberg[18], Jaana Lindström[12], Markku Peltonen[12], Richard Abraham[19], Ammar Al-Chalabi[20], Nicholas J. Bass[21], Carol Brayne[22], Kristelle S. Brown[23], John Collinge[24], David Craig[25], Pangiotis Deloukas[26], Nick Fox[27], Amy Gerrish[27], Michael Gill[28,29], Rhian Gwilliam[26], John Hardy[30], Denise Harold[31], Paul Hollingworth[19], Jarret A, Johnston[25], Lesley Jones[19], Brian Lawlor[28,29], Gill Livingston[32], Simon Lovestone[33], Michelle Lupton[34,35], Aoibhinn Lynch[28,29], David Mann[36], Andrew McQuillin[33], Bernadette McGuinness[25], Andrew McQuillin[32], Michael C. O'Donovan[19], Michael J. Owen[19], Peter Passmore[25], John F. Powell[34], Petra Proitsi[34], Martin Rossor[27], David C. Rubinsztein[37], Christopher E. Shaw[20], A. David Smith[38], Hugh Gurling[39], Stephen Todd[25], Catherine Mummery[40], Nathalie Ryan[40], Giordano Lacidogna[41], Astrid Daniela Adarmes-Gómez[42], Ana Mauleón[43], Ana Pancho[43], Anna Gailhajenet[43], Asunción Lafuente[43], Daniel Macias-García[42], Elvira Martín[43], Esther Pelejà[43], Fatima Carrillo[42], Isabel Sastre Merlín[44], Lorena Garrote-Espina[42], Liliana Vargas[43], Mario Carrion-Claro[42], Marta Marín[45], Miguel Angel Labrador[42], Mar Buendia[43], María Dolores Alonso[46], Marina Guitart[43], Mariona Moreno[43], Marta Ibarria[43], María Teresa

Periñán[42], Nuria Aguilera[43], Pilar Gómez-Garre[42], Pilar Cañabate[43], Rocio Escuela[42], Rocio Pineda-Sánchez[42], Rocio Vigo-Ortega[42], Silvia Jesús[42], Silvia Preckler[43], Silvia Rodrigo-Herrero[45], Susana Diego[43], Alessandro Vacca[47], Fausto Roveta[47], Nicola Salvadori[47], Elena Chipi[47], Henning Boecker[48,49], Christoph Laske[50,51], Robert Perneczky[52,53,54], Stefan Teipel[55], Costas Anastasiou [56], Daniel Janowitz[57], Rainer Malik[57], Anna Anastasiou[58], Kayenat Parveen[59], Carmen Lage[60], Sara López-García[60], Anna Antonell[61], Kalina Yonkova Mihova[62], Diyana Belezhanska[63], Heike Weber[64], Silvia Kochen[65], Patricia Solis[65], Nancy Medel[65], Julieta Lisso[65], Zulma Sevillano[65], Daniel G Politis[65,66], Valeria Cores[65,66], Carolina Cuesta[65,66], Cecilia Ortiz[67], Juan Ignacio Bacha[67], Mario Rios[68], Aldo Saenz[68], Mariana Sanchez Abalos[69], Eduardo Kohler[70], Dana Lis Palacio[71], Ignacio Etchepareborda[71], Matias Kohler[71], Gisela Novack[72], Federico Ariel Prestia[72], Pablo Galeano[72], Eduardo M. Castaño[72], Sandra Germani[73], Natividad Olivar[73], Carlos Reyes Toso[73], Matias Rojo[73], Carlos Ingino[73], Carlos Mangone[73], Nathalie Fievet[74], Vincent Deramecourt[75], Charlotte Forsell[76,77], Håkan Thonberg[76,77], Ellen De Roeck[78], Maria Bjerke[78], María Teresa Martínez-Larrad[79]

1. Memory Clinic, Department of Neurology, Charles University, 2nd Faculty of Medicine and Motol University Hospital, Czech Republic
2. International Clinical Research Center, St. Anne's University Hospital Brno, Brno, Czech Republic
3. Department of Clinical Biochemistry, Hematology and Immunology, Na Homolce, Czech Republic
4. University of Milan, Milan, Italy
5. Laboratory of Neuropsychiatry, Department of Clinical and Behavioral Neurology, IRCCS Santa Lucia Foundation, Rome, Italy
6. School of Medicine and Surgery, University of Milano-Bicocca, Italy
7. Neurology Unit, "San Gerardo" hospital, Monza, Italy
8. Department of Biomedical Sciences, Section of Neuroscience and Clinical Pharmacology, University of Cagliari, Italy
9. Centre for Neurodegenerative Disorders, Department of Clinical and Experimental Sciences, University of Brescia, Brescia, Italy
10. Institute of Gerontology and Geriatrics, Department of Medicine, University of Perugia Perugia, Italy
11. Institute of Biomedicine, University of Eastern Finland, Finland
12. Public Health Promotion Unit, Finnish Institute for Health and Welfare, Helsinki, Finland
13. Institute of Public Health and Clinical Nutrition, University of Eastern Finland, Kuopio, Finland
14. JointMunicipal Authority for North Karelia Social and Health Services (Siun Sote), Joensuu, Finland
15. Department of Public Health, University of Helsinki, Helsinki, Finland
16. National School of Public Health, Madrid, Spain
17. Center for Life Course Health Research, University of Oulu, Oulu, Finland
18. Medical Research Center Oulu, Oulu University Hospital, Oulu, Finland
19. Division of Psychological Medicine and Clinial Neurosciences, MRC Centre for Neuropsychiatric Genetics and Genomics, Cardiff University, UK
20. Kings College London, Institute of Psychiatry, Psychology and Neuroscience, London, UK
21. Division of Psychiatry, University College London, UK
22. Institute of Public Health, University of Cambridge, Cambridge, UK
23. Institute of Genetics, Queens Medical Centre, University of Nottingham, Nottingham, UK
24. MRC Prion Unit at UCL, Institute of Prion Diseases, London
25. Ageing Group, Centre for Public Health, School of Medicine, Dentistry and Biomedical Sciences, Queen's University Belfast, UK
26. The Wellcome Trust Sanger Institute, Wellcome Trust Genome Campus, Hinxton, Cambridge, UK.
27. Dementia Research Centre, Department of Neurodegenerative Disease, UCL Institute of Neurology, London, UK
28. Mercer's Institute for Research on Ageing, St James' Hospital, Dublin, Ireland
29. James Hospital and Trinity College, Dublin, Ireland
30. Department of Molecular Neuroscience, UCL, Institute of Neurology, London, UK
31. School of Biotechnology, Dublin City University, Dublin, Ireland.
32. Division of Psychiatry, University College London, UK

33. Department of Psychiatry, University of Oxford, Oxford, UK
34. Department of Basic and Clinical Neuroscience, Institute of Psychiatry, Psychology and Neuroscience, Kings College London, London UK
35. Genetic Epidemiology, QIMR Berghofer Medical Research Institute, Herston, Queensland, Australia
36. Division of Neuroscience and Experimental Psychology, School of Biological Sciences, Faculty of Biology, Medicine and Health, University of Manchester, Manchester Academic Health Science Centre, Manchester, UK
37. Cambridge Institute for Medical Research and UK Dementia Research Institute, University of Cambridge, Cambridge, UK
38. Oxford Project to Investigate Memory and Ageing (OPTIMA), University of Oxford, Level 4, John Radcliffe Hospital, Oxford, UK
39. Department of Mental Health Sciences, University College London, London, UK
40. Dementia Research Centre, UCL, London, UK
41. Department of Neuroscience, Catholic University of Sacred Heart, Fondazione Policlinico Universitario A. Gemelli IRCCS, Rome, Italy
42. Unidad de Trastornos del Movimiento, Servicio de Neurología y Neurofisiología. Instituto de Biomedicina de Sevilla (IBiS), Hospital Universitario Virgen del Rocío/CSIC/Universidad de Sevilla, Seville, Spain
43. Research Center and Memory clinic Fundació ACE, Institut Català de Neurociències Aplicades, Universitat Internacional de Catalunya, Barcelona, Spain
44. Centro de Biología Molecular Severo Ochoa (UAM-CSIC), Spain
45. Unidad de Demencias, Servicio de Neurología y Neurofisiología. Instituto de Biomedicina de Sevilla (IBiS), Hospital Universitario Virgen del Rocío/CSIC/Universidad de Sevilla, Seville, Spain
46. Servei de Neurologia. Hospital Clínic Universitari de València, Valencia, Spain
47. Centre for Memory Disturbances, Lab of Clinical Neurochemistry, Section of Neurology, University of Perugia, Italy
48. German Center for Neurodegenerative Diseases (DZNE), Bonn, Venusberg-Campus 1, 53127 Bonn, Germany
49. Department of Radiology, University Hospital Bonn, Bonn, Germany
50. German Center for Neurodegenerative Diseases (DZNE), Tübingen, Germany
51. Section for Dementia Research, Hertie Institute for Clinical Brain Research and Department of Psychiatry, Tübingen, Germany
52. German Center for Neurodegenerative Diseases (DZNE, Munich), Feodor-Lynen-Strasse 17, 81377 Munich, Germany
53. Department of Psychiatry and Psychotherapy, University Hospital, LMU Munich, Munich, Germany
54. Munich Cluster for Systems Neurology (SyNergy) Munich, Munich, Germany
55. German Center for Neurodegenerative Diseases (DZNE), Rostock, Germany
56. Department of Nutrition and Diatetics, Harokopio University, Athens, Greece
57. Institute for Stroke and Dementia Research (ISD),University Hospital, Ludwig-Maximilian University Munich, Germany
58. 1st Department of Neurology, Medical school, Aristotle University of Thessaloniki, Thessaloniki, Makedonia, Greece
59. Division of Neurogenetics and Molecular Psychiatry, Department of Psychiatry and Psychotherapy, University of Cologne, Medical Faculty, 50937 Cologne, Germany.
60. Service of Neurology, University Hospital Marqués de Valdecilla, IDIVAL, University of Cantabria, Santander, Spain
61. Neurology department-Hospital Clínic, IDIBAPS, Universitat de Barcelona, Barcelona, Spain.
62. Molecular Medicine Center, Department of Medical chemistry and biochemistry, Medical University of Sofia, Bulgaria
63. Clinic of Neurology, UH "Alexandrovska", Medical University - Sofia, Sofia, Bulgaria
64. Department of Psychiatry, Psychosomatics and Psychotherapy, Center of Mental Health, University Hospital of Würzburg, Germany
65. ENYS (Estudio en Neurociencias y Sistemas Complejos) CONICET- Hospital El Cruce "Nestor Kirchner"- UNAJ, Argentina
66. HIGA Eva Perón, Buenes Aires, Agentina
67. Neurología Clinica, Buenes Aires, Agentina

68. Dirección de Atención de Adultos Mayores del Min. Salud Desarrollo Social y Deportes de la Pcia. de Mendoza, Argentina
69. Laboratorio de Genética Forense del Ministerio Público de la Pcia. de La Pampa, Argentina
70. Fundacion Sinapsis, Santa Rosa, Argentina
71. Hospital Dr. Lucio Molas, Santa Rosa; Fundacion Ayuda Enfermo Renal y Alta Complejidad (FERNAC), Santa Rosa, Argentina
72. Laboratory of Brain Aging and Neurodegeneration- FIL, Buneos Aires, Argentina
73. Centro de Neuropsiquiatría y Neurología de la Conducta (CENECON), Facultad de Medicina, Universidad de Buenos Aires (UBA), C.A.B.A, Buenos Aires, Argentina.
74. Univ. Lille, Inserm, CHU Lille, Institut Pasteur de Lille, U1167-RID-AGE facteurs de risque et déterminants moléculaires des maladies liés au vieillissement, Lille, France.
75. Univ Lille Inserm 1171, CHU Clinical and Research Memory Research Centre (CMRR) of Distalz Lille France.
76. Karolinska Institutet, Center for Alzheimer Research, Department NVS, Division of Neurogeriatrics, 171 64 Stockholm Sweden
77. Unit for Hereditary Dementias, Theme Aging, Karolinska University Hospital-Solna, 171 64 Stockholm Sweden
78. Center for Neurosciences, Vrije Universiteit Brussel (VUB), Brussels, Belgium
79. Centro de Investigación Biomédica en Red de Diabetes y Enfermedades Metabólicas Asociadas, CIBERDEM, Spain, Hospital Clínico San Carlos, Madrid, Spain

#### ADGC

Erin Abner[1], Perrie Adams[2], Alyssa Aguirre[3], Marilyn Albert[4], Roger Albin[5,6], Mariet Allen[7], Lisa Alvarez[8], Liana Apostolova[9,10,11], Steven Arnold[12], Sanjay Asthana[13,14,15], Craig Atwood[14,15], Gayle Ayres[3], Robert Barber[8], Lisa Barnes[16,17,18], Sandra Barral[19,20,21], Thomas Beach[22], James Becker[23], Gary Beecham[24,25], Bruno Benitez[26], David Bennett[16,18], John Bertelson[27], Eileen Bigio[28,29], Thomas Bird[30,31], Deborah Blacker[32,33], Bradley Boeve[34], James Bowen[35], Adam Boxer[36], James Brewer[37], James Burke[38], Jeffrey Burns[39], Joseph Buxbaum[40,41,42], Laura Cantwell[43], Chuanhai Cao[44], Cynthia Carlsson[13,14,15], Regina Carney[45], Minerva Carrasquillo[7], Scott Chasse[46], Marie-Francoise Chesselet[47], Nathaniel Chin[13,14], Helena Chui[48], Jaeyoon Chung[49], Suzanne Craft[50], Paul Crane[51], Elizabeth Crocco[45], Carlos Cruchaga[26], Michael Cuccaro[24,25,26], Munro Cullum[2,52,53], Eveleen Darby[54], Barbara Davis[53], Philip De Jager[55,56], Charles DeCarli[57], John DeToledo[58], Dennis Dickson[7], Ranjan Duara[59], Nilufer Ertekin-Taner[7,60], Denis Evans[61], Kelley Faber[10], Thomas Fairchild[62], Kenneth Fallon[63], David Fardo[64], Martin Farlow[11], John Farrell[49], Victoria Fernandez-Hernandez[26], Tatiana Foroud[10], Matthew Frosch[65], Douglas Galasko[37], Adriana Gamboa[66], Marla Gearing[67,68], Daniel Geschwind[47], Bernardino Ghetti[69], John Gilbert[24,25], Alison Goate[40], Thomas Grabowski[30], Neill Graff-Radford[7,60], Robert Green[70], John Growdon[71], Hakon Hakonarson[72], James Hall[8], Ronald Hamilton[73], Oscar Harari[26], Lindy Harrell[74], Elizabeth Head[75], Victor Henderson[76,77], Michelle Hernandez[58], Lawrence Honig[19,20], Ryan Huebinger[52], Matthew Huentelman[78], Christine Hulette[79], Bradley Hyman[71], Linda Hynan[53], Laura Ibanez[26], Gail Jarvik[80,81], Suman Jayadev[30], Lee-Way Jin[82], Kim Johnson[58], Leigh Johnson[8], Gyungah Jun[83,84], M. Ilyas Kamboh[85,86], Anna Karydas[36], Mindy Katz[87], John Kauwe[88], Jeffrey Kaye[89,90], C. Dirk Keene[91], Aisha Khaleeq[54], Ronald Kim[75], Janice Knebl[66], Neil Kowall[92,93], Joel Kramer[94], Walter Kukull[95], Amanda Kuzma[43], Frank LaFerla[96], James Lah[97], Eric Larson[51,98], Alan Lerner[99], Yuk Yee Leung[43], James Leverenz[100], Allan Levey[97], Andrew Lieberman[101], Richard Lipton[87], Oscar Lopez[86], Kathryn Lunetta[83], Constantine Lyketsos[102], Douglas Mains[103,104], John Malamon[43], Logue Mark[49,105], Daniel Marson[74], Eliezer Masliah[37], Paul Massman[54], Arjun Masurkar[106], Wayne McCormick[51], Susan McCurry[107], Stefan McDonough[108], Ann McKee[92,93], Marsel Mesulam[28,109], Jesse Mez[92], Bruce Miller[110], Carol Miller[36], Thomas Montine[111], Edwin Monuki[75,112], John Morris[113,114], Shubhabrata Mukherjee[51], Amanda Myers[45], Trung Nguyen[53], Sid O'Bryant[66], John Olichney[57], Marcia Ory[115], Raymond Palmer[116], Joseph Parisi[117], Henry Paulson[5], Valory Pavlik[54], David Paydarfar[3], Victoria Perez[58], Ronald Petersen[33], Marsha Polk[116], Huntington

Potter[118], Liming Qu[43], Mary Quiceno[119], Joseph Quinn[89,90], Ashok Raj[44], Eric Reiman[78,120,121], Joan Reisch[53], Christiane Reitz[19,20,21], John Ringman[48], Erik Roberson[74], Monica Rodriguear[54], Ekaterina Rogaeva[122], Howard Rosen[36], Roger Rosenberg[123], Donald Royall[124], Mark Sager[14], Mary Sano[41], Andrew Saykin[9,10], Julie Schneider[16,18], Lon Schneider[48], William Seeley[36], Scott Small[19,20], Amanda Smith[53], Janet Smith[44], Salvatore Spina[69], Peter St George-Hyslop[122,125], Robert Stern[92], Alan Stevens[115], Stephen Strittmatter[126], David Sultzer[127], Russell Swerdlow[39], Rudolph Tanz[171], Jeffrey Tilson[46], Giuseppe Tosto[19,20], John Trojanowski[43], Juan Troncoso[128], Debby Tsuang[31,129], Otto Valladares[43], Vivianna Van Deerlin[43], Linda Van Eldik[119], Jeffery Vance[24], Badri Vardarajan[19,20,21], Robert Vassar[29,109], Harry Vinters[130], Jean Paul Vonsattel[131], Sandra Weintraub[28,29], Kathleen Welsh-Bohmer[38,132,133], Ellen Wijsman[80,81,134], Kirk Wilhelmsen[46], Benjamin Williams[119], Jennifer Williamson[19], Henrick Wilms[58], Thomas Wingo[97], Thomas Wisniewski[106,133], Randall Woltjer[134], Martin Woon[27], Steven Younkin[7], Lei Yu[16,18], Xiongwei Zhou[99], Congcong Zhu[49]

1. Sanders-Brown Center on Aging, College of Public Health, Department of Epidemiology, University of Kentucky, Lexington, Kentucky, USA
2. Department of Psychiatry, University of Texas Southwestern Medical Center, Dallas, Texas, USA
3. Department of Neurology, University of Texas at Austin/Dell Medical School, Austin, Texas,
4. Department of Neurology, Johns Hopkins University, Baltimore, Maryland,
5. Department of Neurology, University of Michigan, Ann Arbor, Michigan,
6. Geriatric Research, Education and Clinical Center (GRECC), VA Ann Arbor Healthcare System (VAAAHS), Ann Arbor, Michigan,
7. Department of Neuroscience, Mayo Clinic, Jacksonville, Florida,
8. Department of Pharmacology and Neuroscience, University of North Texas Health Science Center, Fort Worth, Texas,
9. Department of Radiology, Indiana University, Indianapolis, Indiana,
10. Department of Medical and Molecular Genetics, Indiana University, Indianapolis, Indiana,
11. Indian Alzheimer's Disease Center, Indiana University, Indianapolis, Indiana
12. Department of Psychiatry, University of Pennsylvania Perelman School of Medicine, Philadelphia, Pennsylvania,
13. Geriatric Research, Education and Clinical Center (GRECC), University of Wisconsin, Madison, Wisconsin,
14. Department of Medicine, University of Wisconsin, Madison, Wisconsin,
15. Wisconsin Alzheimer's Disease Research Center, Madison, Wisconsin,
16. Department of Neurological Sciences, Rush University Medical Center, Chicago, Illinois,
17. Department of Behavioral Sciences, Rush University Medical Center, Chicago, Illinois,
18. Rush Alzheimer's Disease Center, Rush University Medical Center, Chicago, Illinois,
19. Taub Institute on Alzheimer's Disease and the Aging Brain, Department of Neurology, Columbia University, New York, New York,
20. Gertrude H. Sergievsky Center, Columbia University, New York, New York,
21. Department of Neurology, Columbia University, New York, New York
22. Civin Laboratory for Neuropathology, Banner Sun Health Research Institute, Phoenix, Arizona,
23. Departments of Psychiatry, Neurology, and Psychology, University of Pittsburgh School of Medicine, Pittsburgh,
24. The John P. Hussman Institute for Human Genomics, University of Miami, Miami, Florida,
25. Dr. John T. Macdonald Foundation Department of Human Genetics, University of Miami, Miami, Florida,
26. Department of Psychiatry and Hope Center Program on Protein Aggregation and Neurodegeneration, Washington University School of Medicine, St. Louis, Missouri,
27. Department of Psychiatry, University of Texas at Austin/Dell Medical School, Austin, Texas
28. Department of Pathology, Northwestern University Feinberg School of Medicine, Chicago, Illinois
29. Cognitive Neurology and Alzheimer's Disease Center, Northwestern University Feinberg School of Medicine, Chicago, Illinois,
30. Department of Neurology, University of Washington, Seattle, Washington,
31. VA Puget Sound Health Care System/GRECC, Seattle, Washington,

32. Department of Epidemiology, Harvard School of Public Health, Boston, Massachusetts,
33. Department of Psychiatry, Massachusetts General Hospital/Harvard Medical School, Boston, Massachusetts,
34. Department of Neurology, Mayo Clinic, Rochester, Minnesota,
35. Swedish Medical Center, Seattle, Washington,
36. Department of Neurology, University of California San Francisco, San Francisco, California,
37. Department of Neurosciences, University of California San Diego, La Jolla, California,
38. Department of Medicine, Duke University, Durham, North Carolina,
39. University of Kansas Alzheimer’s Disease Center, University of Kansas Medical Center, Kansas City, Kansas,
40. Department of Neuroscience, Mount Sinai School of Medicine, New York, New York,
41. Department of Psychiatry, Mount Sinai School of Medicine, New York, New York,
42. Mount Sinai School of Medicine, New York, New York,
43. Penn Neurodegeneration Genomics Center, Department of Pathology and Laboratory Medicine, University of Pennsylvania Perelman School of Medicine, Philadelphia, Pennsylvania,
44. USF Health Byrd Alzheimer's Institute, University of South Florida, Tampa, Florida,
45. Department of Psychiatry and Behavioral Sciences, Miller School of Medicine, University of Miami, Miami, Florida,
46. Department of Genetics, University of North Carolina Chapel Hill, Chapel Hill, North Carolina,
47. Neurogenetics Program, University of California Los Angeles, Los Angeles, California,
48. Department of Neurology, University of Southern California, Los Angeles, California,
49. Department of Medicine (Biomedical Genetics), Boston University, Boston, Massachusetts,
50. Gerontology and Geriatric Medicine Center on Diabetes, Obesity, and Metabolism, Wake Forest School of Medicine, Winston-Salem, North Carolina,
51. Department of Medicine, University of Washington, Seattle, Washington,
52. Department of Surgery, University of Texas Southwestern Medical Center, Dallas, Texas,
53. Department of Clinical Sciences, University of Texas Southwestern Medical Center, Dallas, Texas,
54. Alzheimer's Disease and Memory Disorders Center, Baylor College of Medicine, Houston, Texas,
55. Program in Translational Neuro-Psychiatric Genomics, Institute for the Neurosciences, Department of Neurology & Psychiatry, Brigham and Women's Hospital and Harvard Medical School, Boston, Massachusetts,
56. Program in Medical and Population Genetics, Broad Institute, Cambridge, Massachusetts,
57. Department of Neurology, University of California Davis, Sacramento, California,
58. Departments of Neurology, Pharmacology & Neuroscience, Texas Tech University Health Science Center, Lubbock, Texas,
59. Wien Center for Alzheimer's Disease and Memory Disorders, Mount Sinai Medical Center, Miami Beach, Florida,
60. Department of Neurology, Mayo Clinic, Jacksonville, Florida,
61. Rush Institute for Healthy Aging, Department of Internal Medicine, Rush University Medical Center, Chicago, Illinois,
62. Office of Strategy and Measurement, University of North Texas Health Science Center, Fort Worth, Texas,
63. Department of Pathology, University of Alabama at Birmingham, Birmingham, Alabama,
64. Sanders-Brown Center on Aging, Department of Biostatistics, University of Kentucky, Lexington, Kentucky,
65. C.S. Kubik Laboratory for Neuropathology, Massachusetts General Hospital, Charlestown, Massachusetts,
66. Internal Medicine, Division of Geriatrics, University of North Texas Health Science Center, Fort Worth, Texas,
67. Department of Pathology and Laboratory Medicine, Emory University, Atlanta, Georgia,
68. Emory Alzheimer's Disease Center, Emory University, Atlanta, Georgia,
69. Department of Pathology and Laboratory Medicine, Indiana University, Indianapolis, Indiana,
70. Division of Genetics, Department of Medicine and Partners Center for Personalized Genetic Medicine, Brigham and Women's Hospital and Harvard Medical School, Boston, Massachusetts,

71. Department of Neurology, Massachusetts General Hospital/Harvard Medical School, Boston, Massachusetts,
72. Center for Applied Genomics, Children's Hospital of Philadelphia, Philadelphia, Pennsylvania,
73. Department of Pathology (Neuropathology), University of Pittsburgh, Pittsburgh, Pennsylvania,
74. Department of Neurology, University of Alabama at Birmingham, Birmingham, Alabama,
75. Department of Pathology and Laboratory Medicine, University of California Irvine, Irvine, California,
76. Department of Epidemiology and Population Health, Stanford University, Stanford, California,
77. Department of Neurology & Neurological Sciences, Stanford University, Stanford, California,
78. Neurogenomics Division, Translational Genomics Research Institute, Phoenix, Arizona,
79. Department of Pathology, Duke University, Durham, North Carolina,
80. Department of Genome Sciences, University of Washington, Seattle, Washington,
81. Department of Medicine (Medical Genetics), University of Washington, Seattle, Washington,
82. Department of Pathology and Laboratory Medicine, University of California Davis, Sacramento, California,
83. Department of Biostatistics, Boston University, Boston, Massachusetts,
84. Department of Ophthalmology, Boston University, Boston, Massachusetts,
85. Department of Human Genetics, University of Pittsburgh, Pittsburgh, Pennsylvania,
86. University of Pittsburgh Alzheimer's Disease Research Center, Pittsburgh, Pennsylvania,
87. Department of Neurology, Albert Einstein College of Medicine, New York, New York,
88. Department of Biology, Brigham Young University, Provo, Utah,
89. Department of Neurology, Oregon Health & Science University, Portland, Oregon,
90. Department of Neurology, Portland Veterans Affairs Medical Center, Portland, Oregon,
91. Department of Pathology, University of Washington, Seattle, Washington,
92. Department of Neurology, Boston University, Boston, Massachusetts,
93. Department of Pathology, Boston University, Boston, Massachusetts,
94. Department of Neuropsychology, University of California San Francisco, San Francisco, California,
95. Department of Epidemiology, University of Washington, Seattle, Washington,
96. Department of Neurobiology and Behavior, University of California Irvine, Irvine, California,
97. Department of Neurology, Emory University, Atlanta, Georgia,
98. Department of Medicine, University of Washington, Seattle, Washington,
99. Department of Epidemiology and Biostatistics, Case Western Reserve University, Cleveland, Ohio,
100. Cleveland Clinic Lou Ruvo Center for Brain Health, Cleveland Clinic, Cleveland, Ohio,
101. Department of Pathology, University of Michigan, Ann Arbor, Michigan,
102. Department of Psychiatry, Johns Hopkins University, Baltimore, Maryland,
103. Department of Health Behavior and Health Systems, University of North Texas Health Science Center, Fort Worth, Texas,
104. Health Management and Policy Department, School of Public Health, University of North Texas Health Science Center, Fort Worth, Texas,
105. National Center for PTSD at Boston VA Healthcare System, Boston, Massachusetts,
106. Department of Psychiatry, New York University, New York, New York,
107. School of Nursing Northwest Research Group on Aging, University of Washington, Seattle, Washington,
108. harmaTherapeutics Clinical Research, Pfizer Worldwide Research and Development, Cambridge, Massachusetts,
109. Department of Neurology, Northwestern University Feinberg School of Medicine, Chicago, Illinois,
110. Department of Pathology, University of Southern California, Los Angeles, California,
111. Department of Pathology, Stanford University School of Medicine, Stanford, California, USA.
112. Department of Developmental and Cell Biology, UC Irvine, Irvine,
113. Department of Neurology, Washington University, St. Louis, Missouri,
114. Department of Pathology and Immunology, Washington University, St. Louis, Missouri,
115. Center for Population Health & Aging, Texas A&M University Health Science Center, Lubbock Texas,

116. Department of Family and Community Medicine, University of Texas Health Science Center - San Antonio, San Antonio, Texas,
117. Department of Laboratory Medicine and Pathology, Mayo Clinic, Rochester, Minnesota,
118. Department of Neurology, University of Colorado School of Medicine, Aurora, Colorado,
119. Sanders-Brown Center on Aging, Department of Anatomy and Neurobiology, University of Kentucky, Lexington, Kentucky,
120. Arizona Alzheimer's Consortium, Phoenix, Arizona,
121. Banner Alzheimer's Institute, Phoenix, Arizona,
122. Tanz Centre for Research in Neurodegenerative Disease, University of Toronto, Toronto, Ontario,
123. Department of Neurology, University of Texas Southwestern, Dallas, Texas,
124. Departments of Psychiatry, Medicine, Family & Community Medicine, South Texas Veterans Health Administration Geriatric Research Education & Clinical Center (GRECC), UT Health Science Center at San Antonio, San Antonio, Texas,
125. Cambridge Institute for Medical Research and Department of Clinical Neurosciences, University of Cambridge, Cambridge, United Kingdom,
126. Program in Cellular Neuroscience, Neurodegeneration & Repair, Yale University, New Haven, Connecticut,
127. Department of Psychiatry & Human Behavior, University of California Irvine, Irvine, California,
128. Department of Pathology, Johns Hopkins University, Baltimore, Maryland,
129. Department of Psychiatry and Behavioral Sciences, University of Washington School of Medicine, Seattle, Washington,
130. Department of Neurology, University of California Los Angeles, Los Angeles, California,
131. Taub Institute on Alzheimer's Disease and the Aging Brain, Department of Pathology, Columbia University, New York, New York,
132. Department of Psychiatry & Behavioral Sciences, Duke University, Durham, North Carolina,
133. Center for Cognitive Neurology and Departments of Neurology, New York University, School of Medicine, New York,
134. Department of Biostatistics, University of Washington, Seattle, Washington,

#### FinnGen

Aarno Palotie[1], Mark Daly[1], Howard Jacob[2], Athena Matakidou[3], Heiko Runz[4], Sally John[4], Robert Plenge[5], Mark McCarthy[6], Julie Hunkapiller[6], Meg Ehm[7], Dawn Waterworth[7], Caroline Fox[8], Anders Malarstig[9], Kathy Klinger[10], Kathy Call10, Tim Behrens[11], Patrick Loerch[12], Tomi Mäkelä[13], Jaakko Kaprio[1], Petri Virolainen[14], Kari Pulkki[14], Terhi Kilpi[15], Markus Perola[15], Jukka Partanen[16], Anne Pitkäranta[17], Riitta Kaarteenaho[18], Seppo Vainio[18], Miia Turpeinen[18], Raisa Serpi[18], Tarja Laitinen[19], Johanna Mäkelä[19], Veli-Matti Kosma[20], Urho Kujala[21], Outi Tuovila[22], Minna Hendolin[22], Raimo Pakkanen[22], Jeff Waring[2], Bridget Riley-Gillis[2], Jimmy Liu[4], Shameek Biswas[5], Julie Hunkapiller[6], Dorothee Diogo[8], Catherine Marshall[9], Xinli Hu[9], Matthias Gossel[10], Robert Graham[11], Tim Behrens[11], Beryl Cummings[12], Samuli Ripatti[1], Johanna Schleutker[14], Mikko Arvas[16], Olli Carpén[17], Reetta Hinttala[18], Johannes Kettunen[18], Arto Mannermaa[20], Jari Laukkanen[21], Hilkka Soininen[23], Valtteri Julkunen[23], Anne Remes[23], Reetta Kälviäinen[23], Jukka Peltola[24], Pentti Tienari[25], Juha Rinne[26], Adam Ziemann[2], Jeffrey Waring[2], Sahar Esmaeeli[2], Nizar Smaoui[2], Anne Lehtonen[2], Susan Eaton[4], Sanni Lahdenperä[4], Janet van Adelsberg[5], Shameek Biswas[5], John Michon[6], Geoff Kerchner[6], Natalie Bowers[6], Edmond Teng[6], John Eicher[8], Vinay Mehta[8], Padhraig Gormley[8], Kari Linden[9], Christopher Whelan[9], Fanli Xu[7], David Pulford[7], Martti Färkkilä[25], Sampsa Pikkarainen[25], Airi Jussila[27], Timo Blomster[28], Mikko Kiviniemi[29], Markku Voutilainen[26], Bob Georgantas[2], Graham Heap[2], Fedik Rahimov[2], Keith Usiskin[5], Tim Lu[6], Danny Oh[6], Kirsi Kalpala[9], Melissa Miller[9], Linda McCarthy[7], Kari Eklund[25], Antti Palomäki[26], Pia Isomäki[27], Laura Pirilä[26], Oili Kaipiainen-Seppänen[29], Johanna Huhtakangas[28], Bob Georgantas[2], Fedik Rahimov[2], Apinya Lertratanakul[2], Marla Hochfeld[5], Kirsi Kalpala[9], Nan Bing[9], Jorge Esparza Gordillo[7], Nina Mars[1], Margit Pelkonen[29], Paula Kauppi[25], Hannu Kankaanranta[24], Terttu Harju[28], David Close[3], Steven Greenberg[5], Hubert Chen[6], Jo Betts[7], Soumitra Ghosh[7], Veikko Salomaa[30], Teemu Niiranen[30], Markus Juonala[26],

Kaj Metsärinne[26], Mika Kähönen[27], Juhani Junttila[28], Markku Laakso[23], Jussi Pihlajamäki[23], Juha Sinisalo[25], Marja-Riitta Taskinen[25], Tiinamaija Tuomi[25], Ben Challis[3], Andrew Peterson[6], Audrey Chu[8], Jaakko Parkkinen[9], Melissa Miller[9], Anthony Muslin[10], Dawn Waterworth[7], Heikki Joensuu[25], Tuomo Meretoja[25], Lauri Aaltonen[25], Johanna Mattson[25], Annika Auranen[24], Peeter Karihtala[28], Saila Kauppila[28], Päivi Auvinen[23], Klaus Elenius[26], Relja Popovic[2], Jennifer Schutzman[6], Andrey Loboda[8], Aparna Chhibber[8], Heli Lehtonen[9], Stefan McDonough[9], Marika Crohns[10], Diptee Kulkarni[7], Kai Kaarniranta[23], Joni A Turunen[25], Terhi Ollila[25], Sanna Seitsonen[25], Hannu Uusitalo[24], Vesa Aaltonen[26], Hannele Uusitalo-Järvinen[24], Marja Luodonpää[28], Nina Hautala[28], Stephanie Loomis[4], Erich Strauss[6], Hao Chen[6], Anna Podgornaia[8], Joshua Hoffman[7], Kaisa Tasanen[28], Laura Huilaja[28], Katariina Hannula-Jouppi[25], Teea Salmi[27], Sirkku Peltonen[25], Leena Koulu[25], Ilkka Harvima[23], Kirsi Kalpala[9], Ying Wu[9], David Choy[6], Fedik Rahimov[2], Dawn Waterworth[7], Pirkko Pussinen[25], Aino Salminen[25], Tuula Salo[25], David Rice[25], Pekka Nieminen[25], Ulla Palotie[25], Maria Siponen[23], Liisa Suominen[23], Päivi Mäntylä[23], Ulvi Gursoy[26], Vuokko Anttonen[28], Kirsi Sipilä[28], Justin Wade Davis[2], Bridget Riley-Gillis[2], Danjuma Quarless[2], Fedik Rahimov[2], Sahar Esmaeeli[2], Slavé Petrovski[3], Eleonor Wigmore[3], Chia-Yen Chen[4], Paola Bronson4, Ellen Tsai[4], Yunfeng Huang[4], Joseph Maranville[5], Elmutaz Shaikho Elhaj Mohammed[5], Samir Wadhawan[31], Erika Kvikstad[31], Minal Caliskan[31], Diana Chang[6], Tushar Bhangale[6], Natalie Bowers[6], Sarah Pendergrass[6], Emily Holzinger[8], Xing Chen[9], Åsa Hedman[9], Karen S King[7], Clarence Wang[10], Ethan Xu[10], Franck Auge[10], Clement Chatelain[10], Deepak Rajpal[10], Dongyu Liu[10], Katherine Call[10], Tai-he Xia[10], Matt Brauer[11], Mitja Kurki[1], Samuli Ripatti[1], Juha Karjalainen[1], Aki Havulinna[1], Anu Jalanko[1], Priit Palta[1], Pietro della Briotta Parolo[1], Wei Zhou[32], Susanna Lemmelä[1], Manuel Rivas[33], Jarmo Harju[1], Arto Lehisto[1], Andrea Ganna[1], Vincent Llorens[1], Hannele Laivuori[1], Sina Rüeger[1], Mari E Niemi[1], Taru Tukiainen[1], Mary Pat Reeve[1], Henrike Heyne[1], Nina Mars[1], Kimmo Palin[34], Javier Garcia-Tabuenca[35], Harri Siirtola[35], Tuomo Kiiskinen[1], Tuomo Kiiskinen[1], Jiwoo Lee[1], Kristin Tsuo[1], Amanda Elliott[1], Kati Kristiansson[15], Kati Hyvärinen[36], Jarmo Ritari[36], Miika Koskinen[17], Katri Pylkäs[18], Marita Kalaoja[18], Minna Karjalainen[18], Tuomo Mantere[18], Eeva Kangasniemi[19], Sami Heikkinen[20], Sami Heikkinen[21], Eija Laakkonen[21], Csilla Sipeky[37], Samuel Heron[37], Antti Karlsson[14], Dhanaprakash Jambulingam[37], Venkat Subramaniam Rathinakannan[37], Anu Jalanko[1], Risto Kajanne[1], Mervi Aavikko[1], Manuel González Jiménez[1], Mitja Kurki[1], Juha Karjalainen[1], Pietro della Briotta Parola[1], Sina Rüeger[1], Arto Lehistö[1], Masahiro Kanai[32], Hannele Laivuori[1], Aki Havulinna[1], Susanna Lemmelä[1], Tuomo Kiiskinen[1], Mari Kaunisto[1], Jarmo Harju[1], Elina Kilpeläinen[1], Timo P. Sipilä[1], Georg Brein[1], Ghazal Awaisa[1], Anastasia Shcherban[1], Kati Donner[1], Timo P. Sipilä[1], Anu Loukola[17], Päivi Laiho[15], Tuuli Sistonen[15], Essi Kaiharju[15], Markku Laukkanen[15], Elina Järvensivu[15], Sini Lähteenmäki[15], Lotta Männikkö[15], Regis Wong[15], Hannele Mattsson[15], Kati Kristiansson[15], Susanna Lemmelä[1], Tero Hiekkalinna[15], Teemu Paajanen[15], Priit Palta[1], Kalle Pärn[1], Harri Siirtola[35], Javier Gracia-Tabuenca[35]

1. Institute for Molecular Medicine Finland, HiLIFE, University of Helsinki, Finland
2. Abbvie, Chicago, IL, United States
3. Astra Zeneca, Cambridge, United Kingdom
4. Biogen, Cambridge, MA, United States
5. Celgene, Summit, NJ, United States
6. Genentech, San Francisco, CA, United States
7. GlaxoSmithKline, Brentford, United Kingdom
8. Merck, Kenilworth, NJ, United States
9. Pfizer, New York, NY, United States
10. Sanofi, Paris, France
11. Maze Therapeutics, San Francisco, CA, United States
12. Janssen Biotech, Beerse, Belgium
13. HiLIFE, University of Helsinki, Finland, Finland

14. Auria Biobank / University of Turku / Hospital District of Southwest Finland, Turku, Finland
15. THL Biobank / The National Institute of Health and Welfare Helsinki, Finland
16. Finnish Red Cross Blood Service / Finnish Hematology Registry and Clinical Biobank, Helsinki
17. Helsinki Biobank / Helsinki University and Hospital District of Helsinki and Uusimaa, Helsinki
18. Northern Finland Biobank Borealis / University of Oulu / Northern Ostrobothnia Hospital District, Oulu, Finland
19. Finnish Clinical Biobank Tampere **/** University of Tampere / Pirkanmaa Hospital District, Tampere, Finland
20. Biobank of Eastern Finland / University of Eastern Finland / Northern Savo Hospital District, Kuopio, Finland
21. Central Finland Biobank / University of Jyväskylä / Central Finland Health Care District, Jyväskylä, Finland
22. Business Finland, Helsinki, Finland
23. Northern Savo Hospital District, Kuopio, Finland
24. Pirkanmaa Hospital District, Tampere, Finland
25. Hospital District of Helsinki and Uusimaa, Helsinki, Finland
26. Hospital District of Southwest Finland, Turku, Finland
27. Pirkanmaa Hospital District, Tampere, Finland
28. Northern Ostrobothnia Hospital District, Oulu, Finland
29. Northern Savo Hospital District, Kuopio, Finland
30. The National Institute of Health and Welfare Helsinki, Finland
31. Bristol-Meyers-Squibb
32. Broad Institute, Cambridge, MA, United States
33. University of Stanford, Stanford, CA, United States
34. University of Helsinki, Helsinki, Finland
35. University of Tampere, Tampere, Finland
36. Finnish Red Cross Blood Service, Helsinki, Finland
37. University of Turku, Turku, Finland

#### 15. Supplementary Figures

**Supplementary Figure 1.** Stage I QQ Plot. QQ Plot of Stage I meta-analysis results (excludes the APOE locus). Genomic inflation factors ($\lambda$) were slightly inflated ($\lambda$ =1.08 overall and 1.17 when restricted to variants with minor allele frequency (MAF) above 1%. However, linkage disequilibrium score (LDSC) regression estimate indicated that the majority of this inflation was due to a polygenic signal, with the intercept being close to 1 (intercept=1.05, s.e=0.01 versus $\lambda$=1.2 on the variants considered in the LDSC analysis).

**Supplementary Figure 2.** LocusZoom and forest plots for (a) SORT1, (b) CR1 and (c) ADAM17 loci. The LocusZoom plot is based on the results from Stage I, and the variant in purple is the best associated variant in the Stage I + II meta-analysis. OR: odds ratio, CI: confidence interval, EA: effect allele, EAF: effect allele frequency range across all studies, HetP: heterogeneity P value, HetISq: heterogeneity statistic.

**Supplementary Figure 3.** LocusZoom and forest plots for (a) PRKD3, (b) NCK2 and (c) BIN1 loci. The LocusZoom plot is based on the results from Stage I, and the variant in purple is the best associated variant in the Stage I + II meta-analysis. OR: odds ratio, CI: confidence interval, EA: effect allele, EAF: effect allele frequency range across all studies, HetP: heterogeneity P value, HetISq: heterogeneity statistic.

**Supplementary Figure 4.** LocusZoom and forest plots for (a) WDR12, (b) INPP5D and (c) MME (1) loci. The LocusZoom plot is based on the results from Stage I, and the variant in purple is the best associated variant in the Stage I + II meta-analysis. OR: odds ratio, CI: confidence interval, EA: effect allele, EAF: effect allele frequency range across all studies, HetP: heterogeneity P value, HetISq: heterogeneity statistic.

**Supplementary Figure 5.** LocusZoom and forest plots for (a) MME (2), (b) IDUA and (c) CLNK loci. The LocusZoom plot is based on the results from Stage I, and the variant in purple is the best associated variant in the Stage I + II meta-analysis. OR: odds ratio, CI: confidence interval, EA: effect allele, EAF: effect allele frequency range across all studies, HetP: heterogeneity P value, HetISq: heterogeneity statistic.

**Supplementary Figure 6.** LocusZoom and forest plots for (a) RHOH, (b) ANKH and (c) COX7C loci. The LocusZoom plot is based on the results from Stage I, and the variant in purple is the best associated variant in the Stage I + II meta-analysis. OR: odds ratio, CI: confidence interval, EA: effect allele, EAF: effect allele frequency range across all studies, HetP: heterogeneity P value, HetISq: heterogeneity statistic.

**Supplementary Figure 7.** LocusZoom and forest plots for (a) TNIP1, (b) RASGF1C and (c) HLA-DQA1 loci. The LocusZoom plot is based on the results from Stage I, and the variant in purple is the best associated variant in the Stage I + II meta-analysis. OR: odds ratio, CI: confidence interval, EA: effect allele, EAF: effect allele frequency range across all studies, HetP: heterogeneity P value, HetISq: heterogeneity statistic.

**Supplementary Figure 8.** LocusZoom and forest plots for (a) UNC5CL, (b) TREM2 (R62H) and (c) TREM2 (R47H) loci. The LocusZoom plot is based on the results from Stage I, and the variant in purple is the best associated variant in the Stage I + II meta-analysis. OR: odds ratio, CI: confidence interval, EA: effect allele, EAF: effect allele frequency range across all studies, HetP: heterogeneity P value, HetISq: heterogeneity statistic.

**Supplementary Figure 9.** LocusZoom and forest plots for (a) TREML2, (b) CD2AP and (c) HS3ST5 loci. The LocusZoom plot is based on the results from Stage I, and the variant in purple is the best associated variant in the Stage I + II meta-analysis. OR: odds ratio, CI: confidence interval, EA: effect allele, EAF: effect allele frequency range across all studies, HetP: heterogeneity P value, HetISq: heterogeneity statistic.

**Supplementary Figure 10.** LocusZoom and forest plots for (a) UMAD1, (b) ICA1 and (c) TMEM106B loci. The LocusZoom plot is based on the results from Stage I, and the variant in purple is the best associated variant in the Stage I + II meta-analysis. OR: odds ratio, CI: confidence interval, EA: effect allele, EAF: effect allele frequency range across all studies, HetP: heterogeneity P value, HetISq: heterogeneity statistic.

**Supplementary Figure 11.** LocusZoom and forest plots for (a) JAZF1, (b) NME8 and (c) SEC61G loci. The LocusZoom plot is based on the results from Stage I, and the variant in purple is the best associated variant in the Stage I + II meta-analysis. OR: odds ratio, CI: confidence interval, EA: effect allele, EAF: effect allele frequency range across all studies, HetP: heterogeneity P value, HetISq: heterogeneity statistic.

**Supplementary Figure 12.** LocusZoom and forest plots for (a) ZCWPW1/NYAP1, (b) EPHA1 and (c) CTSB loci. The LocusZoom plot is based on the results from Stage I, and the variant in purple is the best associated variant in the Stage I + II meta-analysis. OR: odds ratio, CI: confidence interval, EA: effect allele, EAF: effect allele frequency range across all studies, HetP: heterogeneity P value, HetISq: heterogeneity statistic.

**Supplementary Figure 13.** LocusZoom and forest plots for (a) PTK2B, (b) CLU and (c) SHARPIN loci. The LocusZoom plot is based on the results from Stage I, and the variant in purple is the best associated variant in the Stage I + II meta-analysis. OR: odds ratio, CI: confidence interval, EA: effect allele, EAF: effect allele frequency range across all studies, HetP: heterogeneity P value, HetISq: heterogeneity statistic.

**Supplementary Figure 14.** LocusZoom and forest plots for (a) ABCA1, (b) USP6NL and (c) ANK3 loci. The LocusZoom plot is based on the results from Stage I, and the variant in purple is the best associated variant in the Stage I + II meta-analysis. OR: odds ratio, CI: confidence interval, EA: effect allele, EAF: effect allele frequency range across all studies, HetP: heterogeneity P value, HetISq: heterogeneity statistic.

**Supplementary Figure 15.** LocusZoom and forest plots for (a) TSPAN14, (b) BLNK and (c) PLEKHA1 loci. The LocusZoom plot is based on the results from Stage I, and the variant in purple is the best associated variant in the Stage I + II meta-analysis. OR: odds ratio, CI: confidence interval, EA: effect allele, EAF: effect allele frequency range across all studies, HetP: heterogeneity P value, HetISq: heterogeneity statistic.

**Supplementary Figure 16.** LocusZoom and forest plots for (a) CELF1/SPI1, (b) MS4A and (c) PICALM loci. The LocusZoom plot is based on the results from Stage I, and the variant in purple is the best associated variant in the Stage I + II meta-analysis. OR: odds ratio, CI: confidence interval, EA: effect allele, EAF: effect allele frequency range across all studies, HetP: heterogeneity P value, HetISq: heterogeneity statistic.

**Supplementary Figure 17.** LocusZoom and forest plots for (a) SORL1 (1), (b) SORL1 (2) and (c) TPCN1 loci. The LocusZoom plot is based on the results from Stage I, and the variant in purple is the best associated variant in the Stage I + II meta-analysis. OR: odds ratio, CI: confidence interval, EA: effect allele, EAF: effect allele frequency range across all studies, HetP: heterogeneity P value, HetISq: heterogeneity statistic.

**Supplementary Figure 18.** LocusZoom and forest plots for (a) FERMT2, (b) SLC24A4/RIN3 (1) and (c) SLC24A4/RIN3 (2) loci. The LocusZoom plot is based on the results from Stage I, and the variant in purple is the best associated variant in the Stage I + II meta-analysis. OR: odds ratio, CI: confidence interval, EA: effect allele, EAF: effect allele frequency range across all studies, HetP: heterogeneity P value, HetISq: heterogeneity statistic.

**Supplementary Figure 19.** LocusZoom and forest plots for (a) IGH gene cluster (1), (b) IGH gene cluster (2) and (c) SPPL2A loci. The LocusZoom plot is based on the results from Stage I, and the variant in purple is the best associated variant in the Stage I + II meta-analysis. OR: odds ratio, CI: confidence interval, EA: effect allele, EAF: effect allele frequency range across all studies, HetP: heterogeneity P value, HetISq: heterogeneity statistic.

**Supplementary Figure 20.** LocusZoom and forest plots for (a) ADAM10, (b) APH1B and (c) SNX1 loci. The LocusZoom plot is based on the results from Stage I, and the variant in purple is the best associated variant in the Stage I + II meta-analysis. OR: odds ratio, CI: confidence interval, EA: effect allele, EAF: effect allele frequency range across all studies, HetP: heterogeneity P value, HetISq: heterogeneity statistic.

**Supplementary Figure 21.** LocusZoom and forest plots for (a) CTSH, (b) DOC2A and (c) KAT8 loci. The LocusZoom plot is based on the results from Stage I, and the variant in purple is the best associated variant in the Stage I + II meta-analysis. OR: odds ratio, CI: confidence interval, EA: effect allele, EAF: effect allele frequency range across all studies, HetP: heterogeneity P value, HetISq: heterogeneity statistic.

**Supplementary Figure 22.** LocusZoom and forest plots for (a) IL34, (b) MAF and (c) PLCg2 (1) loci. The LocusZoom plot is based on the results from Stage I, and the variant in purple is the best associated variant in the Stage I + II meta-analysis. OR: odds ratio, CI: confidence interval, EA: effect allele, EAF: effect allele frequency range across all studies, HetP: heterogeneity P value, HetISq: heterogeneity statistic.

**Supplementary Figure 23.** LocusZoom and forest plots for (a) PLCg2 (2), (b) FOXF1 and (c) PRDM7 loci. The LocusZoom plot is based on the results from Stage I, and the variant in purple is the best associated variant in the Stage I + II meta-analysis. OR: odds ratio, CI: confidence interval, EA: effect allele, EAF: effect allele frequency range across all studies, HetP: heterogeneity P value, HetISq: heterogeneity statistic.

**Supplementary Figure 24.** LocusZoom and forest plots for (a) WDR81, (b) SCIMP/RABEP1 and (c) MYO15A loci. The LocusZoom plot is based on the results from Stage I, and the variant in purple is the best associated variant in the Stage I + II meta-analysis. OR: odds ratio, CI: confidence interval, EA: effect allele, EAF: effect allele frequency range across all studies, HetP: heterogeneity P value, HetISq: heterogeneity statistic.

**Supplementary Figure 25.** LocusZoom and forest plots for (a) GRN, (b) MAPT and (c) ABI3 loci. The LocusZoom plot is based on the results from Stage I, and the variant in purple is the best associated variant in the Stage I + II meta-analysis. OR: odds ratio, CI: confidence interval, EA: effect allele, EAF: effect allele frequency range across all studies, HetP: heterogeneity P value, HetISq: heterogeneity statistic.

**Supplementary Figure 26.** LocusZoom and forest plots for (a) TSPOAP1, (b) ACE and (c) ABCA7 loci. The LocusZoom plot is based on the results from Stage I, and the variant in purple is the best associated variant in the Stage I + II meta-analysis. OR: odds ratio, CI: confidence interval, EA: effect allele, EAF: effect allele frequency range across all studies, HetP: heterogeneity P value, HetISq: heterogeneity statistic.

**Supplementary Figure 27.** LocusZoom and forest plots for (a) KLF16, (b) SIGLEC11 and (c) LILRB2 loci. The LocusZoom plot is based on the results from Stage I, and the variant in purple is the best associated variant in the Stage I + II meta-analysis. OR: odds ratio, CI: confidence interval, EA: effect allele, EAF: effect allele frequency range across all studies, HetP: heterogeneity P value, HetISq: heterogeneity statistic.

**Supplementary Figure 28.** LocusZoom and forest plots for (a) RBCK1, (b) CASS4 and (c) SLC2A4RG loci. The LocusZoom plot is based on the results from Stage I, and the variant in purple is the best associated variant in the Stage I + II meta-analysis. OR: odds ratio, CI: confidence interval, EA: effect allele, EAF: effect allele frequency range across all studies, HetP: heterogeneity P value, HetISq: heterogeneity statistic.

**Supplementary Figure 29.** LocusZoom and forest plots for (a) APP and (b) ADAMTS1 loci. The LocusZoom plot is based on the results from Stage I, and the variant in purple is the best associated variant in the Stage I + II meta-analysis. OR: odds ratio, CI: confidence interval, EA: effect allele, EAF: effect allele frequency range across all studies, HetP: heterogeneity P value, HetISq: heterogeneity statistic.

**Supplementary Figure 30.** Comparison of ORs observed in the diagnosed cases only analysis and estimated in Stage I including ADD-proxy cases for the genome-wide significant loci. OR: odds-ratio

**Supplementary Figure 31.** Forest plots of the 8 HLA alleles associated with ADD (FDR P below 0.05). AF: allele frequency, HetP: heterogeneity P value, HetISq: heterogeneity statistic.

**Supplementary Figure 32.** Forest plots of the 3 three-locus haplotypes associated with ADD (FDR P below 0.05). AF: allele frequency, HetP: heterogeneity P value, HetISq: heterogeneity statistic.

**Supplementary Figure 33.** Z-scores for neurodegenerative and AD related diseases for the risk allele of the ADD associated variants. Colors represent the direction of association: the ADD-risk allele increases risk of the disease (red), decreases risk of the disease (blue) or shows no effect (white). Euclidian distances that were clustered according to unweighted pair group method with arithmetic mean (UPGMA) were used (columns and rows). P values (uncorrected) are denoted with $ ($P<5x10^{-8}$), X ($P<1x10^{-5}$), # ($P<1x10^{-3}$), * (P<0.05). The following variants were not shown as they were not found (or a proxy) in more than half of the traits: rs141749679:SORT1, rs143332484:TREM2, rs75932628:TREM2, rs1160871:JAZF1, rs143080277:NCK2, rs35048651:WDR81, rs139643391:WDR12, rs616338:ABI3, rs149080927:KLF16. These are mainly the rarer variants or indels. Creutzfeldt-Jakob disease (CJD), Dementia with Lewy-Bodies (DLB), amyotrophic lateral sclerosis (ALS), Frontotemporal dementia (FTD), Parkinson's disease (PD), ischemic brain infarcts (MRI defined) and ischemic stroke (clinical), white matter lesions (WML).

**Supplementary Figure 34.** eQTL effects of lead variants within novel ADD risk loci in (a) AD-relevant brain regions, LCL, microglia, blood, and (b) in naïve state and stimulated macrophages and monocytes. Overlap of novel lead variants with significant eQTL variants affecting the expression of genes within 1 Mb in n=26 different eQTL catalogues. Absolute slope (beta) values of eQTL associations are indicated in increasing scale of point size. Gene expression increasing effect of the risk allele of the lead variant with eQTL association is colored red, and decreasing effect is colored blue. For stimulated macrophage and monocyte eQTL catalogues, "h" stands for hours of exposure to the stimulant, and the stimulants are Influenza, Listeria, Salmonella, IFNg (Interferon gamma), LPS (Lipopolysaccharides), Pam3CSK4 (Pam3CysSerLys4) and R848 (Resiquimod). Index number of novel loci are shown in parentheses.

**Supplementary Figure 35.** sQTL effects of lead variants within novel ADD risk loci in AD-relevant brain regions, LCL, microglia, and blood. Overlap of novel lead variants with significant sQTL variants affecting the alternative splicing of genes within 1 Mb in n=18 different sQTL catalogues. Absolute slope (beta) values of sQTL associations are indicated in increasing scale of point size. Increasing splice junction preference effect of the risk allele of the lead variant with sQTL association is colored red, and decreasing effect is colored blue. Index number of novel loci are shown in parentheses.

**Supplementary Figure 36.** (a) mQTL and haQTL effects of lead variants within novel ADD risk loci. Overlap of novel lead variants with significant mQTL and haQTL variants affecting respectively methylation & histone acetylation of the features within 1 Mb in ROSMAP DLPFC mQTL & haQTL catalogues (xQTL Serve). Shapes indicate the positional annotation of the feature for the related gene. Absolute Spearman's rho values of QTL associations are labelled below the shapes. Methylation or histone acetylation increasing effect of the risk allele of the lead variant is colored red, and decreasing effect is colored blue. Index number of novel loci are shown in parentheses. (b) ADD-associated predicted methylation results using MetaMeth. Results for ADD-associated significant (after Bonferroni correction) CpG features are shown, where Z-score of association is indicated in a heatmap scale from -7.5 to +7.5. Each CpG is paired with its annotated gene(s) and respective positional annotation of CpGs are shown in different shapes on the figure. MetaMeth hits are grouped by their percentile and direction of blood-brain methylation correlation estimates across 3 brain regions that were obtained from BECon website. Index number of novel loci are shown in parentheses.

**Supplementary Figure 37.** Colocalization between eQTL signals for genes and ADD association signals. Colocalization probability results for ADD signal with the eQTL signals of genes within 1 Mb of lead variants of novel ADD risk loci in n=12 different eQTL catalogues. Only the genes with at least one significant eQTL (in any included tissues or cell groups) overlapping with at least one suggestively significant (p =< 1E-5) ADD risk variant were tested. Colocalization probability PP4 estimate is indicated in increasing scale of point size and opacity for all tested colocalizations, and scores with at least eQTL coloc PP4 ≥ 70% is labelled on the figure as well. Only colocalized hits at an eQTL coloc PP4 ≥ 70% level in at least one catalogue are shown in this figure. Index number of novel loci are shown in parentheses.

**Supplementary Figure 38.** Colocalization between sQTL signals for splice junctions and ADD association signals. Colocalization probability results for ADD signal with the sQTL signals of splice junctions within 1 Mb of lead variants of novel ADD risk loci in n=12 different sQTL catalogues. Only the splice junctions with at least one significant sQTL (in any included tissues or cell groups) overlapping with at least one suggestively significant (p =< 1E-5) AD risk variant were tested. Colocalization probability PP4 estimate is indicated in increasing scale of point size and opacity for all tested colocalizations, and scores with at least sQTL coloc PP4 ≥ 70% is labelled on the figure as well. Only colocalized hits at a sQTL coloc PP4 ≥ 70% level in at least one catalogue are shown in this figure. Index number of novel loci are shown in parentheses.

**Supplementary Figure 39.** TWAS of ADD using Expression Reference Panels. Expression TWAS (eTWAS) results for genes within 1 Mb of lead variants in n=13 different expression reference panels used. For each expression reference panel, expression models that are not available in the respective panel are shown as dark gray. TWAS Z-score of association is indicated in a heatmap scale from -10 to +10. Significant eTWAS associations that are passing Bonferroni-corrected significance level threshold per reference panel are labelled with asterisk ("*"), fine-mapped eTWAS associations are labelled with a dagger ("†") along with PIP value, eQTL colocalizations with coloc PP4 ≥ 70% are labelled with "C". Associations are only illustrated if significant in at least one panel. Index number of novel loci are shown in parentheses.

**Supplementary Figure 40.** TWAS of ADD using Splicing Reference Panels. Splicing TWAS (sTWAS) results for splice junctions within 1 Mb of lead variants in n=13 different splicing reference panels used. For each splicing reference panel, splice junctions that are not available in the respective panel are shown as dark gray. TWAS Z-score of association is indicated in a heatmap scale from -10 to +10. Significant sTWAS associations that are passing Bonferroni-corrected significance level threshold per prediction panel is labelled on the figure with asterisk ("*") and sQTL colocalizations with coloc PP4 ≥ 70% are labelled with "C". Associations are only illustrated if significant in at least one panel. Index number of novel loci are shown in parentheses.

**Supplementary Figure 41.** Fine-mapping of expression TWAS results. Representative examples of fine-mapping of eTWAS results are shown below in the regional plots (generated by FOCUS) when multiple significant eTWAS hits are observed in the same expression reference panel. Posterior inclusion probability (PIP) values are shown in increasing point size, -log10 of marginal TWAS p-value is shown on y-axis, and pairwise predicted expression correlations between genes shown below the plots. (a) *GRN* locus in MayoRNASeq TCX, (b) *KLF16* locus in ROSMAP DLPFC, (c) *TSPAN14* locus in MSBB BA36, and (d) *DOC2A* locus in GTEx hippocampus.

**Supplementary Figure 42.** Mean fluorescence intensity variations (log2 fold-change) of the mCherry signal obtained after the silencing of genes associated with the ADD risk in HEK293 cells stably over-expressing a mCherry-APP$^{695WT}$-YFP in the 42 new loci. To evaluate the impact of each siRNA (SMARTPool), an average of 1,000 cells was analysed in triplicate. The mean fluorescence intensity was normalized to the fold change based on the non-targeting siRNA. Bars indicate the means ± S.D.

**Supplementary Figure 43.** STRING protein interaction analysis. The main networks are shown in a) previous genes, b) prioritized new genes and c) combined datasets. A significantly larger than expected network of interacting genes was observed in the set of previously identified genes (P value<$2x10^{-5}$, in the prioritized gene set in the new loci (P value=$3.9x10^{-3}$) and also in the combination of these two gene sets (P value<$2x10^{-5}$).

**Supplementary Figure 44.** Association of PRS with the risk of progression to dementia starting from either (a) normal cognition or (b) mild cognitive impairment (MCI) in non e4-bearers. PRS was based on the genetic data of 83 variants (see Methods and Supplementary Table 31).

**Supplementary Figure 45.** Association of PRS with the risk of progression to dementia starting from either (a) normal cognition or (b) mild cognitive impairment (MCI) in e4-bearers. PRS was based on the genetic data of 83 variants (see Methods and Supplementary Table 31).

**Supplementary Figure 46.** EADB sample quality control.

**Supplementary Figure 47.** EADB variant quality control.

**Supplementary Figure 1:** Stage I QQ Plot. QQ Plot of Stage I meta-analysis results (excludes the APOE locus).

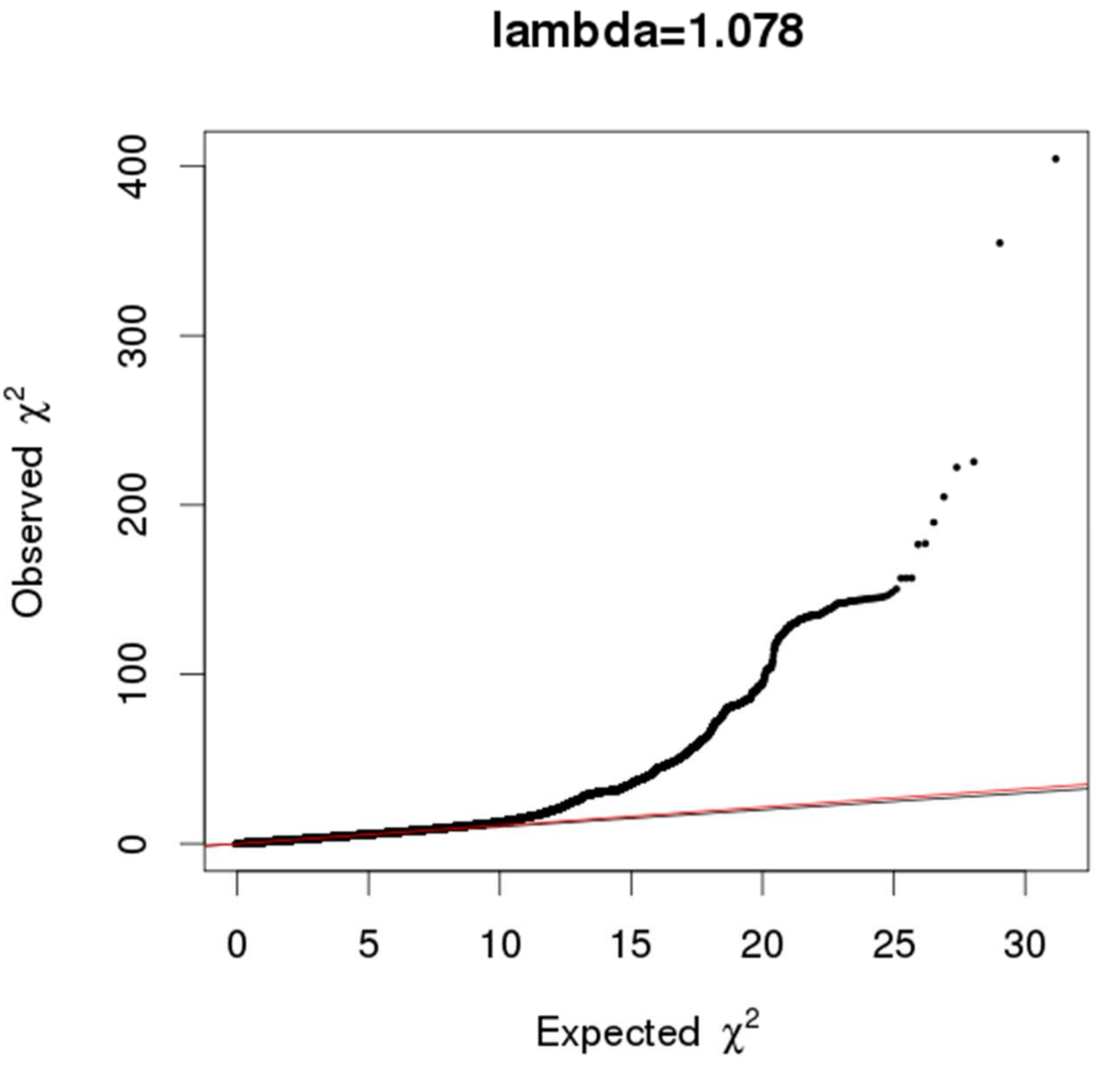

**Supplementary Figure 2**: LocusZoom and forest plots for (a) SORT1, (b) CR1 and (c) ADAM17 loci.

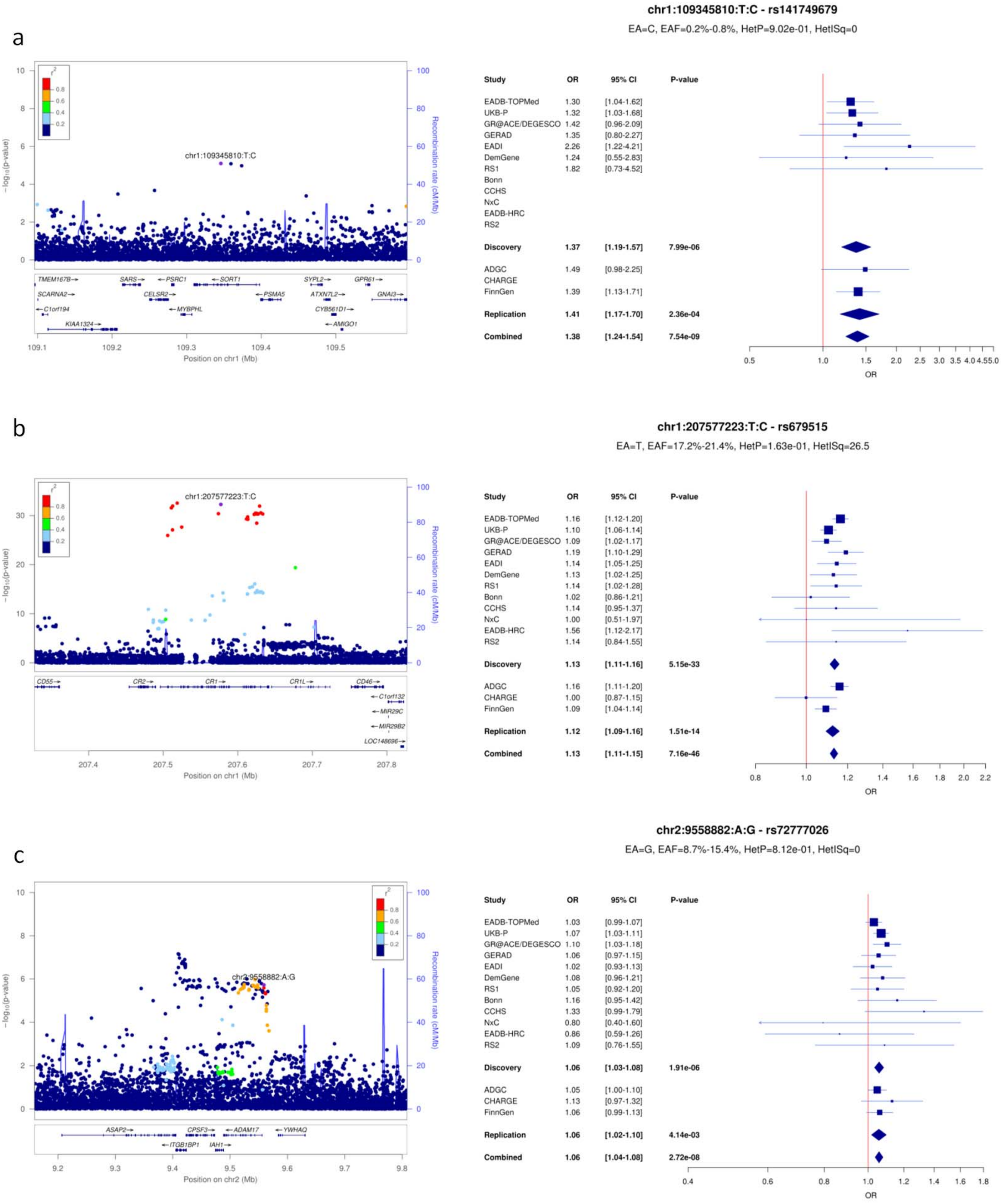

**Supplementary Figure 3**: LocusZoom and forest plots for (a) PRKD3, (b) NCK2 and (c) BIN1 loci.

a

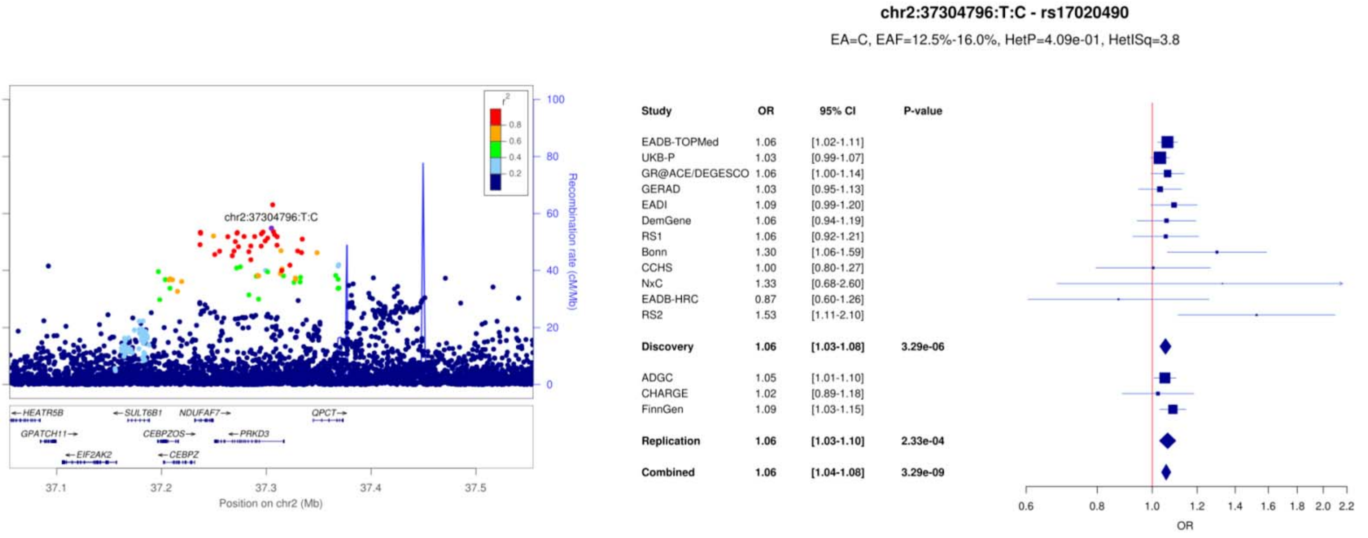


b

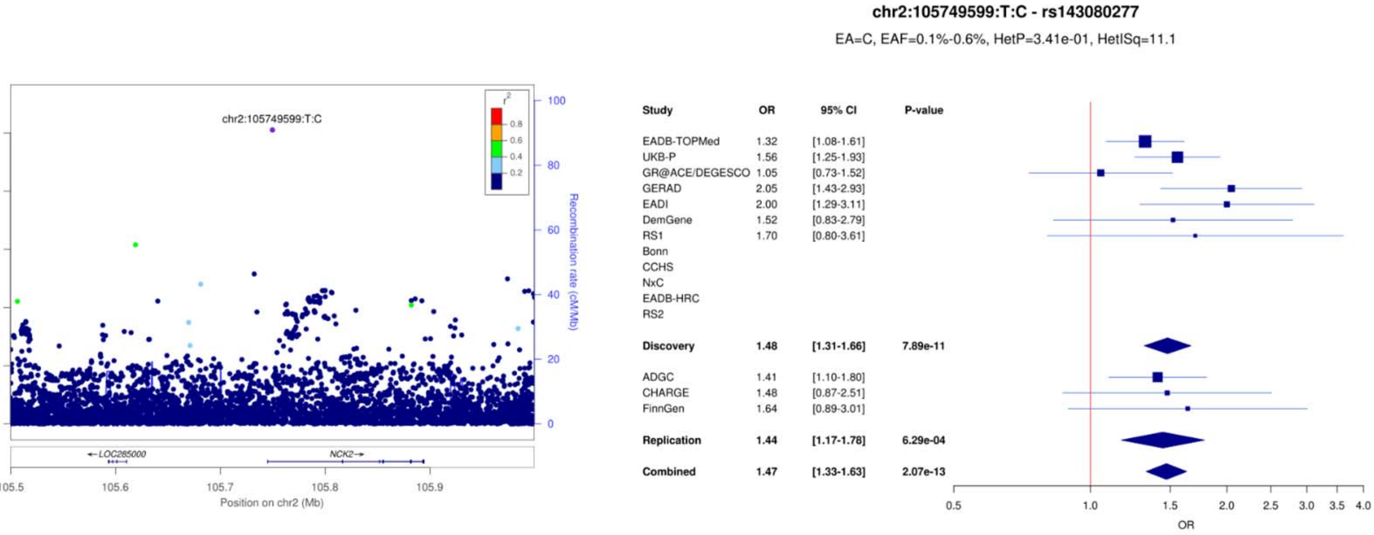


c

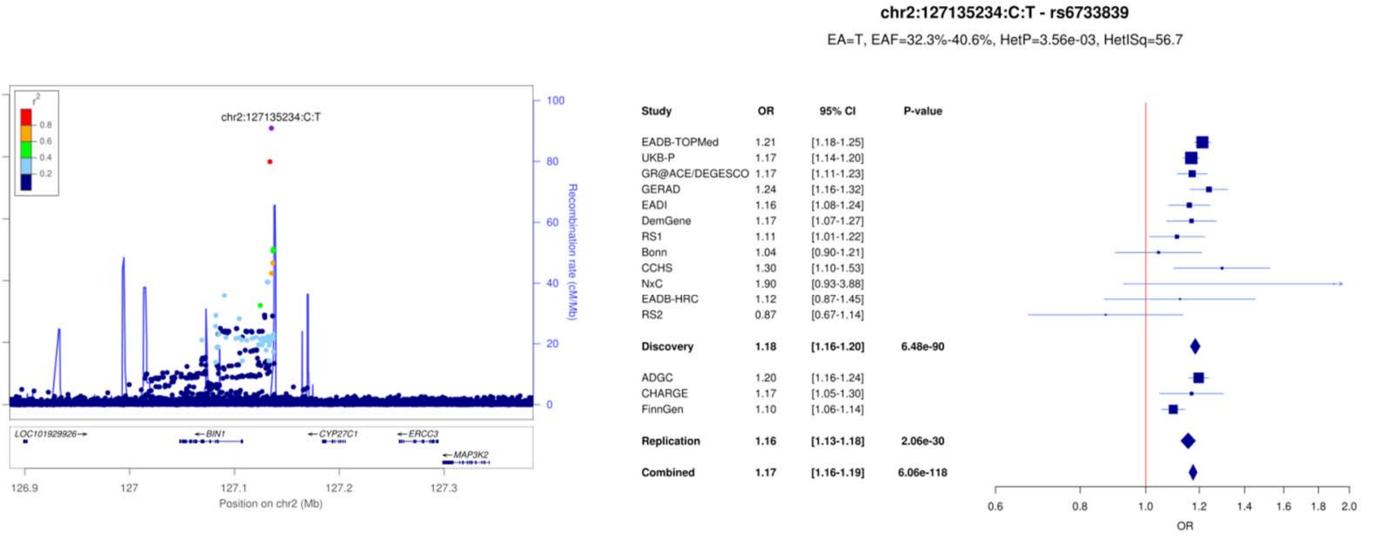

**Supplementary Figure 4**: LocusZoom and forest plots for (a) WDR12, (b) INPP5D and (c) MME (1) loci.

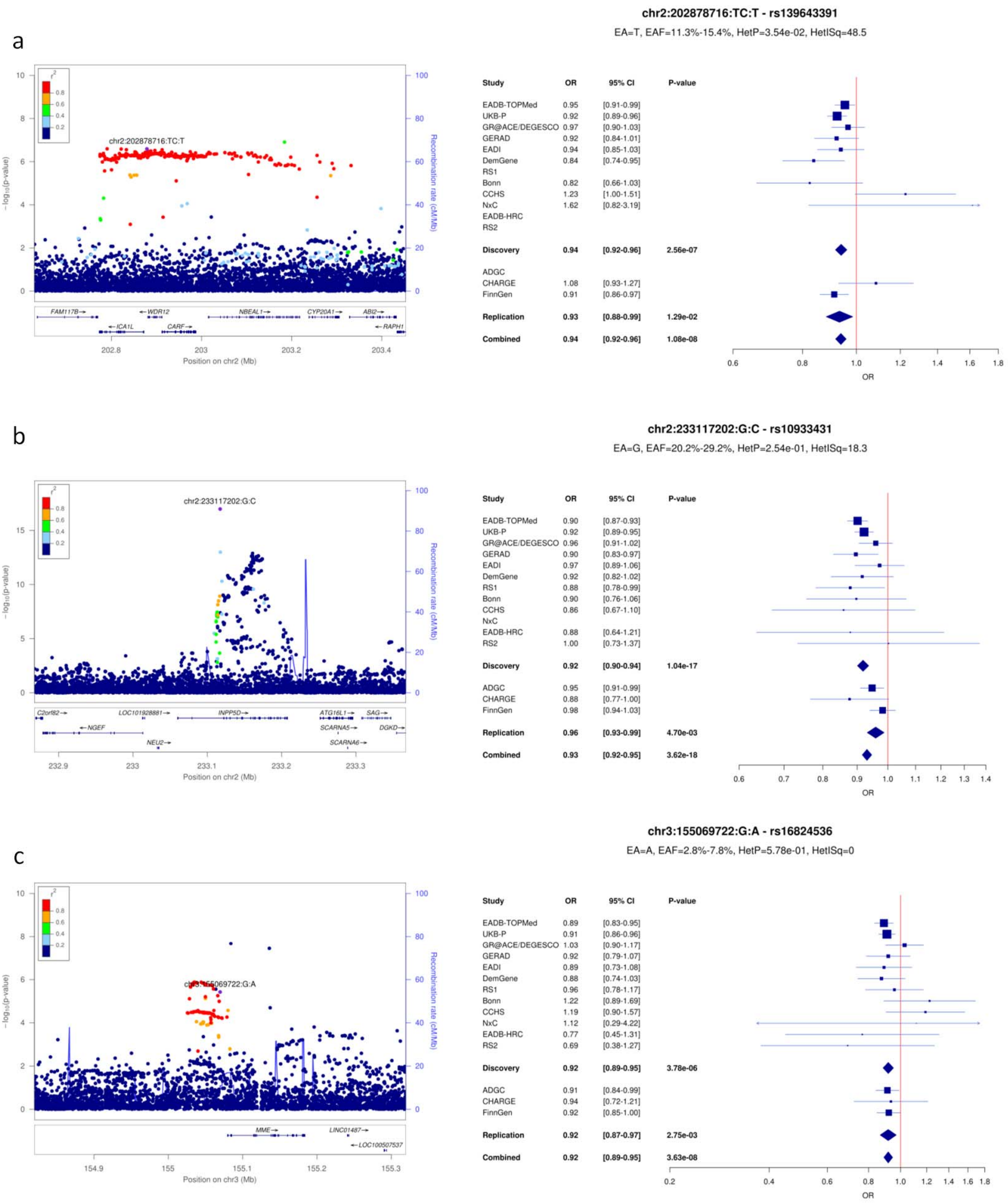

**Supplementary Figure 5**: LocusZoom and forest plots for (a) MME (2), (b) IDUA and (c) CLNK loci.

a

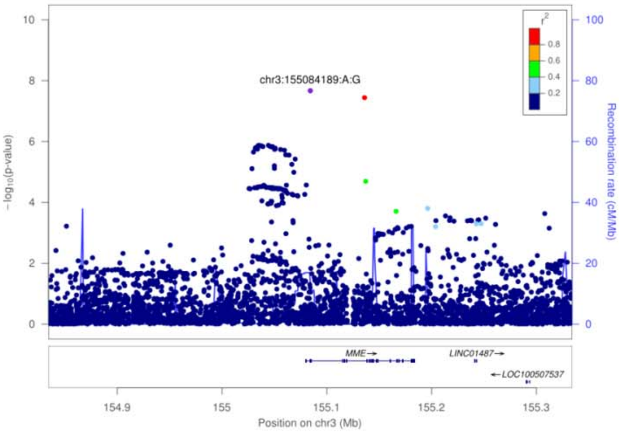


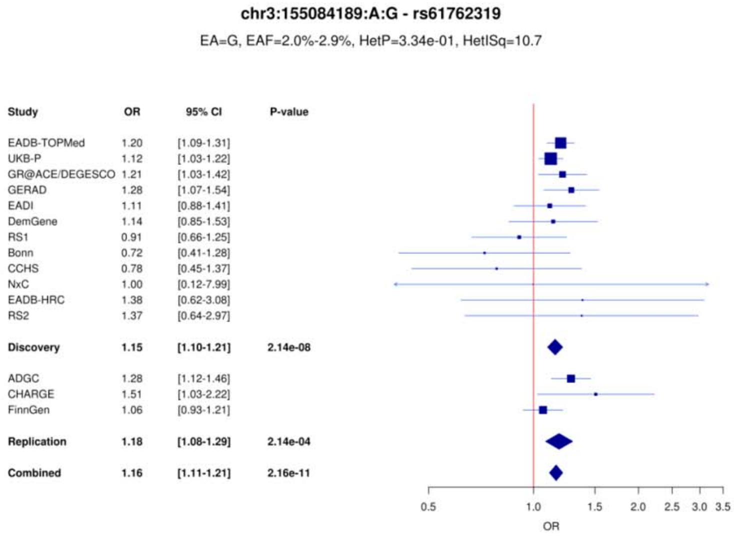


b

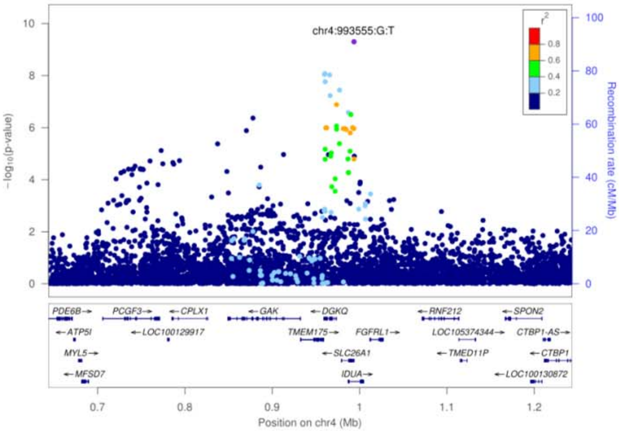


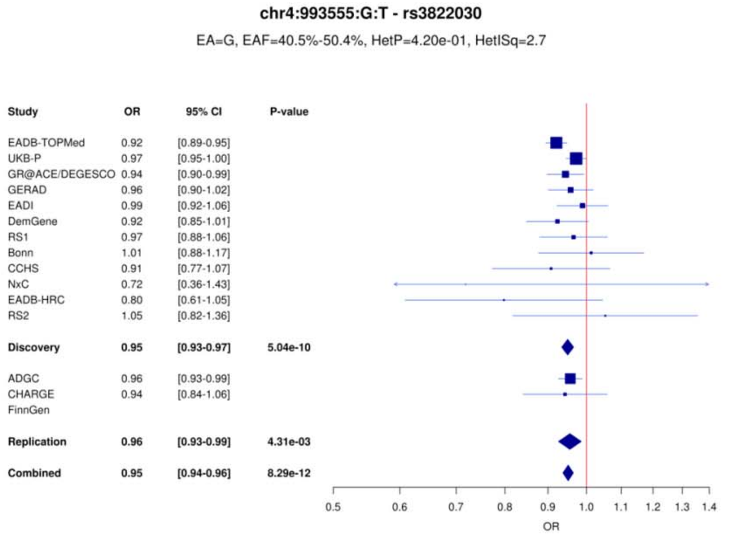


c

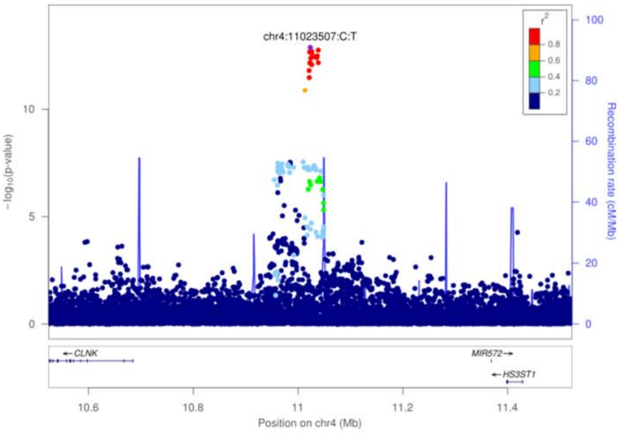


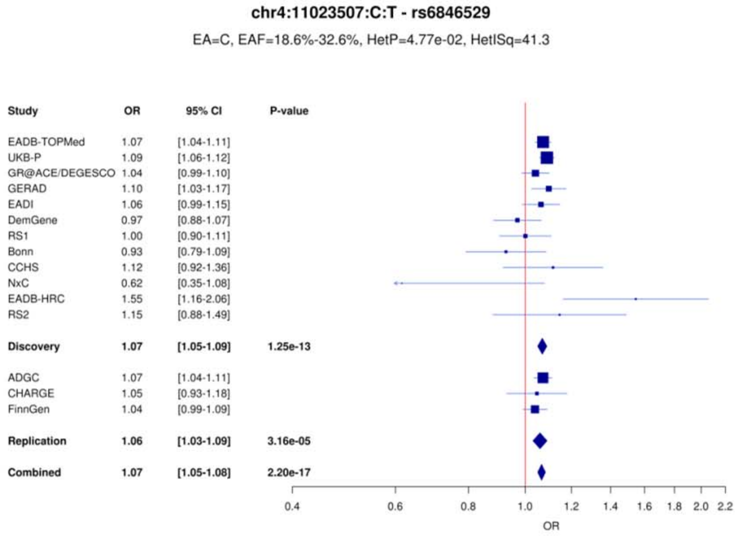

**Supplementary Figure 6**: LocusZoom and forest plots for (a) RHOH, (b) ANKH and (c) COX7C loci.

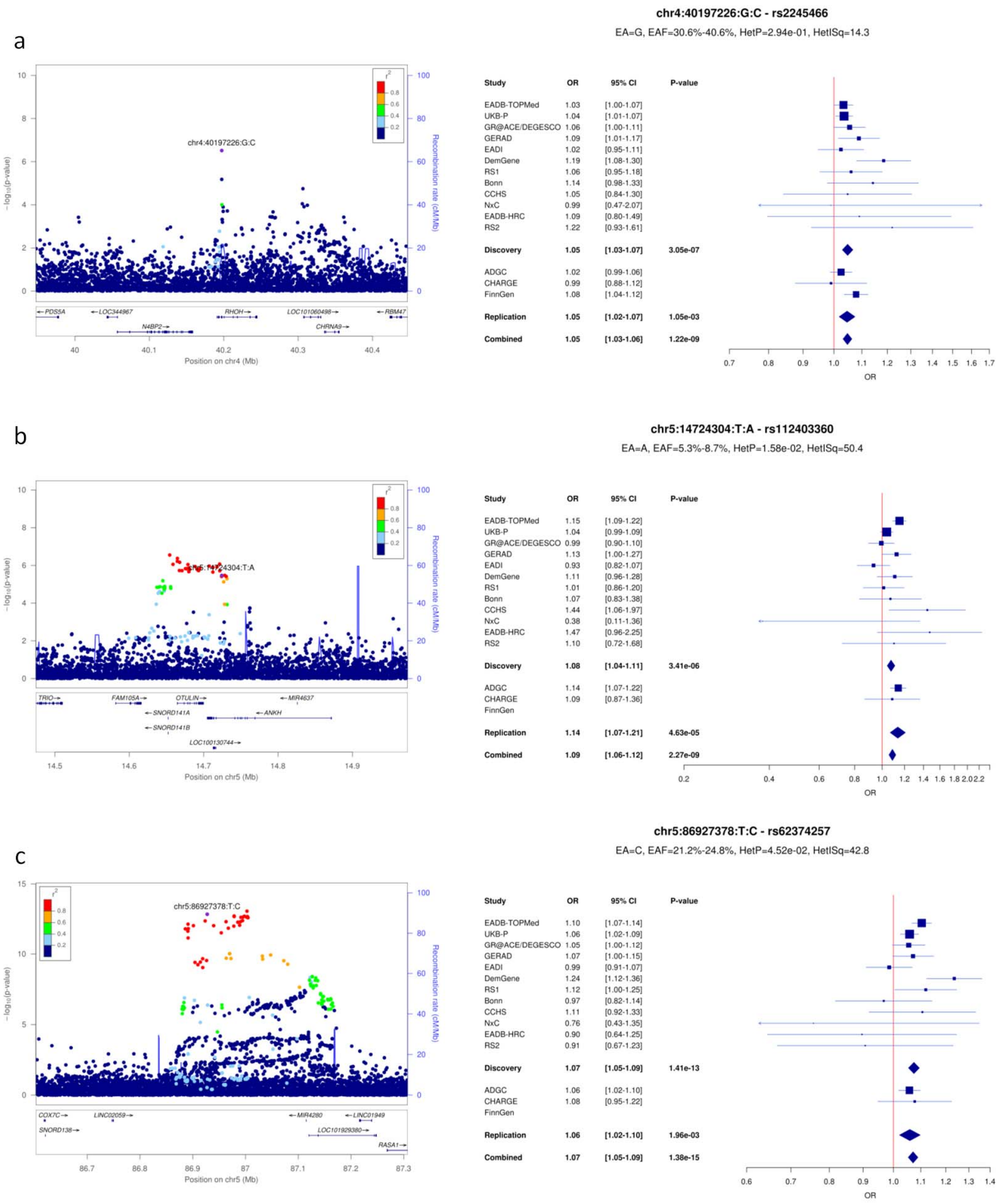

**Supplementary Figure 7**: LocusZoom and forest plots for (a) TNIP1, (b) RASGF1C and (c) HLA-DQA1 loci.

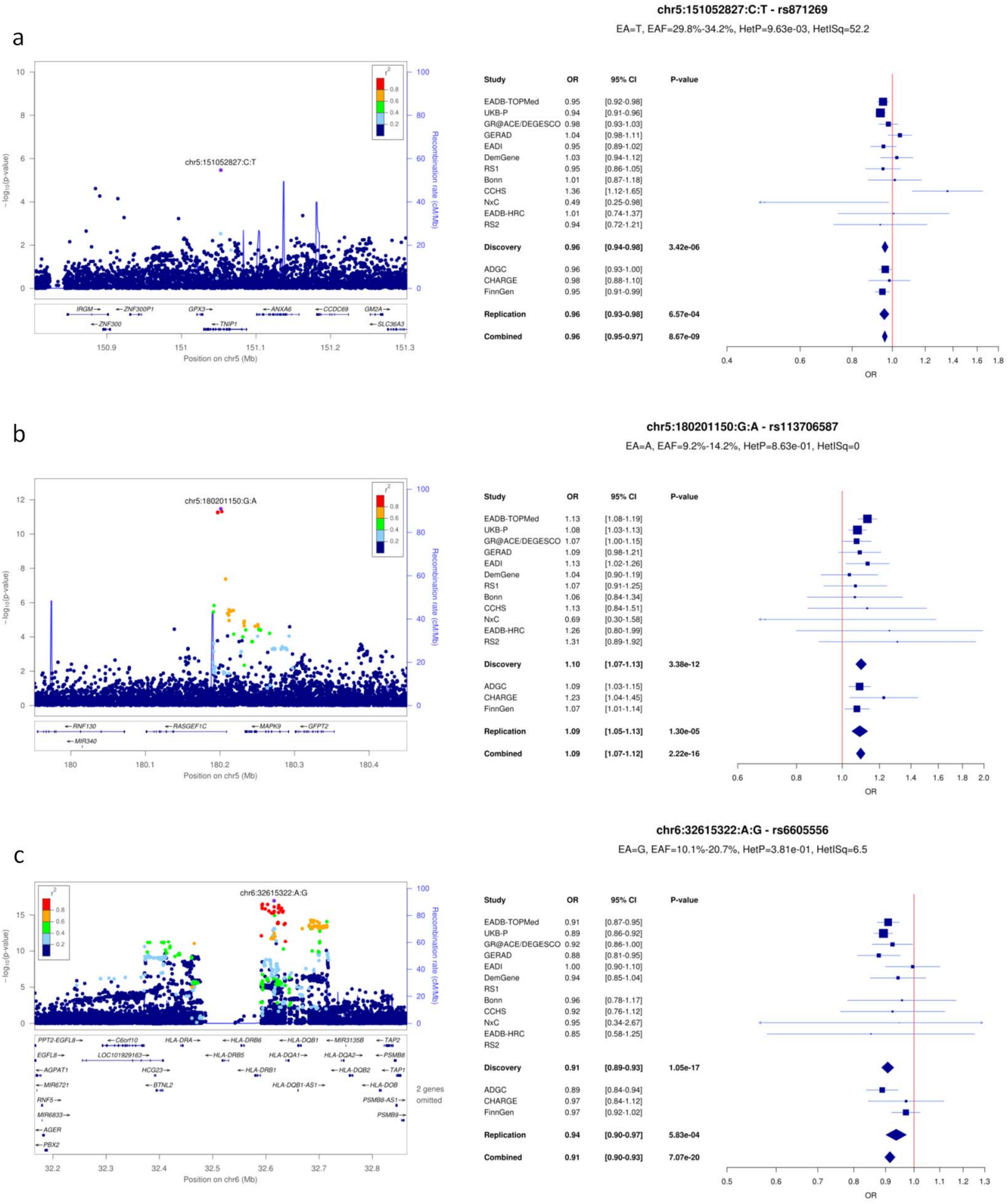

**Supplementary Figure 8**: LocusZoom and forest plots for (a) UNC5CL, (b) TREM2 (R62H) and (c) TREM2 (R47H) loci.

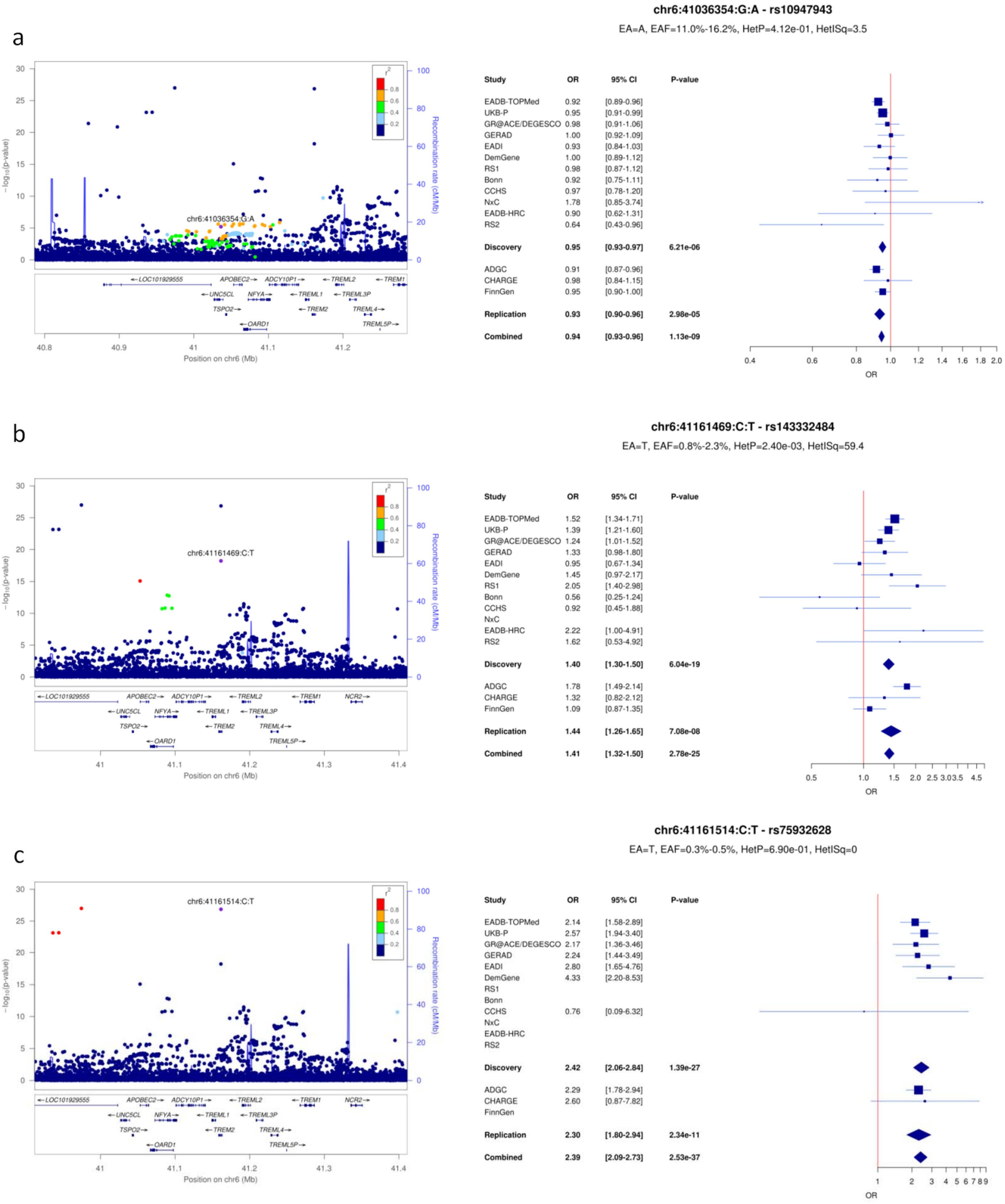

**Supplementary Figure 9:** LocusZoom and forest plots for (a) TREML2, (b) CD2AP and (c) HS3ST5 loci.

a

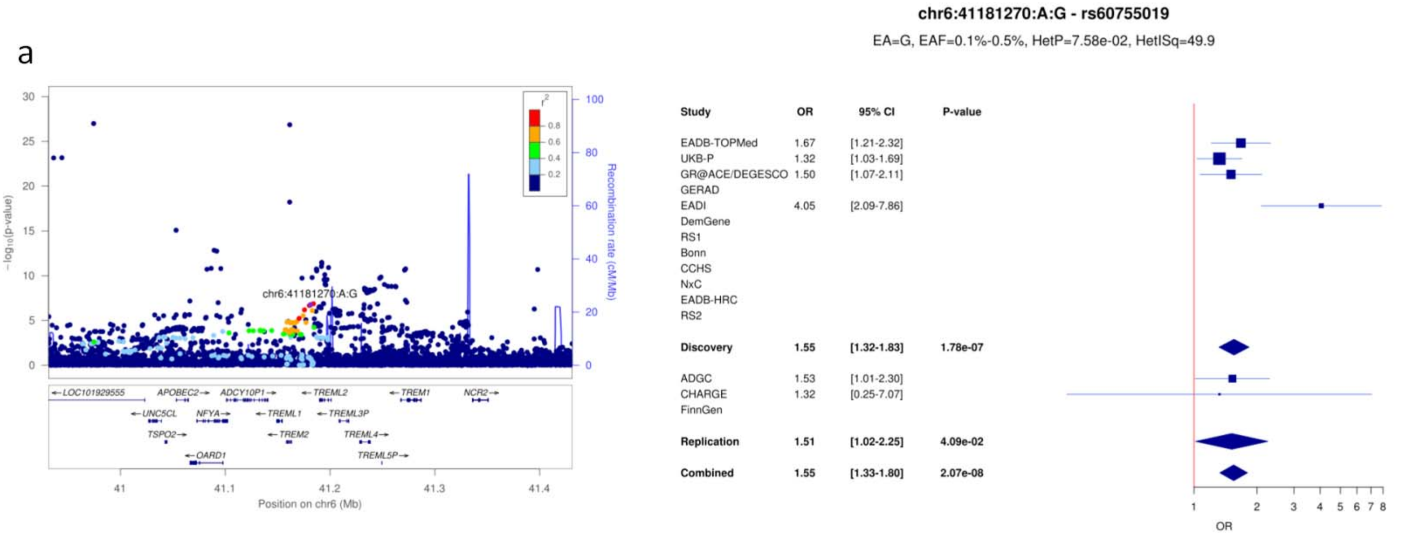


b

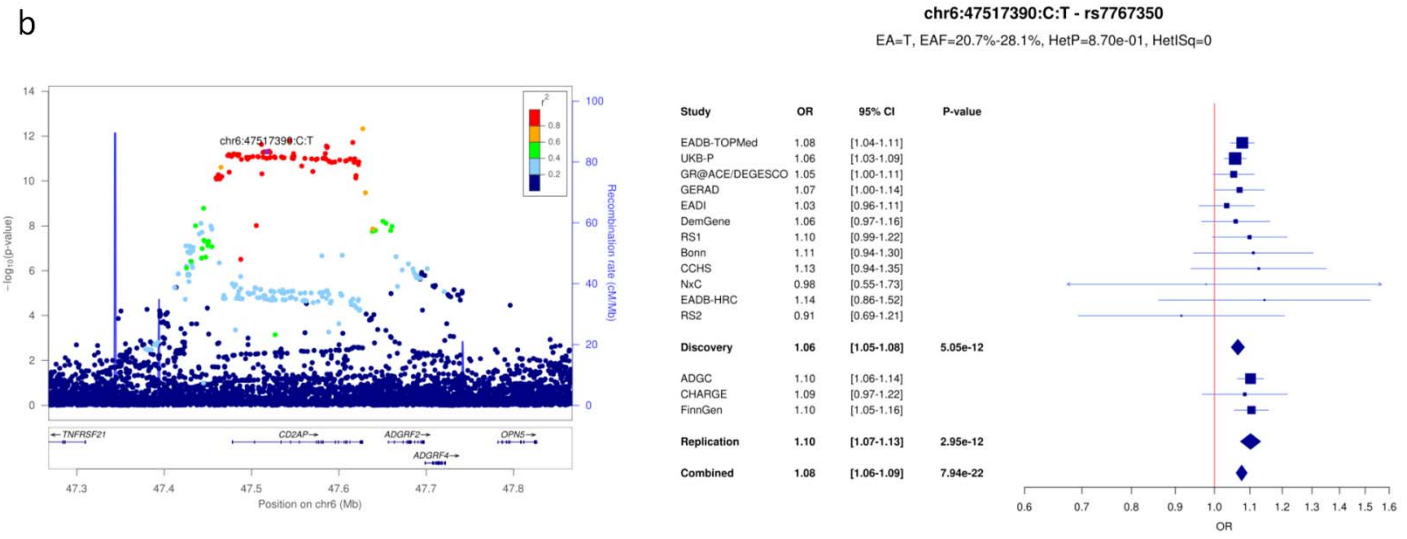


c

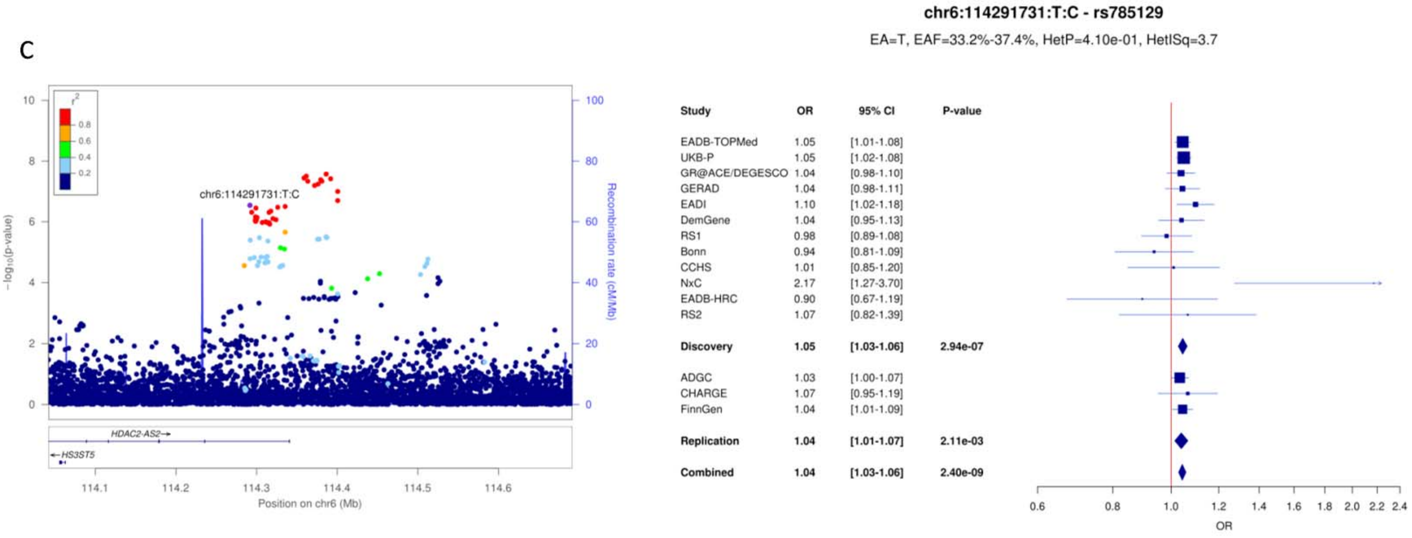

**Supplementary Figure 10**: LocusZoom and forest plots for (a) UMAD1, (b) ICA1 and (c) TMEM106B loci.

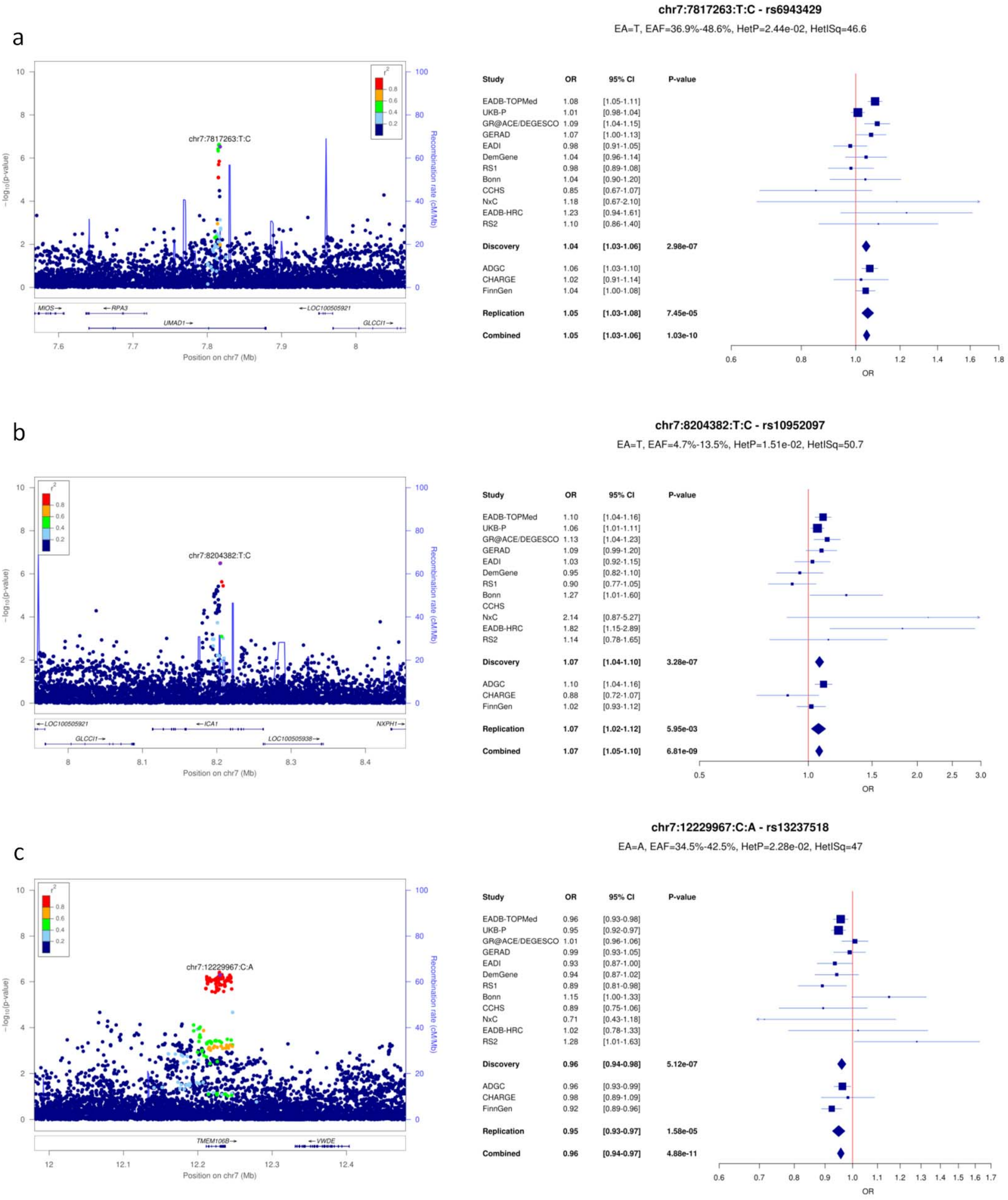

**Supplementary Figure 11**: LocusZoom and forest plots for (a) JAZF1, (b) NME8 and (c) SEC61G loci.

a

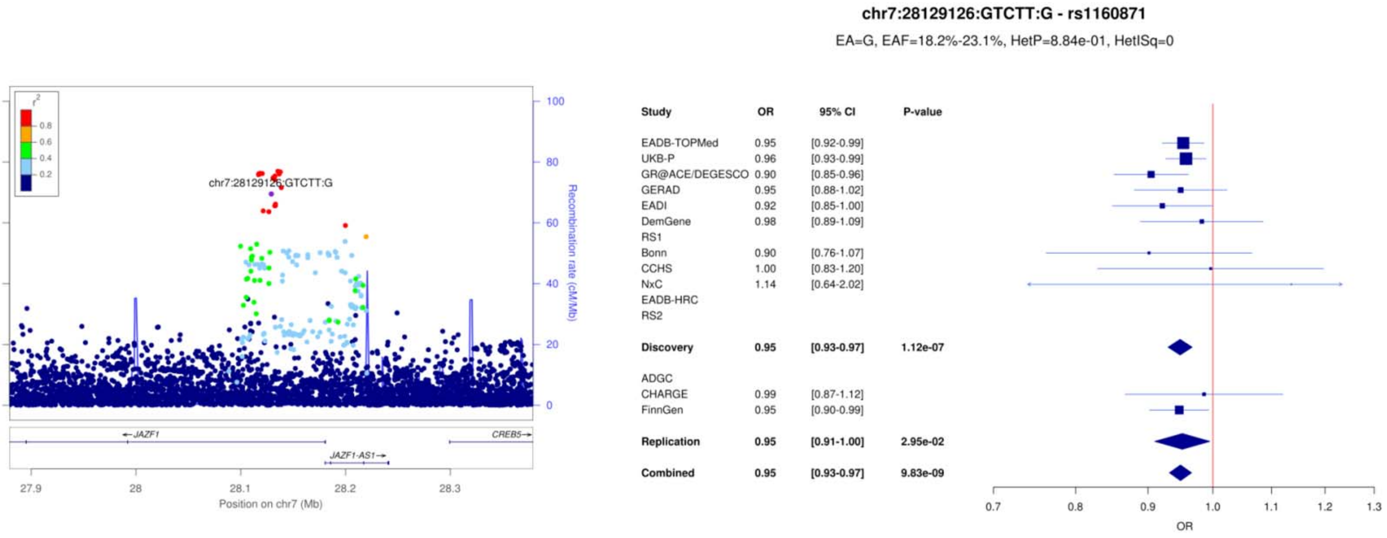


b

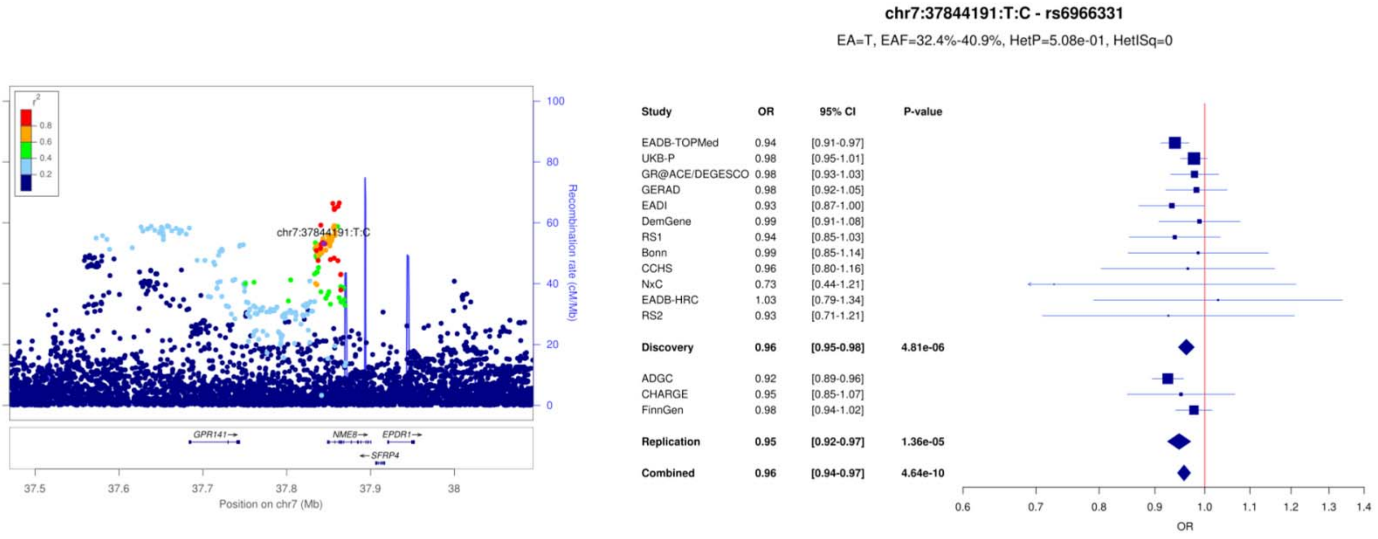


c

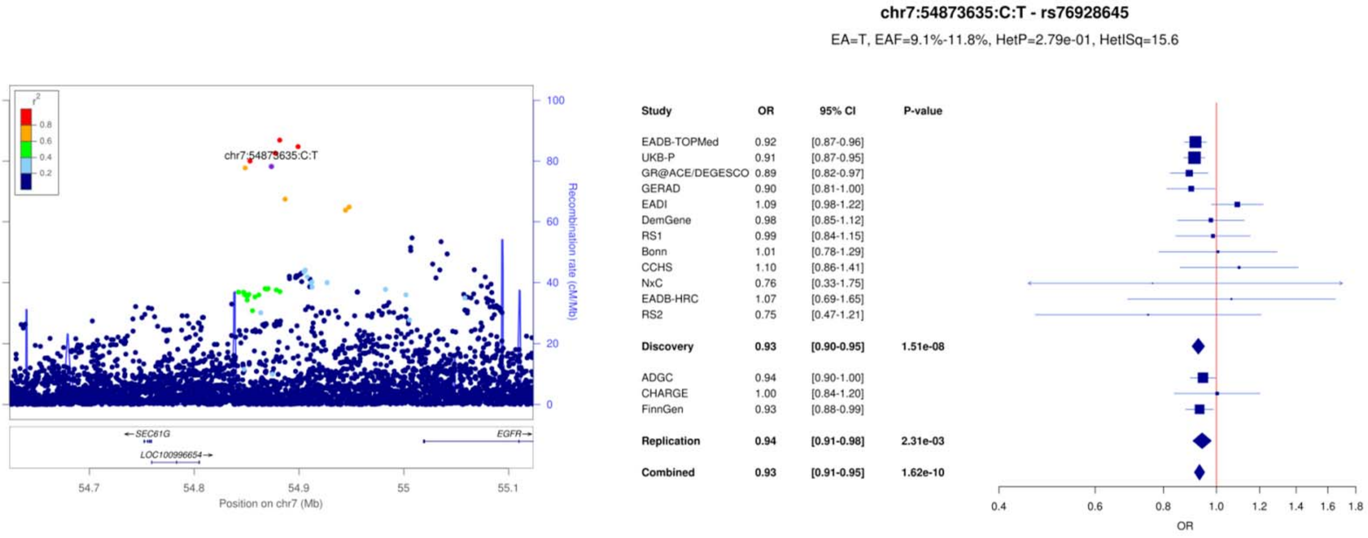

**Supplementary Figure 12**: LocusZoom and forest plots for (a) ZCWPW1/NYAP1, (b) EPHA1 and (c) CTSB loci.

a

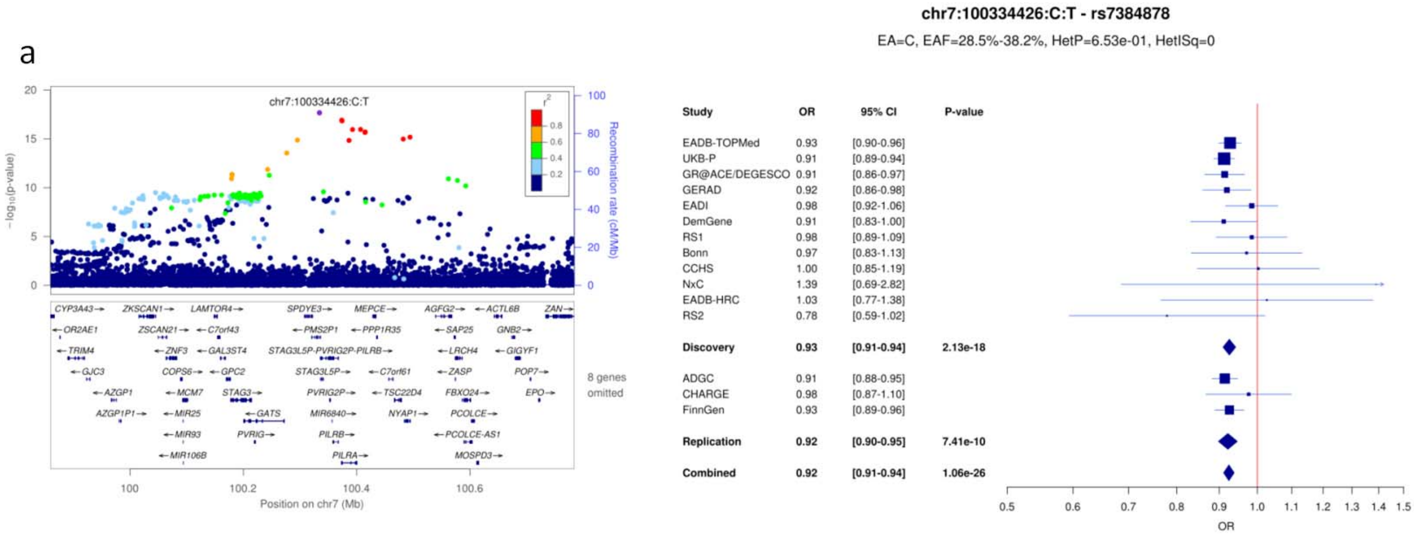


b

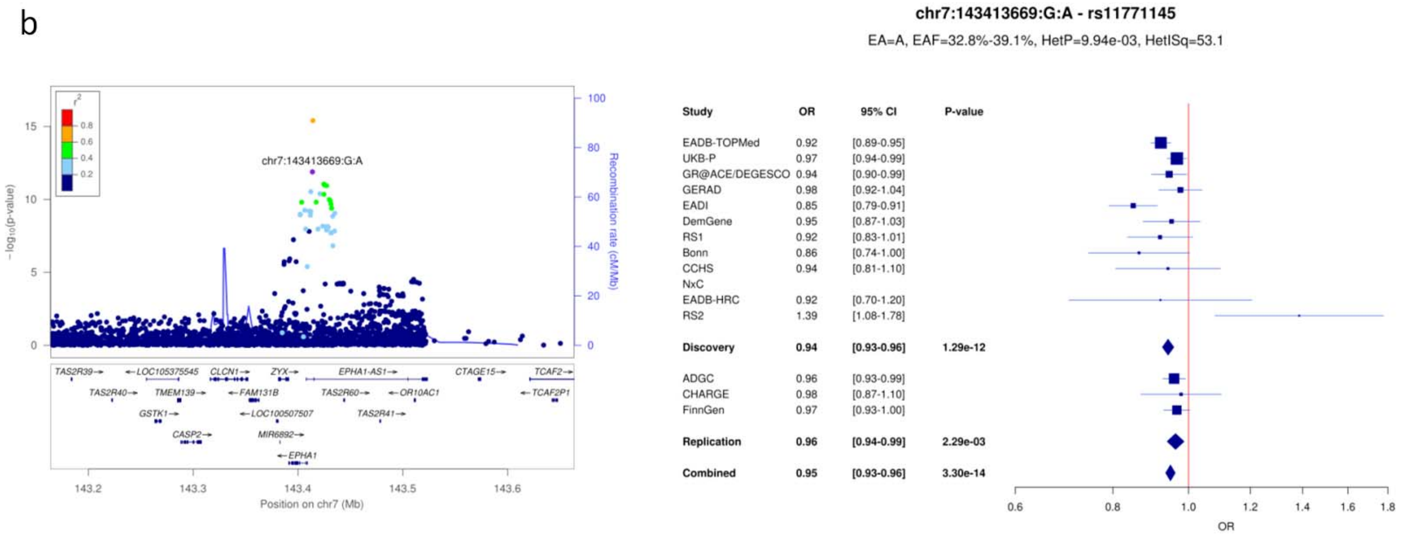


c

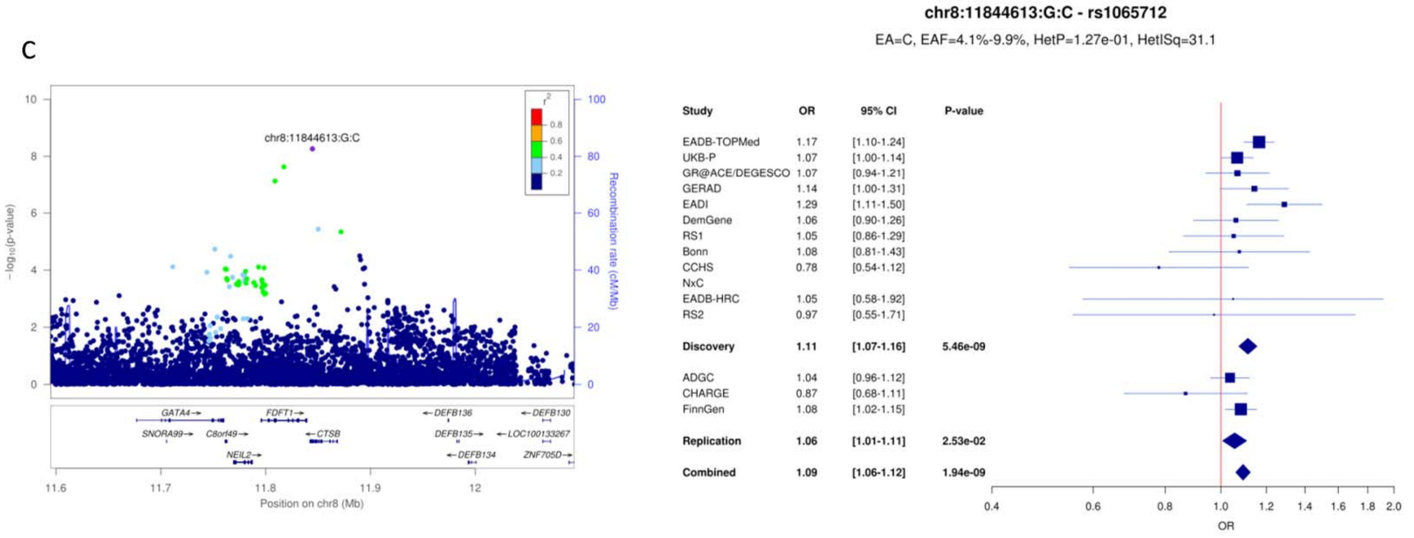

**Supplementary Figure 13**: LocusZoom and forest plots for (a) PTK2B, (b) CLU and (c) SHARPIN loci.

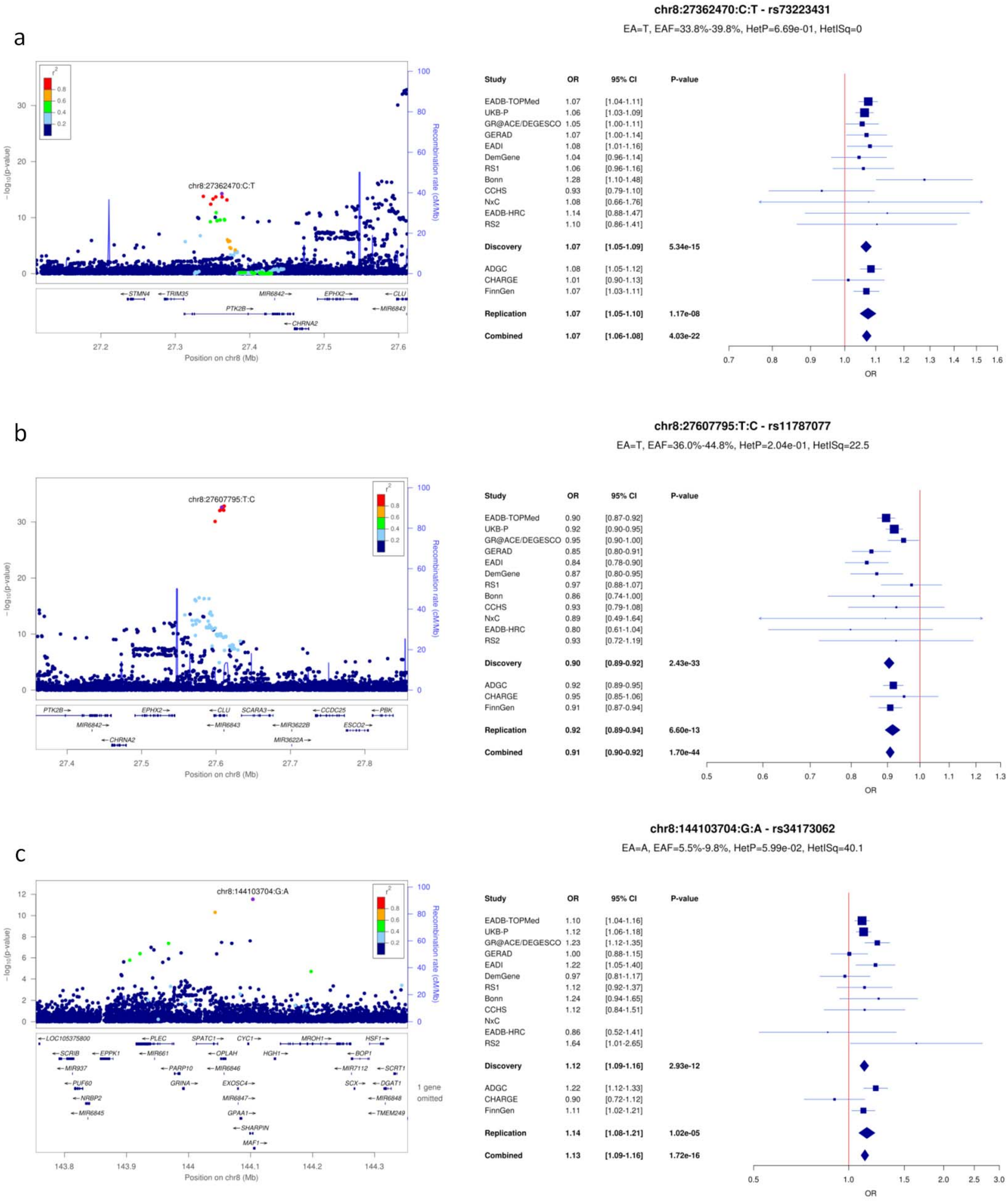

**Supplementary Figure 14**: LocusZoom and forest plots for (a) ABCA1, (b) USP6NL and (c) ANK3 loci.

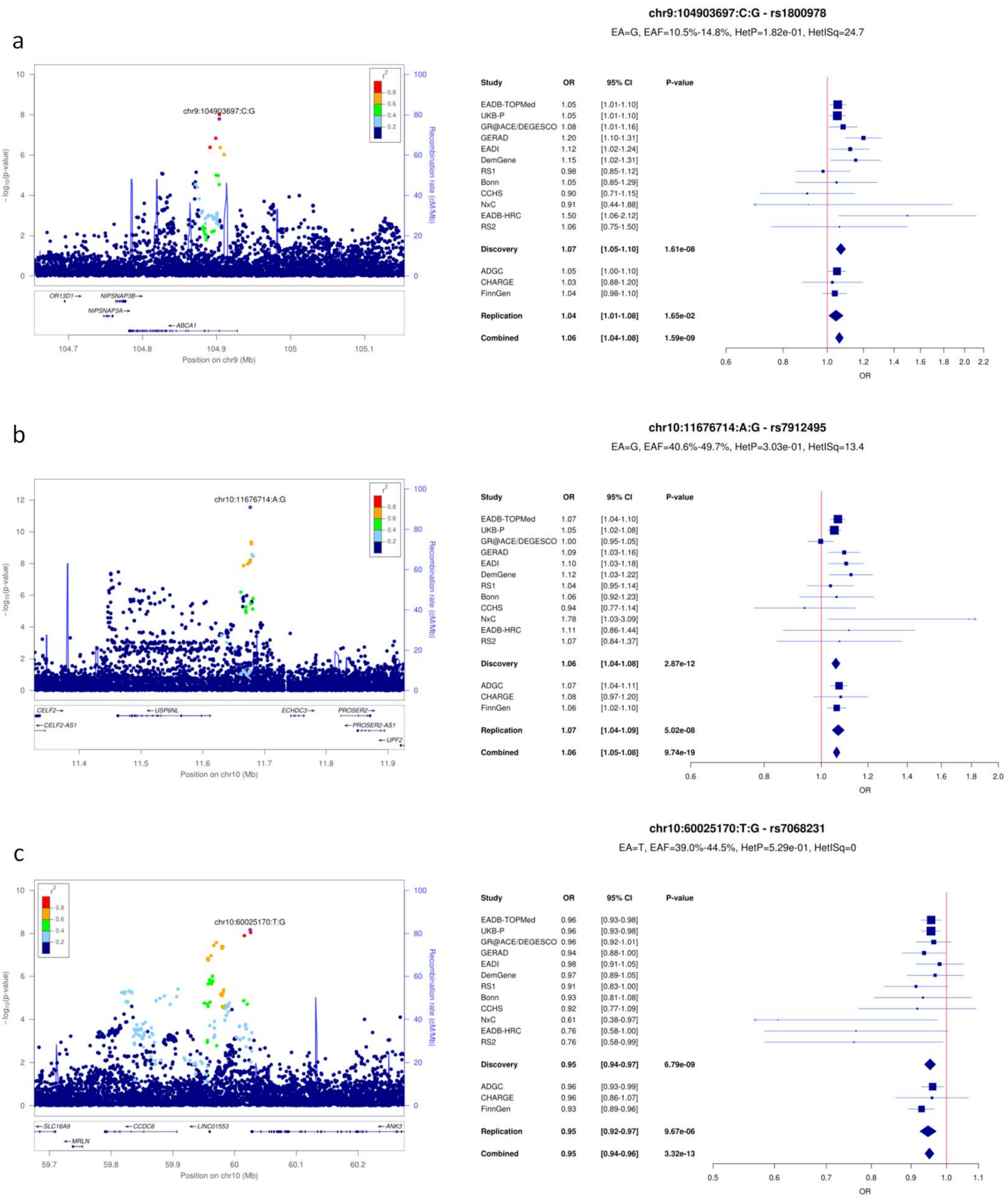

**Supplementary Figure 15**: LocusZoom and forest plots for (a) TSPAN14, (b) BLNK and (c) PLEKHA1 loci.

a

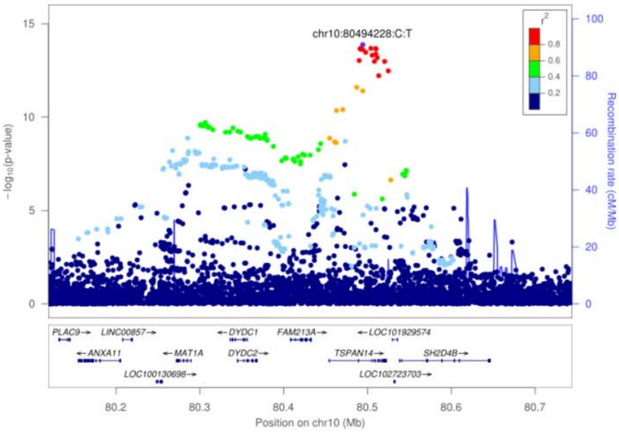


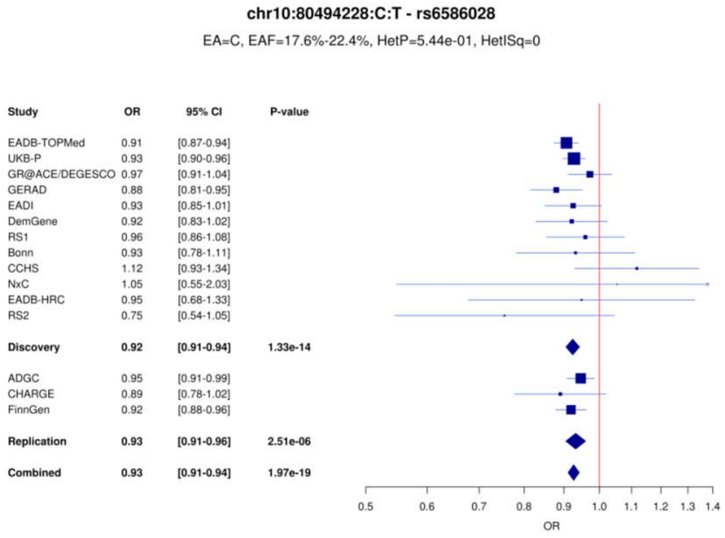


b

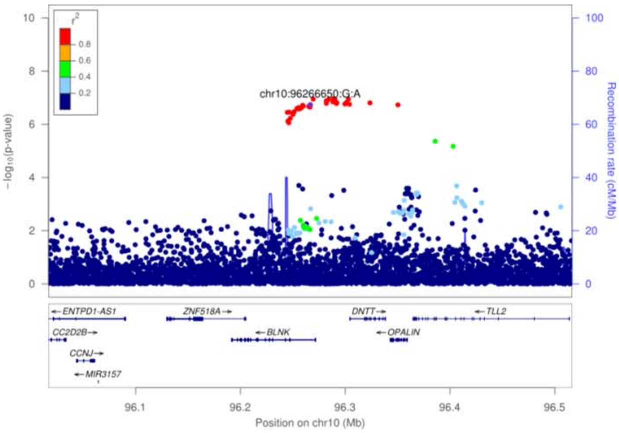


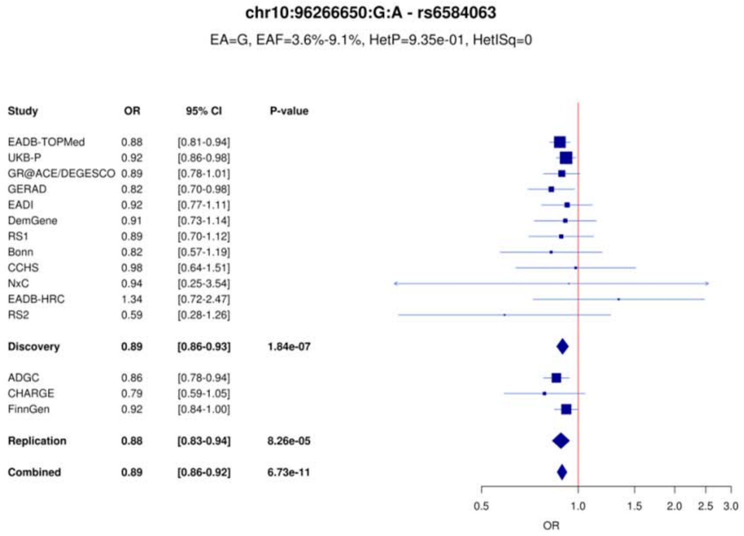


c

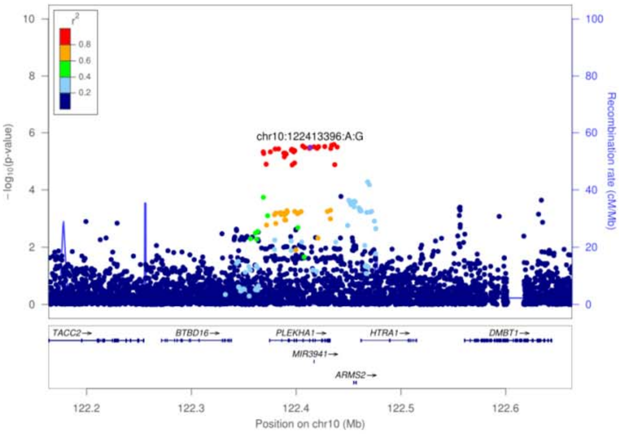


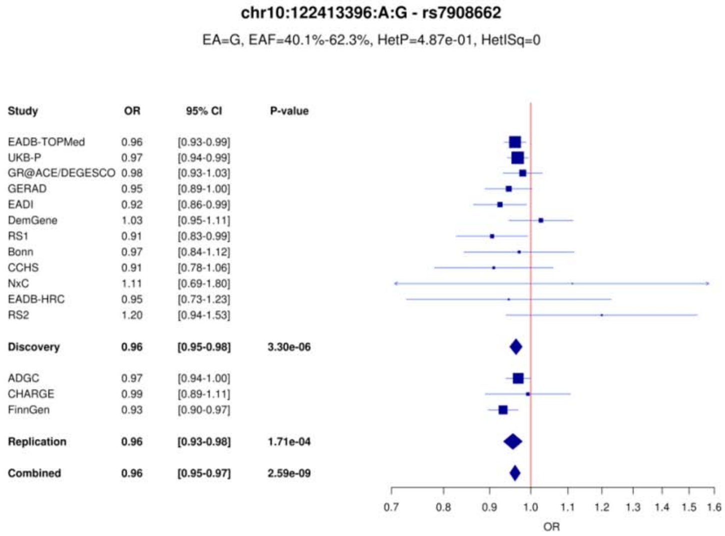

**Supplementary Figure 16**: LocusZoom and forest plots for (a) CELF1/SPI1, (b) MS4A and (c) PICALM loci.

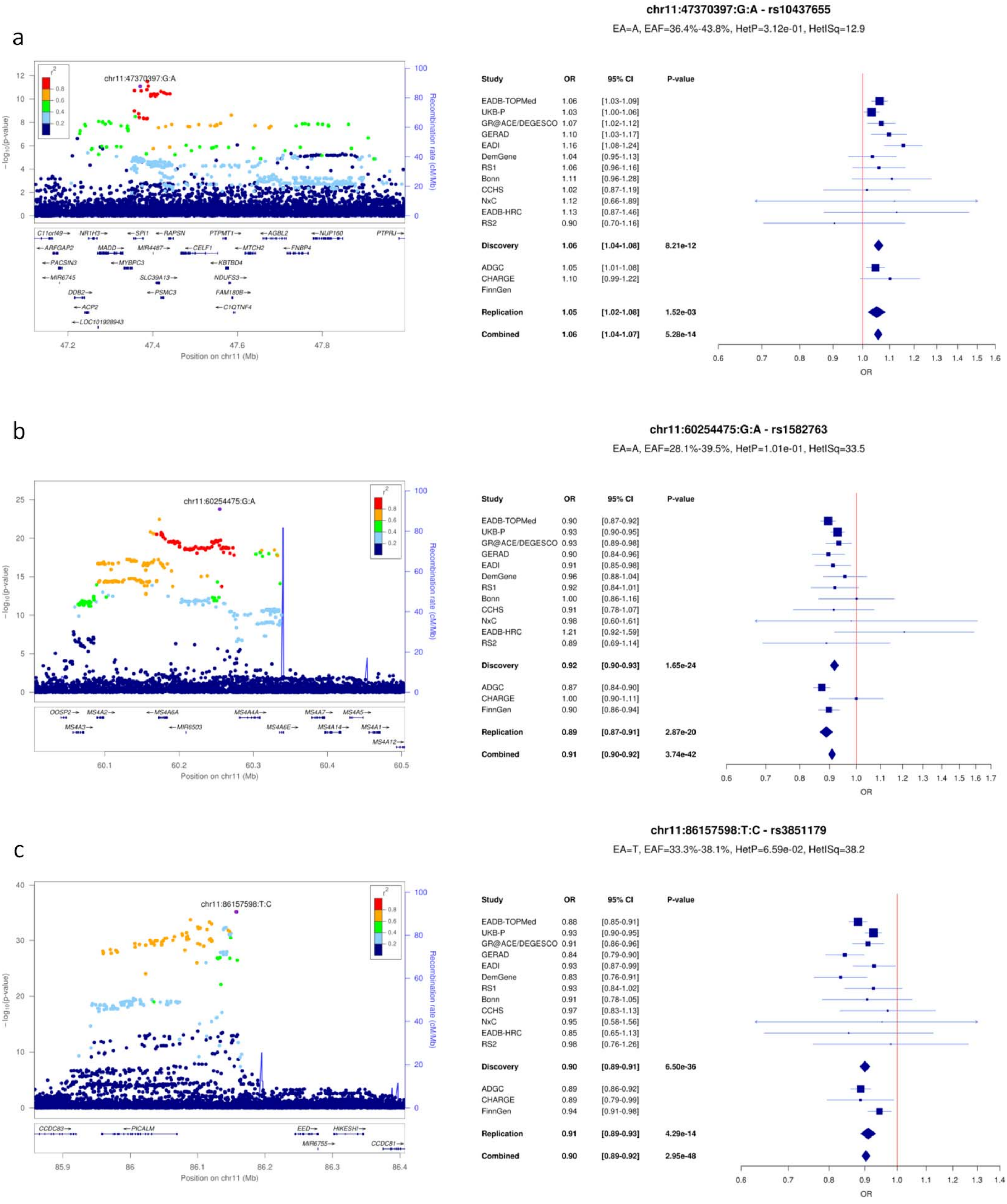

**Supplementary Figure 17**: LocusZoom and forest plots for (a) SORL1 (1), (b) SORL1 (2) and (c) TPCN1 loci.

a

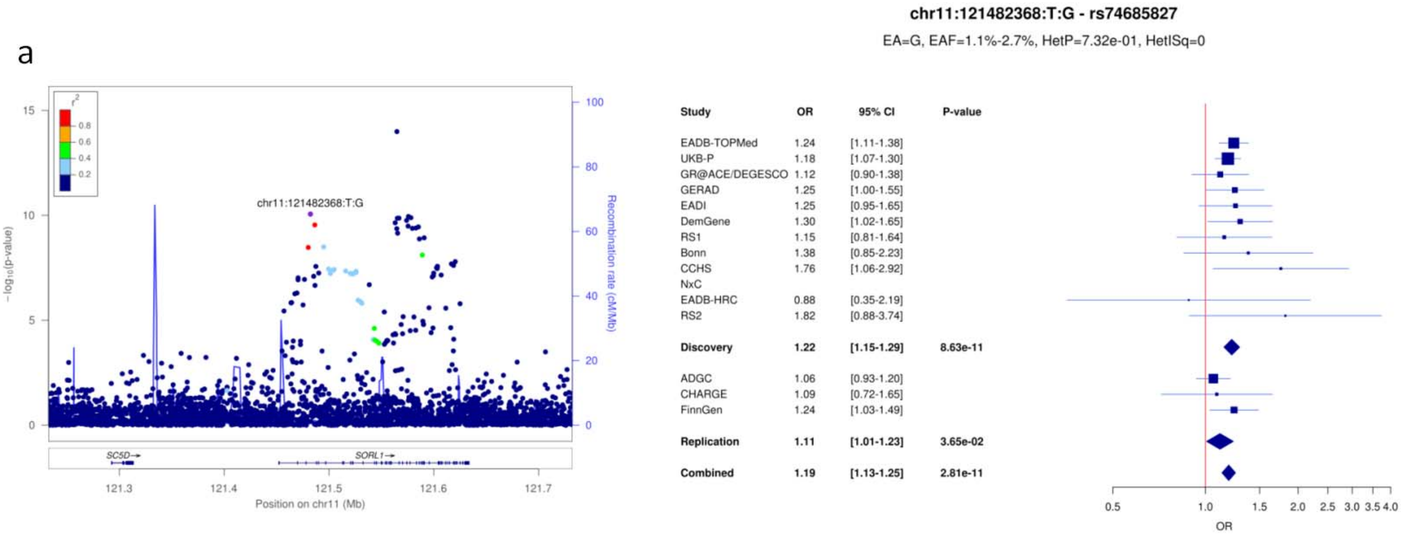


b

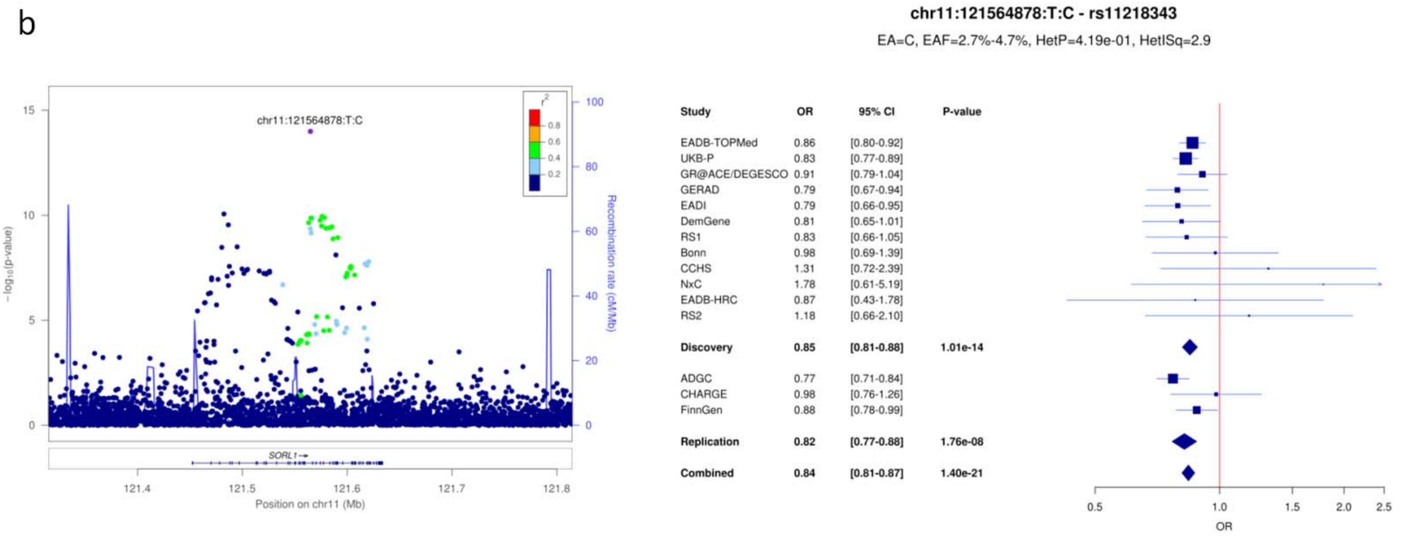


c

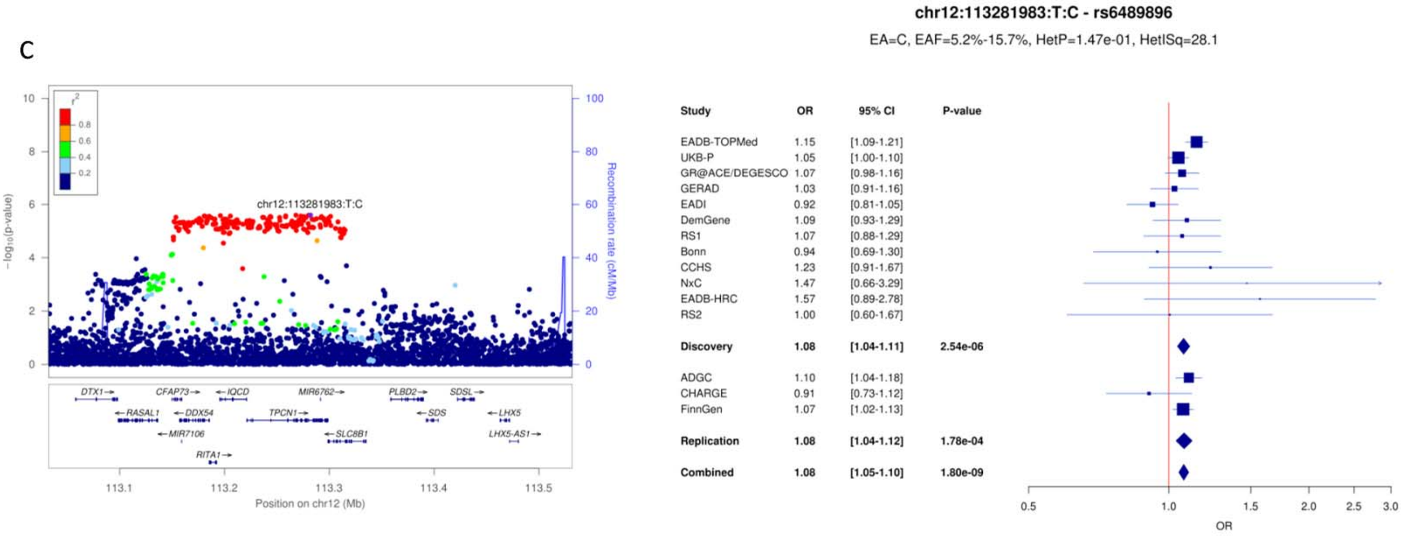

**Supplementary Figure 18**: LocusZoom and forest plots for (a) FERMT2, (b) SLC24A4/RIN3 (1) and (c) SLC24A4/RIN3 (2) loci.

a

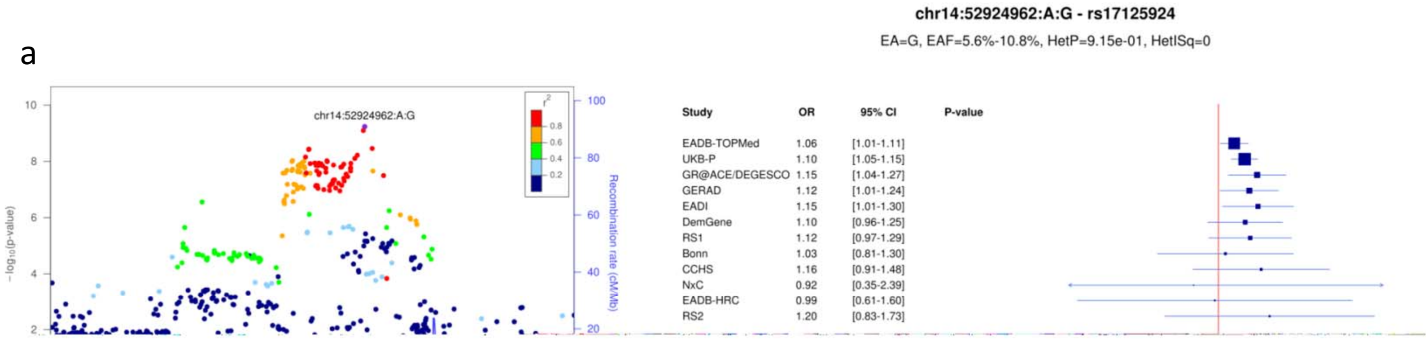


b

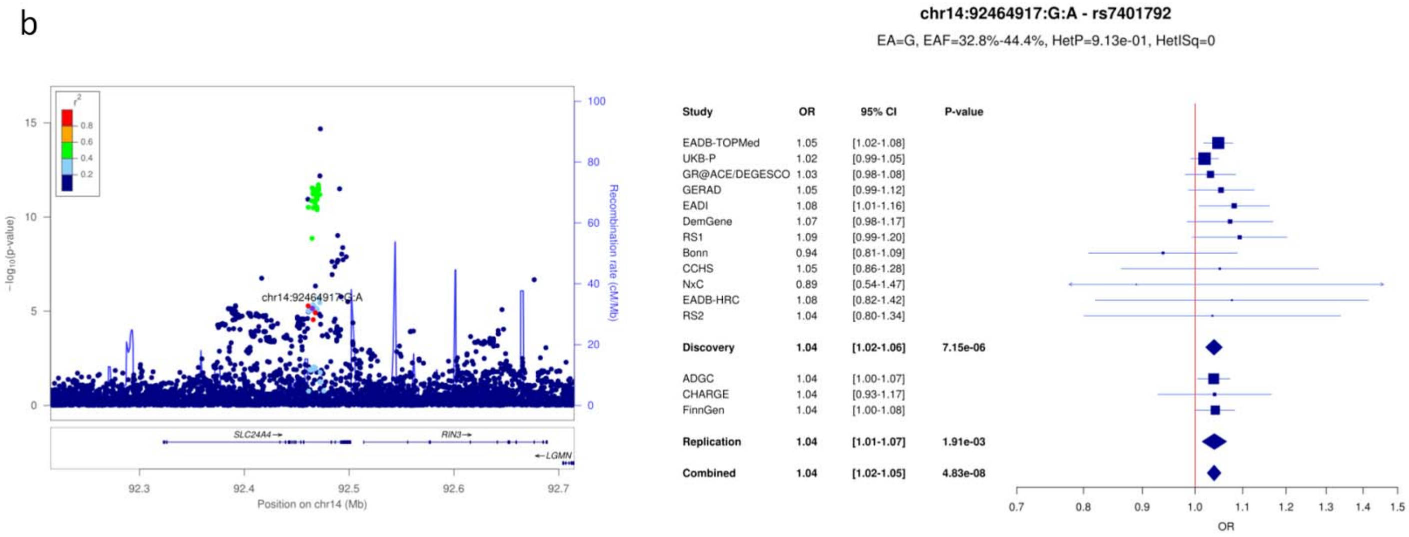


c

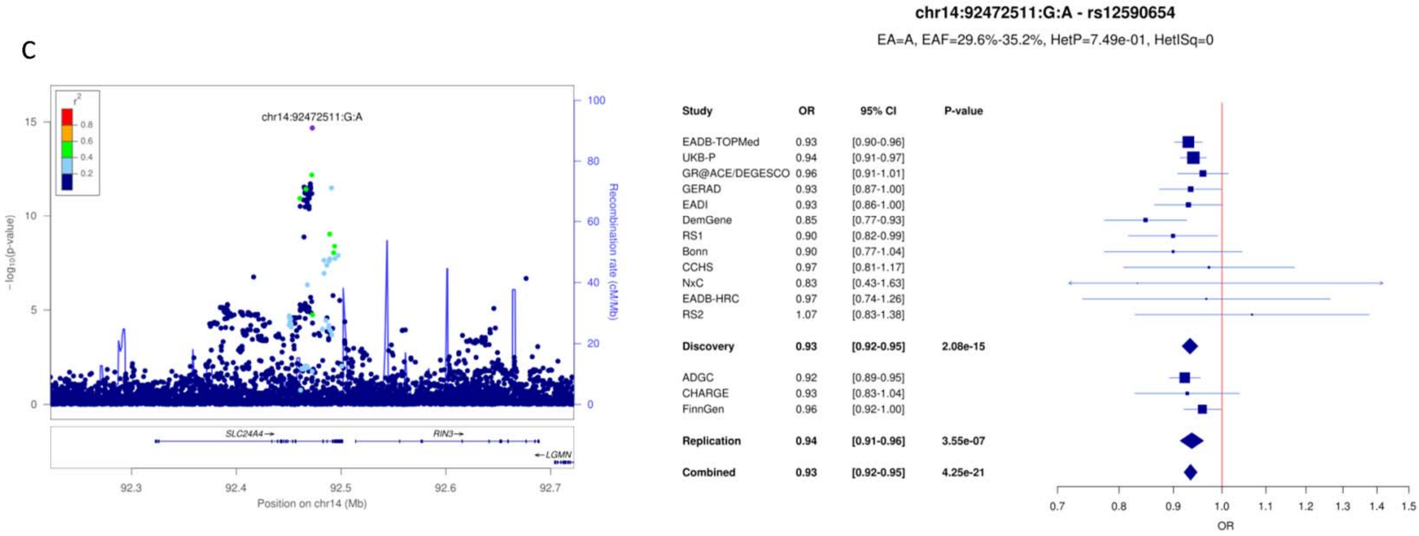

**Supplementary Figure 19**: LocusZoom and forest plots for (a) IGH gene cluster (1), (b) IGH gene cluster (2) and (c) SPPL2A loci.

a

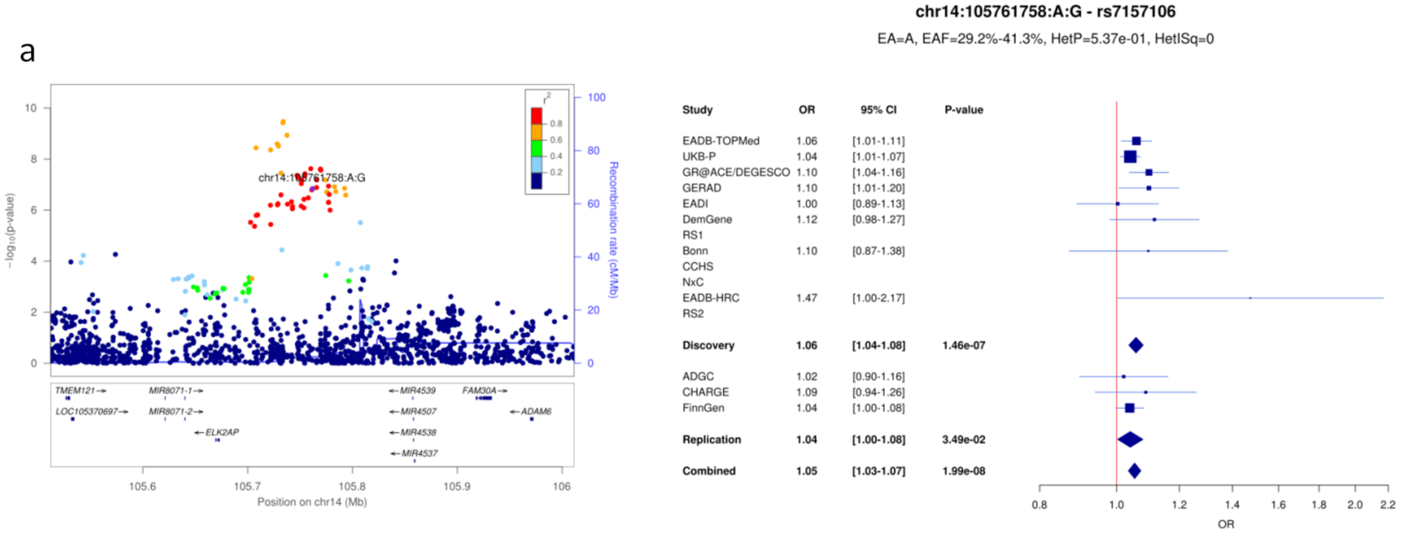


b

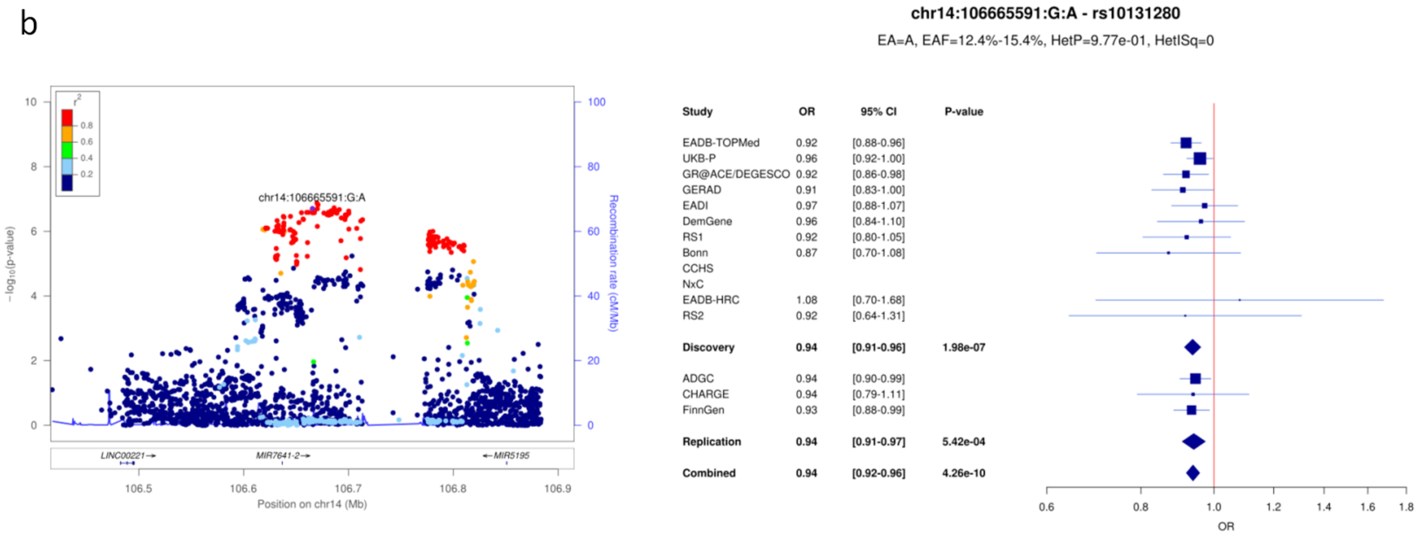


c

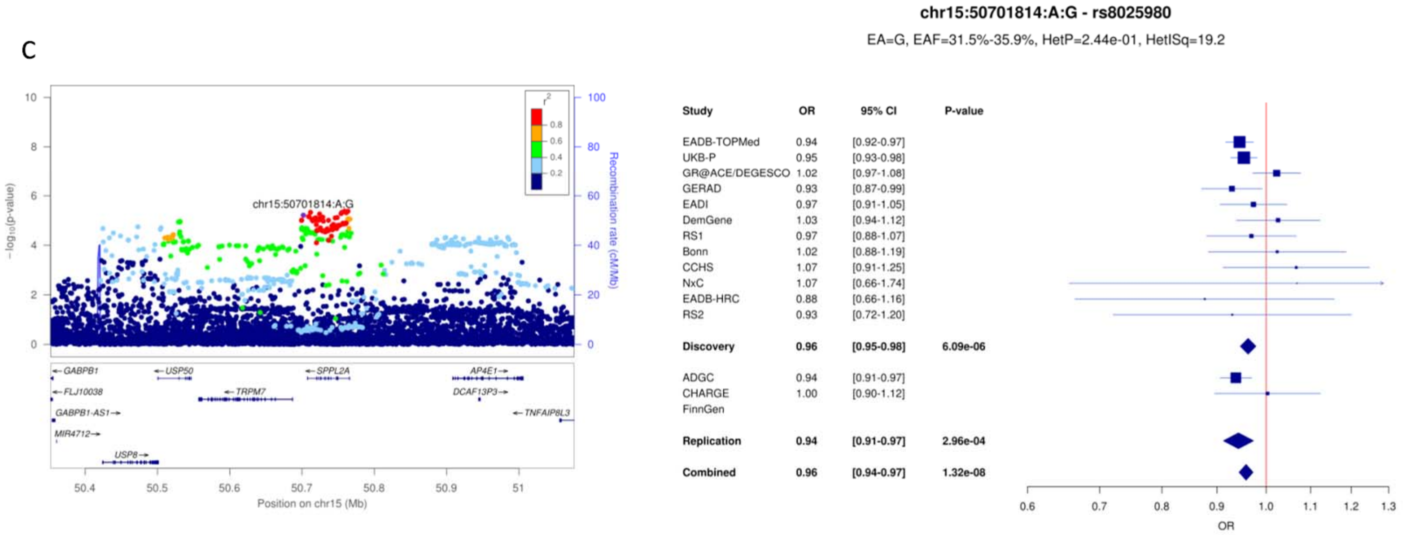

**Supplementary Figure 20**: LocusZoom and forest plots for (a) ADAM10, (b) APH1B and (c) SNX1 loci.

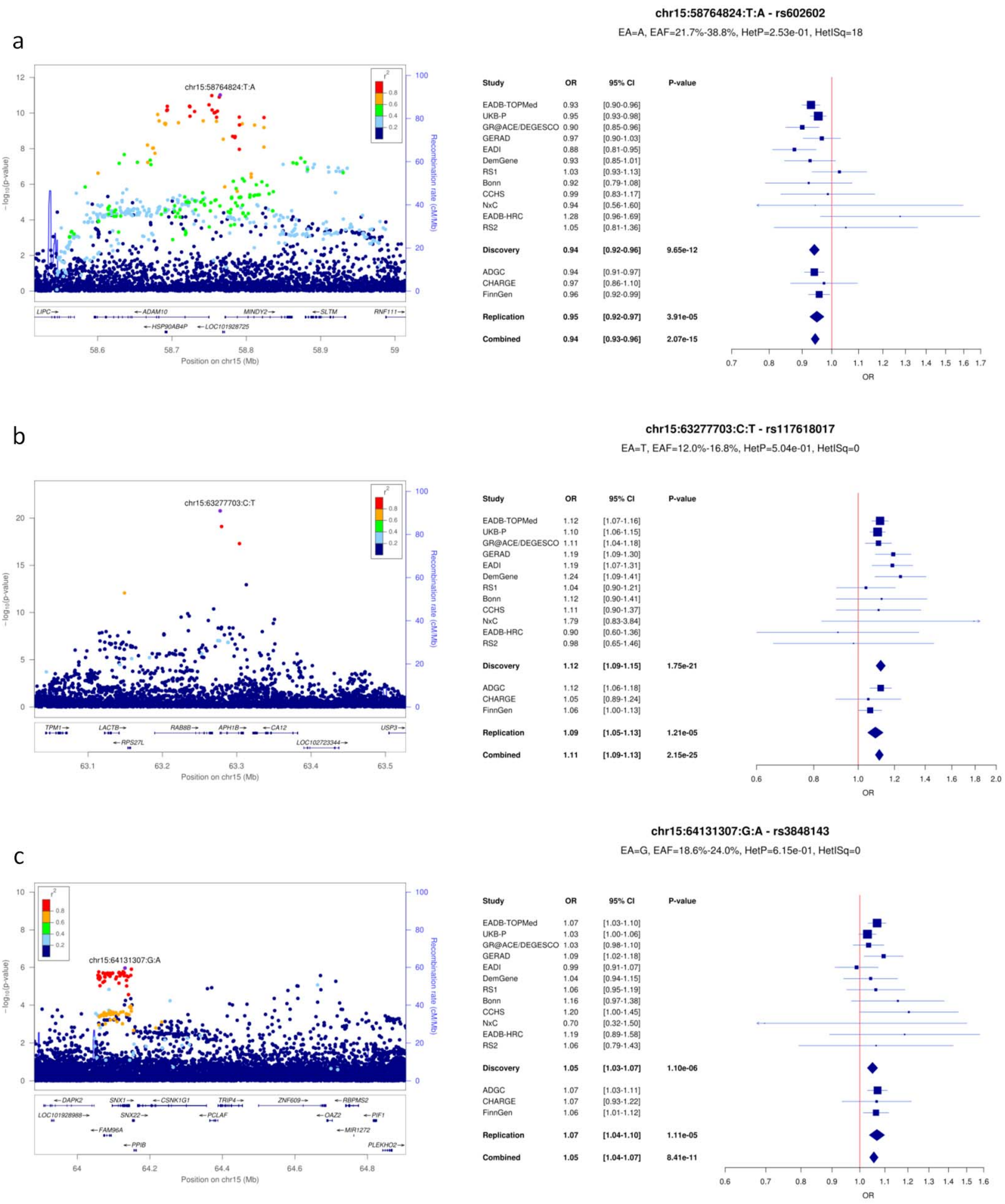

**Supplementary Figure 21**: LocusZoom and forest plots for (a) CTSH, (b) DOC2A and (c) KAT8 loci.

a

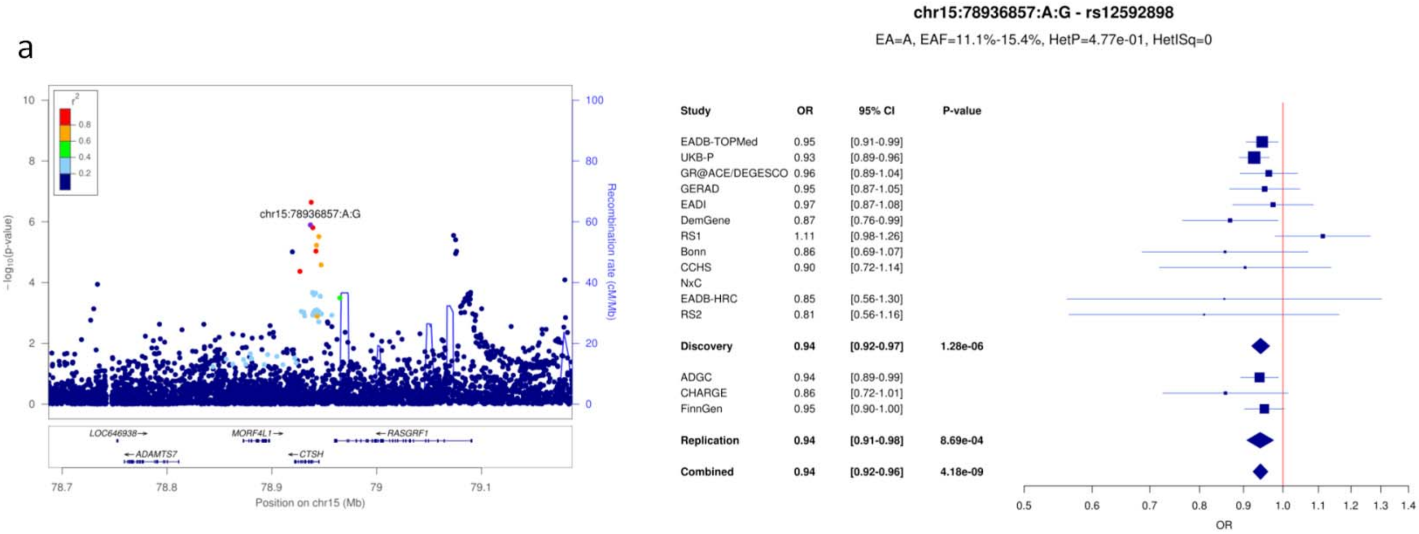


b

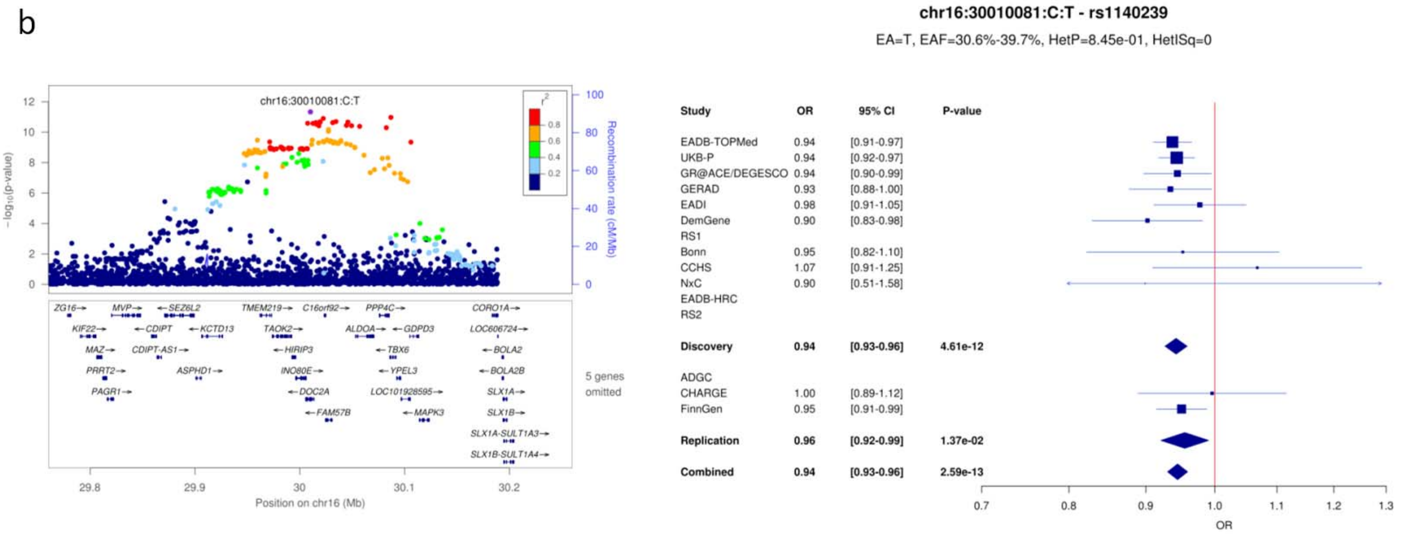


c

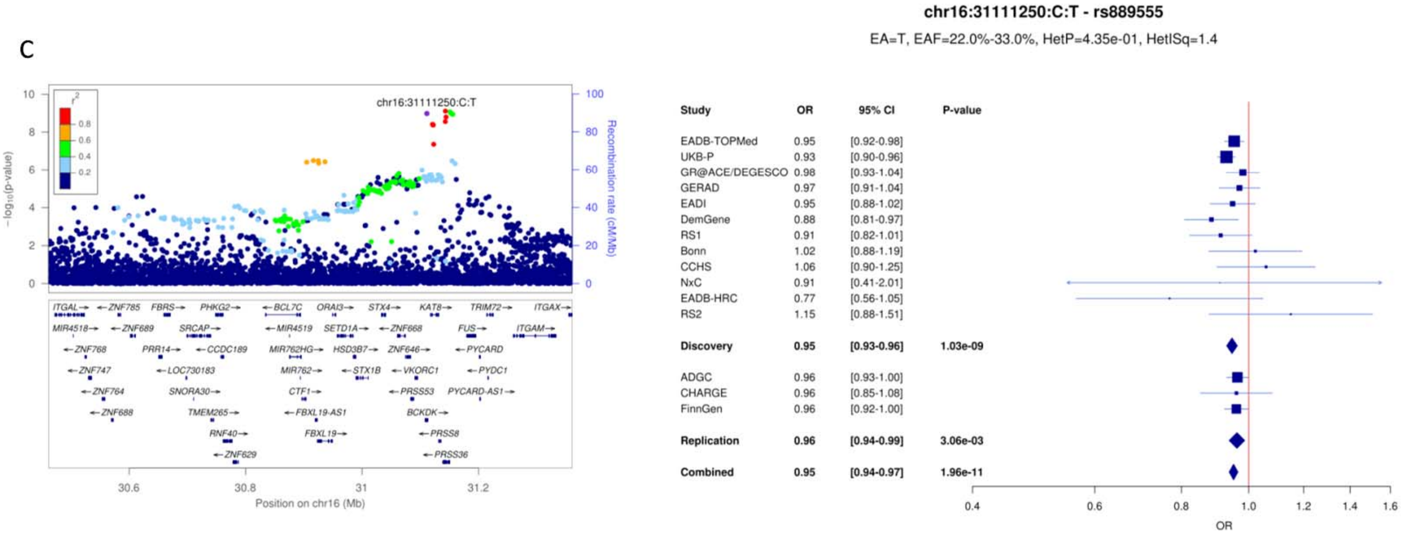

**Supplementary Figure 22**: LocusZoom and forest plots for (a) IL34, (b) MAF and (c) PLCγ2 (1) loci.

a

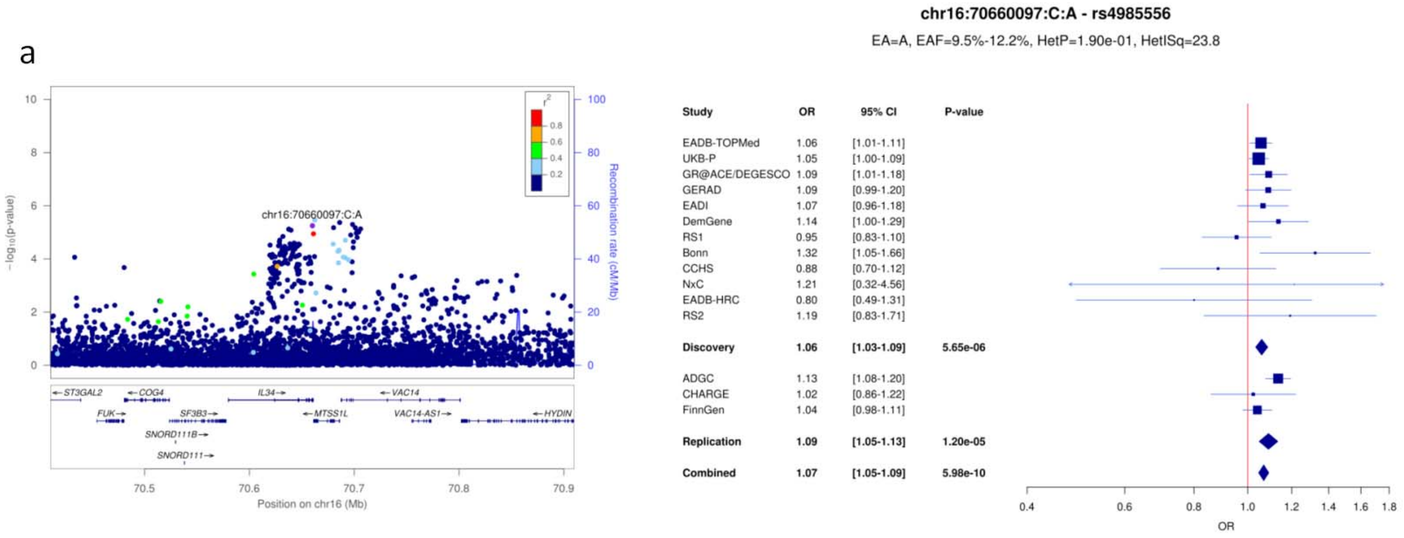


b

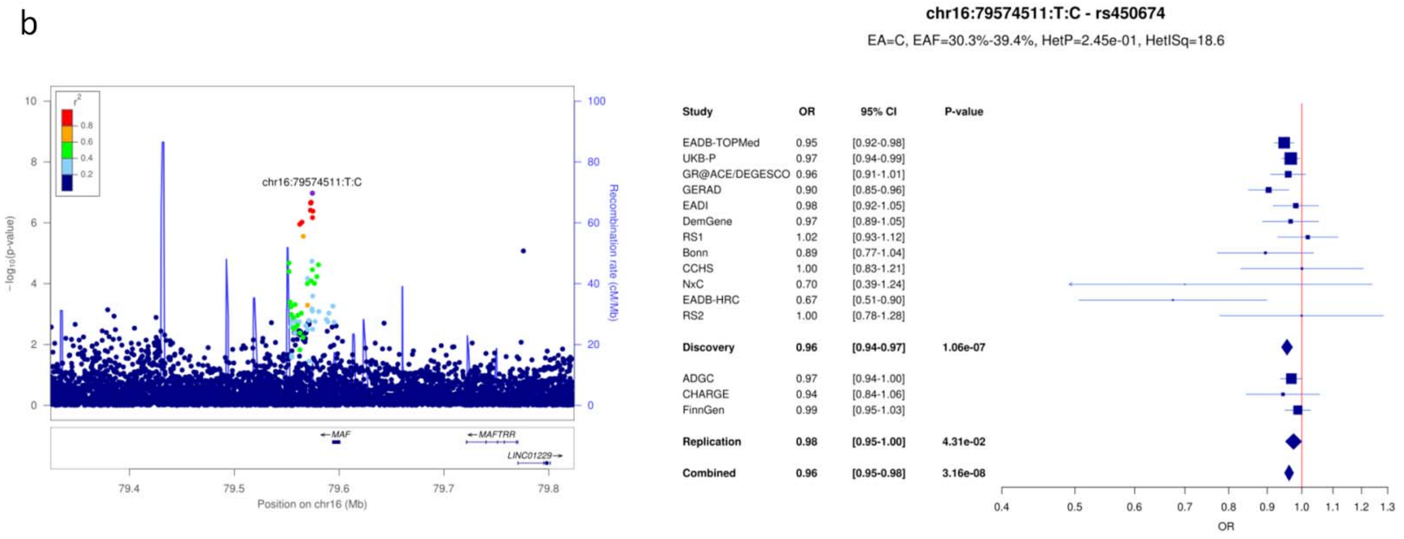


c

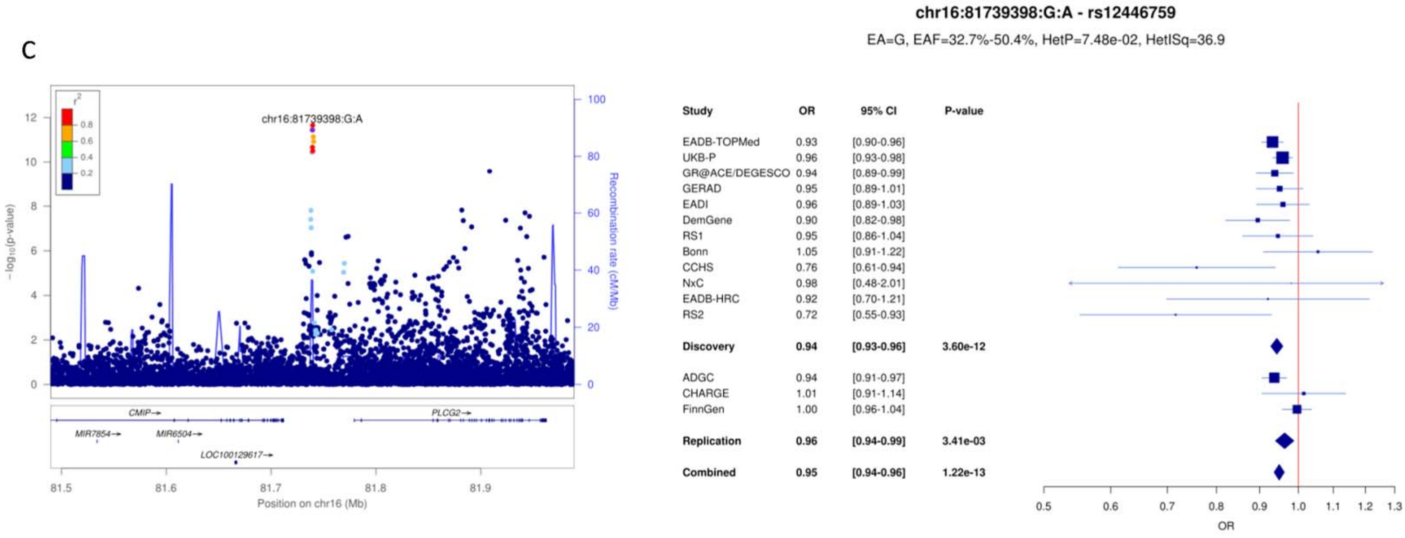

**Supplementary Figure 23**: LocusZoom and forest plots for (a) PLCγ2 (2), (b) FOXF1 and (c) PRDM7 loci.

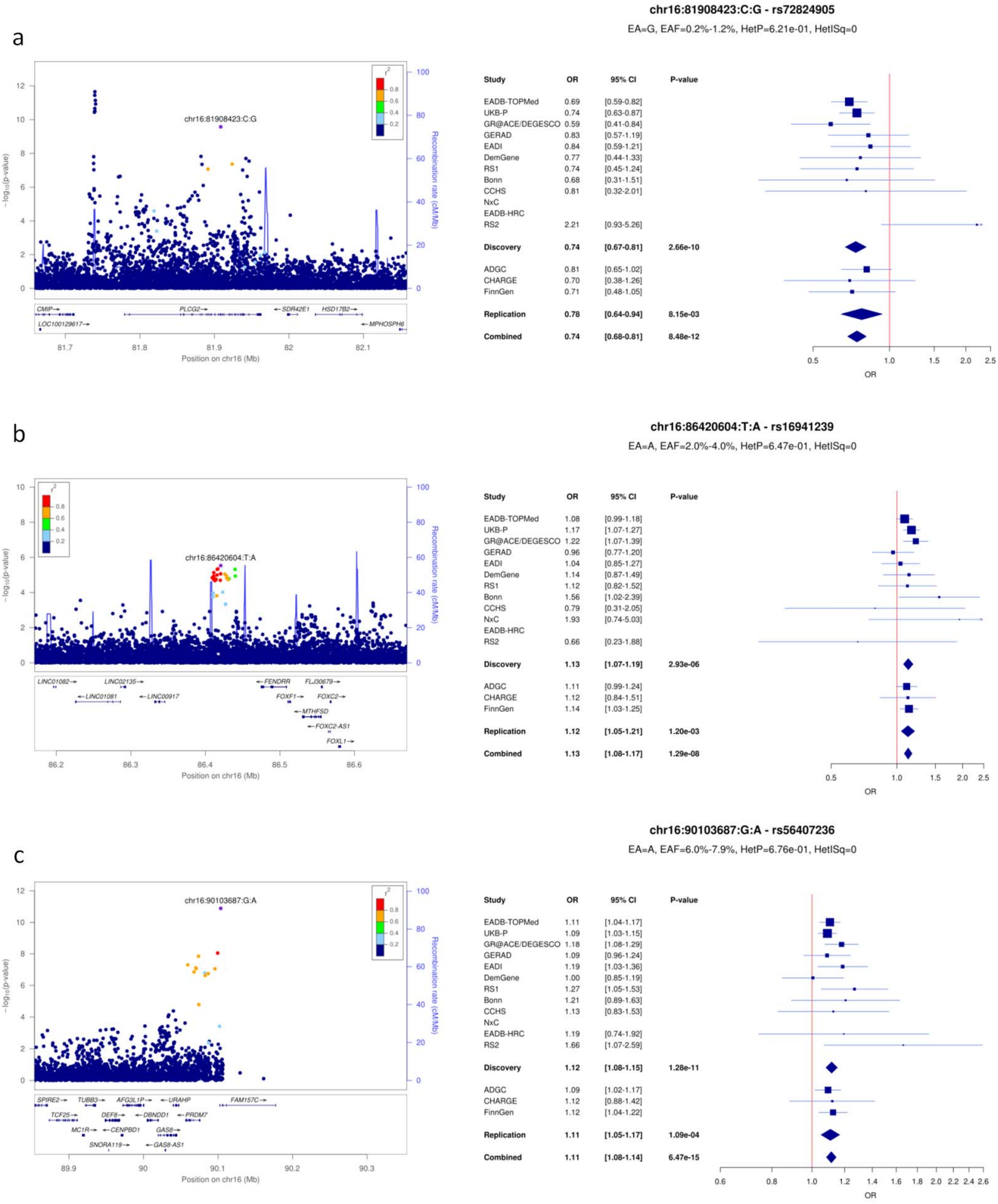

**Supplementary Figure 24**: LocusZoom and forest plots for (a) WDR81, (b) SCIMP/RABEP1 and (c) MYO15A loci.

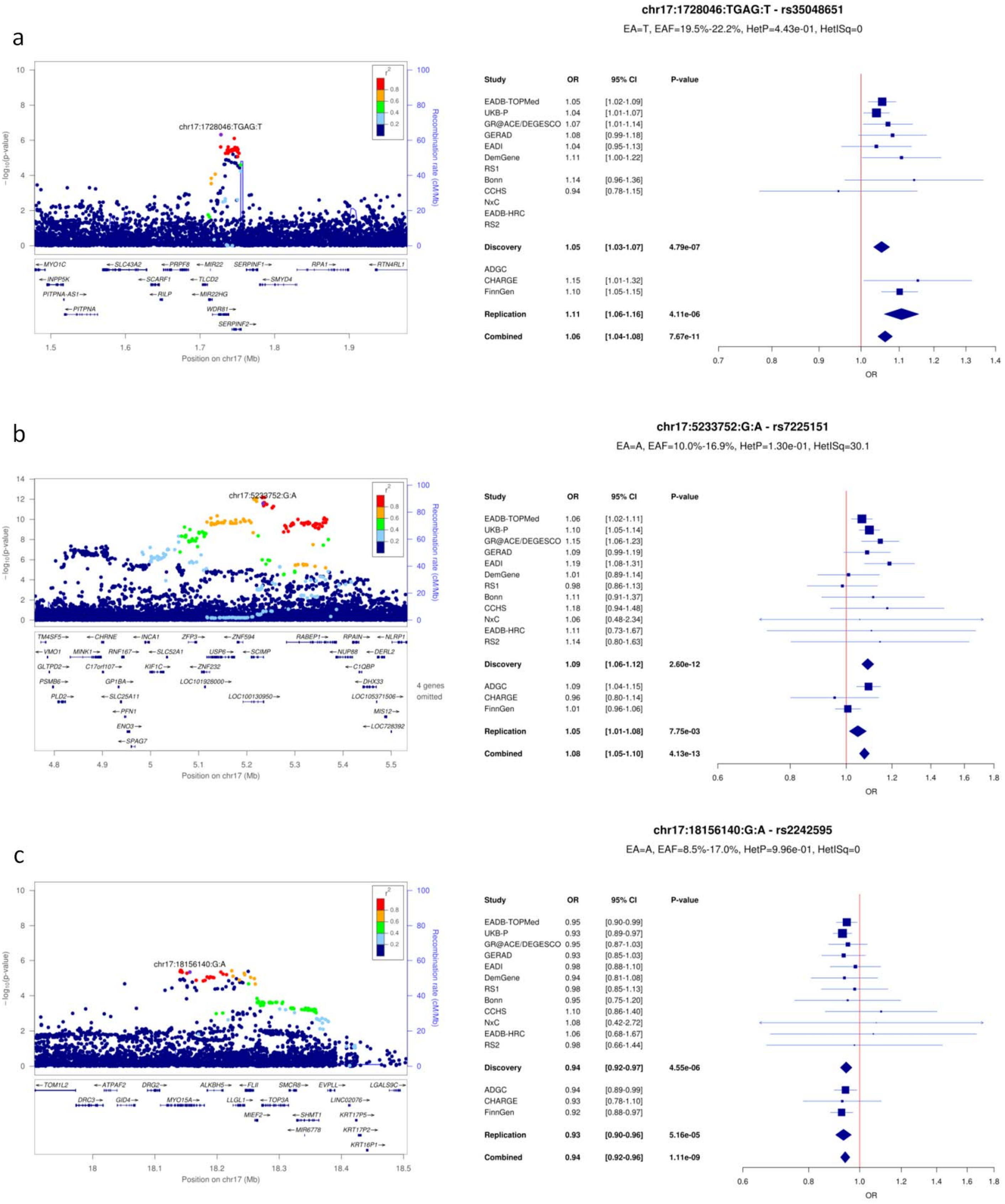

**Supplementary Figure 25**: LocusZoom and forest plots for (a) GRN, (b) MAPT and (c) ABI3 loci.

a

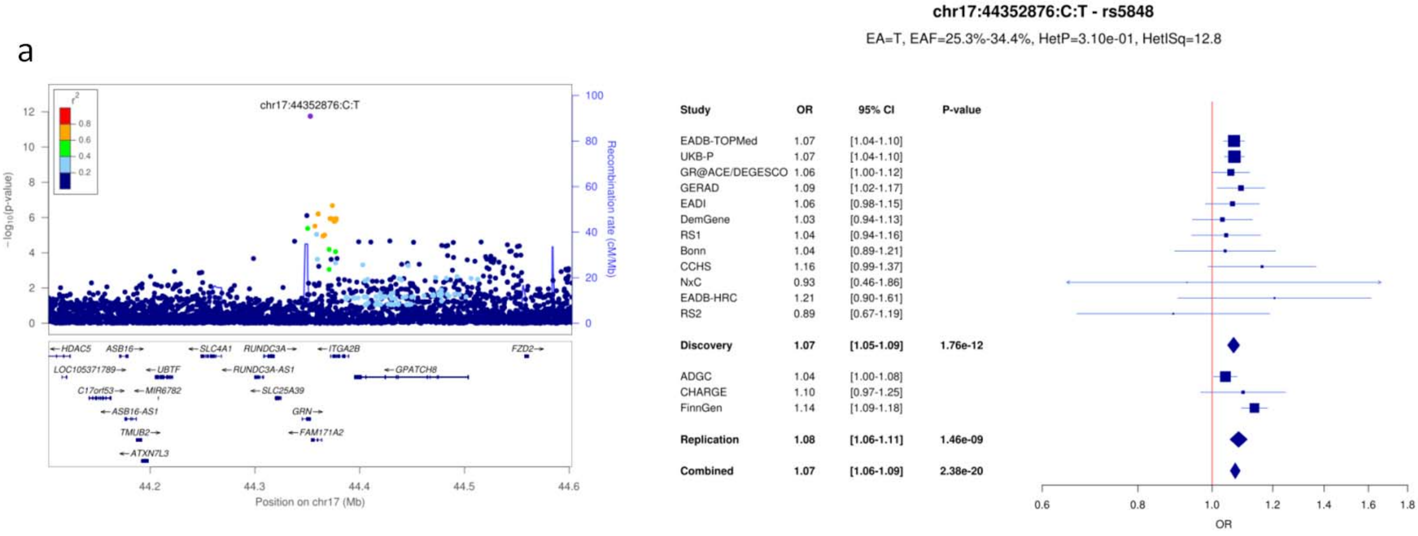


b

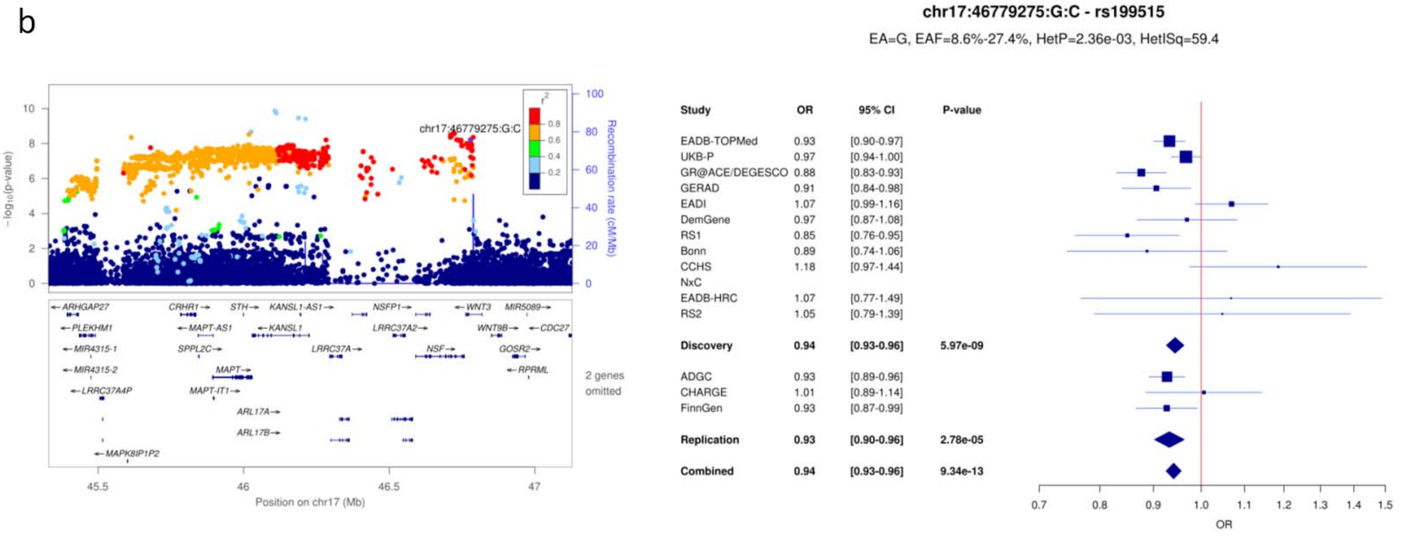


c

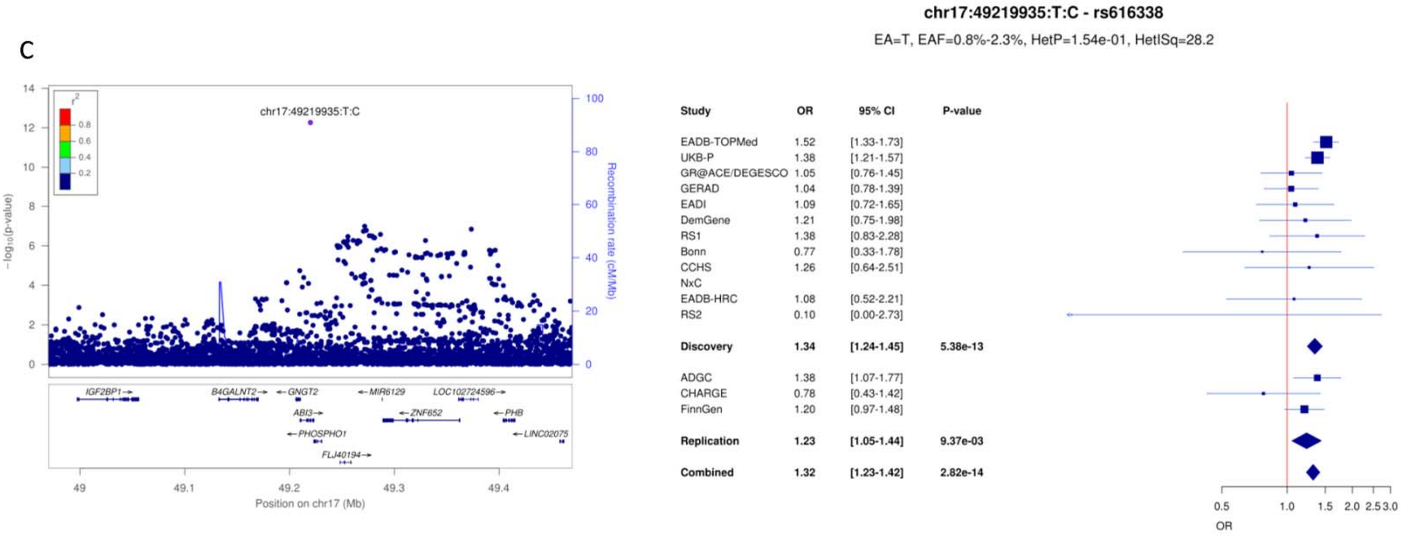

**Supplementary Figure 26**: LocusZoom and forest plots for (a) TSPOAP1, (b) ACE and (c) ABCA7 loci.

a

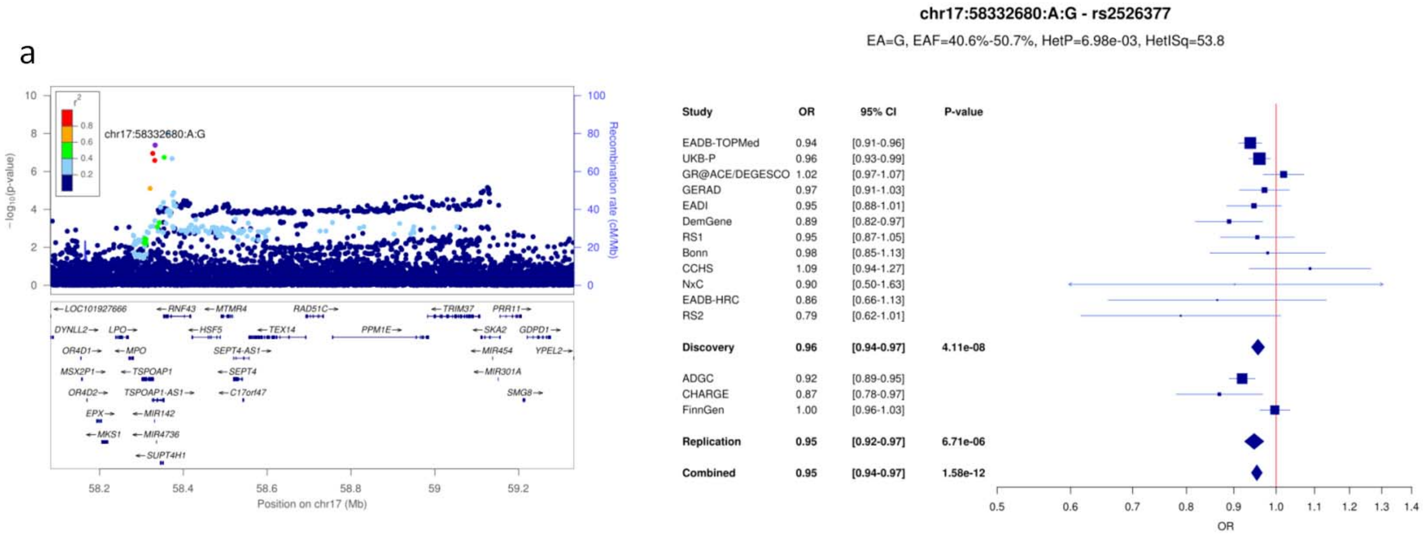


b

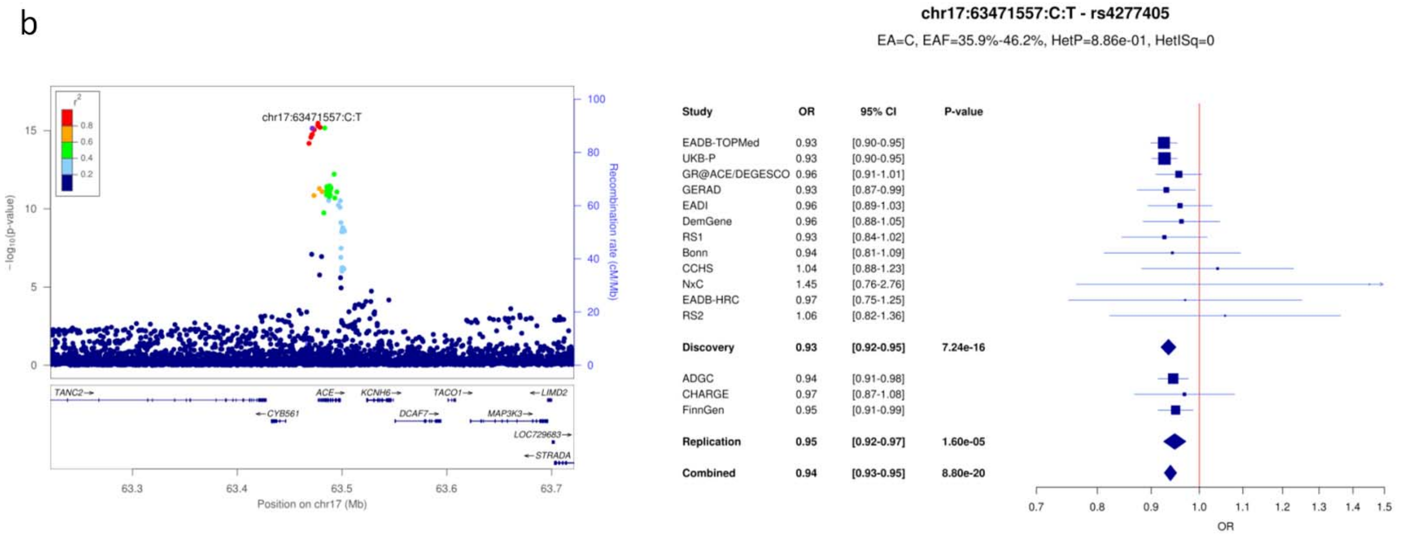


c

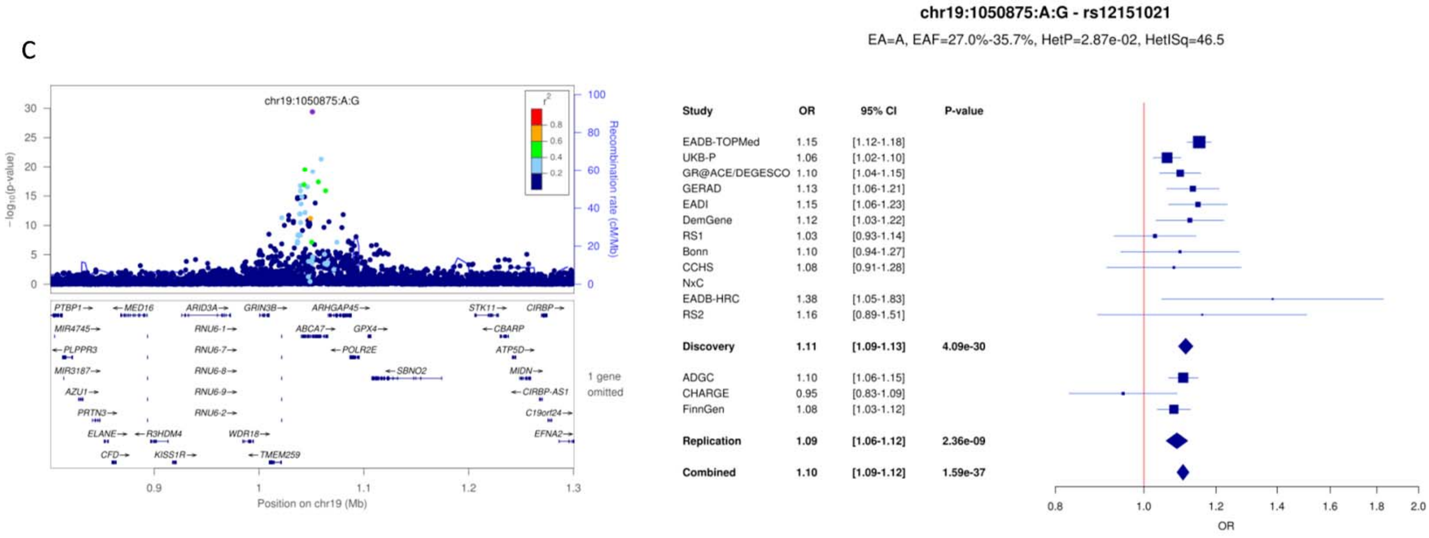

**Supplementary Figure 27**: LocusZoom and forest plots for (a) KLF16, (b) SIGLEC11 and (c) LILRB2 loci.

a

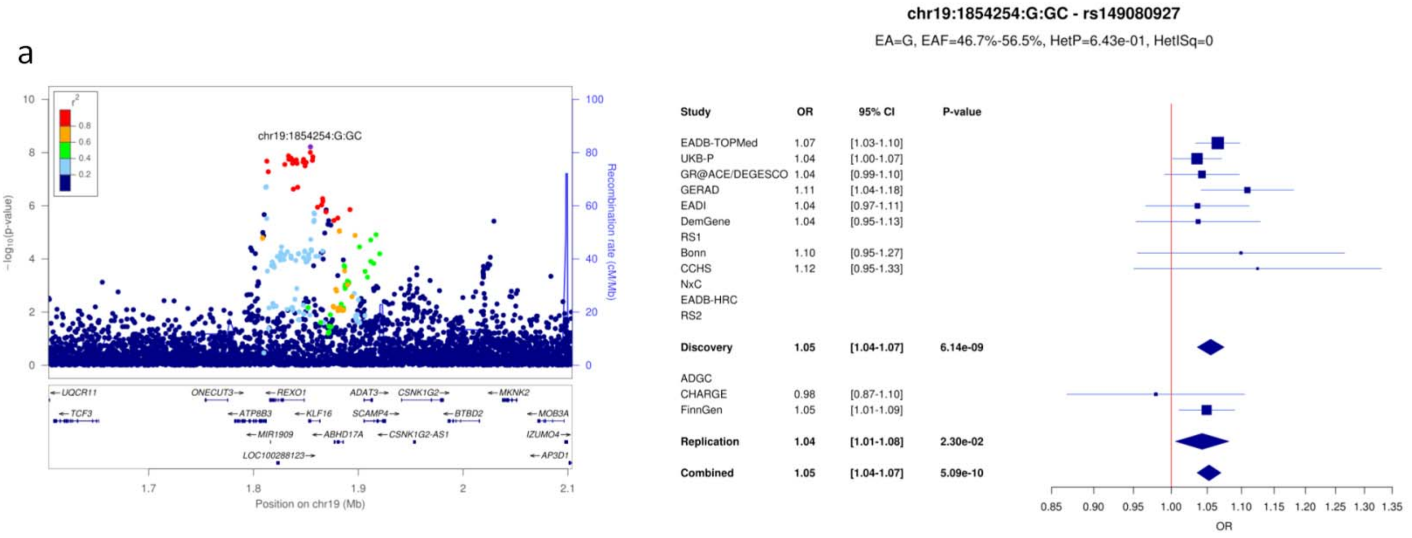


b

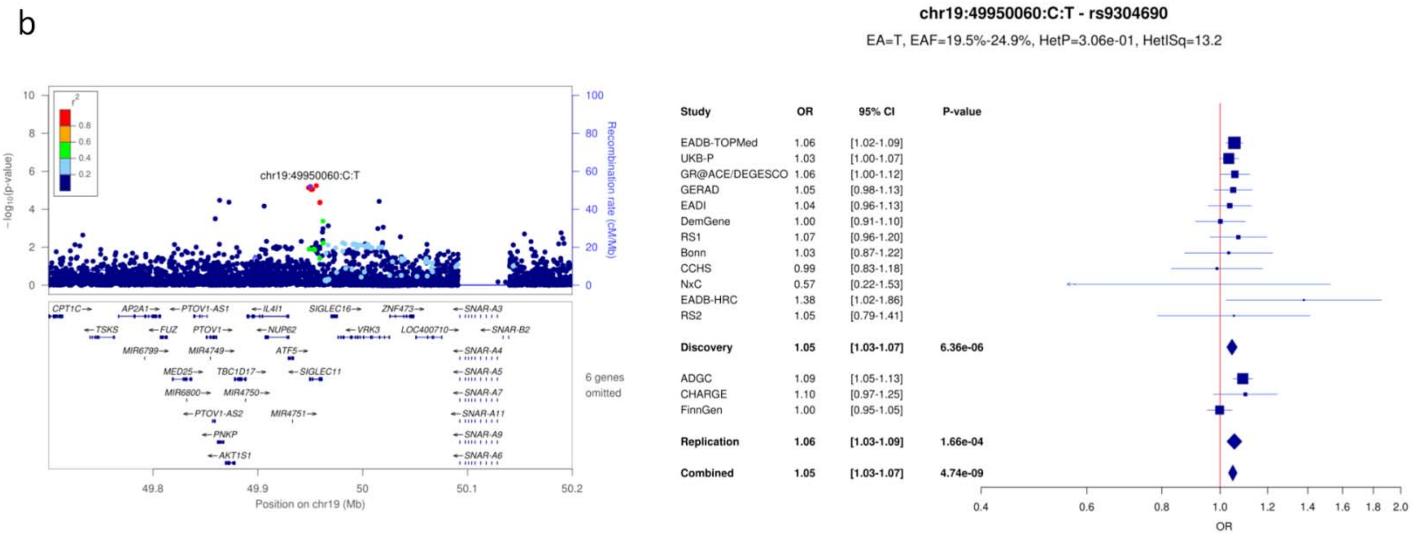


c

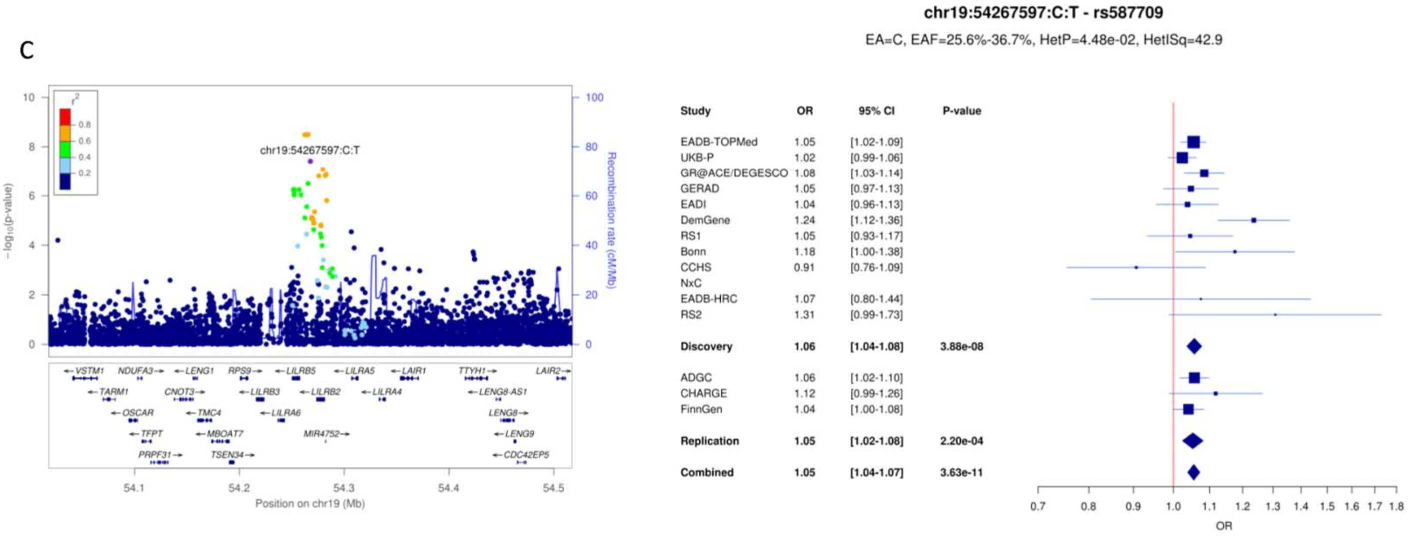

**Supplementary Figure 28**: LocusZoom and forest plots for (a) RBCK1, (b) CASS4 and (c) SLC2A4RG loci.

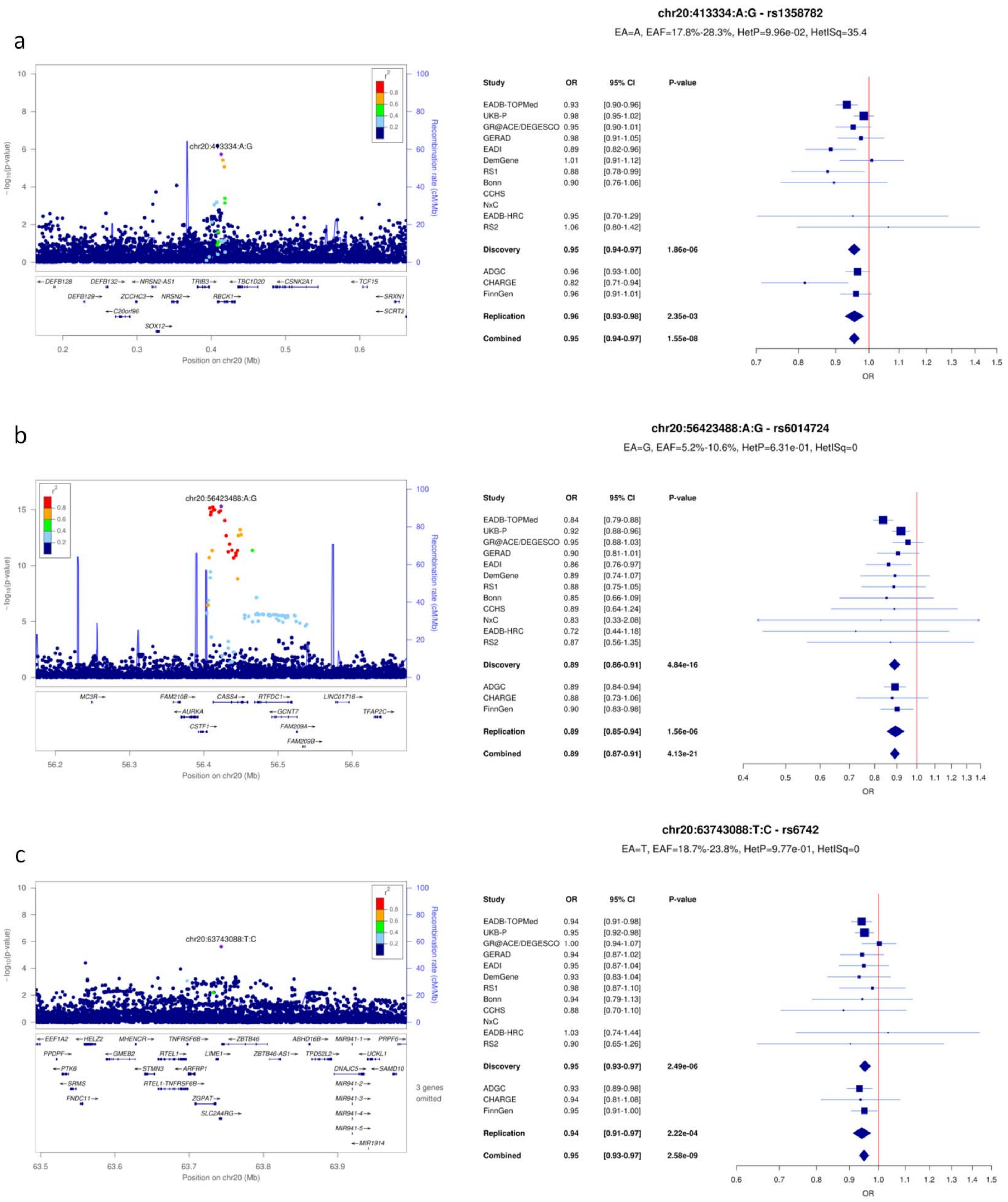

**Supplementary Figure 29**: LocusZoom and forest plots for (a) APP and (b) ADAMTS1 loci.

a

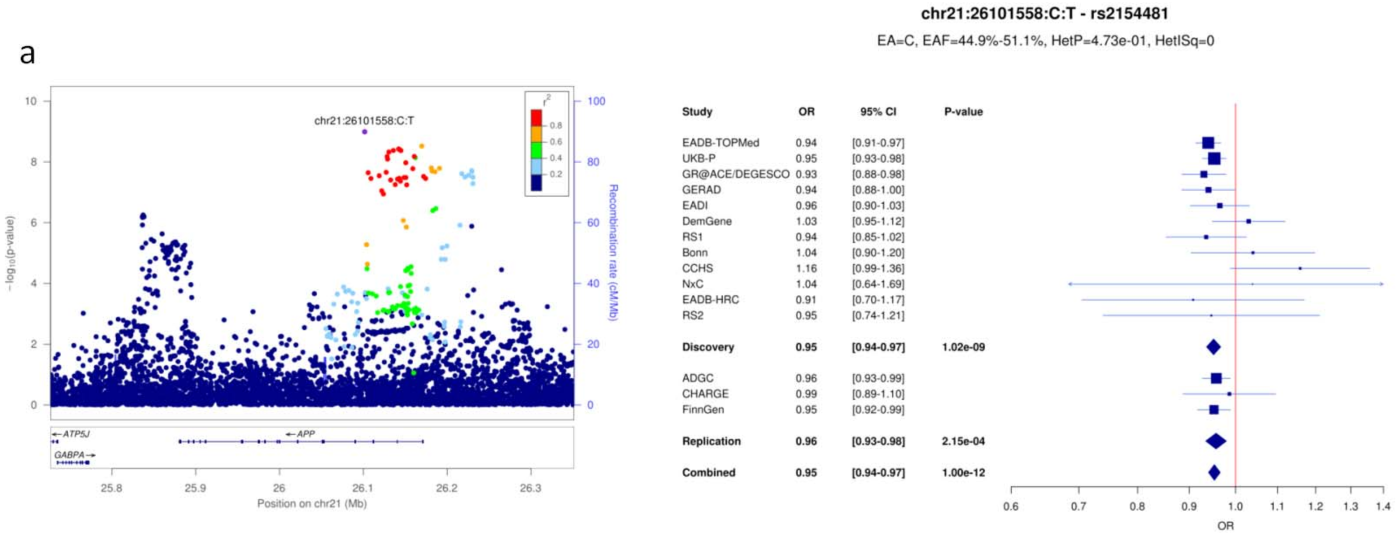


b

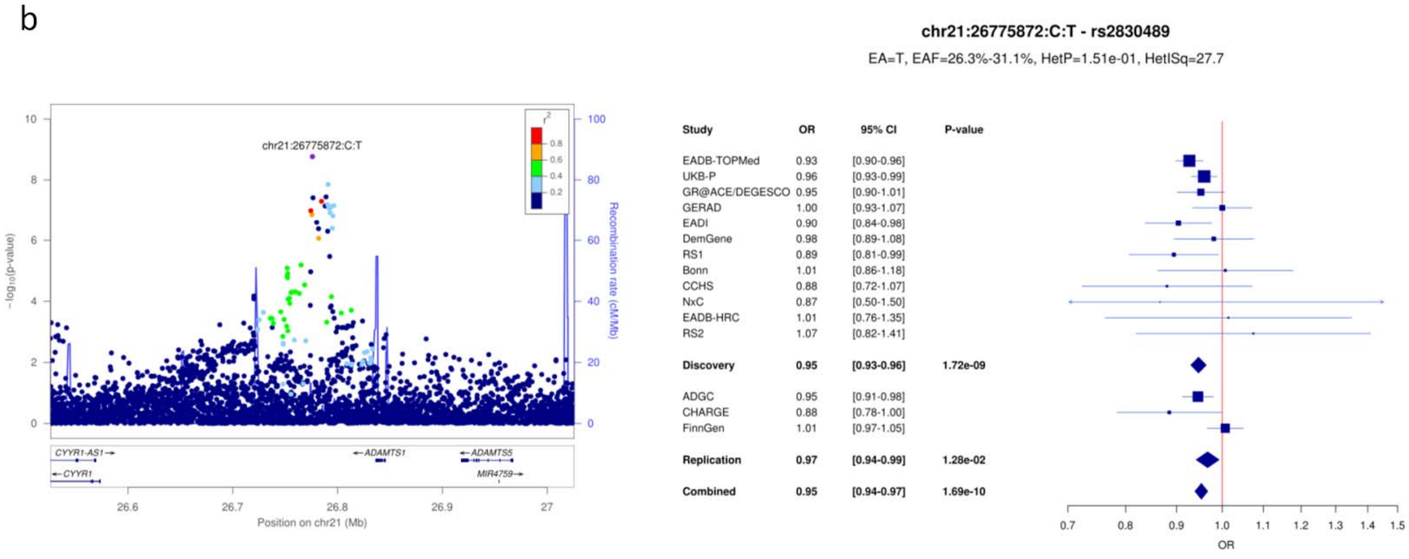

**Supplementary Figure 30.** Comparison of ORs of the genome-wide significant loci a) observed in the diagnosed cases only analysis and estimated in the UK-Biobank only including ADD-proxy cases (UKB-P). b) observed in the diagnosed cases only analysis and estimated in Stage I including ADD-proxy cases. OR: odds-ratio

a

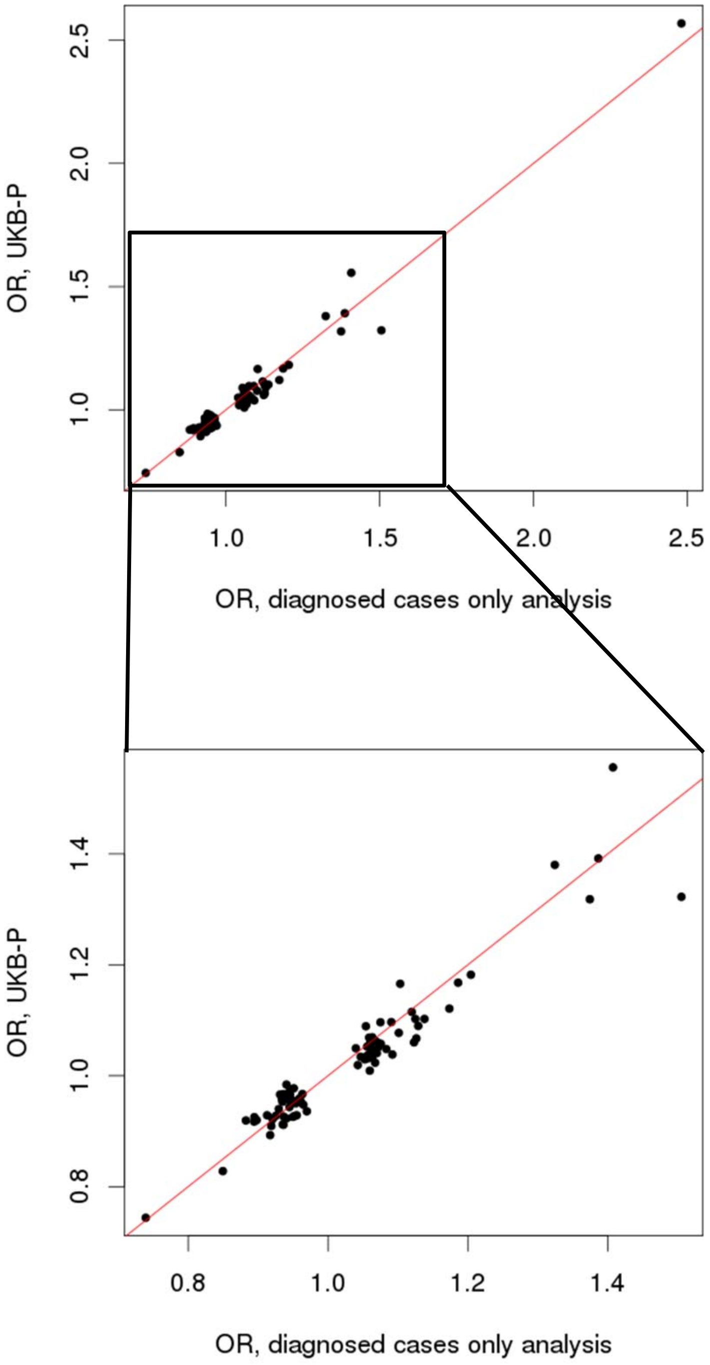

**Supplementary Figure 30 continued**

b

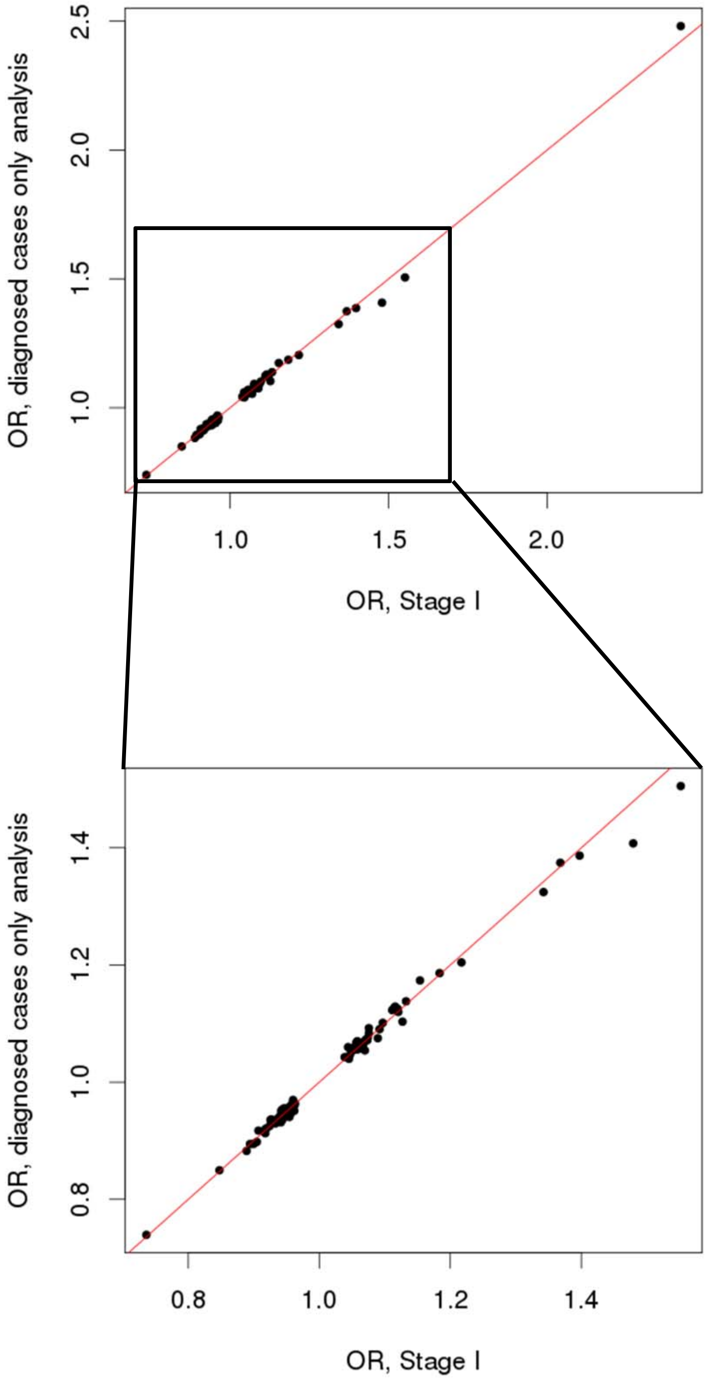

**Supplementary Figure 31.** Forest plots of the 8 HLA alleles associated with ADD (FDR P below 0.05).

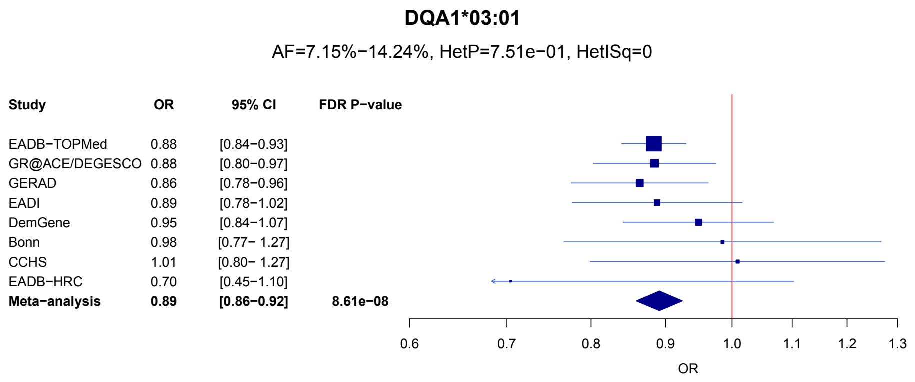


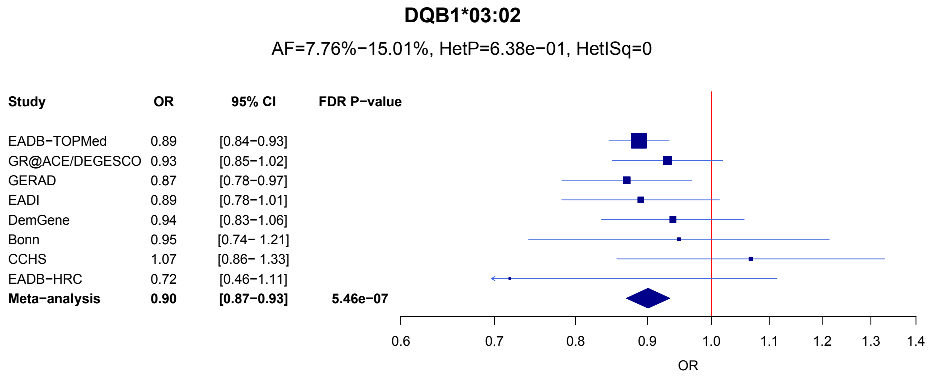


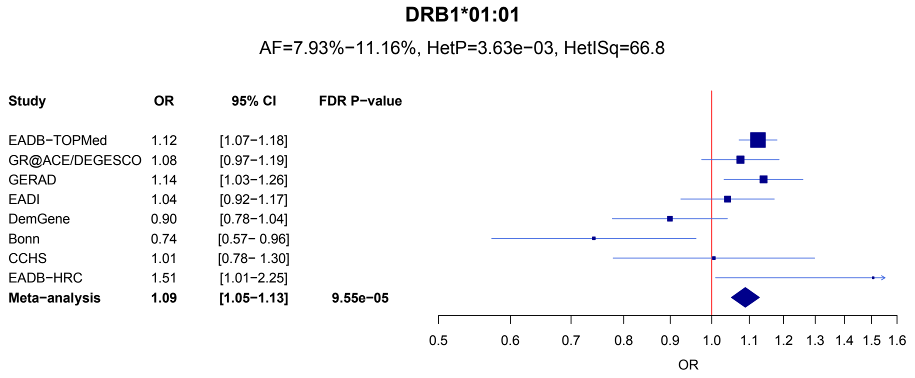


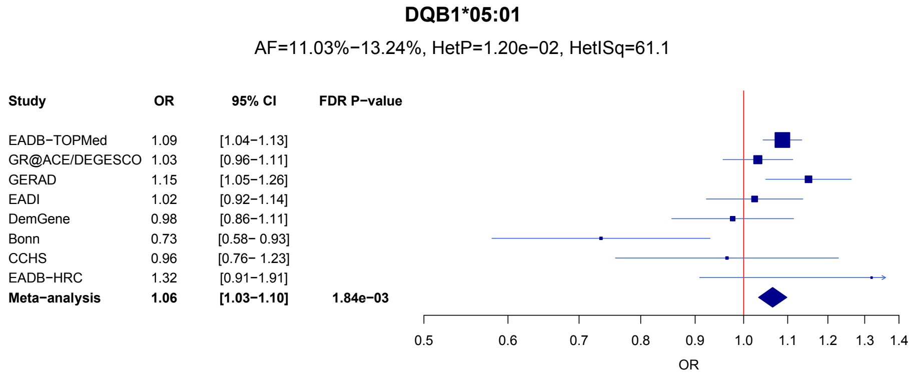

#### Supplementary Figure 31 continued

##### DRB1*04:04

AF=1.55%−5.86%, HetP=1.07e−01, HetISq=40.7

| Study | OR | 95% CI | FDR P−value |
|---|---|---|---|
| EADB−TOPMed | 0.85 | [0.77−0.94] | |
| GR@ACE/DEGESCO | 0.94 | [0.80−1.11] | |
| GERAD | 0.76 | [0.64−0.90] | |
| EADI | 0.94 | [0.75−1.18] | |
| DemGene | 0.94 | [0.79−1.13] | |
| Bonn | 0.70 | [0.41− 1.21] | |
| CCHS | 1.30 | [0.91− 1.86] | |
| EADB−HRC | 0.34 | [0.08−1.49] | |
| **Meta−analysis** | **0.88** | **[0.82−0.94]** | **2.77e−03** |

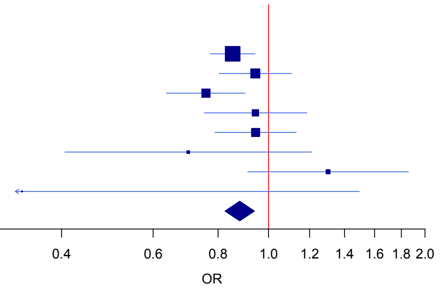


##### DQA1*01:01

AF=10.29%−11.97%, HetP=2.20e−03, HetISq=68.7

| Study | OR | 95% CI | FDR P−value |
|---|---|---|---|
| EADB−TOPMed | 1.10 | [1.05−1.15] | |
| GR@ACE/DEGESCO | 1.01 | [0.94−1.10] | |
| GERAD | 1.13 | [1.03−1.25] | |
| EADI | 1.01 | [0.90−1.12] | |
| DemGene | 0.94 | [0.82−1.08] | |
| Bonn | 0.71 | [0.55− 0.90] | |
| CCHS | 0.96 | [0.74− 1.23] | |
| EADB−HRC | 1.42 | [0.97−2.08] | |
| **Meta−analysis** | **1.06** | **[1.03−1.10]** | **4.96e−03** |

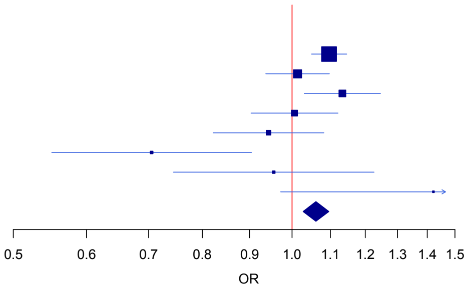


### B*57:01

AF=2.5%−4.1%, HetP=2.00e−01, HetISq=28.5

| Study | OR | 95% CI | FDR P−value |
|---|---|---|---|
| EADB−TOPMed | 1.10 | [1.01−1.20] | |
| GR@ACE/DEGESCO | 1.11 | [0.93−1.31] | |
| GERAD | 0.95 | [0.81−1.12] | |
| EADI | 1.15 | [0.93−1.41] | |
| DemGene | 1.26 | [1.01−1.59] | |
| Bonn | 1.44 | [0.97− 2.15] | |
| CCHS | 1.41 | [0.98− 2.02] | |
| EADB−HRC | 1.71 | [0.85−3.46] | |
| **Meta−analysis** | **1.11** | **[1.04−1.18]** | **1.21e−02** |

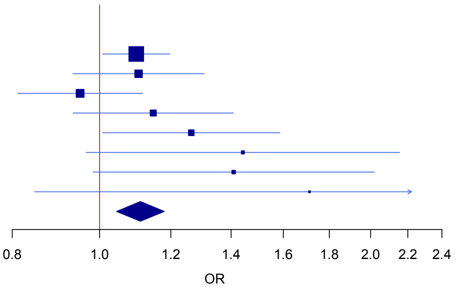


### A*02:01

AF=24.22%−33.45%, HetP=2.68e−01, HetISq=20.3

| Study | OR | 95% CI | FDR P−value |
|---|---|---|---|
| EADB−TOPMed | 1.04 | [1.00−1.07] | |
| GR@ACE/DEGESCO | 1.04 | [0.98−1.10] | |
| GERAD | 1.09 | [1.02−1.17] | |
| EADI | 1.02 | [0.95−1.10] | |
| DemGene | 0.99 | [0.91−1.08] | |
| Bonn | 1.00 | [0.86− 1.18] | |
| CCHS | 1.01 | [0.86− 1.19] | |
| EADB−HRC | 0.72 | [0.53−0.98] | |
| **Meta−analysis** | **1.03** | **[1.01−1.06]** | **4.92e−02** |

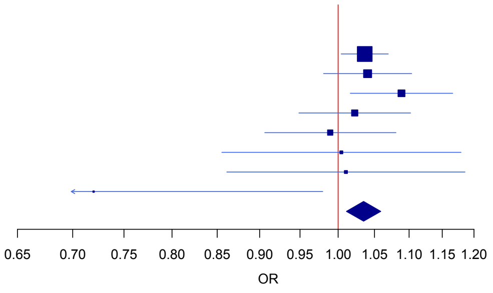

**Supplementary Figure 32.** Forest plots of the 3 three-locus haplotypes associated with ADD (FDR P below 0.05).

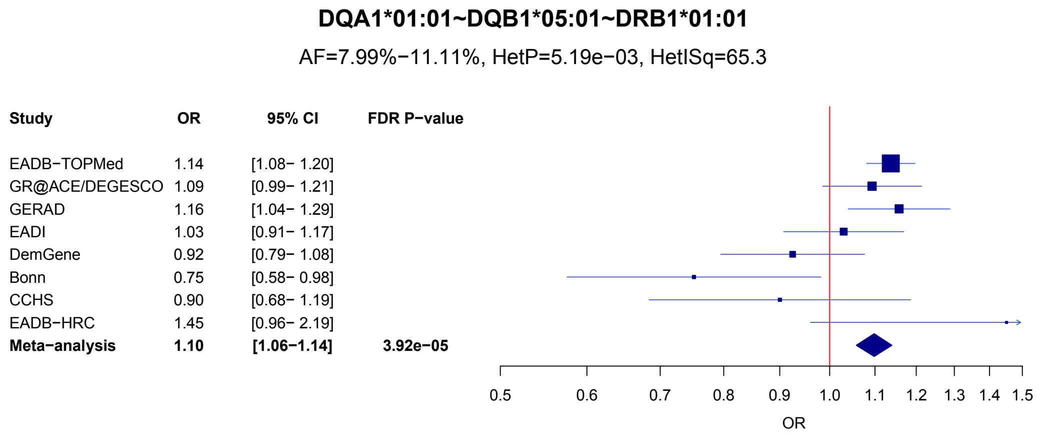


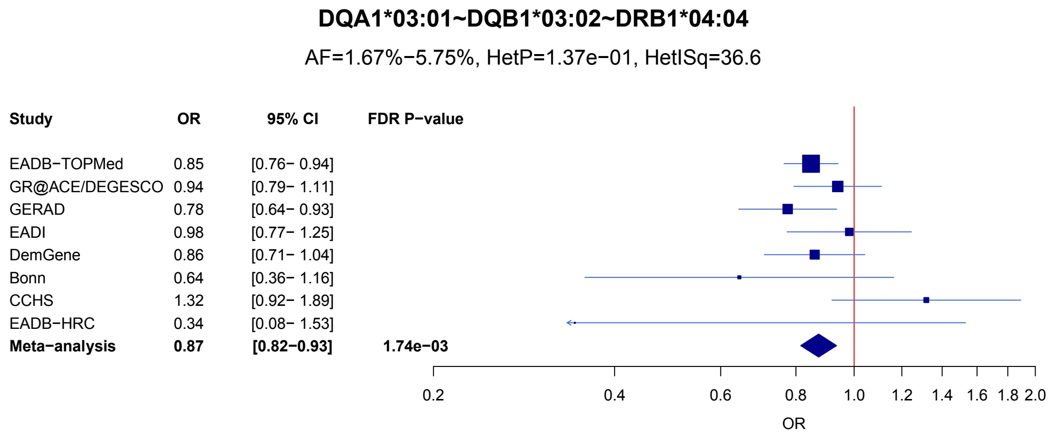


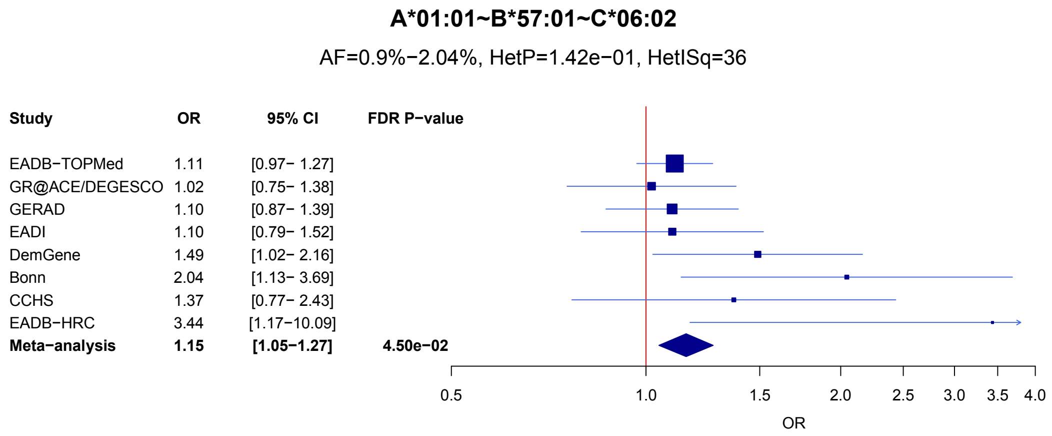

**Supplementary Figure 33.** Z-scores for neurodegenerative and AD related diseases for the risk allele of the ADD associated variants. P values (uncorrected) are denoted with $ (P < 5x10$^{-8}$), X (P < 1x10$^{-5}$), # (P < 1x10$^{-3}$), * (P < 0.05).

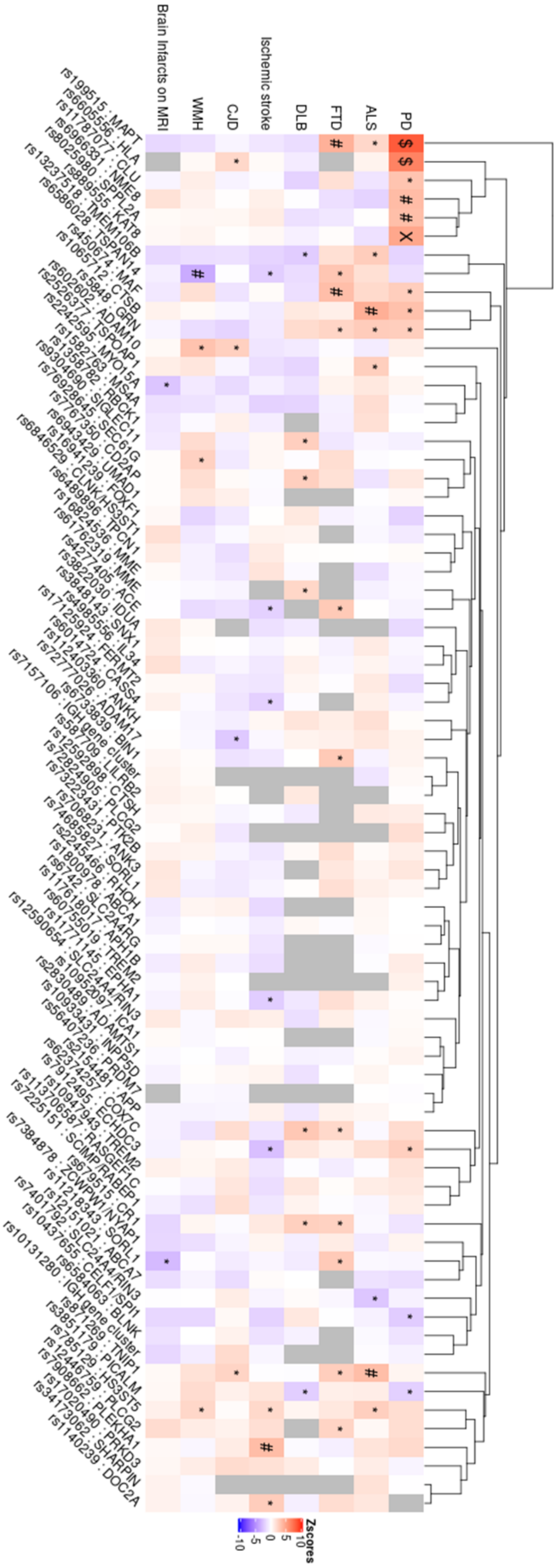

**Supplementary Figure 34**: eQTL effects of lead variants within novel ADD risk loci in (a) AD-relevant brain regions, LCL, microglia, blood, and (b) in naïve state and stimulated macrophages and monocytes.

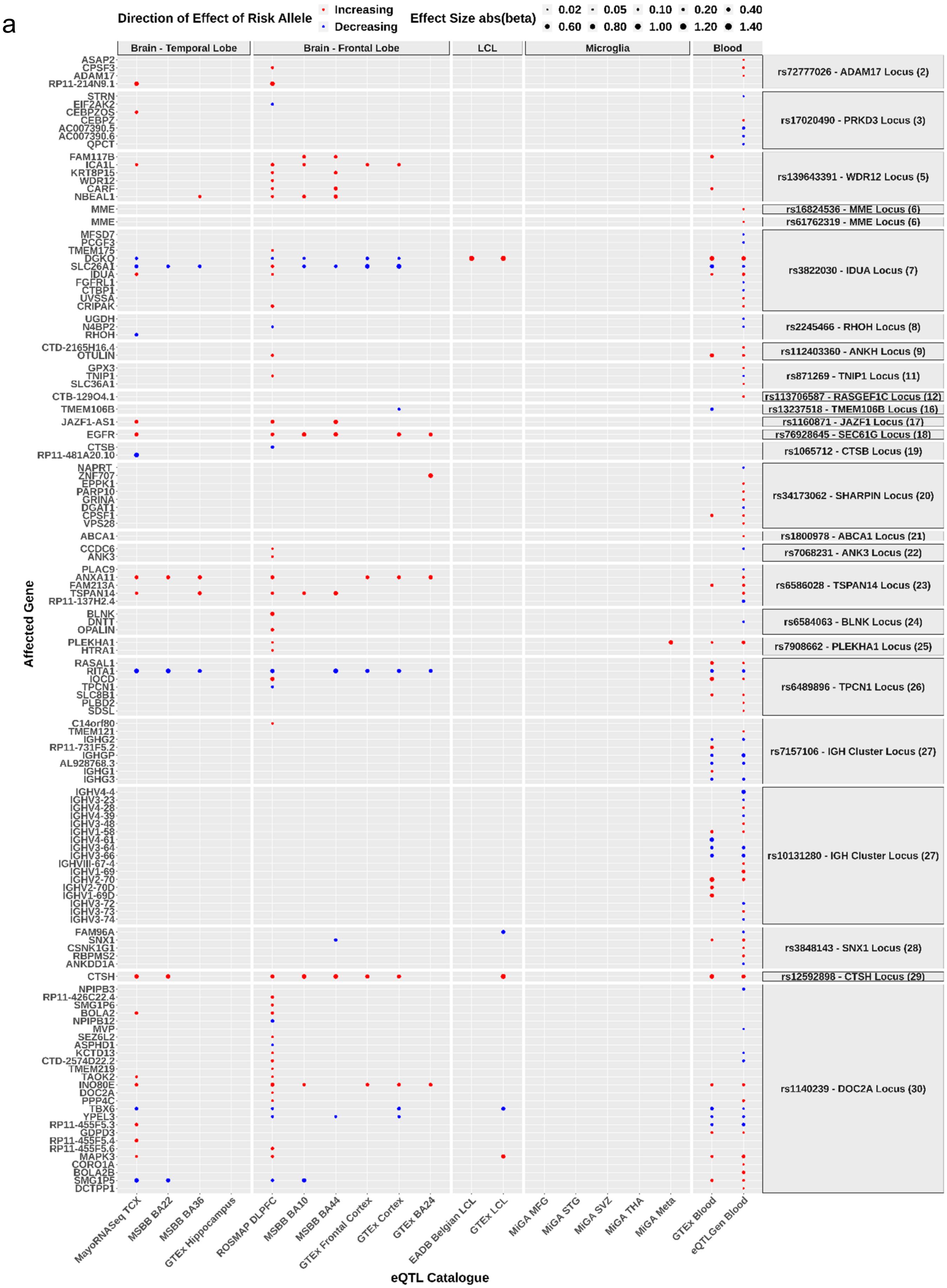

**Supplementary Figure 34 continued**

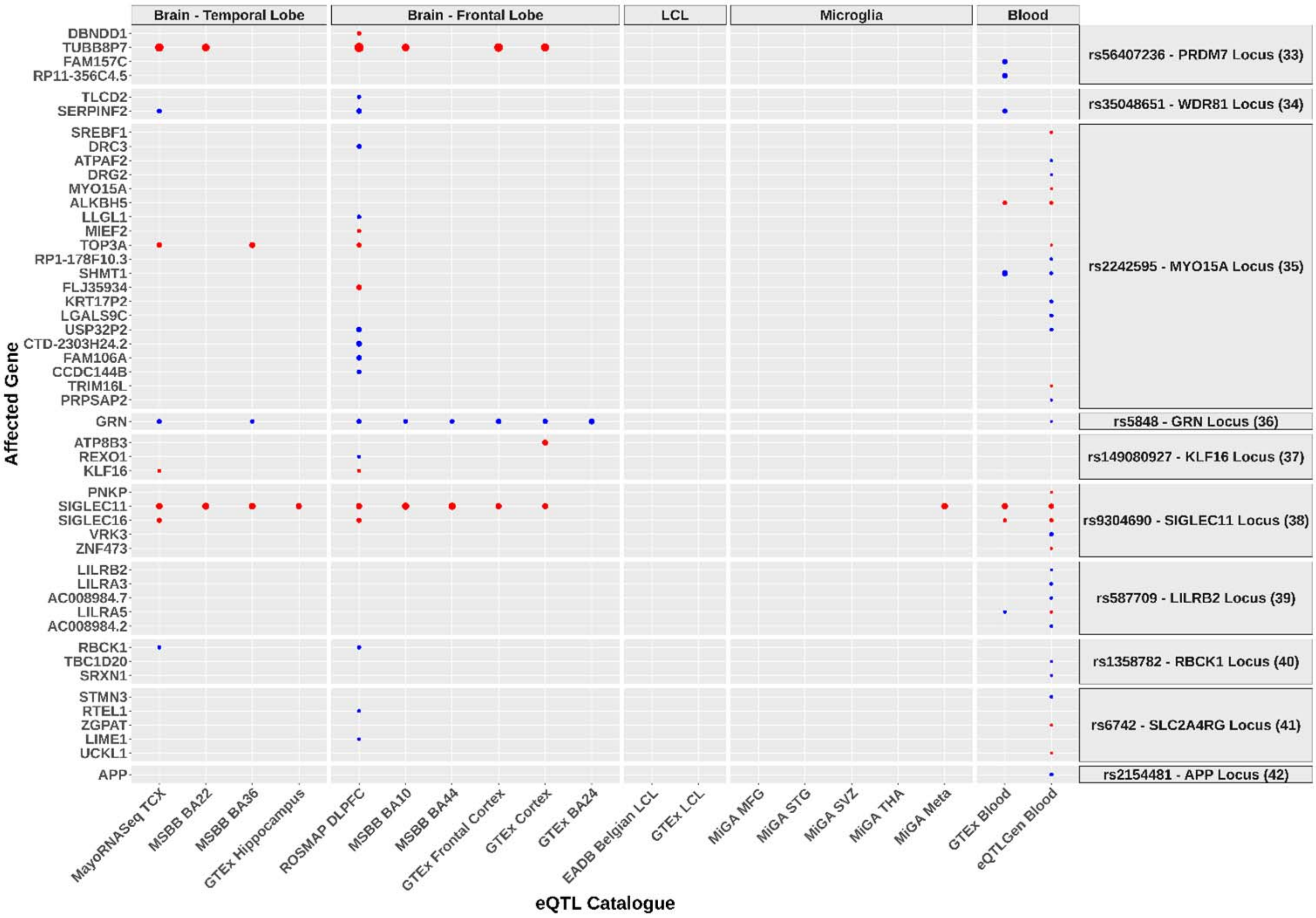


b

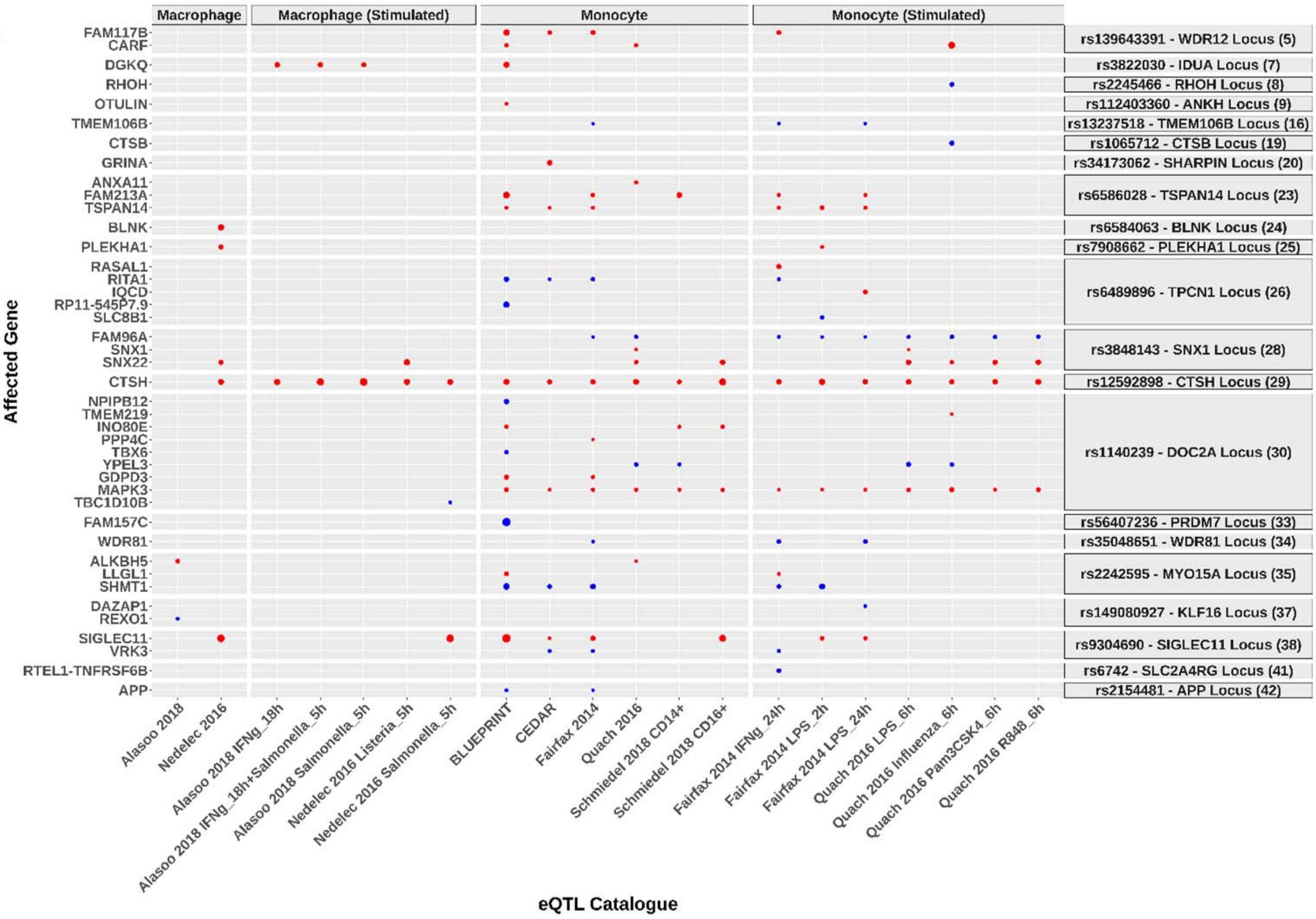

**Supplementary Figure 35**: sQTL effects of lead variants within novel ADD risk loci in AD-relevant brain regions, LCL, microglia, and blood.

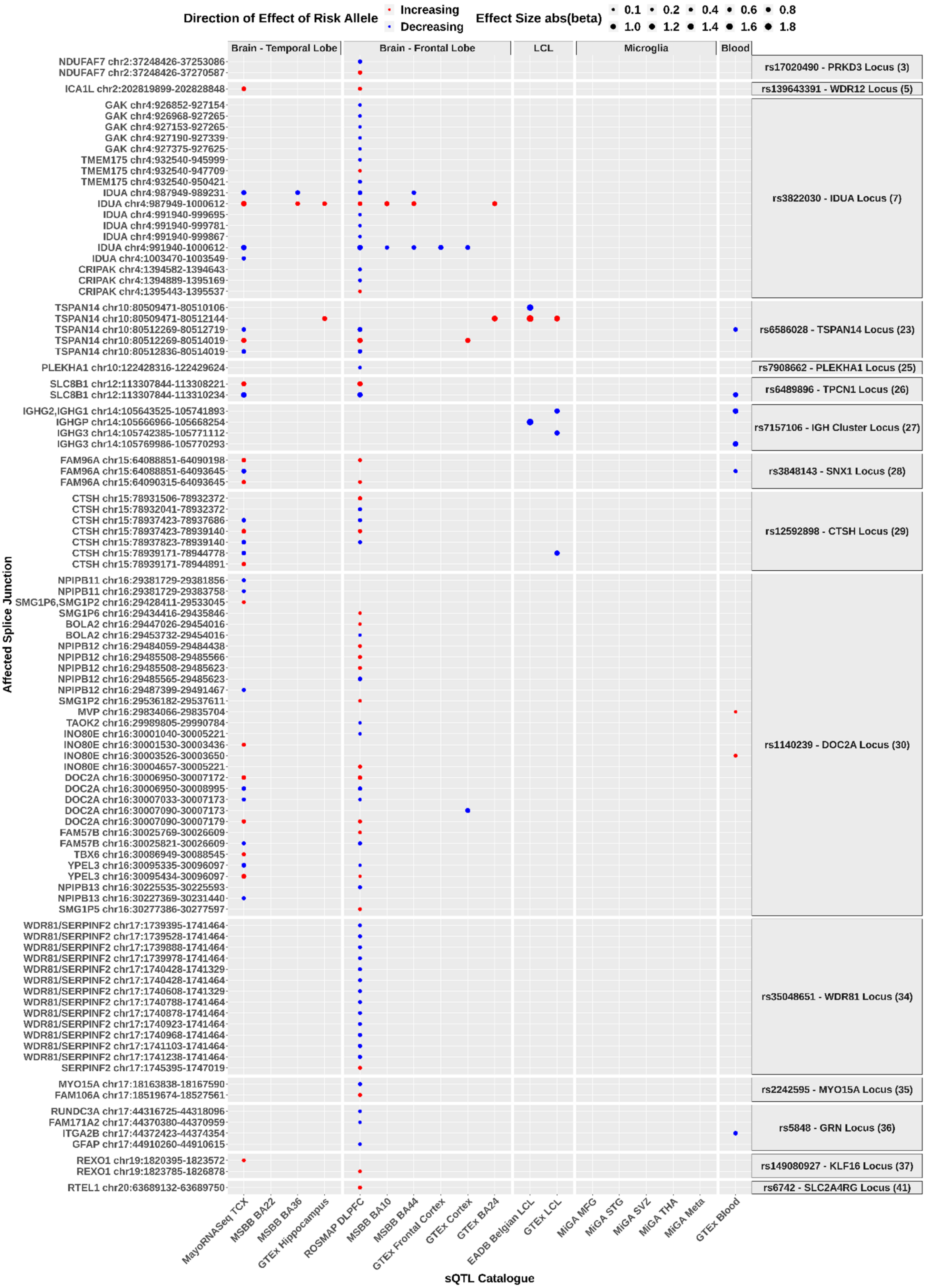

**Supplementary Figure 36**: (a) mQTL and haQTL effects of lead variants within novel ADD risk loci. (b) ADD-associated predicted methylation results using MetaMeth.

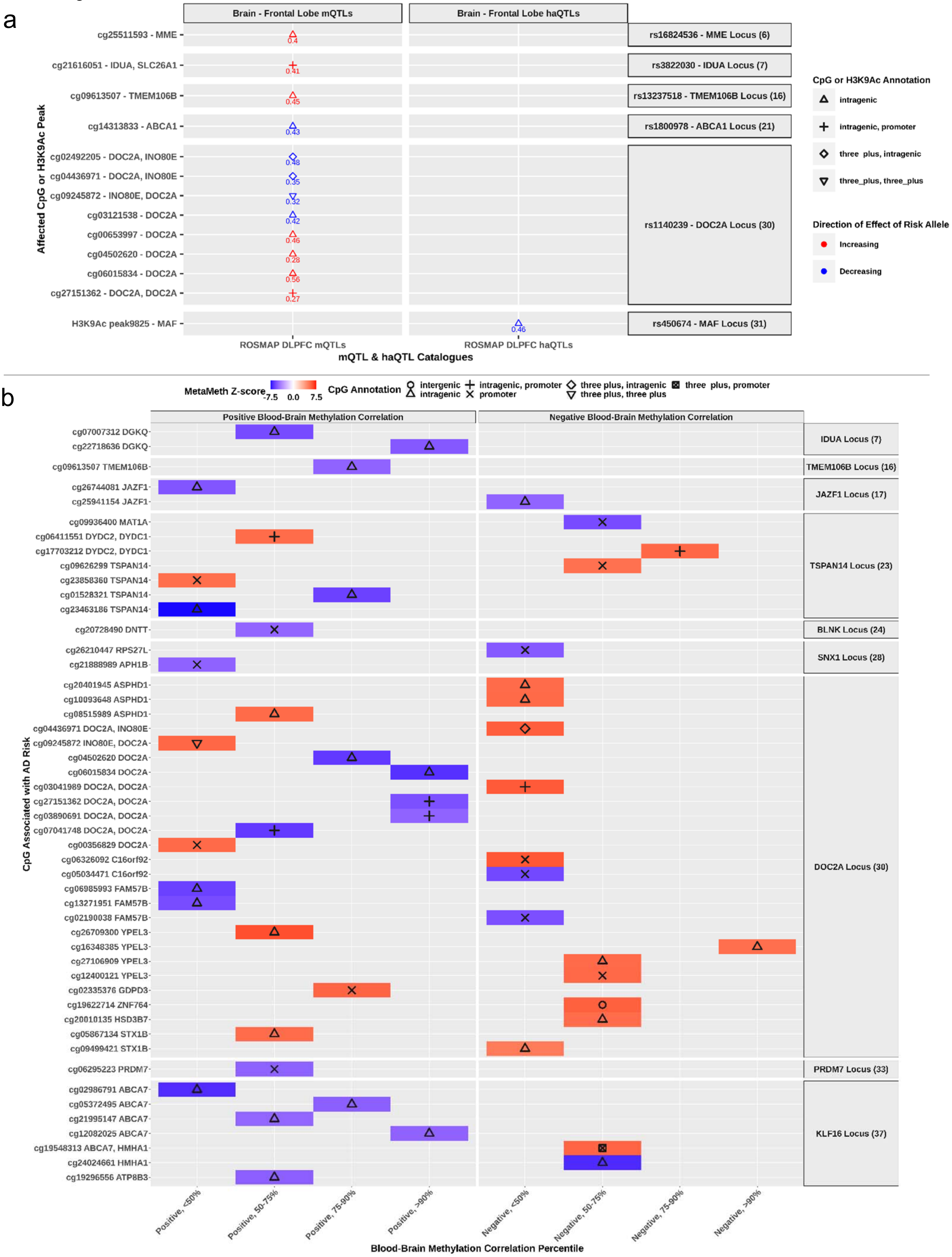

**Supplementary Figure 37**: Colocalization between eQTL signals for genes and ADD association signals.

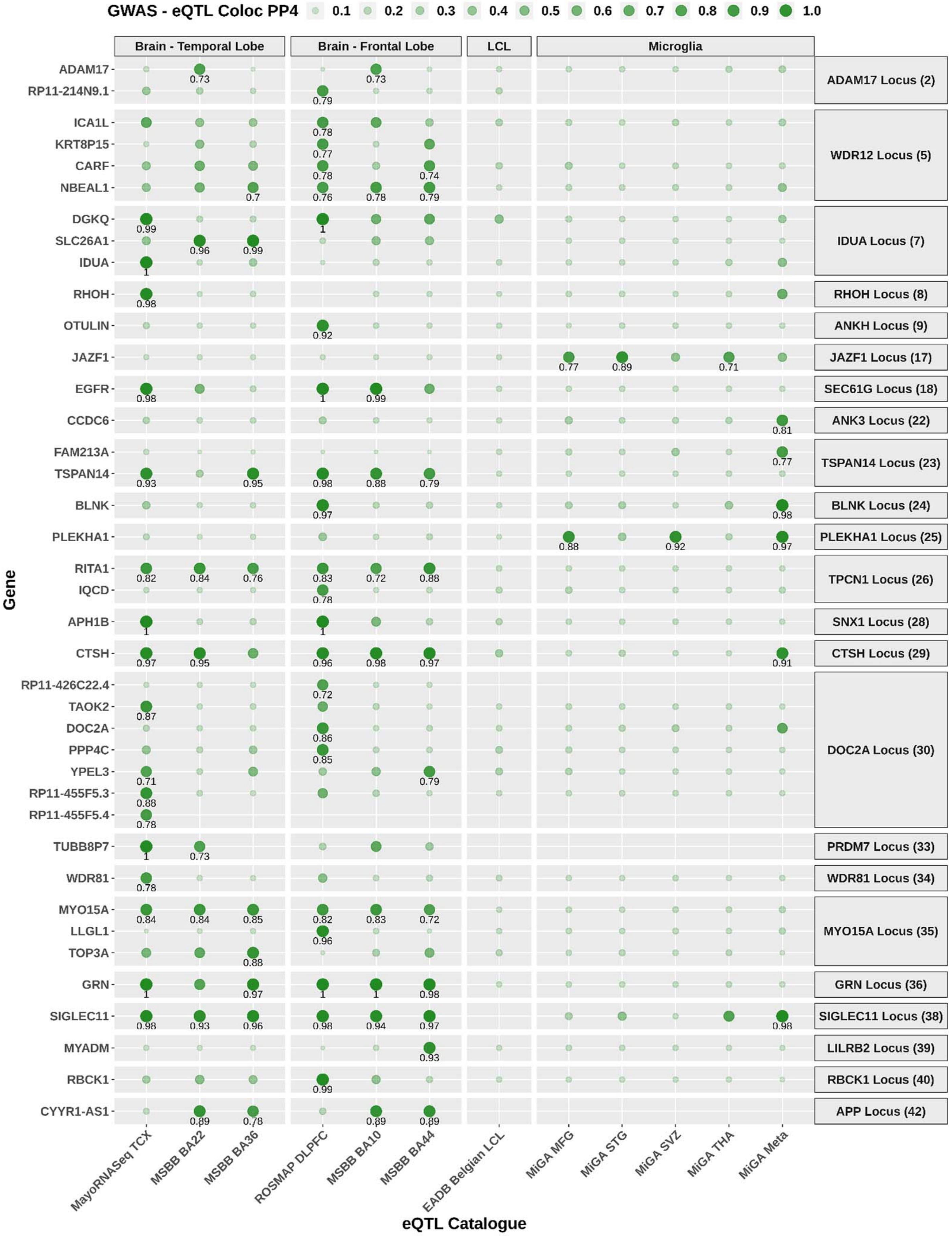

**Supplementary Figure 38**: Colocalization between sQTL signals for splice junctions and ADD association signals.

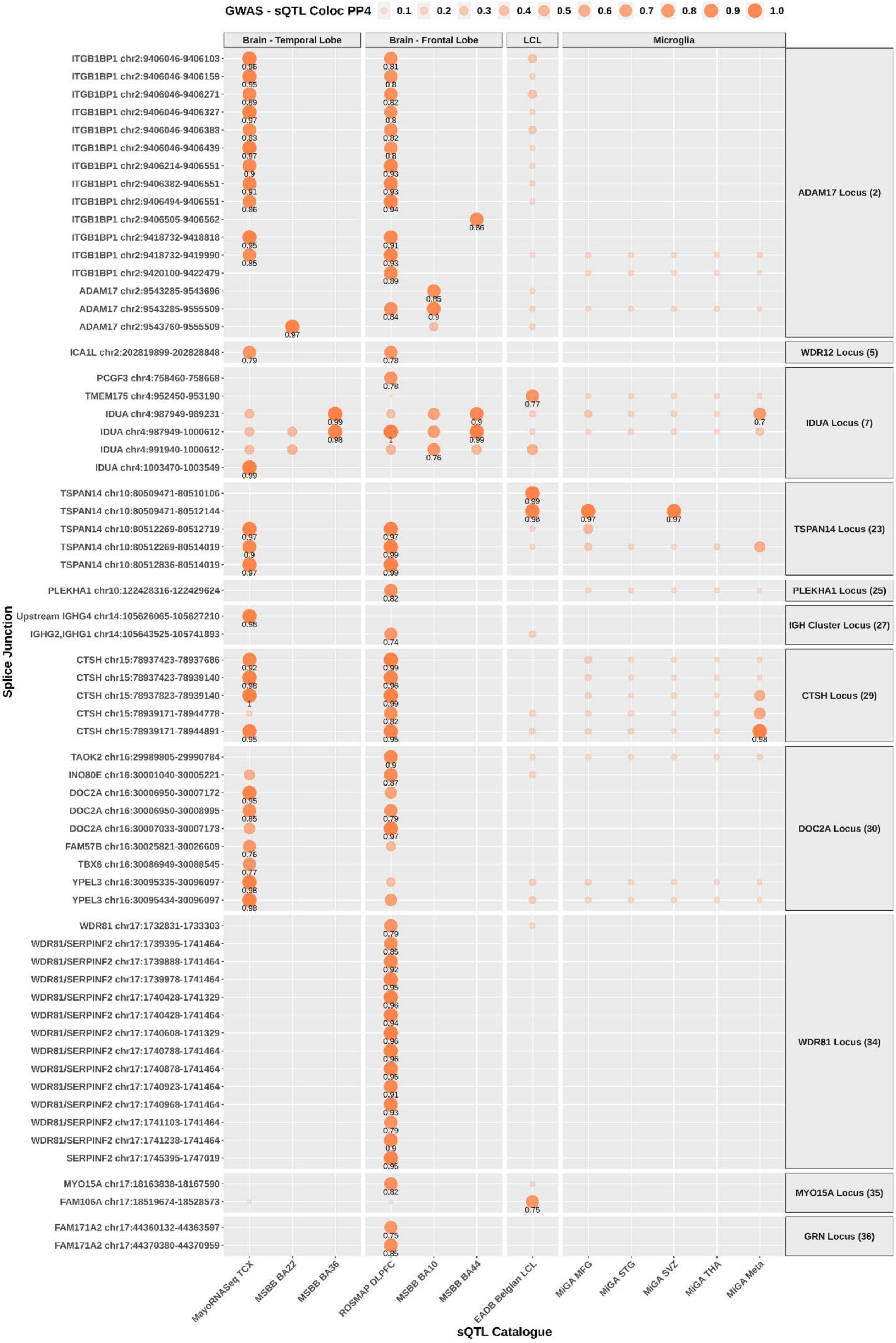

**Supplementary Figure 39**: TWAS of ADD using Expression Reference Panels.

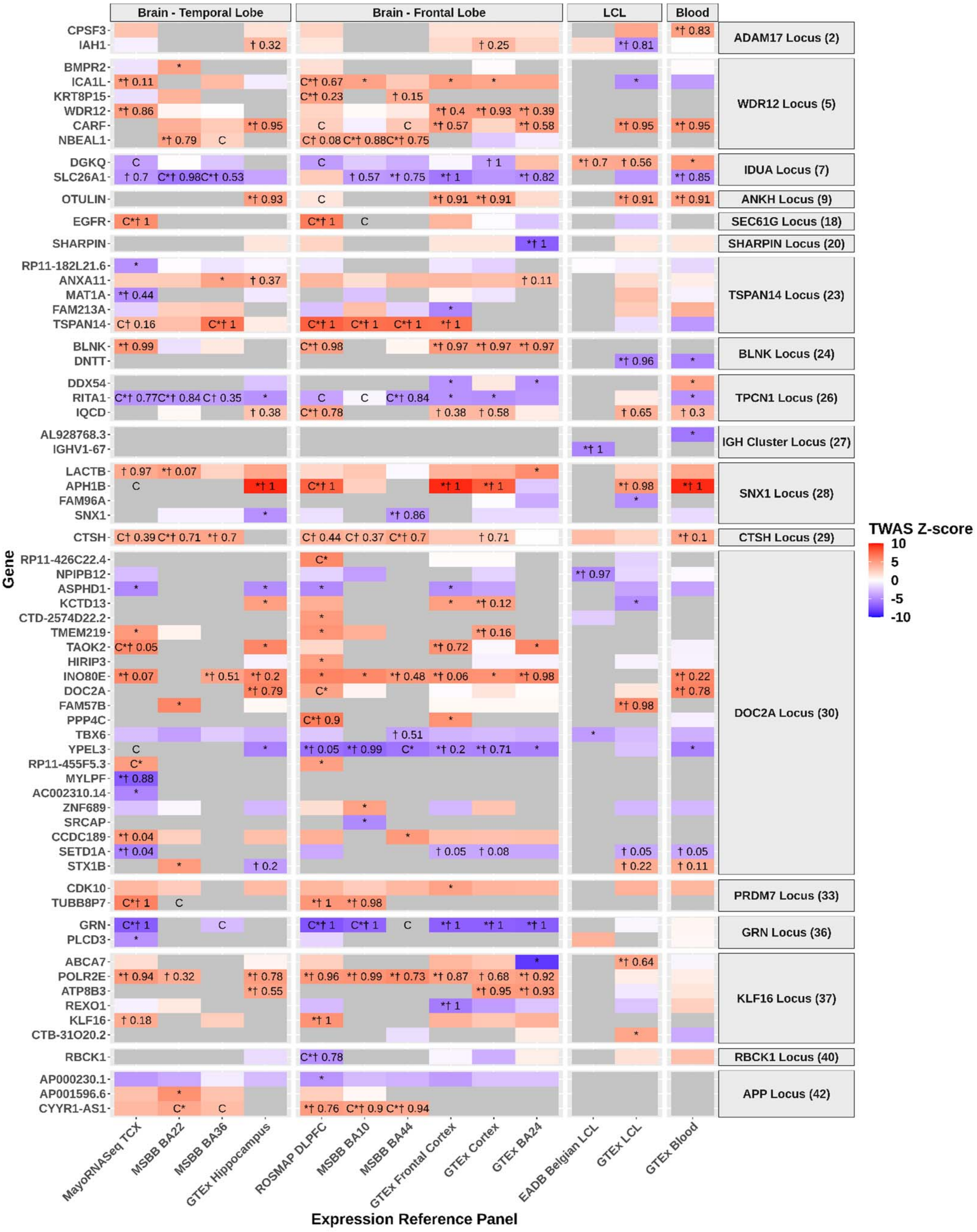

**Supplementary Figure 40**: TWAS of ADD using Splicing Reference Panels.

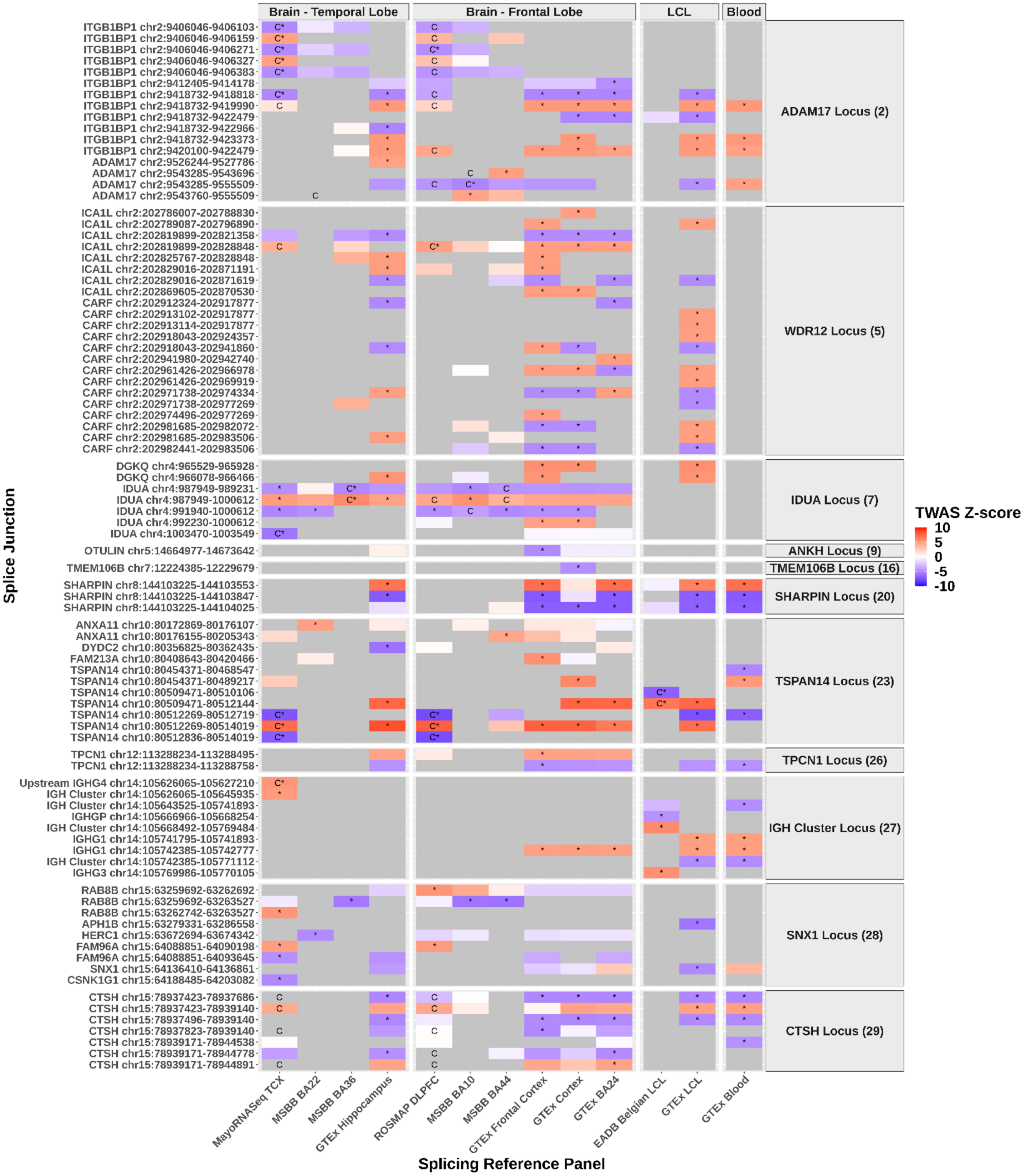

**Supplementary Figure 40 continued**

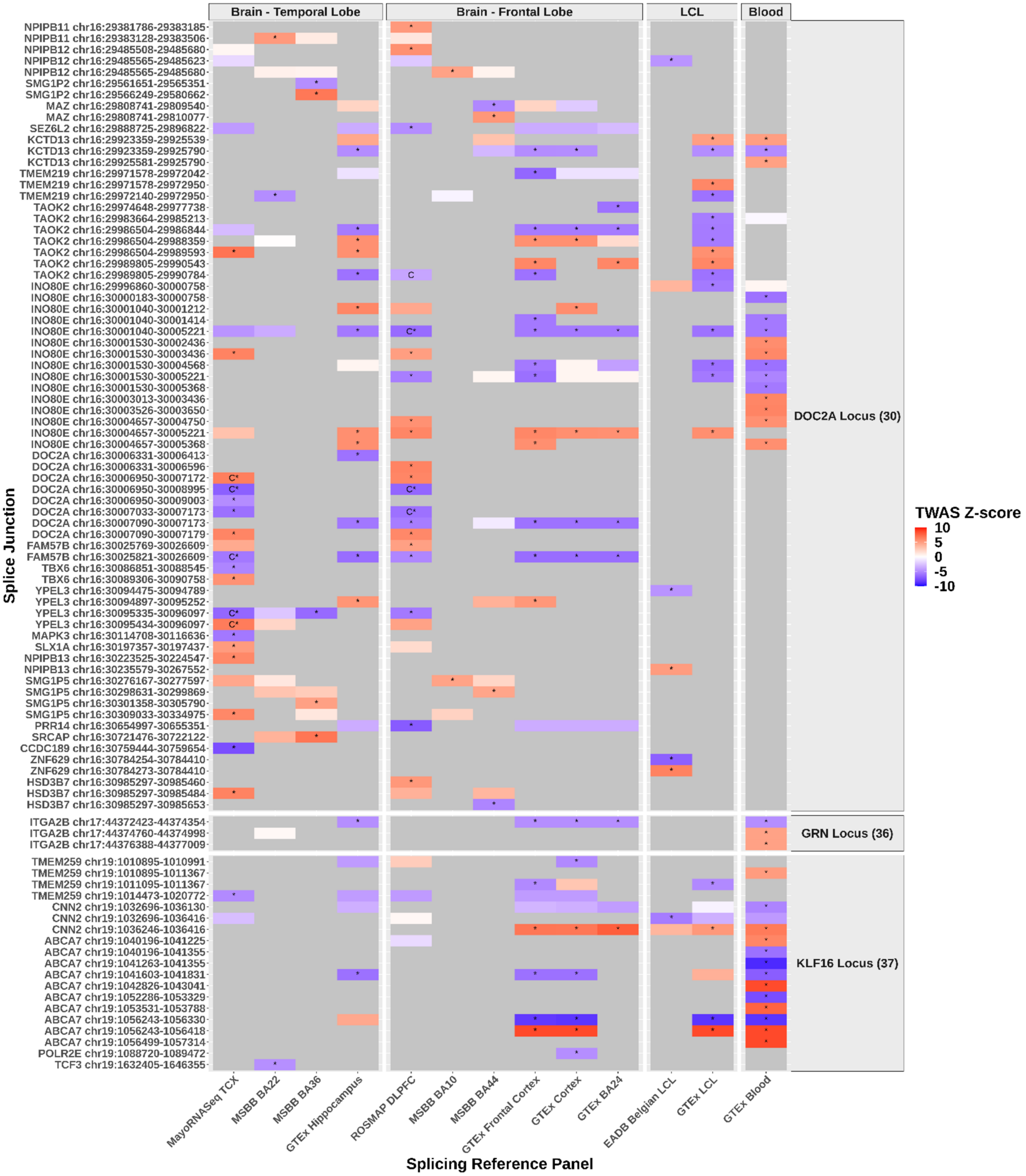

**Supplementary Figure 41**: Fine-mapping of expression TWAS results. (a) *GRN* locus in MayoRNASeq TCX, (b) *KLF16* locus in ROSMAP DLPFC, (c) *TSPAN14* locus in MSBB BA36, and (d) *DOC2A* locus in GTEx hippocampus.

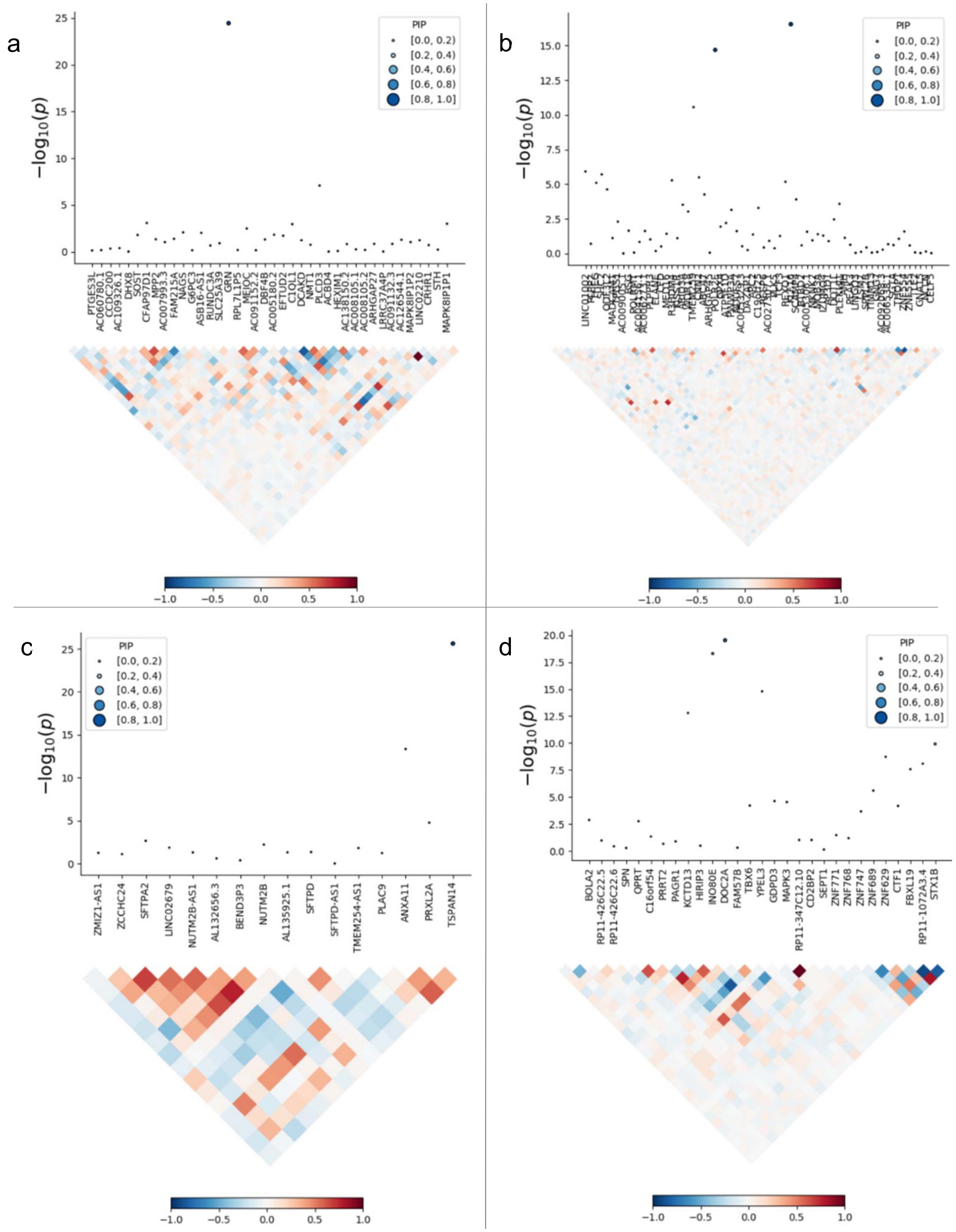

**Supplementary Figure 42.** Mean fluorescence intensity variations (log2 fold-change) of the mCherry signal obtained after the silencing of genes associated with the ADD risk in HEK293 cells stably over-expressing a mCherry-APP695WT-YFP in the 42 new loci.

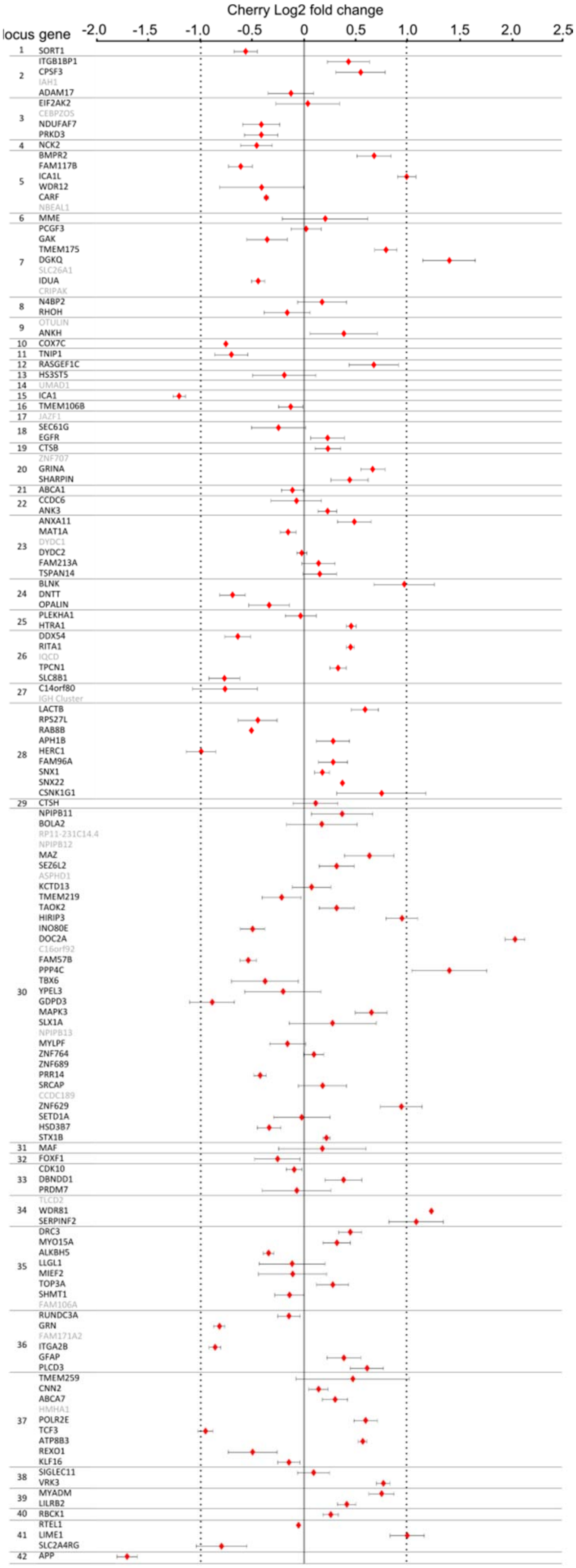

**Supplementary Figure 43.** STRING protein interaction analysis. The main networks are shown in a) previous genes, b) prioritized new genes and c) combined datasets.

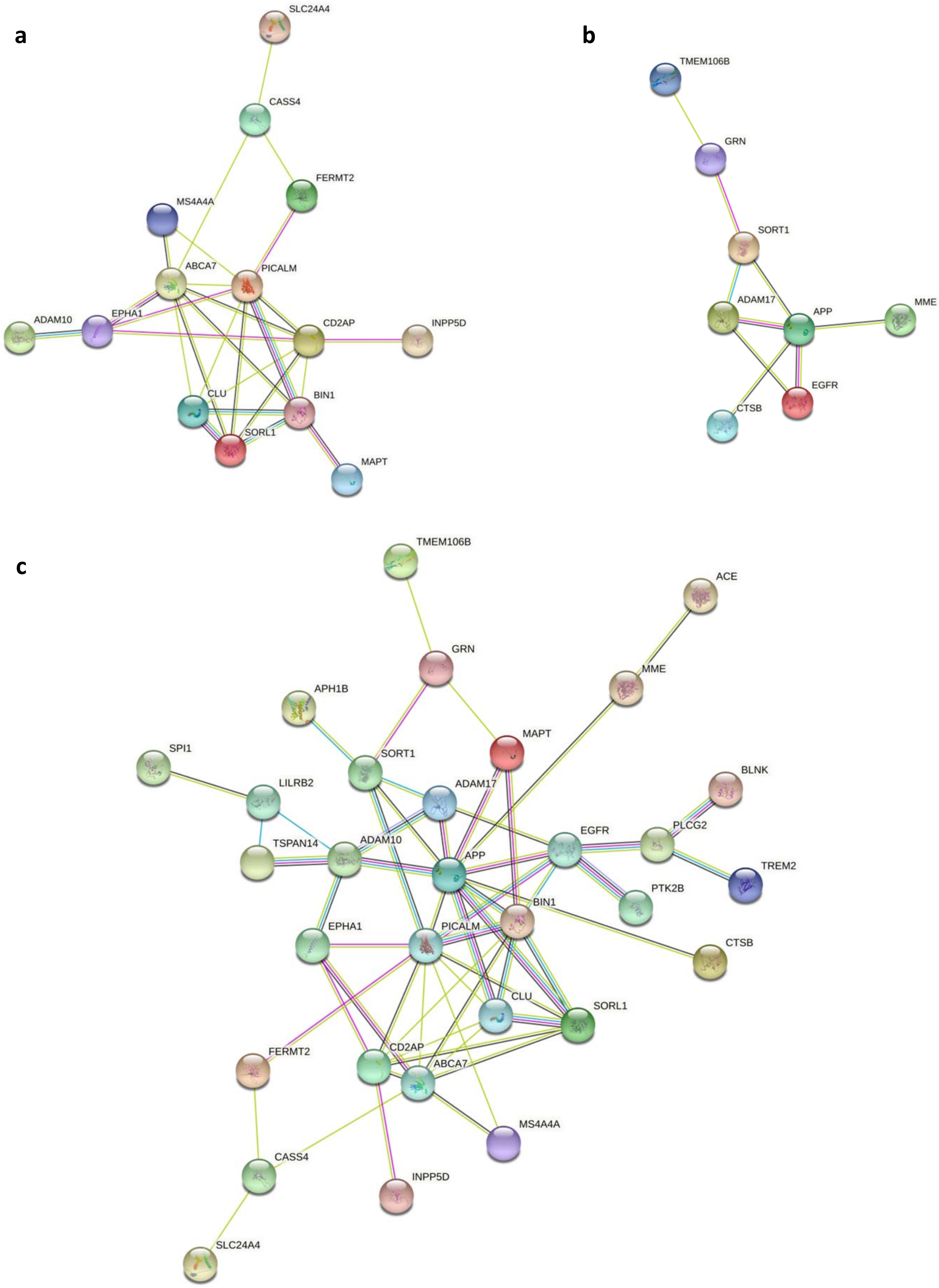

**Supplementary Figure 44.** Association of PRS with the risk of progression to dementia starting from either (a) normal cognition or (b) mild cognitive impairment (MCI) in non e4-bearers. PRS was based on the genetic data of 83 variants (see Methods and Supplementary Table 31).

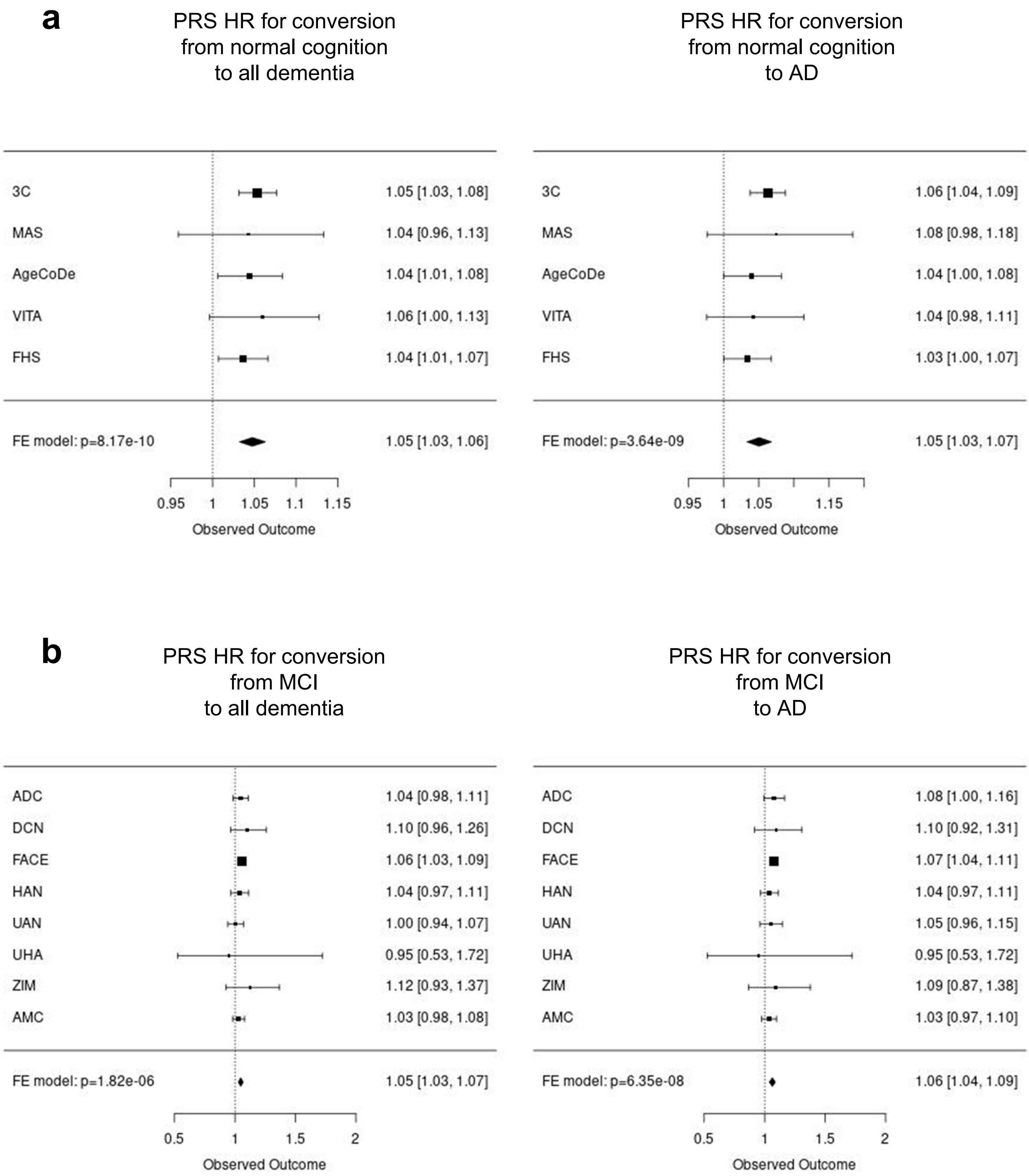

**Supplementary Figure 45.** Association of PRS with the risk of progression to dementia starting from either (a) normal cognition or (b) mild cognitive impairment (MCI) in e4-bearers. PRS was based on the genetic data of 83 variants (see Methods and Supplementary Table 31).

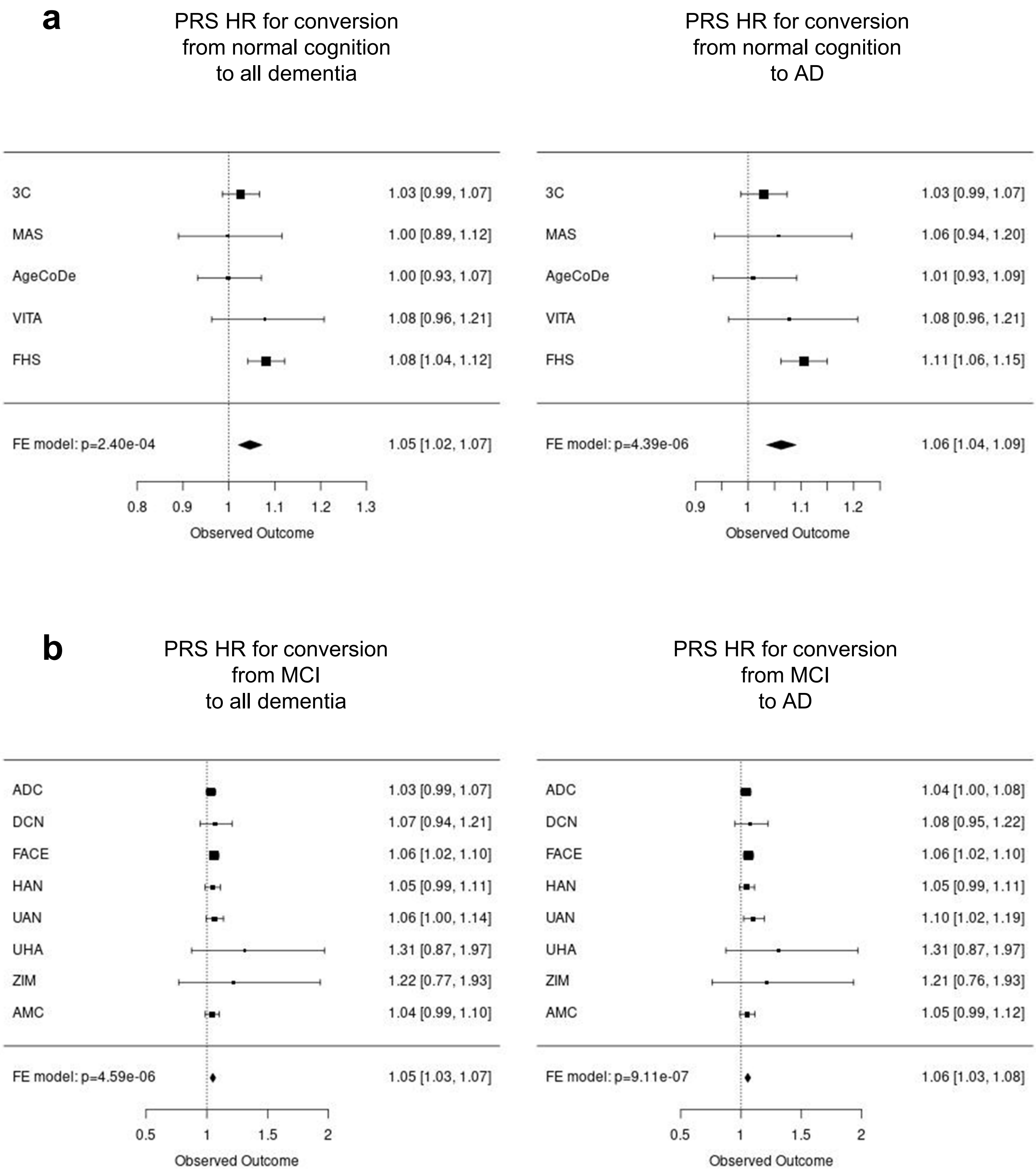

**Supplementary Figure 46.** EADB sample quality control

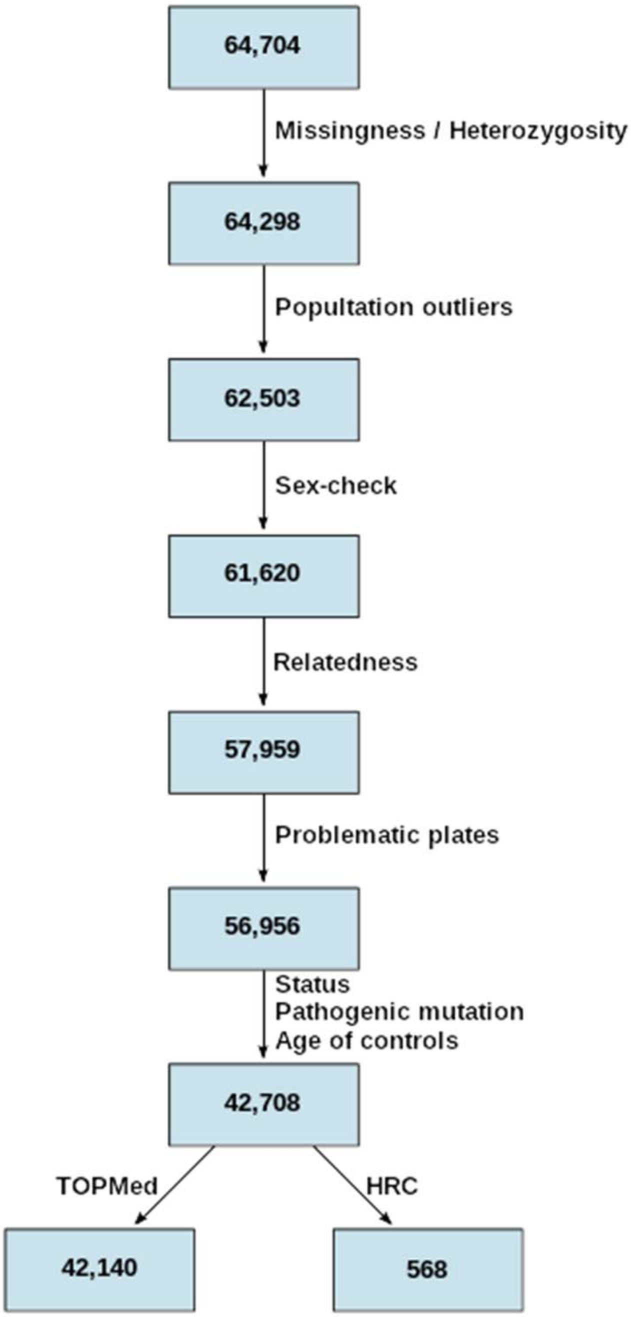

**Supplementary Figure 47.** EADB variant quality control

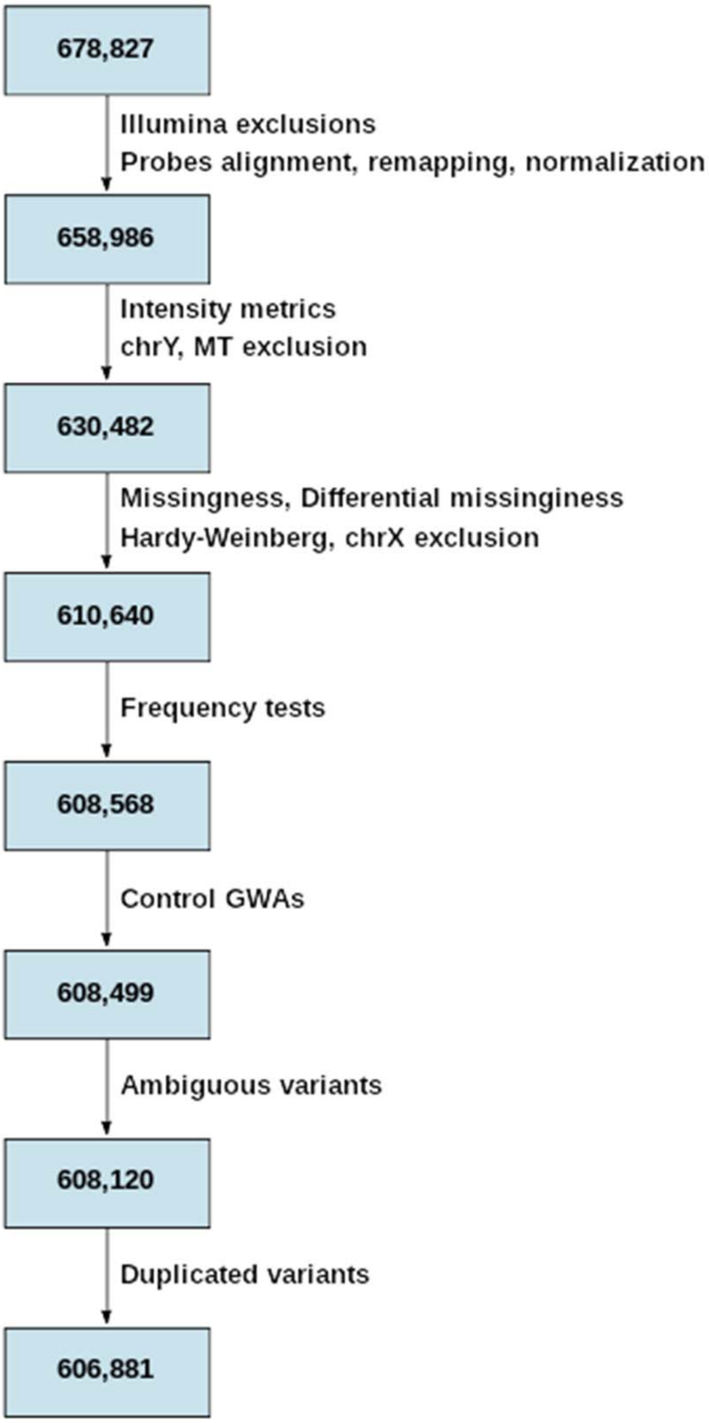
